# Tracing the international arrivals of SARS-CoV-2 Omicron variants after Aotearoa New Zealand reopened its border

**DOI:** 10.1101/2022.07.12.22277518

**Authors:** Jordan Douglas, David Winter, Xiaoyun Ren, Andrea McNeill, Michael Bunce, Nigel French, James Hadfield, Joep de Ligt, David Welch, Jemma L Geoghegan

## Abstract

Recently there has been a surge in emergent SARS-CoV-2 lineages that are able to evade both vaccine induced immunity as well as prior infection from the founding Omicron BA.1 and BA.2 lineages. These highly transmissible and evasive lineages are on the rise and include Omicron variants BA.2.12.1, BA.4, and BA.5. Aotearoa New Zealand recently reopened its borders to many travellers, without their need to enter quarantine. By generating 10,403 complete SARS-CoV-2 genomes classified as Omicron, we show that New Zealand is observing an influx of these immune-evasive variants through the border. Specifically, there has been a recent surge of BA.5 and BA.2.12.1 introductions into the community and these can be explained by the gradual return to pre-pandemic levels of international traveller arrival rates. We estimate there is one Omicron transmission event from the border to the community for every ∼5,000 passenger arrivals into the country, or around one introduction event per day at the current levels of travel. Given the waning levels of population immunity, this rate of importation presents the risk of a large wave in New Zealand during the second half of 2022. Genomic surveillance, coupled with modelling the rate at which new variants cross the border into the community, provides a lens on the rate at which new variants might gain a foothold and trigger new waves of infection.

## Introduction

At the beginning of the coronavirus disease 2019 (COVID-19) pandemic, Aotearoa New Zealand closed its borders in order to quell the addition of further outbreaks in the community [1,2] (March 2020). These border control measures greatly limited entries and required those who were able to enter to spend at least 14 days at a dedicated managed isolation and quarantine (MIQ) facility upon arrival [3]. Due to its geographical isolation, the New Zealand border was able to be tightly regulated. Coupled with a stringent local response (including stay-at-home orders, contact tracing, and isolation of cases [4]), this strategy resulted in the elimination of COVID-19 in New Zealand by May 2020 [5,6]. This elimination phase, which lasted until late 2021, saw several small but quickly contained outbreaks, which leaked from MIQ facilities, cargo vessels, and other channels through the border [3]. Between May 2020 and July 2021, the country recorded a total of 1390 cases and five deaths. Real-time genomic surveillance played a pivotal role in sustaining this state of elimination [3,7].

The border restrictions remained until the trans-Tasman travel ‘bubble’ opened in April 2021, which enabled quarantine-free travel between New Zealand and Australia (Figure 1), who at the time was also pursuing an elimination strategy [8,9]. However, the travel bubble was suspended in July 2021 due to Australia’s difficulty in controlling the emergent Delta variant of concern (VoC). Shortly afterwards, the Delta variant entered the New Zealand community; it likely leaked from an MIQ facility via a traveller from Australia [10]. Unlike previous variants, Delta spread widely and quickly and was unable to be fully controlled, thus leading New Zealand (following a nationwide vaccine rollout, Figure 1) to abandon its elimination strategy in favour of suppression by early October 2021 [11]. By early 2022, the even-more infectious Omicron VoC (BA.1 and BA.2) had entered the community and quickly outcompeted Delta as it had done globally [12-14]. Border controls were gradually relaxed and the MIQ system was abandoned in favour of pre-departure and on-arrival testing.

**Figure 1.**
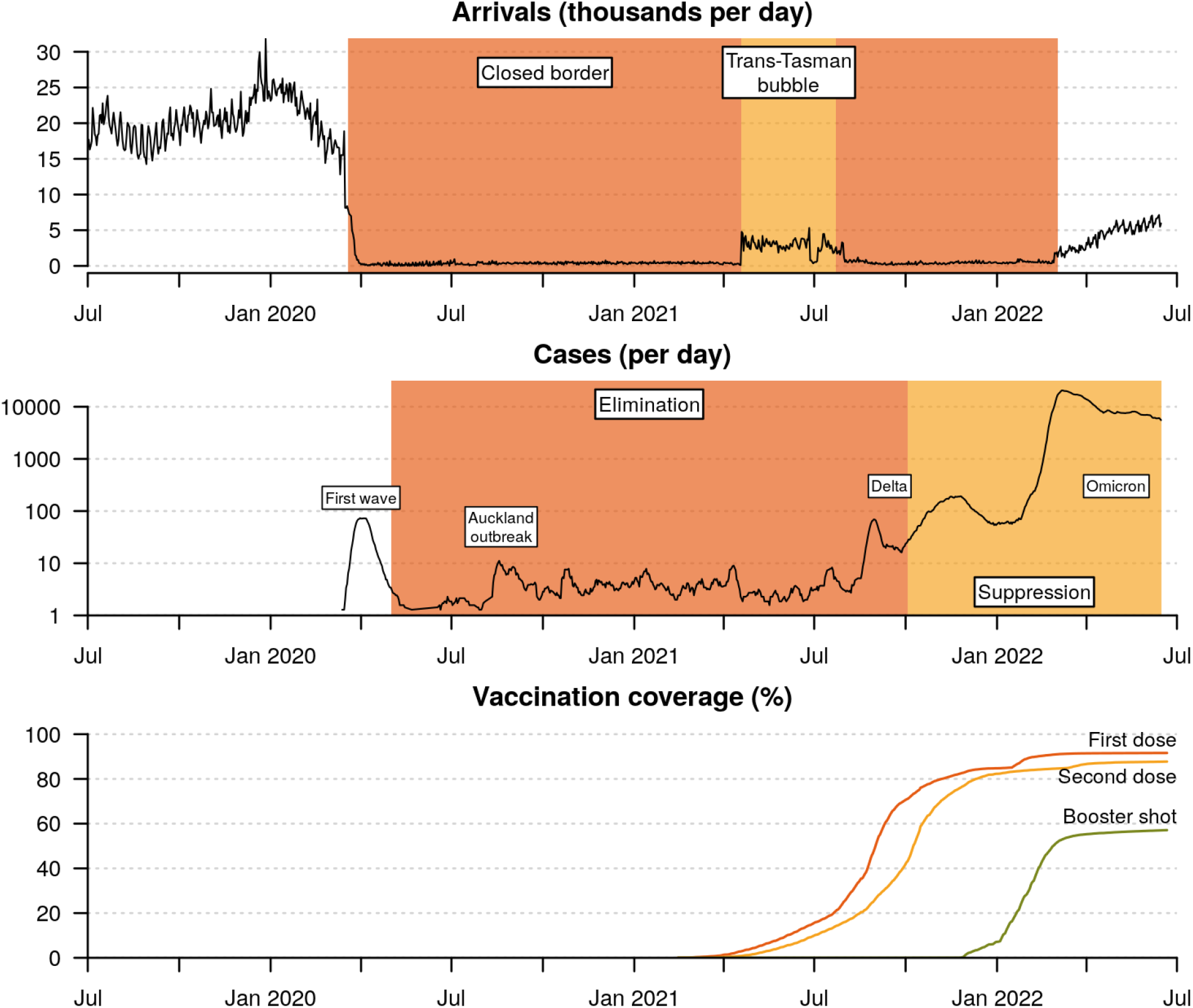
Timeline of the New Zealand COVID-19 pandemic. Daily cases are averaged across a one week period, and include cases in both MIQ facilities as well as the community. Vaccination coverage is expressed as a percentage of the eligible community (5+ years of age; or 94% of the total population). The genomic epidemiology of the first three waves have been characterised elsewhere - First wave: [1]; Auckland August outbreak: [7]; Delta wave: [10].

Unlike other *Severe acute respiratory syndrome coronavirus 2* (SARS-CoV-2) variants, Omicron includes multiple subvariants, termed BA.1 - BA.5. Omicron variants are characterised by at least 50 nonsynonymous mutations compared with ancestral genomes, with a large proportion of these concentrated in the receptor binding domain of the spike protein, resulting in Omicron having a growth advantage of ∼2-4 times over Delta [15]. In the first half of 2022 New Zealand had recorded ∼1.2 million COVID-19 cases - genomic surveillance estimated that around 22% of these infections were BA.1 and 74% BA.2.

Shortly after BA.2 triggered additional waves across the globe, three further Omicron lineages - BA.2.12.1, BA.4 and BA.5 - were linked to another rise in cases globally [16,17]. Both BA.4 and BA.5, which only differ from one another outside of the spike protein, possess spike mutations L452R and F486V, offering both increased binding affinity and enhanced immune escape with an estimated growth advantage of 0.08 and 0.14 over BA.2, respectively [18]. The ability to seemingly evade vaccine- and infection-induced immunity provides BA.4 and BA.5 with this growth advantage. The apparent continued genomic diversification of Omicron lineages highlights the need to tightly monitor its evolution and dispersal.

From March 2022, MIQ ceased for fully vaccinated New Zealand citizens, residents, and work visa holders. People arriving into New Zealand were instead required to undertake COVID-19 rapid antigen tests on days 0-1 and 5-6 of arrival, without the need to isolate. Arrivees who test positive, are required to self-isolate for seven days (previously ten, or fourteen) and undergo a swab and PCR test, which could also be sent for whole genome sequencing. Household contacts are also required to self isolate. Under these more relaxed border settings, the rate of international arrivals rose from fewer than 500 to over 5,000 per day between March and June 2022 (Figure 1).

The move from elimination to suppression and then reopening of borders was justified not just by elimination no longer being obtainable, but also by recognising that the New Zealand population was well vaccinated and boosted. By late 2021, over 80% of the eligible population (5 years+) had received two doses of the Pfizer BioNTech vaccine. By 15 Jun 2022, the figure was 88% with two doses and 57% had received at least one booster, albeit with some significant waning of immunity given the plateau since early 2022 (Figure 1).

Reopening the border, as expected, increased the risk of emergent COVID-19 variants more rapidly entering the community. By 1 August 2022, New Zealand will be fully open to tourists and travellers from anywhere in the world meaning that daily arrivals are expected to quickly return to pre-pandemic levels of 15,000-30,000 over the coming months. In this study we evaluate the impact that recent changes to the border are having on New Zealand’s ability to control COVID-19. Specifically, we monitor the arrival of Omicron variants (BA.1, BA.2, BA.2.12.1, BA.4, and BA.5) from overseas into the New Zealand community and we present a framework for monitoring future variants of concern.

## Results

### Omicron genomics

We generated 10,403 complete SARS-CoV-2 genomes, designated as the Omicron VoC, sampled between 8 Dec 2021 and 15 Jun 2022 (Figure 2). Lineages were designated as BA.1 (n=2,273), BA.2 (n=7,686), BA.2.12.1 (n=175), BA.4 (n=81), and BA.5 (n=188). Here, and throughout the remainder of this article, when we report BA.2 lineages, we are excluding BA.2.12.1 unless specified otherwise. This sample represents 0.8% of the 1,247,900 reported cases in this period. Nasopharyngeal samples were randomly selected to undergo genomic sequencing, however cases linked to the border and those admitted to hospital were sequenced with high priority.

**Figure 2.**
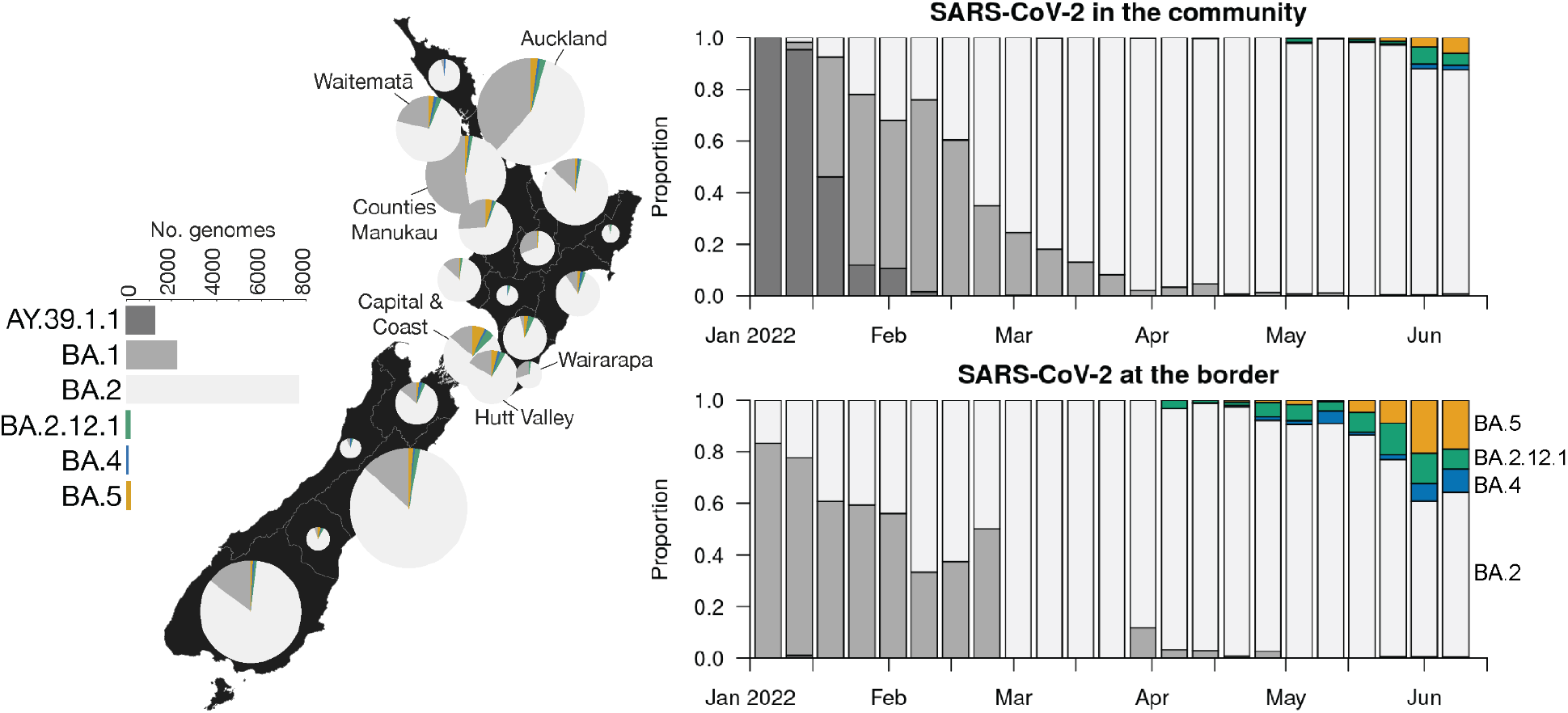
*Left*: Omicron variant distribution by New Zealand district health board, for cases reported between 8 Dec 2021 and 15 Jun 2022. The Delta lineage (AY.39.1.1) is omitted from the map. *Right*: New Zealand genome sequencing, coloured by lineage. A border case is one either in managed isolation after arriving in New Zealand (MIQ era), or one with overseas travel history in the past seven days (post-MIQ era). BA.2 metrics are non-inclusive of BA.2.12.1.

By February 2022, Omicron BA.1 and BA.2 had outcompeted the prevailing Delta VoC (B.1.617.2; lineage AY.39.1.1) in the community, and BA.2 subsequently outcompeted BA.1. At the time of writing, BA.2.12.1, BA.4, and BA.5 are on the rise in New Zealand, but BA.2 continues to dominate (Figure 2).

### Counting Omicron introductions

We describe a framework for tracing SARS-CoV-2 introductions from overseas into the community. Here, we define a ‘*global’* case as one who tested positive either overseas, during their managed isolation period after arriving in New Zealand, or within seven days of arriving in New Zealand (after the MIQ system was abolished). We define a *‘community’* case as one based in New Zealand, and without any recent (i.e. within the past seven days) overseas travel history. The New Zealand Ministry of Health has annotated all of its cases with such labelling. Thus, we define an *‘introduction’* (or an *‘arrival’*) as a transmission event from *global* to the *community*.

We estimated the number of SARS-CoV-2 introductions into the New Zealand community using both global and local genomic sequences (as described in Methods). These results show that BA.1 and BA.4 were only introduced a few times, while BA.2, BA.2.12.1, and BA.5 were introduced significantly more frequently (Figure 3). The majority of these introductions did not lead to any secondary infections in the community. BA.1 and BA.2 were each associated with 1 or 2 large outbreaks represented by over 100 genomic samples, while the outbreaks detected among the younger variants have been smaller (Figure 4). There were several instances of New Zealand border cases who appear to have acquired their infection within the New Zealand community (depicted by red stars in the tree at the top of Figure 3), but the majority (75% across all five analyses) of transitions were in the reverse direction (i.e. introductions). Most of these export events were BA.2 (Figure S2), which is an intuitive result given that a recent arrivee is probably about as likely to receive a BA.2 infection from New Zealand as they were from overseas.

**Figure 3.**
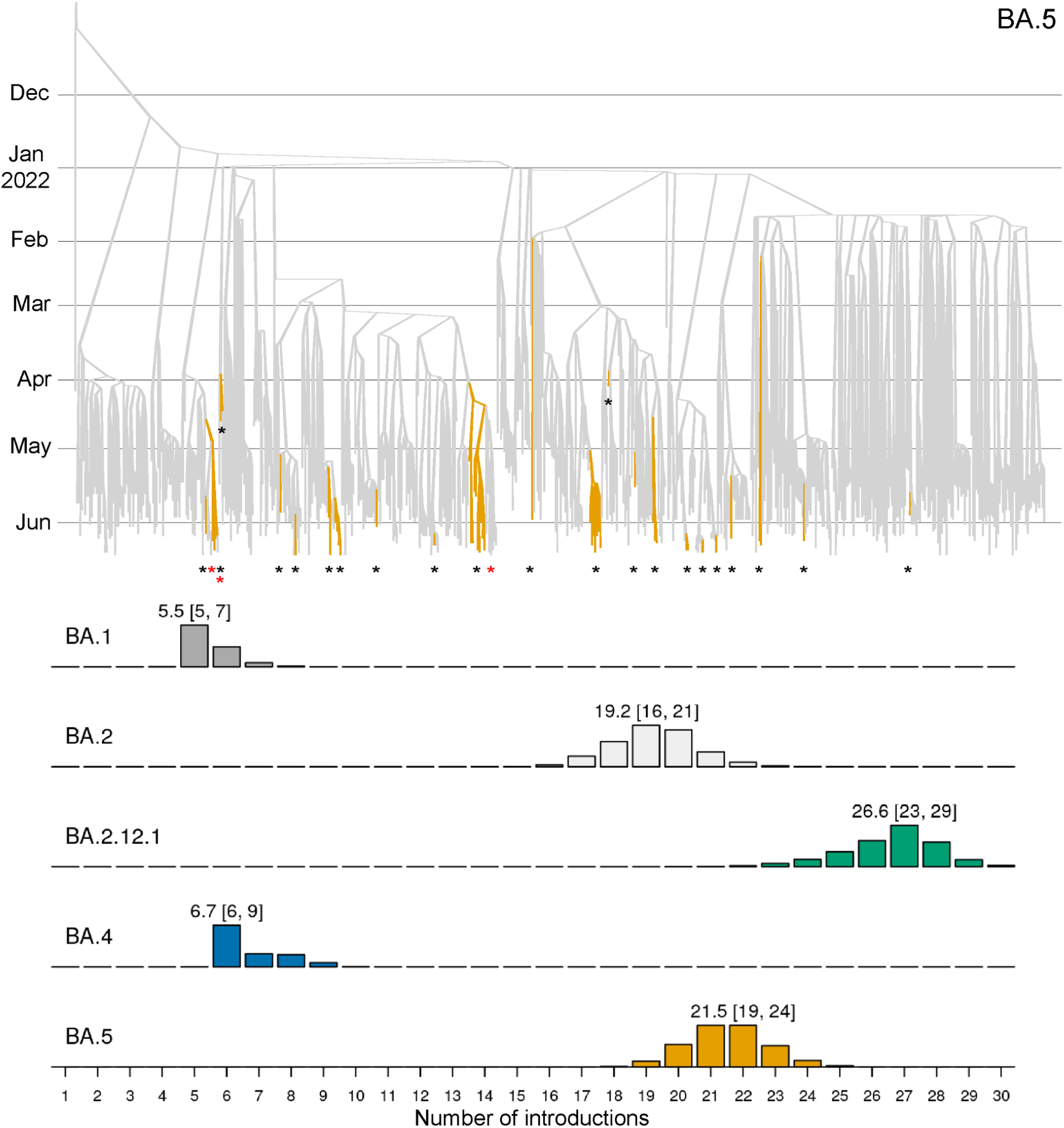
*Top*: summary tree of the BA.5 analysis. Lineages are coloured by world (grey) or community (orange). Introductions from the world into the community are indicated by black stars *, while export events from the community to the world or border and indicated by red stars *. Additional trees can be found in Supporting Information. *Bottom*: posterior distribution of introduction counts (across all trees in the posterior distributions). The y-axes are proportional to Bayesian posterior support. The means [and 95% credible intervals] are indicated.

**Figure 4.**
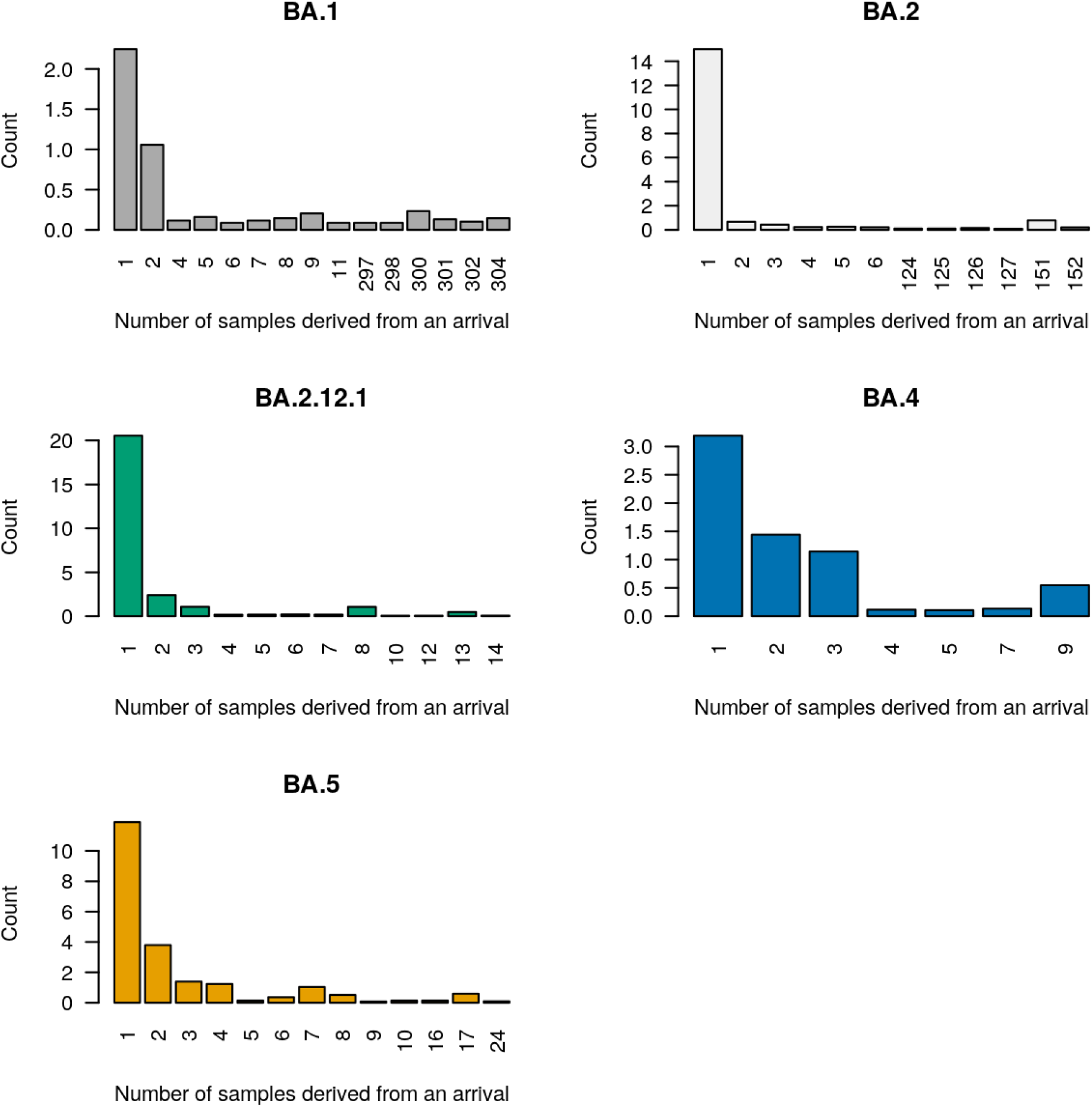
Posterior distribution of sample count (or tree leaf count) resulting from each introduction event. Zero counts are not shown on the horizontal axis. For each variant, 60-80% of all introductions were singletons and did not lead to any detected secondary infection.

### Tracing Omicron arrivals from the border

The first quarter of 2022 was characterised by several BA.1 and BA.2 introductions (Figure 5), some of which spread widely through the New Zealand population. In contrast, the second quarter was characterised by BA.2.12.1, BA.4, and BA.5 introductions. Notably, we estimated 19-24 introductions of BA.5, and 23-29 for BA.2.12.1, into the community since they first arrived in April 2022. Remarkably, these are higher than the 16-21 BA.2 introductions despite its ongoing introductions since 2021. In our sample, there was an average delay of 35 days, and a 95% credible interval of 1 - 105 days, between an estimated introduction time and the lineage being detected by genomic surveillance (Figure 5). This is reflective of our global sampling protocol - a more comprehensive global genome sample would likely reduce this lag.

**Figure 5.**
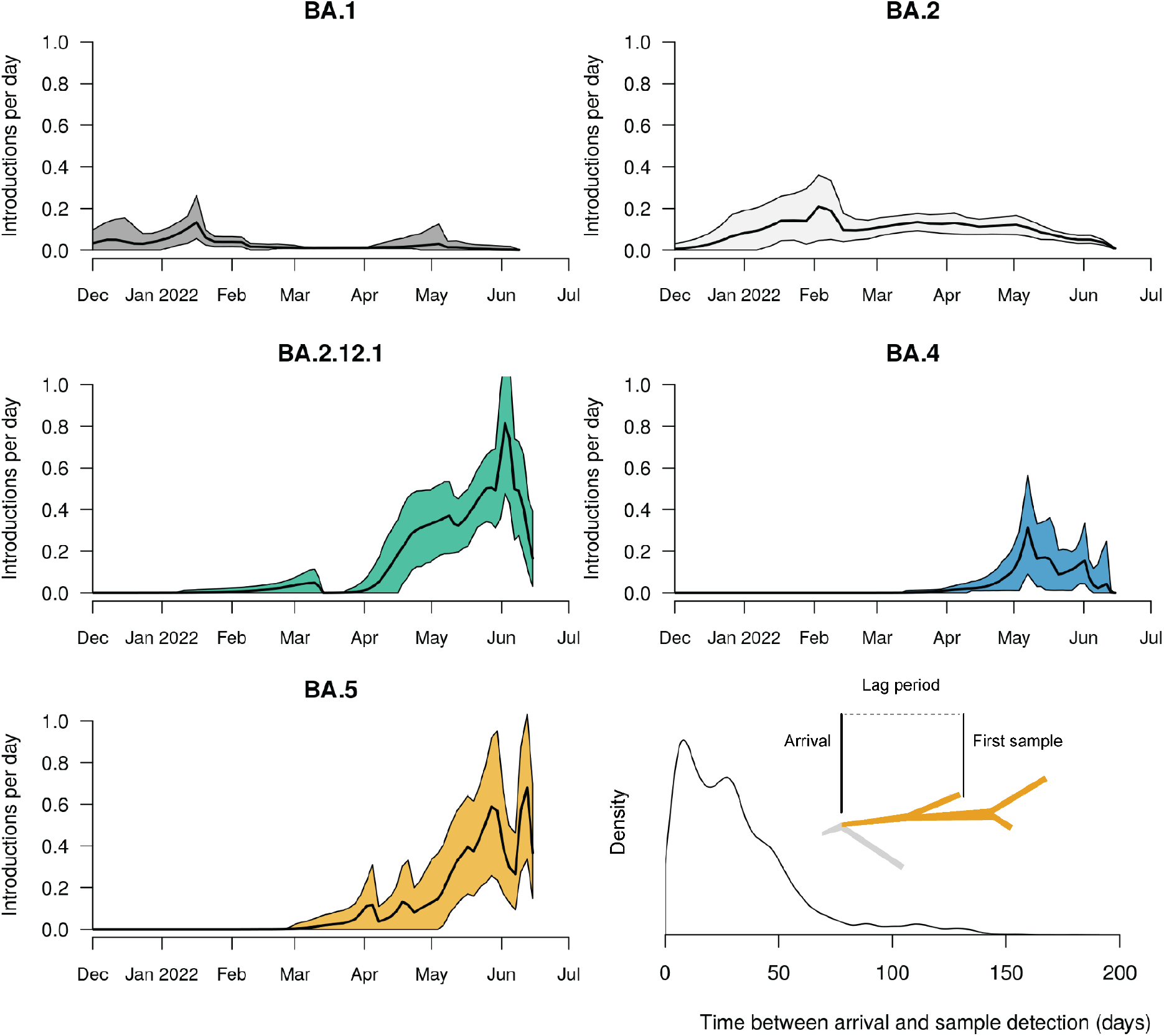
Estimated arrival rate of Omicron subvariants into New Zealand. Mean estimates (black line) and 95% credible intervals (shaded) are indicated. *Bottom right*: Lag time between when a lineage is estimated to have entered the community, and when the first case is detected by genomic surveillance. The meaning of lag time is illustrated in the figure.

The most recent surge of Omicron introductions into the community (namely BA.2.12.1 and BA.5) can be largely explained by the relaxation of New Zealand’s border restrictions, including an end to the MIQ system in early March. We compared the estimated rate of Omicron arrivals with the recorded number of border crossings into New Zealand (i.e. passenger arrivals). These results show a strong correlation between the daily arrival rate of passengers into the country and the estimated daily arrival rate of Omicron into the community (Figure 6). We fit a linear regression model to these data and identified a strong positive linear relationship between the two (R^2^ = 0.76). The slope coefficient was 0.000209, indicating that, under the currently enforced border control measures, there is ∼1 Omicron arrival per 5,000 passenger arrivals, or around one per day at the current levels of travel.

**Figure 6.**
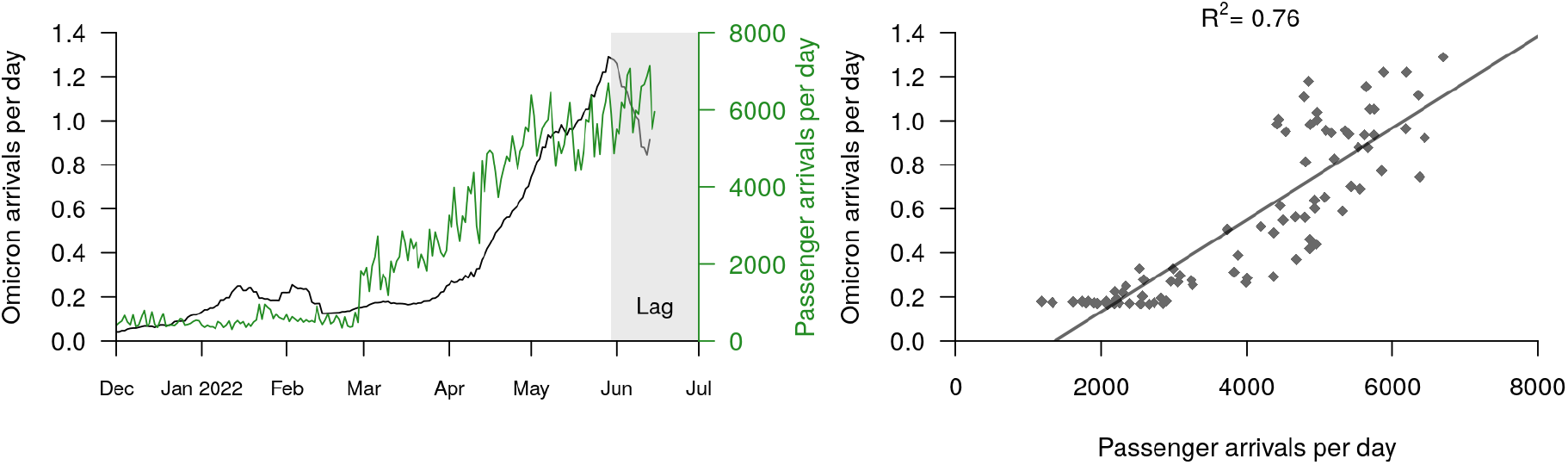
*Left*: Estimated Omicron arrivals (into the community) and recorded passenger arrivals (into the country). The black curve is equal to a smoothed sum of the four black curves in Figure 5. The Omicron arrival rate appears to drop off at the start of June, but this is simply due to a lag between lineages arriving at the border and then being detected in the community by genomic surveillance. *Right*: Omicron arrivals can be explained by passenger arrivals into New Zealand. This linear model was built from the two curves in the left panel, restricted from after the border opened (3 Mar) until available genomic data starts to lag (30 May).

## Discussion

Since the start of the COVID-19 pandemic, highly transmissible and immune-evasive SARS-CoV-2 variants have emerged worldwide [19,20]. The Omicron BA.2 lineage is currently dominant at the global scale. However BA.2.12.1, BA.4, BA.5, as well as recent subvariants such as BA.2.75, are on the rise, and possess mutations that are thought to offer enhanced evasion against immunity induced by vaccines and prior infection [16,17]. Although their severity in humans remains unclear, infection experiments on hamsters suggest that BA.4 and BA.5 may spread more efficiently through lung cells and may be more pathogenic than BA.2 [21]. At the time of writing, BA.5 is rapidly rising and appears it will be the successor of BA.2 as the globally dominant lineage, and has already become predominant in many parts of Africa, North America, and Europe [12].

In this study, we focused on the introduction of emergent variants (BA.2.12.1, BA.4, and BA.5) into New Zealand since its borders reopened to quarantine-free travel. The reopening has led to the international traveller arrival rate increasing by an order of magnitude since the start of the year, and could potentially increase a further three-to-four fold following the border’s full reopening to tourists later in the year. In order to evaluate how these, and other potential lineages, are entering the community, we described a framework (using genomic data) for identifying the trajectories of variants from overseas, through the New Zealand border pre-departure and on-arrival testing requirements, and into the community. We distinguish between New Zealand based cases with recent overseas travel (i.e. the border), compared to those without (i.e. the community) and can therefore trace introductions directly into the population. This framework is based on real-time genomic surveillance coupled with Bayesian phylogenetic inference. Recent computational advancements - such as the BICEPS, ORC, and online packages for BEAST 2 [22,23,24] - have made rapid Bayesian phylogenetic inference on large genomic datasets more feasible.

The first quarter of 2022 was characterised by the introduction, and widespread transmission, of Omicron BA.1 and BA.2 into the country (Figure 2), while the second quarter was characterised by multiple introductions of BA.2.12.1, BA.4, and BA.5. We estimated between six (for BA.1) and 27 (BA.2.12.1) introductions of each variant (Figure 3). The preponderance of recent introductions were of the BA.2.12.1 and BA.5 variants, reflecting trends in overseas ‘feeder’ countries. This may also reflect their abilities to evade immunity from vaccinations and previous BA.1 or BA.2 infections [16,17] - each of which are high in the New Zealand population. Community introductions of Omicron variants surged after the New Zealand borders reopened in March 2022, and grew roughly linearly with the daily international arrival rate. Under the current border settings we estimated there is approximately one transmission event into the community for every 5,000 passenger arrivals into the country (Figure 6). Epidemiological models from earlier in the year predicted that a second wave was likely to arise in August or September 2022 due to waning population immunity, but they noted that a variant with a growth advantage could bring that wave forward [25,26]. This could occur imminently after BA.5 gains a foothold in the community, and this is likely to happen sooner with increased international travel.

We have demonstrated that pathogen surveillance at the border can measure the effectiveness of border control measures and provide advance warning of potential outbreaks. However, this approach is not restricted to COVID-19 – it can also be applied to seasonal influenza, respiratory syncytial virus, or the ongoing monkeypox outbreak [27], for example. As new pathogens continue to emerge around the world, monitoring their global transmission and tracing their arrival into unexposed communities remain important tasks for genomic surveillance.

## Methods

### Genomic sequencing of SARS-CoV-2

For cases reported between 8 Dec 2021 and 15 Jun 2022, ∼0.8% (10,403 genomes) of all COVID-19 cases were referred to the Institute of Environmental Science and Research, New Zealand. In brief, viral extracts were prepared from respiratory tract samples in which SARS-CoV-2 was detected by rRT-PCR. Extracted RNA was subjected to whole-genome sequencing using the Oxford Nanopore Technologies R9.4 chemistry by following the Midnight protocol v6 [28], which contains a 1200-bp primer set tiling the SARS-CoV-2 genome.

Consensus genomes were generated through a standardised pipeline (https://github.com/ESR-NZ/NZ_SARS-CoV-2_genomics) based on the original ARTIC bioinformatics pipeline (https://artic.network/ncov-2019/ncov2019-bioinformatics-sop.html). Genomes were designated into lineages using pangolin version 4.0.6 [29].

### Inferring SARS-CoV-2 introductions

We infer introductions from genomic data as follows:

1. Retrieve complete SARS-CoV-2 genomes from around the world (e.g. from the GISAID EpiCov database [30]). These genomes should be from the respective lineage and during an appropriate time frame (in our case 1 Jan – 15 Jun 2022).
2. Sample, without replacement, *N*_*1*_ complete SARS-CoV-2 genomes from this global pool. In order to reduce geographical sampling bias, genomes are sampled uniformly across locations (e.g. England is equally likely to be sampled as Hong Kong). New Zealand genomes are omitted from this sample. These genomes are added to the *global* pool.
3. Sample, without replacement, *N*_*2*_ complete SARS-CoV-2 genomes from the available New Zealand genomes. In order to reduce population sampling bias, genomes are sampled linearly through time (e.g. 5 Jan 2022 is equally likely to be sampled as 5 May 2022). The genomes which are labelled as *border* cases are added to the *global* pool, and the remainder are added to the *community* pool. This labelling was provided as epidemiological case metadata by the New Zealand Ministry of Health, and we are unable to verify its accuracy.
4. Generate a multiple sequence alignment from the two sampled pools (here we used NextAlign [31] with Wuhan-Hu-1 (NC_045512.2) as a reference).
5. Run a Bayesian two deme discrete phylogeographic analysis on the alignment (described in next subsection).
6. Omicron introductions are estimated as transitions from the *global* deme to the *community* deme in the inferred phylogenetic trees.

We applied this procedure for each of BA.1, BA.2, BA.2.12.1, BA.4, and BA.5, where *N*_*1*_=400 and *N*_*2*_=400. It is important that the *community* pool is not significantly larger than the *global* pool, else the discrete phylogeography model can become unreliable [2,32]. We have used a similar procedure for inferring introductions into New Zealand in previous work [2], as have others for Brazil [33], Rwanda [34], and Europe [35], and has been reviewed elsewhere [36].

### Phylogenetic analysis

Bayesian phylogenetic inference was performed using BEAST 2.6 [37]. We modelled transitions between the two demes (*global* and *community*) using a discrete phylogeography (DPG) model [38]. Under this model, the geographic transition rate had a LogNormal(−0.738, 0.3) prior distribution, the relative transition rate from *global* to the *community* was sampled from LogNormal(4.29, 0.8), while the reverse rate was fixed at 1. This prior assumption means that imports into the community are expected to be significantly more frequent than exports back to the *global* deme, and is used to prevent back-and-forth transitions from appearing too often in the tree. The root of the phylogenetic tree is assumed to belong to the *global* deme. We used an efficient implementation of the Bayesian skyline tree prior implemented in the BICEPS package [22], where the first effective population size is drawn from a Gamma(rate=b, shape=2) distribution, where b ∼ LogNormal(−2.43, 0.5). Nucleotide substitution was modelled using an HKY model [39] with frequencies estimated from a Dirichlet(1,1,1,1) distribution, and a transition-transversion ratio drawn from a LogNormal(1, 1.25) prior. The molecular substitution rate was estimated from a LogNormal(−6.9, 0.05) prior. We used adaptive-weight operators from the ORC package [23] and adaptive variance multivariate normal distribution operators [40] to improve convergence during Bayesian Markov chain Monte Carlo (MCMC). Two independent MCMC chains were run under each lineage, and their convergences were diagnosed using Tracer [41] Each analysis had over 200 effective samples for all relevant parameters. Our BEAST 2 XML file template is uploaded as supplementary data. Phylogenetic tree posterior distributions were summarised as the maximum clade credibility tree [42] and visualised using UglyTrees [43].

## Supporting information

BEAST2 XML file

BA.1 GISAID acknowledgements

BA.2.12.1 GISAID acknowledgements

BA.2 GISAID acknowledgements

BA.4 GISAID acknowledgements

BA.5 GISAID acknowledgements

## Data Availability

All data produced will be made available online

## Data Availability

All SARS-CoV-2 genomic sequence data are available on GISAID. Our BEAST 2 XML file template (with sequence data removed) is uploaded as supplementary data. Case, death, and vaccination data were taken from a New Zealand Ministry of Health GitHub repository (https://github.com/minhealthnz/nz-covid-data; accessed 30 Jun 2022). New Zealand passenger arrival data were taken from the Statistics New Zealand International travel provisional records (https://www.stats.govt.nz/indicators/international-travel-provisional; accessed 30 Jun 2022).

## Acknowledgements

Funding for genome sequencing of SARS-CoV-2 was provided by the Ministry of Health of New Zealand. We thank the diagnostic laboratories that performed the initial RT-PCRs and referred samples for sequencing as well as the public health units for providing epidemiological data. We thank all those who have contributed SARS-CoV-2 sequences to GISAID. JLG is funded by a New Zealand Royal Society Rutherford Discovery Fellowship (RDF-20-UOO-007) and New Zealand Health Research Council Grant (22/138). The authors wish to acknowledge the use of New Zealand eScience Infrastructure (NeSI) high performance computing facilities, consulting support and/or training services as part of this research. New Zealand’s national facilities are provided by NeSI and funded jointly by NeSI’s collaborator institutions and through the Ministry of Business, Innovation & Employment’s Research Infrastructure programme. URL https://www.nesi.org.nz.

## Ethics statement

Nasopharyngeal samples that had positive results for SARS-CoV-2 by real-time reverse transcription PCR were obtained from medical diagnostic laboratories located throughout New Zealand. Under contract for the New Zealand Ministry of Health, the Institute of Environmental Science and Research has approval to conduct genomic sequencing and phylogenetic analysis for surveillance of notifiable diseases.

## Supporting Information

**Figure S1.**
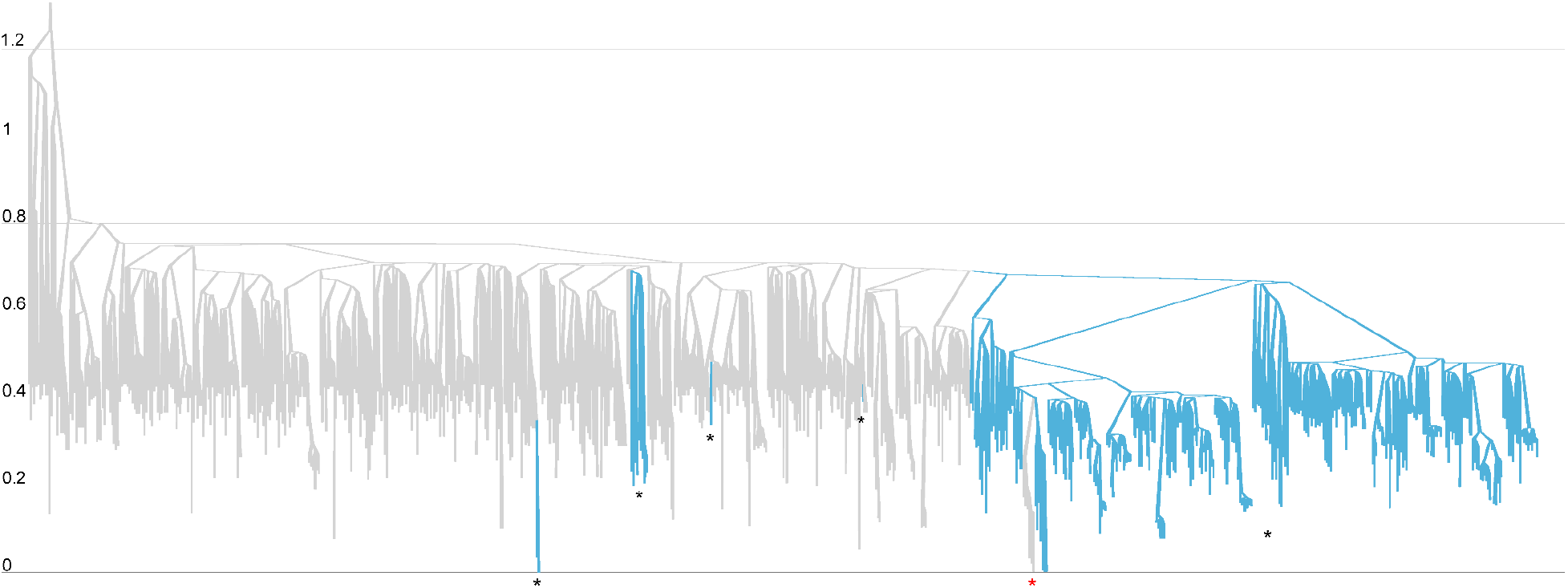
Summary tree of the BA.1 analysis. Lineages are coloured by world (grey) or community (blue). Tree heights are in units for years, where time 0 is 15 Jun 2022. Introduction events are indicated by black stars *, while the reverse events are indicated by red stars *.

**Figure S2.**
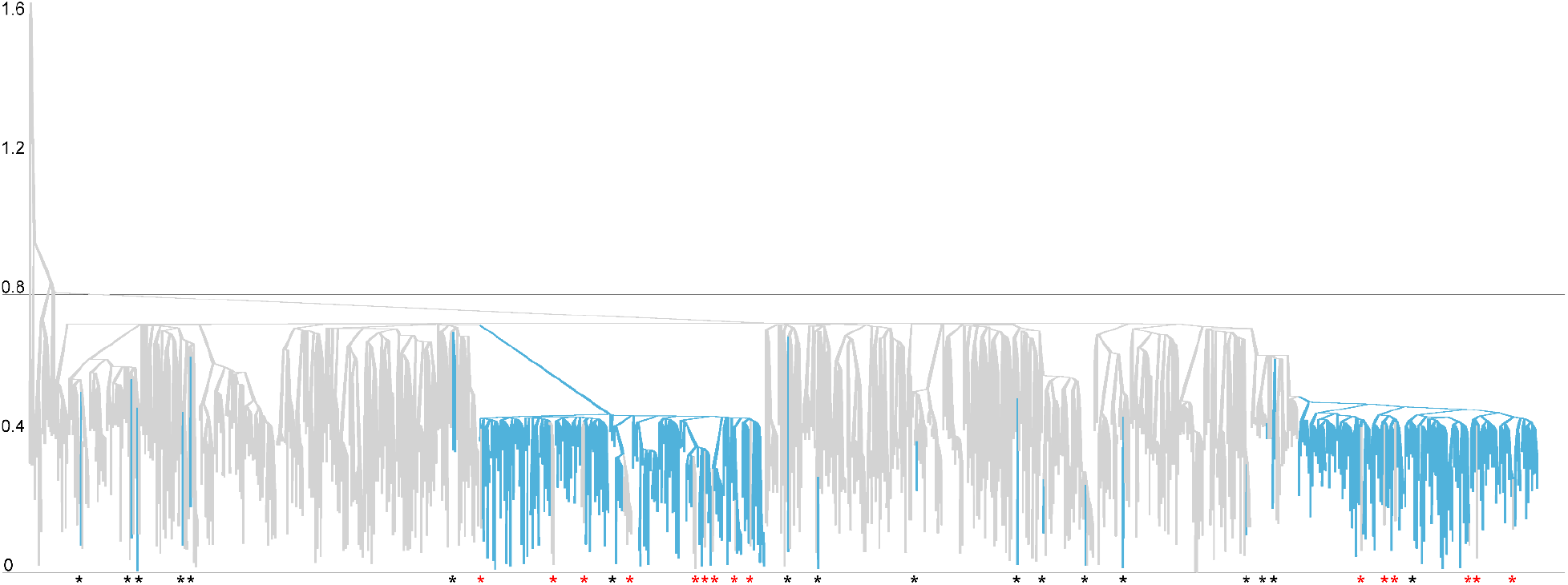
Summary tree of the BA.2 analysis. Lineages are coloured by world (grey) or community (blue). Tree heights are in units for years, where time 0 is 15 Jun 2022. Introduction events are indicated by black stars *, while the reverse events are indicated by red stars *.

**Figure S3.**
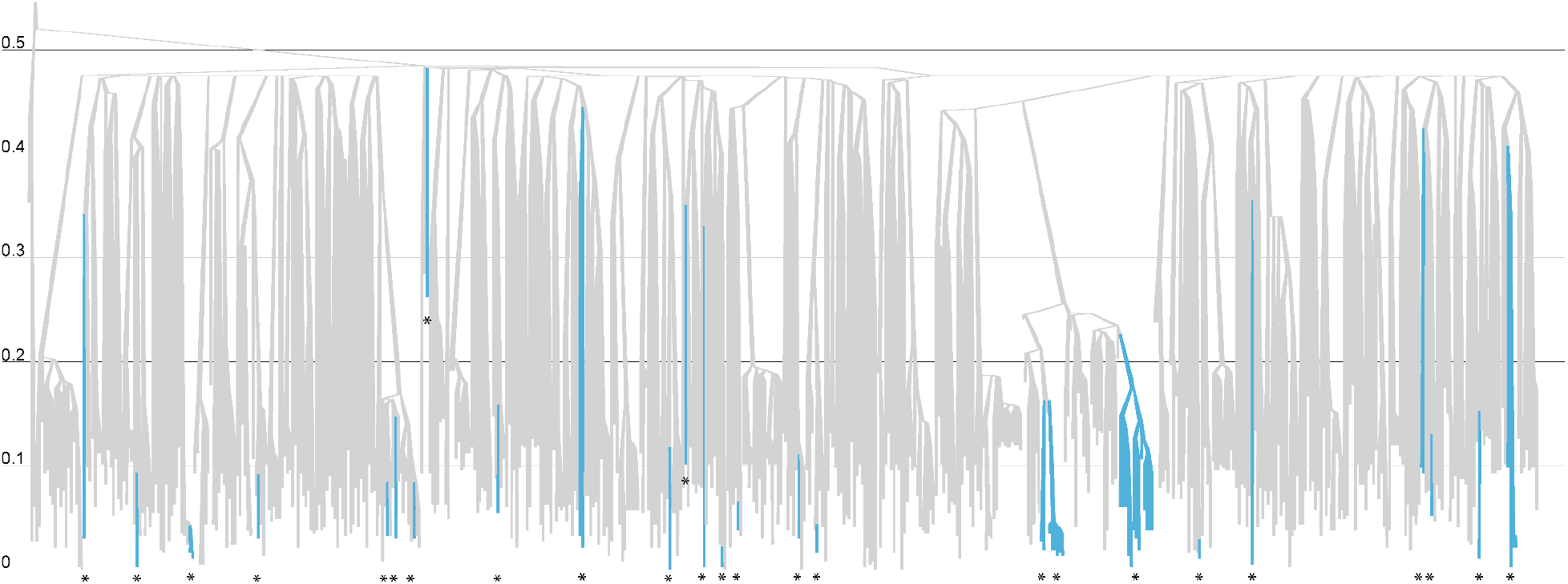
Summary tree of the BA.2.12.1 analysis. Lineages are coloured by world (grey) or community (blue). Tree heights are in units for years, where time 0 is 15 Jun 2022. Introduction events are indicated by black stars *.

**Figure S4.**
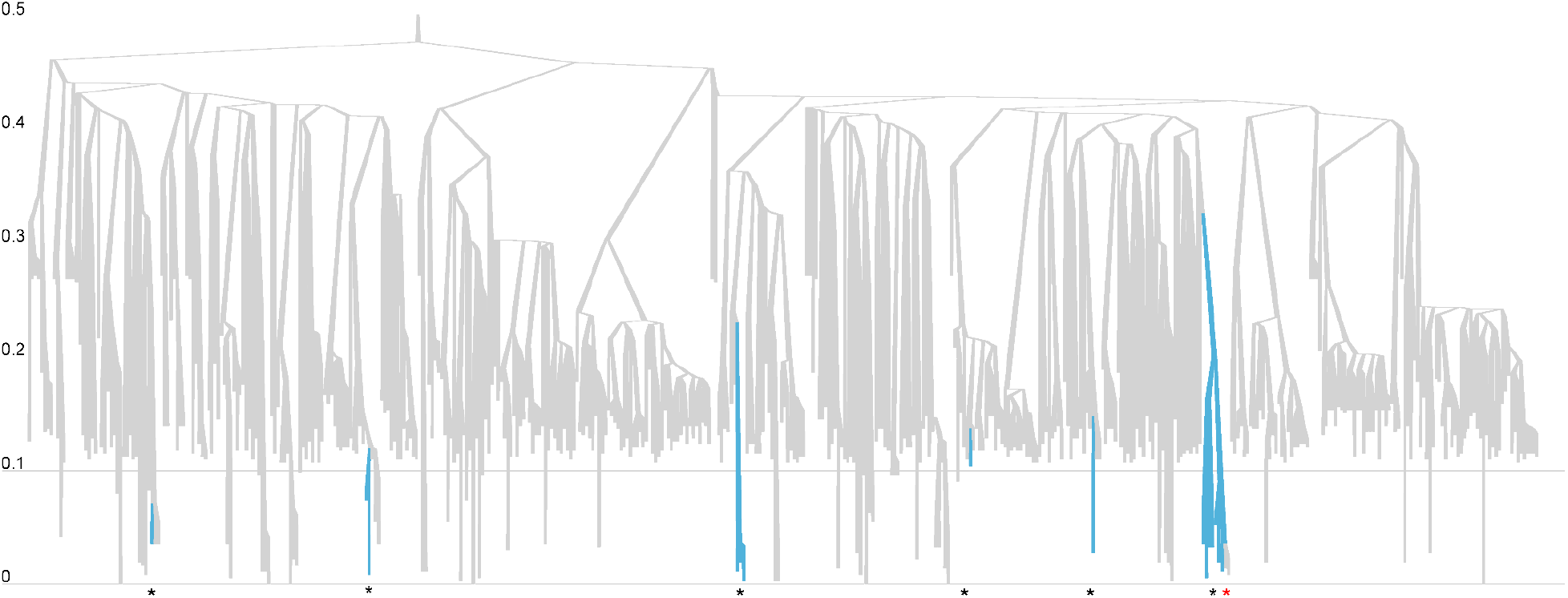
Summary tree of the BA.4 analysis. Lineages are coloured by world (grey) or community (blue). Tree heights are in units for years, where time 0 is 15 Jun 2022. Introduction events are indicated by black stars *, while the reverse events are indicated by red stars *.

## Notes

### Competing Interest Statement

The authors have declared no competing interest.

### Author Declarations

The New Zealand Ministry of Health gave ethical approval for this work.

