## Supplementary material for "Tracing the international arrivals of SARS-CoV-2 Omicron variants after Aotearoa New Zealand reopened its border": BA.1 GISAID acknowledgements

We gratefully acknowledge the following Authors from the Originating laboratories responsible for obtaining the specimens, as well as the Submitting laboratories where the genome data were generated and shared via GISAID, on which this research is based.

All Submitters of data may be contacted directly via www.gisaid.org

Authors are sorted alphabetically.

Acknowledgement EPI_SET Identifier: EPI_SET_20220706be

| Accession ID | Originating Laboratory | Submitting Laboratory | Authors |
|---|---|---|---|
| EPI_ISL_11686030 | ACT Pathology | Schwessinger Lab, Research School of Biology, College of Science | Abigail Graetz; Ashley Jones; Austin Bird; Bayantes Dagvadorj; Benjamin Schwessinger; Carl McCombe; Catalina Barragán Quintero; Craig Kennedy; Evie Hodgson; Gabrielle Smith; Karina Kennedy; Makenna Short; Rachel Leonard; Rene Riedelbauch; Rita Tam; Robyn Hall; Scott Ferguson |
| EPI_ISL_10864979 | AZDelta | AZ Delta Medical Laboratories in Roeselare, Belgium | Dieter De Smet; Frederik Van Hoecke; Geert Martens; Patrick Descheemaeker; on behalf of AZ Delta COVID-19 Genomics core (member of Genomic surveillance of SARS-CoV-2 in Belgium network) |
| EPI_ISL_8886635 | Addu Equitorial Hospital | Addu Equitorial Hospital | D. Fathmath Nazla Rafeeq; Dr. Ibrahim Afzal; Mr. Ibrahim Nishan Ahmed; Ms. Aishath Shuhudha; Ms. Aminath Shazleena Abdul Rahman; Ms. Soafy Mohamed |
| EPI_ISL_9510038 | BBMP- West | inStem, NCBS-INSACOG | Aswin SaiNarain; Dasaradhi Palakodeti; Uma Ramakrishnan |
| EPI_ISL_9648072, EPI_ISL_9896672, EPI_ISL_11992199 | Bayerisches Landesamt fuer Gesundheit und Lebensmittelsicherheit (LGL) | Bayerisches Landesamt fuer Gesundheit und Lebensmittelsicherheit (LGL) | Alexandra Dangel; Andreas Sing; Annika Sprenger; Carola Berger; Laura Weise; Lisa Falk; Nikolaus Ackermann; Sabrina Hepner |
| EPI_ISL_13331313 | Bheri Hospital | Center for Molecular Dynamics Nepal | Bheri Hospital; Birat Nepal Medical Trust; Center for Molecular Dynamics Nepal; Epidemic Intelligence Team; Koshi Hospital; Liverpool School of Tropical Medicine; Nepal Health Research Council; OUCRU Nepal; Sukraraj Tropical & Infectious Disease Hospital; University of Cambridge |
| EPI_ISL_10511959 | Bio Molecular Laboratory | National Institute of Health, Department of Medical Sciences, Ministry of Public Health, Thailand | Archawin Rojanawiwat; Natchaya Khiadsang; Nuttida Thongpramul; Pakorn Piromtong; Pilailuk Okada; Sirikanda Wimol; Siripaporn Phuygun; Sunthareeya Waicharoen; Suratchana Mitrat; Thanutsapa Thanadachakul |
| EPI_ISL_12702789 | BioneXt Lab | Laboratoire national de sante, Microbiology, Microbial Genomics Platform | Anke Wienecke-Baldacchino; Cynthia Oxacelay; Elodie Solarino; Eric Hugoson; Fatu Djabi; Jessica Tapp; Lise Pignon; Raoul Salmon; SibelCatherine Ragimbeau; Tamir Abdelrahman; Virginie Jover |
| EPI_ISL_11325766 | Biotecnology and Genetics Laboratory - Instituto Nacional de Saúde | Biotecnology and Genetics Laboratory - Instituto Nacional de Saúde | Aventina Macuacua; Cacildo Magul; Mirela Pale; Nalia Ismael; Nuro Abilio; Samuel Chumane; Sofia Viegas |
| EPI_ISL_11905689 | Bushehr University of Medical University | National Influenza Center | A Nejati; Adel Abedi; J Yavarian; K Sadeghi; Marzieh Faraji-Zonouz; NZ Shafiei Jandaghi; Nastaran Ghavami; Sevrin Zadheidar; V Salimi; and T Mokhtari Azad |
| EPI_ISL_9210610 | CENTOGENE Frankfurt Laboratory: Niederlassung Industriepark Höchst | Robert Koch Institute | |
| EPI_ISL_9704077 | CERBALLIANCE CENTRE VAL DE LOIRE | CERBA HealthCare | Bénédicte Roquebert; Laura Verdurme; Mathilde Roussel; Sabine Trombert; Stéphanie Haim-Boukobza |
| EPI_ISL_10314479 | CERBALLIANCE COTE D'AZUR | CERBA HealthCare | Bénédicte Roquebert; Laura Verdurme; Mathilde Roussel; Sabine Trombert; Stéphanie Haim-Boukobza |
| EPI_ISL_9470063 | CERBALLIANCE PARIS ET IDF EST | CERBA HealthCare | Bénédicte Roquebert; Laura Verdurme; Mathilde Roussel; Sabine Trombert; Stéphanie Haim-Boukobza |
| EPI_ISL_9300755 | CHTMAD - Vila Real | Instituto Nacional de Saude (INSA) | Borges et al |
| EPI_ISL_9156927 | Center for Laboratory Medicine | Center for Laboratory Medicine, ZLMSG | Yannick Gerth |
| EPI_ISL_10635619 | Central Health Laboratory(Virology Dept) , Ministry of Health and Wellness | Central Health Laboratory(Virology Dept) , Ministry of Health and Wellness | Bahadoor BS; Janoo N; Mathur H; Ramuth M; Sonoo J; Sujeewon C; Ubheeram A |
| EPI_ISL_11895191, EPI_ISL_11895203 | Central Public Health Laboratory - LACEN -Bahia, Salvador, Brazil | Central Public Health Laboratory - LACEN -Bahia, Salvador, Brazil | Arabela Leal; Felicidade Pereira; Gabriela Menezes; Jaqueline Gomes; Jessica Araujo; Luciana Oliveira; Luiz Alcantara; Marcela Gómez; Marta Giovanetti; Vagner Fonseca; Vanessa Nardy |
| EPI_ISL_11579838 | Centre hospitalier de Châtellerault | Centre Hospitalier Universitaire (CHU) Poitiers | Agnes Beby-Defaux; Jean Philippe Da Mota; Luc Deroche; Magali Garcia; Manon Prat; Nicolas Leveque |
| EPI_ISL_9431841 | Centrum Medyczne Medyk Sp. z o. o., Sp. k. Zakład Diagnostyki Medycznej | 1. Academic Center for Pathomorphological and Genetic-Molecular Diagnostics ltd, Bialystok, Poland 2. National Institute of Public Health - National Institute of Hygiene, Warsaw, Poland | Anetta Sulewska; Jacek Nikliński; Janusz Dzięcioł; Joanna Kiśluk; Katarzyna Zacharczuk; Konrad Raczkowski; Magdalena Nowakowska; Małgorzata Sadkowska-Todys; Piotr Karabowicz; Piotr Majewski; Przemysław Biecek. Joanna Reszeć; Radosław Charkiewicz; Tomasz Wołkowicz |
| EPI_ISL_8931581, EPI_ISL_8931629 | Cliniques universitaires Saint-Luc | UCLouvain/IREC/MBLG-CTMA | Benoit Kabamba Mukadi; Bertrand Bearzatto; Jean-Luc Gala; Nicolas Pinte; Paul Blanpain; Simon Ophélie; Valentin Coste |
| EPI_ISL_9227259 | Department Medical Microbiology, Baerum Hospital, Vestre Viken Health Trust | Norwegian Institute of Public Health, Department of Virology | Atiya R Ali; Debech Nadia; Engebretsen Serina Beate; Garcia Llorente Ignacio; Hilde Elshaug; Hilde Nordby Falkenhaug; Hilde Vollan; Jon Bråte; Kamilla Heddeland Instefjord; Karoline Bragstad; Kathrine Stene-Johansen; Line Victoria Moen; Marie Paulsen Madsen; Olav Hungnes; Pedersen Benedikte Nevjen; Rasmus Riis Kopperud |
| EPI_ISL_8582273, EPI_ISL_8927075, EPI_ISL_9120233, EPI_ISL_9453635, EPI_ISL_9499975 | Department of Bacteria, Parasites and Fungi, Statens Serum Institut, Copenhagen, Denmark | Statens Serum Institut Bioinformatics and Microbial Genomics | Danish Covid-19 Genome Consortium |
| EPI_ISL_10204396 | Department of Clinical Microbiology and Center for Genomic Medicine, Rigshospitalet, Copenhagen, Denmark | Statens Serum Institut Bioinformatics and Microbial Genomics | Danish Covid-19 Genome Consortium |
| EPI_ISL_12418400 | Department of Health Technology and Informatics, The Hong Kong Polytechnic University | Department of Health Technology and Informatics, The Hong Kong Polytechnic University | Alan Ka-Lun Wu; Alex Yat-Man Ho; Barry Kin-Chung Wong; Chloe Toi-Mei Chan; David Ho-Keung Shum; Gilman Kit-Hang Siu; Hiu-Yin Lao; Ivan Tak-Fai Wong; Jake Siu-Lun Leung; Kam-Tong Yip; Kenneth Siu-Sing Leung; Kingsley King-Gee Tam; Kitty Sau-Chun Fung; Kristine Luk; Lam-Kwong Lee; Miranda Chong-Yee Yau; Sandy Ka-Yee Chau; Shea Ping Yip; Tak-Lun Que; Timothy Ting-Leung Ng; Wing Cheong Yam; Wing-Hei Lo; Wing-Kin To; Yvette Wai-Man Lai |
| EPI_ISL_11539207, EPI_ISL_11539208, EPI_ISL_11725850, EPI_ISL_12005205 | Department of Laboratory Medicine, Shinshu University Hospital | Department of Laboratory Medicine, Shinshu University Hospital | Akari Miyazaki; Saori Konno; Shohei Shigeto; Tatsuya Negishi; Wakaba Iha |
| EPI_ISL_12650021 | Department of Medical Microbiology - section Molde, Molde Hospital | Norwegian Institute of Public Health, Department of Virology | Atiya R Ali; Debech Nadia; Engebretsen Serina Beate; Garcia Llorente Ignacio; Hilde Elshaug; Hilde Nordby Falkenhaug; Hilde Vollan; Jon Bråte; Kamilla Heddeland Instefjord; Karoline Bragstad; Kathrine Stene-Johansen; Line Victoria Moen; Marie Paulsen Madsen; Olav Hungnes; Pedersen Benedikte Nevjen; Rasmus Riis Kopperud |
| EPI_ISL_10844401, EPI_ISL_11055860, EPI_ISL_11260546, EPI_ISL_12009114 | Department of Microbiology, Laboratoire Hospitalier Universitaire de Bruxelles, Université Libre de Bruxelles | Department of Microbiology, Laboratoire Hospitalier Universitaire de Bruxelles, Université Libre de Bruxelles | Charlotte Michel; Olivier Vandenberg; Sigi Van Den Wijngaert |
| EPI_ISL_11154406 | Department of Public Health Bacau | National Institute of Infectious Diseases-Prof. Dr. Matei Bals Molecular Diagnostics Laboratory | Corina Casangiu; Dan Otelea; Leontina Banica; Marius Surleac; Ovidiu Vlaicu; Petre Milu; Robert Hohan; Simona Paraschiv |
| EPI_ISL_11154299 | Department of Public Health Braila | National Institute of Infectious Diseases-Prof. Dr. Matei Bals Molecular Diagnostics Laboratory | Corina Casangiu; Dan Otelea; Leontina Banica; Marius Surleac; Ovidiu Vlaicu; Petre Milu; Robert Hohan; Simona Paraschiv |
| EPI_ISL_11154559 | Department of Public Health Buzau | National Institute of Infectious Diseases-Prof. Dr. Matei Bals Molecular Diagnostics Laboratory | Corina Casangiu; Dan Otelea; Leontina Banica; Marius Surleac; Ovidiu Vlaicu; Petre Milu; Robert Hohan; Simona Paraschiv |

| | | | |
|---|---|---|---|
| EPI_ISL_11154317 | Department of Public Health Constanta | National Institute of Infectious Diseases-Prof. Dr. Matei Bals Molecular Diagnostics Laboratory | Corina Casangiu; Dan Otelea; Leontina Banica; Marius Surleac; Ovidiu Vlaicu; Petre Milu; Robert Hohan; Simona Paraschiv |
| EPI_ISL_9967428, EPI_ISL_11154380, EPI_ISL_11154698, EPI_ISL_11154704 | Department of Public Health Iasi | National Institute of Infectious Diseases-Prof. Dr. Matei Bals Molecular Diagnostics Laboratory | Corina Casangiu; Dan Otelea; Leontina Banica; Marius Surleac; Ovidiu Vlaicu; Petre Milu; Robert Hohan; Simona Paraschiv |
| EPI_ISL_9967316, EPI_ISL_9967343, EPI_ISL_9967468, EPI_ISL_9967469 | Department of Public Health Mures | National Institute of Infectious Diseases-Prof. Dr. Matei Bals Molecular Diagnostics Laboratory | Corina Casangiu; Dan Otelea; Leontina Banica; Marius Surleac; Ovidiu Vlaicu; Petre Milu; Robert Hohan; Simona Paraschiv |
| EPI_ISL_9967533 | Department of Public Health Timis | National Institute of Infectious Diseases-Prof. Dr. Matei Bals Molecular Diagnostics Laboratory | Corina Casangiu; Dan Otelea; Leontina Banica; Marius Surleac; Ovidiu Vlaicu; Petre Milu; Robert Hohan; Simona Paraschiv |
| EPI_ISL_8648923, EPI_ISL_9614456 | Dept. of Microbiology and Infection Control, Akershus University Hospital HF | Dept. of Microbiology and Infection Control, Akershus University Hospital HF | Alexander Hesselberg Løvestad; Hanne Berggreen; Hege Vangstein Aamot |
| EPI_ISL_9658302, EPI_ISL_10230041 | Diagnostyka. Laboratoria Medyczne. | 1. ViroGenetics - BSL3 Laboratory of Virology, Małopolska Centre of Biotechnology, Jagiellonian University; 2. Diagtron Laboratoria Łukasz Rąbalski | Gromowski, T.; Kowalski, M.; Labaj; Maciej Kosinski; Mazur-Panasiuk, N.; Natalia Derewonko; P.P.; Pyrc, K.; Rabalski L.; Rogalska-Kupiec M.; Swadzba J.; Sylwia Januszczak; Szulc, P.; Wydmanski, W. |
| EPI_ISL_8998400 | Division of Emerging Infectious Diseases, Bureau of Infectious Diseases Diagnosis Control, Korea Disease Control and Prevention Agency | Division of Emerging Infectious Diseases, Bureau of Infectious Diseases Diagnosis Control, Korea Disease Control and Prevention Agency | Ae Kyung Park; Chae Young Lee; Eun-Jin Kim; Hyuck Jin Lee; Il-Hwan Kim; Jeong-Ah Kim |
| EPI_ISL_8929209, EPI_ISL_10773177 | Dutch COVID-19 response team | Medical Microbiology, Maastricht University Medical Centre | Brian van der Veer*; Carmen Reumkens; Christian Hoebe; Erik Beuken; Jozef Dingemans*; Lieke van Alphen; Paul Savelkoul |
| EPI_ISL_9085204, EPI_ISL_10036258, EPI_ISL_10036374, EPI_ISL_10036377, EPI_ISL_11659847, EPI_ISL_11840841 | Dutch COVID-19 response team | National Institute for Public Health and the Environment (RIVM) | Adam Meijer; Afke Vogelzang; AnneMarie van den Brandt; Annelies Kroneman; Bas van der Veer; Chantal Reusken; Dennis Schmitz; Dirk Eggink; Florian Zwagemaker; Harry Vennema; Ivo van Walle; Jeroen Cremer; Jil Kocken; Jordy de Bakker; Karim Hajji; Kim Freriks; Linda van Someren; Lisa Wijsman; Lynn Aarts; Ryanne Jaarsma; Sanne Bos; Sharon van den Brink; on behalf of the national COVID-19 response team |
| EPI_ISL_9234082 | GMERS Medical College, Valsad | Gujarat Biotechnology Research Centre | Akhilesh Modi; Apurvasinh Puvar; Bhadreshsinh Gohil; Chaitanya Joshi; Disha Vora; Janvi Raval; Jaykumar Rangani; Madhvi Joshi; Nimesh Patel; Nitin Savaliya; NitinShukla; Priyank Chavda; Ramesh Pandit; Roshani Mishra; Sonal Sharma; Tasnim Trivedi; Vicky Gandhi; Zarna Patel |
| EPI_ISL_8527573 | Gandhi Medical College and Hospital (GMCH), Secunderabad | NIV Influenza | D.R.Manisha Rani; Dr.G.Sushma Rajya Lakshmi; Dr.K.Nagamani; Dr.Sunitha Pakalapaty |
| EPI_ISL_9191078, EPI_ISL_9709602, EPI_ISL_10807135, EPI_ISL_12695408, EPI_ISL_12695413, EPI_ISL_12739731, EPI_ISL_12739732, EPI_ISL_12740210, EPI_ISL_12982112 | | | |
| see above | Genetica Molecular and Subdepartamento de Virologia ISP Chile | Instituto de Salud Publica de Chile | Andres Castillo; Barbara Parra; Constanza Campano; Ivan Ponce; Jorge Fernandez; Karen Orostica; Marcelo Rojas; Matias Pezoa; Patricia Bustos; Paulo covarrubias; Rodrigo Fasce |
| EPI_ISL_11855026, EPI_ISL_11855027, EPI_ISL_11855030, EPI_ISL_11855042, EPI_ISL_11859468, EPI_ISL_11859477, EPI_ISL_11859490, EPI_ISL_11859500 | | | |
| see above | Gorgas Memorial Institute of Health Studies | Gorgas Memorial Institute of Health Studies | Abdiel Menacho; Alexander A Martinez; Ambar Moreno; Brechla Moreno; Claudia Gonzalez; Danilo Franco; Jessica Gondola; Leyda Abrego; Oris Chavarria |
| EPI_ISL_9850712 | H Divino Espirito Santo - Ponta Delgada | Instituto Nacional de Saude (INSA) | Borges et al |
| EPI_ISL_13169662 | HD-SANTA RITA | Laboratorio Central de Salud Pública | Analía Rojas; Andrea Gómez de la Fuente; Cynthia Vazquez; César Cantero; César Ojeda; Emmanuel Céspedes; Fátima Fleitas; Ivana Fernández; Juan Torales; Julio Barrios; Maria Liz Gamarra; María Jose Duarte; Sandra González; Shirley Villalba; Tania Alfonzo; Wilson Benítez. |
| EPI_ISL_12958919 | HOSP. ALVAREZ BUYLLA | Laboratorio de Virología HUCA | ; Alba L; Alvarez-Arguelles ME; Boga JA; Costales I; Coto E; González-Alba JM; Gómez de Oña J; Martín-Rodríguez G; Melón S; Perez-Martínez Z; Rojo S; Sandoval M |
| EPI_ISL_13169672 | HOSPITAL DISTRITAL-HERNANDARIAS | Laboratorio Central de Salud Pública | Analía Rojas; Andrea Gómez de la Fuente; Cynthia Vazquez; César Cantero; César Ojeda; Emmanuel Céspedes; Fátima Fleitas; Ivana Fernández; Juan Torales; Julio Barrios; Maria Liz Gamarra; María Jose Duarte; Sandra González; Shirley Villalba; Tania Alfonzo; Wilson Benítez. |
| EPI_ISL_13169649, EPI_ISL_13169651, EPI_ISL_13169654, EPI_ISL_13169663, EPI_ISL_13169667 | HR-CIUDAD DEL ESTE | Laboratorio Central de Salud Pública | Analía Rojas; Andrea Gómez de la Fuente; Cynthia Vazquez; César Cantero; César Ojeda; Emmanuel Céspedes; Fátima Fleitas; Ivana Fernández; Juan Torales; Julio Barrios; Maria Liz Gamarra; María Jose Duarte; Sandra González; Shirley Villalba; Tania Alfonzo; Wilson Benítez. |
| EPI_ISL_12422409 | Hospital Alemán | Área de Secuenciación del Laboratorio de Virología del Hospital de Niños Dr. Ricardo Gutierrez on behalf of 'Proyecto Argentino Interinstitucional de genomica de SARS-CoV-2' (PAIS Consortium) | Acuña; D; Eugenia Ibañez; Goya; LE; Lusso; MI; MS; María Paula Della Latta; Nabaes Jodar; Natale; Natalia García allende; S; Valinotto; Viegas, M. |
| EPI_ISL_12476734 | Hospital Borga Roma Verona | Laboratory of Medical Microbiology, University of Antwerp | Basil Britto Xavier; Evelina Tacconelli; Mathias Smet; Matilda Berkell; Surbhi Malhotra-Kumar |
| EPI_ISL_11579837, EPI_ISL_11579849 | Hospital Center de Niort | Centre Hospitalier Universitaire (CHU) Poitiers | Agnes Beby-Defaux; Jean Philippe Da Mota; Luc Deroche; Magali Garcia; Manon Prat; Nicolas Leveque |
| EPI_ISL_11580270 | Hospital Center de Saintonge | Centre Hospitalier Universitaire (CHU) Poitiers | Agnes Beby-Defaux; Jean Philippe Da Mota; Luc Deroche; Magali Garcia; Manon Prat; Nicolas Leveque |
| EPI_ISL_11788147, EPI_ISL_12184538 | Hospital Clínico Universitario Virgen de la Arrixaca | Hospital Clínico Universitario Virgen de la Arrixaca | Laura Moreno Parrado and Luis Javier Gil-Gallardo.; Marina Simón Páez |
| EPI_ISL_11160306, EPI_ISL_11160321 | Hospital General Universitario Gregorio Marañón | Hospital General Universitario Gregorio Marañón | Cristina Rodriguez-Grande; Darío García de Viedma; Jorge Rodríguez-Grande; Julia Suárez; Laura Pérez-Lago; Marta Herranz Martin; Patricia Muñoz; Pedro Sola Campoy; Pilar Catalán; Rosalía Palomino Cabrera; Sergio Buenestado Serrano |
| EPI_ISL_13156314 | Hospital Nacional del Cáncer | Laboratorio Central de Salud Pública | Andrea Gómez de la Fuente; Cesar Cantero; Cynthia Vazquez; Juan Torales; Julio Barrios; Maria Liz Gamarra; Sandra Gonzalez; Shirley Villalba; Tania Alfonzo |
| EPI_ISL_12173793, EPI_ISL_12573062, EPI_ISL_12883432 | Hospital Sao Rafael - H.S.R/BA | Plataforma de Vigilancia Molecular (PVM) - FIOCRUZ/BA | Ana Verena Almeida Mendes; Bruno Bezerril Andrade; Bruno Solano de Freitas Souza; Camila Araujo de Lorenzo Barcia; Camila I. de Oliveira; Carolina Kymie Vasques Nonaka; Clarissa Araujo Gurgel; Eduardo Oyama; Icaro Strobel; Izabela Jesus; Laise de Moraes; Leonardo Paiva Farias; Luisa Pedrosa; Marilia Miranda Franco; Marina Cucco; Ricardo Khouri on behalf of the Fiocruz COVID-19 Genomic Surveillance Network; Tiago Graf |
| EPI_ISL_10607983, EPI_ISL_10608192, EPI_ISL_10823591, EPI_ISL_10823791 | Houston Methodist Hospital | Houston Methodist Hospital | Akanksha Batajoo; James J. Davis; James M. Musser; Jessica Cambric; Jimmy Gollihar; Jordan Pachuca; Kristina Reppond; Madison N. Shyer; Matthew Ojeda Saavedra; Nicole Kanellopoulos; Paul A. Christensen; Randall J. Olsen; Rashi M. Thakur; Regan Mangham; Richard Snehal; Robert Olson; Ryan Gadd; S. Wesley Long; Sindy Pena; Sinjini Gupta; Yuvanesh Vedaraju |
| EPI_ISL_9419339, EPI_ISL_9494928 | ICMR-National Institute of Virology - INSACOG | NIV Influenza | Dr. Varsha Potdar and NIC Team |
| EPI_ISL_12220146 | INSPI-Dirección_de_Idi | INSPI-Dirección_de_Idi | Andrés Carrazco-Montalvo; Ariana León; Clara Lucía Tello; David Guizado; Leandro Patiño |
| EPI_ISL_10826351 | IZSM | TIGEM | Antonio Grimaldi Patrizia Annunziata Francesco Panariello Claudia Tiberio Teresa Giuliano Michela Daniele Valentina Bouche Ilaria Cimmino Chiara Colantuono Lucio Di Filippo Anna Manfredi Marcello Salvi Antonio Limone Luigi Atripaldi Andrea Ballabio Davide Cacchiarelli |
| EPI_ISL_9112451 | Idaho Bureau of Laboratories | Idaho Bureau of Laboratories | Aimee Ceniseros; Christian Loera; Christopher Ball; Matthew Charles Burns; R. Beukelman; Robert L. Voermans |
| EPI_ISL_8886600 | Indira Gandhi Memorial Hospital | Indira Gandhi Memorial Hospital | D. Fathmath Nazla Rafeeq; Dr. Ibrahim Afzal; Mr. Ibrahim Nishan Ahmed; Ms. Aishath Shuhudha; Ms. Aminath Shazleena Abdul Rahman; Ms. Soafy Mohamed |
| EPI_ISL_12359245 | Innlandet Hospital Trust, Division Lillehammer, Department for Medical Microbiology | Norwegian Institute of Public Health, Department of Virology | Atiya R Ali; Debech Nadia; Engebretsen Serina Beate; Garcia Llorente Ignacio; Hilde Elshaug; Hilde Nordby Falkenhaug; Hilde Vollan; Jon Bråte; Kamilla Heddeland Instefjord; Karoline Bragstad; Kathrine Stene-Johansen; Line Victoria Moen; Marie Paulsen Madsen; Olav Hungnes; Pedersen Benedikte Nevjen; Rasmus Riis Kopperud |
| EPI_ISL_9087004, EPI_ISL_10204533, EPI_ISL_10204562, EPI_ISL_10552893, EPI_ISL_10985604, EPI_ISL_10985613, EPI_ISL_10985639, EPI_ISL_11073373 | | | |

| | | | |
|---|---|---|---|
| see above | Institute for Water Quality and Resource Management, Technical University Vienna | Bergthaler laboratory, CeMM Research Center for Molecular Medicine of the Austrian Academy of Sciences | Andreas Bergthaler; Anna Schedl; Bekir Erguner; Benedikt Agerer; Christoph Bock; Fabian Amman; Jan Laine; Lukas Endler; Martin Senekowitsch; Matthew Thornton; Michael Schuster; Michelle Chan; Petr Triska; Thomas Penz |
| EPI_ISL_10204530, EPI_ISL_11073242, EPI_ISL_11073289 | Institute of Legal Medicine, Medical University of Innsbruck | Bergthaler laboratory, CeMM Research Center for Molecular Medicine of the Austrian Academy of Sciences | Andreas Bergthaler; Anna Schedl; Bekir Erguner; Benedikt Agerer; Christoph Bock; Fabian Amman; Jan Laine; Lukas Endler; Martin Senekowitsch; Matthew Thornton; Michael Schuster; Michelle Chan; Petr Triska; Thomas Penz |
| EPI_ISL_9504693, EPI_ISL_9893350, EPI_ISL_9893629, EPI_ISL_9952817, EPI_ISL_9958034, EPI_ISL_11554269, EPI_ISL_11590727, EPI_ISL_11590865, EPI_ISL_11613412, EPI_ISL_11613419, EPI_ISL_12284836, EPI_ISL_12285171, EPI_ISL_12285177, EPI_ISL_13271851 | | | |
| see above | Institute of Microbiology and Immunology, Faculty of Medicine, University of Ljubljana | Institute of Microbiology and Immunology, Faculty of Medicine, University of Ljubljana | Alen Suljič; Andraž Celar; Domen Lazar; Doroteja Vlaj; Mario Poljak; Miša Korva; Patricija Pozvek; Samo Zakotnik; Tatjana Avšič – Županc; Tina Gabrovšek; Tina Živič; Tomaž Mark Zorec; Špela Pleh |
| EPI_ISL_12424913 | Instituto Rene Rachou / Fiocruz Minas | Instituto Rene Rachou / Fiocruz Minas | On behalf of the Fiocruz COVID-19 Genomic Surveillance Network |
| EPI_ISL_12754735 | Irrua Specialist Teaching Hospital (ISTH) | Africa Centre for Excellence for Genomics of Infectious Diseases (ACEGID), Redeemer's University | A.T.; Abechi; Ajogbasile; Akano; C.A.; C.T.; Eromon; F.V.; Folarin, O.; Happi; I.B.; J.N.; J.U.; John; K.O.; Kayode; Nosamiefan, I.; O.G.; Oguzie; Olawoye; Olumade; Oluniyi; P.E.; P.S.; T.J.; Ugwu; Uwanibe |
| EPI_ISL_12771189 | JKN PAHANG | iPROMISE, UiTM | Ariza Adnan; Fadzilah Mohd Nor; Lim Wai Feng; Mohd Asif Mohd Sukri; Mohd Nur Fakhruzzaman Noorizhab; Mohd Zaki Salleh; Sazzli Shahlan Kassim; Siti Farah Alwani Mohd Nawi; Siti Hamimah Sheikh Abdul Kadir; Teh Lay Kek; Wang Seok Mui |
| EPI_ISL_9306388 | Jerusalem Central Laboratories | Israel Central Virology laboratory | Danit Sofer; Efrat Dahan Bucris; Ella Mendelson; Evan Nachum; Hagar Morad; Julia Vainer; Maya Davidovich; Michal Mandelboim; Michal Zak; Miranda Geva; Neta Zuckerman; Or Zilbertzan; Oran Erster; Orna Mor; Rona Grossman |
| EPI_ISL_8589447 | Jessa | Jessa | Severine Berden et al. on behalf of the Jessa_cmdLab |
| EPI_ISL_13360911 | KU Leuven, Rega Institute, Clinical and Epidemiological Virology | KU Leuven, Rega Institute, Clinical and Epidemiological Virology | Anne-Sophie Logist; Bert Vanmechelen; Jens Verlinden; Levi Ysebaert; Piet Maes; Robbe Sinnesael; Tony Wawina-Bokalanga |
| EPI_ISL_11327789 | King Chulalongkorn Memorial Hospital | Thai Red Cross Emerging Infectious Diseases Clinical Center and Faculty of Medicine, Chulalongkorn University | Anthony R. Jones; Anutsara Jitsatja; Chanchanit Phanlop; Chonticha Klungthong; Gompol Suwanpimolkul; Khajohn Joonlasak; Leilani Paitoonpong; Nattakarn Thippamom; Opass Putcharoen; Pattama Torvorapanit; Piyawan Chinnawirotpisan; Sasiprapa Ninwattana; Stefan Fernandez; Supaporn Wacharapluesadee; Watsamon Jantarabenjakul; Wudtichai Manasatienkij |
| EPI_ISL_9143681, EPI_ISL_9143696, EPI_ISL_9143731 | Klinika za infektivne bolesti "Dr. Fran Mihaljević" | Hrvatski zavod za javno zdravstvo | Anita Jurić; Dragan Jurić; Irena Tabain; Ivana Ferenčak; Josipa Kuzle |
| EPI_ISL_9446261 | Klinikum Stuttgart | Robert Koch Institute | |
| EPI_ISL_8886623, EPI_ISL_8886624 | Kudahuvadhoo Atoll Hospital | Kudahuvadhoo Atoll Hospital | D. Fathmath Nazla Rafeeq; Dr. Ibrahim Afzal; Mr. Ibrahim Nishan Ahmed; Ms. Aishath Shuhudha; Ms. Aminath Shazleena Abdul Rahman; Ms. Soafy Mohamed |
| EPI_ISL_8886610 | Kulhudufushi Regional Hospital | Kulhudufushi Regional Hospital | D. Fathmath Nazla Rafeeq; Dr. Ibrahim Afzal; Mr. Ibrahim Nishan Ahmed; Ms. Aishath Shuhudha; Ms. Aminath Shazleena Abdul Rahman; Ms. Soafy Mohamed |
| EPI_ISL_11579864 | LABOffice | Centre Hospitalier Universitaire (CHU) Poitiers | Agnes Beby-Defaux; Jean Philippe Da Mota; Luc Deroche; Magali Garcia; Manon Prat; Nicolas Leveque |
| EPI_ISL_11133791 | LESP Baja California Sur | Instituto de Diagnostico y Referencia Epidemiologicos (INDRE) | Abril Rodriguez-Maldonado; Ana Cota-Magana; Ariadna Medina-Benitez; Claudia Wong-Arambula; Ernesto Ramirez-Gonzalez.; Fernando Gonzalez-Dominguez; Gisela Barrera-Badillo; Irma Lopez-Martinez; Joaquin Quiroz-Mercado; Lucia Hernandez-Rivas; Maribel Gonzalez-Villa; Natividad Cruz-Ortiz; Tatiana Nunez-Garcia; Vanessa Rivero-Arredondo |
| EPI_ISL_9749754 | LESP Chihuahua | Instituto de Diagnostico y Referencia Epidemiologicos (INDRE) | Abril Rodriguez-Maldonado; Ariadna Medina-Benitez; Claudia Wong-Arambula; Ernesto Ramirez-Gonzalez.; Fernando Gonzalez-Dominguez; Gisela Barrera-Badillo; Irma Lopez-Martinez; Joaquin Quiroz-Mercado; Lucia Hernandez-Rivas; Maribel Gonzalez-Villa; Natividad Cruz-Ortiz; Sergio Rangel-Guerrero; Tatiana Nunez-Garcia; Vanessa Rivero-Arredondo |
| EPI_ISL_9749680 | LESP Colima | Instituto de Diagnostico y Referencia Epidemiologicos (INDRE) | Abril Rodriguez-Maldonado; Ariadna Medina-Benitez; Claudia Wong-Arambula; Ernesto Ramirez-Gonzalez.; Fernando Gonzalez-Dominguez; Gisela Barrera-Badillo; Irma Lopez-Martinez; Joaquin Quiroz-Mercado; Lucia Hernandez-Rivas; Maribel Gonzalez-Villa; Natividad Cruz-Ortiz; Sergio Rangel-Guerrero; Tatiana Nunez-Garcia; Vanessa Rivero-Arredondo |
| EPI_ISL_9749779 | LESP Guanajuato | Instituto de Diagnostico y Referencia Epidemiologicos (INDRE) | Abril Rodriguez-Maldonado; Ariadna Medina-Benitez; Claudia Wong-Arambula; Ernesto Ramirez-Gonzalez.; Fernando Gonzalez-Dominguez; Gisela Barrera-Badillo; Irma Lopez-Martinez; Joaquin Quiroz-Mercado; Lucia Hernandez-Rivas; Maribel Gonzalez-Villa; Natividad Cruz-Ortiz; Sergio Rangel-Guerrero; Tatiana Nunez-Garcia; Vanessa Rivero-Arredondo |
| EPI_ISL_9749556, EPI_ISL_9749557 | LESP Jalisco | Instituto de Diagnostico y Referencia Epidemiologicos (INDRE) | Abril Rodriguez-Maldonado; Ariadna Medina-Benitez; Claudia Wong-Arambula; Ernesto Ramirez-Gonzalez.; Fernando Gonzalez-Dominguez; Gisela Barrera-Badillo; Irma Lopez-Martinez; Joaquin Quiroz-Mercado; Lucia Hernandez-Rivas; Maribel Gonzalez-Villa; Natividad Cruz-Ortiz; Sergio Rangel-Guerrero; Tatiana Nunez-Garcia; Vanessa Rivero-Arredondo |
| EPI_ISL_9749573 | LESP Mexico City | Instituto de Diagnostico y Referencia Epidemiologicos (INDRE) | Abril Rodriguez-Maldonado; Ariadna Medina-Benitez; Claudia Wong-Arambula; Ernesto Ramirez-Gonzalez.; Fernando Gonzalez-Dominguez; Gisela Barrera-Badillo; Irma Lopez-Martinez; Joaquin Quiroz-Mercado; Lucia Hernandez-Rivas; Maribel Gonzalez-Villa; Natividad Cruz-Ortiz; Sergio Rangel-Guerrero; Tatiana Nunez-Garcia; Vanessa Rivero-Arredondo |
| EPI_ISL_9749656 | LESP Sonora | Instituto de Diagnostico y Referencia Epidemiologicos (INDRE) | Abril Rodriguez-Maldonado; Ariadna Medina-Benitez; Claudia Wong-Arambula; Ernesto Ramirez-Gonzalez.; Fernando Gonzalez-Dominguez; Gisela Barrera-Badillo; Irma Lopez-Martinez; Joaquin Quiroz-Mercado; Lucia Hernandez-Rivas; Maribel Gonzalez-Villa; Natividad Cruz-Ortiz; Sergio Rangel-Guerrero; Tatiana Nunez-Garcia; Vanessa Rivero-Arredondo |
| EPI_ISL_9749697 | LESP Tabasco | Instituto de Diagnostico y Referencia Epidemiologicos (INDRE) | Abril Rodriguez-Maldonado; Ariadna Medina-Benitez; Claudia Wong-Arambula; Ernesto Ramirez-Gonzalez.; Fernando Gonzalez-Dominguez; Gisela Barrera-Badillo; Irma Lopez-Martinez; Joaquin Quiroz-Mercado; Lucia Hernandez-Rivas; Maribel Gonzalez-Villa; Natividad Cruz-Ortiz; Sergio Rangel-Guerrero; Tatiana Nunez-Garcia; Vanessa Rivero-Arredondo |
| EPI_ISL_11133813, EPI_ISL_11133845 | LESP Veracruz | Instituto de Diagnostico y Referencia Epidemiologicos (INDRE) | Abril Rodriguez-Maldonado; Ana Cota-Magana; Ariadna Medina-Benitez; Claudia Wong-Arambula; Ernesto Ramirez-Gonzalez.; Fernando Gonzalez-Dominguez; Gisela Barrera-Badillo; Irma Lopez-Martinez; Joaquin Quiroz-Mercado; Lucia Hernandez-Rivas; Maribel Gonzalez-Villa; Natividad Cruz-Ortiz; Tatiana Nunez-Garcia; Vanessa Rivero-Arredondo |
| EPI_ISL_11133876 | LESP Yucatan | Instituto de Diagnostico y Referencia Epidemiologicos (INDRE) | Abril Rodriguez-Maldonado; Ana Cota-Magana; Ariadna Medina-Benitez; Claudia Wong-Arambula; Ernesto Ramirez-Gonzalez.; Fernando Gonzalez-Dominguez; Gisela Barrera-Badillo; Irma Lopez-Martinez; Joaquin Quiroz-Mercado; Lucia Hernandez-Rivas; Maribel Gonzalez-Villa; Natividad Cruz-Ortiz; Tatiana Nunez-Garcia; Vanessa Rivero-Arredondo |
| EPI_ISL_10296073 | LKO | Jessa | Severine Berden et al. on behalf of the Jessa_cmdLab |
| EPI_ISL_9058428 | Lab. Microbiologia e Virologia Cotugno A.O. dei Colli | TIGEM | Antonio Grimaldi Patrizia Annunziata Francesco Panariello Claudia Tiberio Teresa Giuliano Valentina Bouche Chiara Colantuono Michela Daniele Lucio Di Filippo Anna Manfredi Marcello Salvi Antonio Limone Luigi Atripaldi Andrea Ballabio Davide Cacchiarelli |
| EPI_ISL_12521761, EPI_ISL_13014643 | Labor ZOTZ\|KLIMAS; MVZ Düsseldorf-Centrum | Robert Koch Institute | |
| EPI_ISL_11293098 | Laboratoire National de Santé Publique – LNSP | Genomics and Proteomics Departament, Gorgas Memorial Institute For Health Studies | Alexander A Martinez; Ambar Moreno; Claudia Gonzalez V; Ito Journel; Jaques Boncy; Jessica Gondola; Oris Chavarria; Patrick Delly |
| EPI_ISL_11110790 | Laboratorio Central de Epidemiologia (LCE) | Unidad de Genomica Avanzada | Alejandro Sanchez-Flores; Alfredo Herrera-Estrella; Alicia Ocana-Mondragon; Angel Gustavo Salas-Lais; Bernardo Martinez-Miguel; Blanca Taboada; Brenda Irasema Maldonado-Meza; Carla Ivon Herrera-Najera; Carlos F. Arias; Celia Boukadida; Clara Esperanza Santacruz-Tinoco; Concepcion Grajales-Muniz; Consorcio Mexicano de Vigilancia Genomica (CoViGen-Mex). Authors (in alphabetical order): Julio Elias Alvarado-Yaah; Fernando Fontove-Herrera; Francisco Pulido; Gloria Elena Espinoza-Ayala; Gloria Maria Molina-Salinas; Gloria Vazquez; Hector Esteban Paz-Juarez; Hector Montoya-Fuentes; Helen Haydee Fernanda Ramirez-Plascencia; Jorge Ivan Salinal-Nevarez; Jose Antonio Enciso-Moreno; Jose Esteban Munoz-Medina; Jose de Jesus Nunez-Contreras; Juan Bautista Chale-Dzul; Luis Alberto Ochoa-Carrera; Margarita Matias-Florentino; Maria Guadalupe Santiago-Mauricio; Maria Guadalupe de Jesus Mireles-Rivera; Nelly Selem-Mojica; Pavel Isa; Ricardo Grande; Santiago Avila-Rios; Victor Hugo Borja-Aburto |
| EPI_ISL_9747755, EPI_ISL_10706152 | Laboratorio Municipal de Biologia Molecular | Instituto Rene Rachou / Fiocruz Minas | Andre Menezes; Anna Salim; Caroline Rocha; Eneida Oliveira; Flavio Araujo; Gabriel Fernandes; On behalf of COVID-19 Fiocruz genomic network; Pedro Alves; Rubens do Monte Neto; Thais Silva |
| EPI_ISL_9801678 | Laboratorio Nacional de Salud (LNS), Guatemala | Laboratorio Nacional de Salud (LNS), Guatemala | García G; Mendoza L |
| EPI_ISL_11629415, EPI_ISL_11629427 | Laboratorio Nacional de Salud, Ministerio de Salud Publica y Asistencia Social | Genomics and Proteomics Departament, Gorgas Memorial Institute For Health Studies | Alexander A Martinez; Ambar Moreno; Claudia Estrada; Claudia Gonzalez V; César Roberto Conde Pereira; Jessica Gondola; Leyda Abrego; Marlene Castillo; Melissa Gaitan; Oris Chavarria |
| EPI_ISL_12422403 | Laboratorio de Virologia del Hospital de Niños Dr Ricardo Gutierrez | Área de Secuenciación del Laboratorio de Virología del Hospital de Niños Dr. Ricardo Gutierrez on behalf of 'Proyecto Argentino Interinstitucional de genomica de SARS-CoV-2' (PAIS Consortium) | Acuña; Alejandra Musto; Alicia Mistchenko; Carla Medina; Cristian Díaz; Cristian Turchiaro; D; Erica Grandis; Erica Luczak; Estela Chascón; Goya; Guillermo Thomas; Isabel Desimone; Jorgelina Caruso; Julián Cipelli; Karina Zacarías; LE; Lorena Serrano; Lusso; MI; MS; Mariana Campal; Mariángeles Barreda Fink; María Cristina Alvarez López; María Elina Acevedo; María Emilia Villegas; Nabaes Jodar; Natale; Natalia Labarta; Omar Grossi; Oscar Jacquez; Oscar Luna; Raquel Barquez; Rubén Pelagamos; S; Sofía Alexay; Streitenberger Cintia; Valinotto; Viegas, M. |
| EPI_ISL_12422397 | Laboratorio del Hospital Interzonal General de Agudos "Evita" | Área de Secuenciación del Laboratorio de Virología del Hospital de Niños Dr. Ricardo Gutierrez on behalf of 'Proyecto Argentino Interinstitucional de genomica de SARS-CoV-2' (PAIS Consortium) | Acuña; Alejandra Musto; D; Erica Luczak; Goya; Isabel Desimone; LE; Lorena Serrano; Lusso; MI; MS; Nabaes Jodar; Natale; Omar Grossi; Rubén Pelagamos; S; Valinotto; Viegas, M. |
| EPI_ISL_12422410 | Laboratorio del Hospital Zonal Especializado Materno Infantil "Argentina Diego" | Área de Secuenciación del Laboratorio de Virología del Hospital de Niños Dr. Ricardo Gutierrez on behalf of 'Proyecto Argentino Interinstitucional de genomica de SARS-CoV-2' (PAIS | Acuña; D; Farinella Bagnozzi y Santiago Hernán Gauna; Goya; Guillermina; LE; Lusso; MI; MS; María Belén Arpaia; Nabaes Jodar; Natale; S; Valinotto; Viegas, M. |

| | | | |
|---|---|---|---|
| | | Consortium) | |
| EPI_ISL_10775606 | Laboratory of Genomics and Bioinformatics, Comenius University Science Park | Laboratory of Genomics and Bioinformatics, Comenius University Science Park | Anna Kaliňáková; Barbora Kotvasová; Diana Rusňáková; Jakub Styk; Jaroslav Budiš; Lucia Ševčíková; Michaela Jakubková Forgáčová; Miroslav Böhmer; Nikola Lipková; Pavol Mišenko; Silvia Bokorová; Tatiana Sedláčková; Terézia Vrabľová; Tomáš Szemes |
| EPI_ISL_13653682 | Laboratory of Respiratory Viruses and Measles, Oswaldo Cruz Institute, FIOCRUZ | Laboratory of Respiratory Viruses and Measles, Oswaldo Cruz Institute, FIOCRUZ | Alice Sampaio Rocha; Bruna Mendonça da Silva; Elisa Cavalcante Pereira; Fernando Motta; Ighor Arantes; Jéssica Graça Macedo de Carvalho; Larissa Macedo Pinto; Luciana Appolinario; Marilda Siqueira on behalf of the Fiocruz COVID-19 Genomic Surveillance Network; Paola Resende; Victor Guimaraes |
| EPI_ISL_12708634, EPI_ISL_12708643 | Laboratory of clinical bacteriology, Russian Children Clinical Hospital, Pirogov Medical University, Moscow, Russian Federation | Center for Precision Genome Editing and Genetic Technologies for Biomedicine, Pirogov Medical University, Moscow, Russian Federation | Denis Rebrikov; Dmitry Korostin; Fedorova Natalia; Iuliia Vasiliadis; Martynenkova Aliya; Natalia Ponikarovskaya; Vera Belova |
| EPI_ISL_11750291 | Lembang General Hospital | West Java Health Laboratory; School of Life Sciences and Technology, Institut Teknologi Bandung | Azzania Fibriani; Cut Nur Cinthia Alamanda; Ema Rahmawati; Hadiana; Karimatu Khoirunnisa; Miftahul Faridl; Rifky Waluyajati Rachman; Rini Robiani; Ryan Bayusantika Ristandi |
| EPI_ISL_10693065, EPI_ISL_10693140, EPI_ISL_11325519, EPI_ISL_11561103 | Lifebrain Covid Labor GmbH | Lifebrain Covid Labor GmbH | Abhishek Mitra; Alexandra Wagner; Anna Edermayr; Felix Valentin Spiegel; Filip Sima; Florian Scharhauser; Hannes Hagen; Kristina Bavrka Kolenc; Lucia Castello; So Jung Han |
| EPI_ISL_8699721, EPI_ISL_8704326, EPI_ISL_8743347, EPI_ISL_9901011, EPI_ISL_10201162, EPI_ISL_10202493, EPI_ISL_10428152, EPI_ISL_10525291, EPI_ISL_10531340, EPI_ISL_10856923, EPI_ISL_11044530, EPI_ISL_11088335, EPI_ISL_11652657 | | | |
| see above | Lighthouse Lab in Glasgow | Wellcome Sanger Institute for the COVID-19 Genomics UK (COG-UK) Consortium | Anna Dominiczak and Alex Alderton; Carol Clugston; Cordelia Langford; David Gray; David K. Jackson; Dominic Kwiatkowski; Ewan Harrison; Harper VanSteenhouse; Ian Johnston; Jeffrey Barrett; John Sillitoe on behalf of the Wellcome Sanger Institute COVID-19 Surveillance Team; Roberto Amato; Sonia Goncalves; Yumi Kasai |
| EPI_ISL_9315229, EPI_ISL_10421352 | Lighthouse Lab in Milton Keynes | Wellcome Sanger Institute for the COVID-19 Genomics UK (COG-UK) Consortium | Cordelia Langford; David K. Jackson; Dominic Kwiatkowski; Ewan Harrison; Ian Johnston; Jeffrey Barrett; John Sillitoe on behalf of the Wellcome Sanger Institute COVID-19 Surveillance Team; Roberto Amato; Sonia Goncalves; The Lighthouse Lab in Milton Keynes and Alex Alderton |
| EPI_ISL_10062804 | Mae Sot General Hospital | Thailand MOPH - U.S. CDC Collaboration (TUC) | Beth Skaggs; Piroon Jenjaroenpun; Pongpun Sawatwong; Thidathip Wongsurawat |
| EPI_ISL_9506904, EPI_ISL_9957098, EPI_ISL_9957184 | Medical Microbiology Unit, Department for Laboratory Medicine, Drammen Hospital, Vestre Viken Health Trust | Norwegian Institute of Public Health, Department of Virology | Atiya R Ali; Debech Nadia; Engebretsen Serina Beate; Garcia Llorente Ignacio; Hilde Elshaug; Hilde Nordby Falkenhaug; Hilde Vollan; Jon Bråte; Kamilla Heddeland Instefjord; Karoline Bragstad; Kathrine Stene-Johansen; Line Victoria Moen; Marie Paulsen Madsen; Olav Hungnes; Pedersen Benedikte Nevjen; Rasmus Riis Kopperud |
| EPI_ISL_10070736 | Medical Virology and BSL3 Laboratory | Centre de Séquençage Génomique | Abbad Anas; Amalou Ghita; Anga Latifa; Barakat Abdelhamid; Bouzidi Aymane; Charoute Hicham; Chgouri Fatima; Dersi Noureddine; EL Hamouchi Adil; El Oualid Abdelmjid; Faouzi Abdellah; Harmak Houda; Maaroufi Abderrahmane; Nadifiyine Saloua; Nourlil Jalal; Omondi Francis Carey; Somda Soro Georgina Charlene; Zemmouri Faouzia; Zouheir Yassine |
| EPI_ISL_12027090 | Medizinisch-Diagnostisches Labor Kempten allgäulab | Robert Koch Institute | |
| EPI_ISL_10079374 | Michigan Department of Health and Human Services, Bureau of Laboratories | Michigan Department of Health and Human Services, Bureau of Laboratories | Blankenship HM; Riner D; Soehnlen MK |
| EPI_ISL_9848150, EPI_ISL_10844464 | Microbiology Department, Laboratori Clínic Metropolitana Nord. Hospital Universitari Germans Trias i Pujol | Can Ruti SARS-CoV-2 Sequencing Hub (HUGTiP/IrsiCaixa/IGTP) | Alexia París; Ana Blanco; Andreu Coello; Antoni E Bordoy; Bonaventura Clotet; David Panisello; Francesc Catala-Moll; Gemma Clara; Ignacio Blanco; Laia Soler; Lauro Sumoy; Marc Noguera-Julian; Mercedes Guerrero; Montserrat Giménez; Pere-Joan Cardona; Pilar Armengol; Roger Paredes; Sara González; Verónica Saludes; and Elisa Martró on behalf of the Can Ruti SARS-CoV-2 Sequencing Hub |
| EPI_ISL_11248019 | Microbiology Department. Complexo Hospitalario Universitario de Vigo | Microbiology Department. Complexo Hospitalario Universitario de Vigo | Alvarez M; Cabrera JJ; Carballo R; Cortizo S; Davina C; Martinez L; Mediero G; Pena I; Perez S; Potel C; Regueiro B; Rey S; Vasallo FJ; del-Campo V |
| EPI_ISL_8843407, EPI_ISL_8929501, EPI_ISL_8929502, EPI_ISL_10443635, EPI_ISL_10720896, EPI_ISL_10720897, EPI_ISL_10720905, EPI_ISL_10721105, EPI_ISL_10721173, EPI_ISL_10721212, EPI_ISL_10721232, EPI_ISL_10858408, EPI_ISL_10858415, EPI_ISL_10858472, EPI_ISL_10858530, EPI_ISL_11507575, EPI_ISL_13113844 | | | |
| see above | Ministry of Health Turkey | Ministry of Health Turkey | Fatma Bayrakdar; Gulay Korukluoglu; Gülay Korukluoğlu; Gültekin Ünal; Suleyman Yalcin; Süleyman Yalcin; Yasemin Cosgun; Yasemin Coşgun |
| EPI_ISL_10008460, EPI_ISL_10008485 | Ministry of Public Health / Hamad Medical Corporation | Biomedical Research Center (BRC), Qatar University / Qatar Genome Project (QGP) | Asmaa A. Al-Thani. MOPH and HMC: Abdullatif Al-Khal; BRC: Heba A. Al-Khatib; Chadi Saad; Dana Al-Batsh; Dina Elgakhlab. QGP: Fatima H. Al-Kuwari; Einas A. E. Al-Kuwari; Fatiha M. Benslimane; Fatma Hassan Ali; Hadeel T. Mohammed; Hadi M. Yassine; Hamad E. Al-Romaihi; Hamda Alromaihi; Maria K. Smatti; Mashael A. Al-Bader; Mohammed Al-Thani; Muna A. S. Al-Maslamani; Ola Al-Jamal; Peter V. Coyle; Reham A. El-Kahlout. QBB: Tasneem Al-Hamad; Roberto Bertollini; Salih Al-Marri; Swapna Thomas |
| EPI_ISL_12949403, EPI_ISL_12949519, EPI_ISL_12949521, EPI_ISL_12949546 | Molecular Genetic Monitoring Group | Molecular Genetic Monitoring Group | Anna S. Gladkikh; Areg A.Totolian; Ekaterina O. Klyuchnikova; Valerya A. Sbarzaglia; Vladimir G. Dedkov |
| EPI_ISL_9457547, EPI_ISL_9860069 | Nastavni zavod za javno zdravstvo Splitsko- Dalmatinske županije | Hrvatski zavod za javno zdravstvo | Anita Jurić; Dragan Jurić; Irena Tabain; Ivana Ferenčak; Josipa Kuzle |
| EPI_ISL_10854133, EPI_ISL_13180065 | National Center of Infectious and Parasitic Diseases | National Center of Infectious and Parasitic Diseases | Alexiev et al |
| EPI_ISL_10083919, EPI_ISL_10083923, EPI_ISL_10083927, EPI_ISL_10083929, EPI_ISL_10083930, EPI_ISL_10083934 | National Influenza Center | National Influenza Center | A Nejati; Adel Abedi; Adel Abedi and T Mokhtari Azad; J Yavarian; K Sadeghi; NZ Shafiei Jandaghi; Sevrin Zadheidar; Sevrin Zadheidar and T Mokhtari Azad; V Salimi |
| EPI_ISL_9417774, EPI_ISL_9823338 | National Influenza Center, Virology Department | National Influenza Center | A Nejati; Adel Abedi and T Mokhtari Azad; J Yavarian; K Sadeghi; NZ Shafiei Jandaghi; Sevrin Zadheidar; V Salimi |
| EPI_ISL_9967402 | National Institute of Infectious Diseases-Prof. Dr. Matei Bals Molecular Diagnostics Laboratory | National Institute of Infectious Diseases-Prof. Dr. Matei Bals Molecular Diagnostics Laboratory | Corina Casangiu; Dan Otelea; Leontina Banica; Marius Surleac; Ovidiu Vlaicu; Petre Milu; Robert Hohan; Simona Paraschiv |
| EPI_ISL_11982371, EPI_ISL_11982379, EPI_ISL_11982383, EPI_ISL_11982390, EPI_ISL_11982393, EPI_ISL_11982434, EPI_ISL_11982438, EPI_ISL_11982462, EPI_ISL_11982489, EPI_ISL_11982516, EPI_ISL_11982542, EPI_ISL_11982584 | | | |
| see above | National Virus Reference Laboratory | National Virus Reference Laboratory | Charlene Bennett; Cillian F De Gascun; Gabriel Gonzalez; Jonathan Dean; Michael Carr; Zoe Yandle |
| EPI_ISL_12631494 | NordLab Kokkola | Expert Microbiology, National Institute for Health and Welfare | Aino Palva; Carita Savolainen-Kopra; Emma Saarinen; Erika Lindh; Haider al-Hello; Jani Halkilahti; Kirsi Liitsola; Niina Ikonen; Olli Vapalahti; Phuoc Truong; Päivi Laurila; Ravi Kant; Sari Hannula; Soile Blomqvist; Suvi Kyttänen; Teemu Smura; Tiina Hannunen |
| EPI_ISL_9695869, EPI_ISL_9815626, EPI_ISL_10261068, EPI_ISL_10264569, EPI_ISL_10264638, EPI_ISL_10583127, EPI_ISL_10863341, EPI_ISL_11006863, EPI_ISL_11231939, EPI_ISL_11232756, EPI_ISL_11320379, EPI_ISL_11751242 | | | |
| see above | Originating lab: Wales Specialist Virology Centre Sequencing lab: Pathogen Genomics Unit | Public Health Wales Microbiology Cardiff Wales Specialist Virology Centre | Alec Birchley; Alexander Adams; Amy Gaskin; Angela Marchbank; Bree Gatica-Wilcox; Catherine Moore; Jason Coombes; Joanne Watkins; Joel Southgate; Johnathan Evans; Laura Gifford; Lauren Gilbert; Lee Graham; Malorie Perry; Matthew Bull; Nicole Pacchiarini; Sally Corden; Sara Kumziene-Summerhayes; Sara Rey; Sarah Taylor; Simon Cottrell; Sophie Jones; Tom Connor |
| EPI_ISL_10190124, EPI_ISL_10826158, EPI_ISL_11023944, EPI_ISL_11055381, EPI_ISL_13030492 | Ospedale Santa Maria delle Grazie | TIGEM | Antonio Grimaldi Patrizia Annunziata Francesco Panariello Claudia Tiberio Teresa Giuliano Michela Daniele Valentina Bouche Ilaria Cimmino Chiara Colantuono Lucio Di Filippo Anna Manfredi Marcello Salvi Antonio Limone Luigi Atripaldi Andrea Ballabio Davide Cacchiarelli; Antonio Grimaldi Patrizia Annunziata Francesco Panariello Claudia Tiberio Teresa Giuliano Tonya Fusco Valentina Bouche Ilaria Cimmino Chiara Colantuono Lucio Di Filippo Anna Manfredi Marcello Salvi Antonio Limone Luigi Atripaldi Andrea Ballabio Davide Cacchiarelli |
| EPI_ISL_11939211 | PKD TIMUR LAUT | iPROMISE, UiTM | Ariza Adnan; Fadzilah Mohd Nor; Lim Wai Feng; Mohd Asif Mohd Sukri; Mohd Nur Fakhruzzaman Noorizhab; Mohd Zaki Salleh; Sazzli Shahlan Kassim; Siti Farah Alwani Mohd Nawi; Siti Hamimah Sheikh Abdul Kadir; Teh Lay Kek; Wang Seok Mui |
| EPI_ISL_12771264 | PKDSPT | iPROMISE, UiTM | Ariza Adnan; Fadzilah Mohd Nor; Lim Wai Feng; Mohd Asif Mohd Sukri; Mohd Nur Fakhruzzaman Noorizhab; Mohd Zaki Salleh; Sazzli Shahlan Kassim; Siti Farah Alwani Mohd Nawi; Siti Hamimah Sheikh Abdul Kadir; Teh Lay Kek; Wang Seok Mui |
| EPI_ISL_9388271 | PPHL Bheri | National Public Health Laboratory | National Public Health Laboratory Team |
| EPI_ISL_10101148 | PRVKP FKUI | National Quality Control Laboratory of Drug and Food | Dillin N; Hana Apsari Pawestri; M. Erdiansyah; Sri Utaminingsih |
| EPI_ISL_9854383, EPI_ISL_10348671, EPI_ISL_10675280, EPI_ISL_10675284, EPI_ISL_10675322, EPI_ISL_10818821, EPI_ISL_10922766, EPI_ISL_11000733, EPI_ISL_12626318 | | | |
| see above | PathWest Laboratory Medicine WA | PathWest Laboratory Medicine WA Microbial Surveillance Unit | PathWest Laboratory Medicine WA Microbial Surveillance Unit |
| EPI_ISL_11332334 | Pathology Queensland and | Public Health Virology - Forensic | Chenwei Wang on behalf of Q-PHIRE Genomics |

| | | | |
|---|---|---|---|
| | Forensic Scientific Services | and Scientific Services | |
| EPI_ISL_10511960 | Piyavate Hospital | National Institute of Health, Department of Medical Sciences, Ministry of Public Health, Thailand | Archawin Rojanawiwat; Natchaya Khiadsang; Nuttida Thongpramul; Pakorn Piromtong; Pilailuk Okada; Sirikanda Wimol; Siripaporn Phuygun; Sunthareeya Waicharoen; Suratchana Mitrat; Thanutsapa Thanadachakul |
| EPI_ISL_12422407 | Plataforma de Servicios Biotecnólogicos: UTTIPP/PSB , Universidad Nacional de Quilmes. | Área de Secuenciación del Laboratorio de Virología del Hospital de Niños Dr. Ricardo Gutierrez on behalf of 'Proyecto Argentino Interinstitucional de genomica de SARS-CoV-2' (PAIS Consortium) | Acuña; Alejandra Zinni; Carla Capobianco; D; Georgina Cardama; Goya; Gustavo Bada; Hernán Farina; Humberto Lamdan; LE; Lusso; MI; MS; Marcelo Mandile; Nabaes Jodar; Natale; Norailys Lorenzo; S; Sandra Goñi; Valinotto; Viegas, M. |
| EPI_ISL_12573083 | Plataforma de Vigilancia Molecular (PVM) - FIOCRUZ/BA | Plataforma de Vigilancia Molecular (PVM) - FIOCRUZ/BA | Bruno Bezerril Andrade; Camila I. de Oliveira; Clarissa Araujo Gurgel; Eduardo Oyama; Icaro Strobel; Izabela Jesus; Laise de Moraes; Leonardo Paiva Farias; Luisa Pedrosa; Marina Cucco; Ricardo Khouri on behalf of the Fiocruz COVID-19 Genomic Surveillance Network; Tiago Graf |
| EPI_ISL_8928596, EPI_ISL_8931071 | Polski Bank Komórek Macierzystych | WSSE w Warszawie | Dorota Wagrocka - Roczniak |
| EPI_ISL_9353081 | Public Health Authority of the Slovak Republic | Regional Authority of Public Health Banska Bystrica | Alžbeta Pristyáková; Diana Rusňáková; Lucia Maďarová; Michaela Mancoš; Miroslav Böhmer; Pavol Mišenko; Soňa Feiková; Terézia Tomajková; Tomáš Szemes |
| EPI_ISL_9561478 | Quadram Institute Bioscience | COVID-19 Genomics UK (COG-UK) Consortium | Alexander J Trotter; Ana Martínez Gascueña; Andrew J. Page; Darcie Burgess; Dave J. Baker; Leonardo de Oliveira Martins; Lizzie Meadows; Martin Lott; Michaela Matthews; Nabil-Fareed Alikhan; Rebecca Sutcliffe; Rhiannon Evans; Thanh Le-Viet |
| EPI_ISL_11449558, EPI_ISL_11449559 | Queen Sirikit National Institute of Child Health | National Institute of Health, Department of Medical Sciences, Ministry of Public Health, Thailand | Archawin Rojanawiwat; Natchaya Khiadsang; Nuttida Thongpramul; Pakorn Piromtong; Pilailuk Okada; Sirikanda Wimol; Siripaporn Phuygun; Sunthareeya Waicharoen; Suratchana Mitrat; Thanutsapa Thanadachakul |
| EPI_ISL_9371494, EPI_ISL_9371898 | REUNILAB | UMR PIMIT | Dr Camille Lebarbenchon; Dr David A Wilkinson; Dr Jonathan Turpin; Dr Patrick Mavingui; Magali Turpin; Marie-Alice Simbi |
| EPI_ISL_9232416 | Rainbow Hospital | CSIR-Centre for Cellular and Molecular Biology | Amareshwar Vodapalli; Ara Sreenivas; Archana Bharadwaj Siva; B Himasri; Divya Tej Sowpati; Jandhyala Sai Krishna; Kalyan Ram Uppaluri; Karthik Bharadwaj Tallapaka; Lamuk Zaveri; Malini Nemalikanti; Priya Nurkurthy; Rakesh K Mishra; Shreekant Verma; Smita Juvvadi; Sreelekshmi MS; Sricharan Devineni; Sumedha Avadhanula; Surabhi Srivastava; Tulasi Nagabandi; Valli Nagalakshmi Undamatla; Vidhyadhari Methuku |
| EPI_ISL_10987670 | Regional Hospital Kladno | National Institute of Public Health | Alexander Nagy; Helena Jirincova; Jan Moskalyk; Jaromira Vecerova; Timotej Suri |
| EPI_ISL_9588925, EPI_ISL_9686940, EPI_ISL_11155050, EPI_ISL_11655400, EPI_ISL_11809578 | Respiratory Virus Unit, Microbiology Services Colindale, Public Health England | COVID-19 Genomics UK (COG-UK) Consortium | PHE Covid Sequencing Team |
| EPI_ISL_11607503 | Rosalind Franklin Laboratory | Wellcome Sanger Institute for the COVID-19 Genomics UK (COG-UK) Consortium | Cordelia Langford; David K. Jackson; Dominic Kwiatkowski; Donald Fraser; Ewan Harrison; Ian Johnston; Jeffrey Barrett; John Sillitoe on behalf of the Wellcome Sanger Institute COVID-19 Surveillance Team; Rob Howes; Roberto Amato; Sonia Goncalves; Suki Lee; The Rosalind Franklin Laboratory and Alex Alderton |
| EPI_ISL_10856693, EPI_ISL_10856789, EPI_ISL_10856799 | SARS-CoV-2 Sequencing Castilla y Leon-Spain Consortium | SARS-CoV-2 Sequencing Castilla y Leon-Spain Consortium | Antonio Orduña-Domingo; Carlos Fuster Foz; Carmen Aldea-Mansilla; Carmen Gimeno Crespo; David Abad; Gabriel March Rosello; Gregoria Megías Lobón; Jose María Eiros Bouza; M. Isabel Fernandez-Natal; Marta Dominguez-Gil; Marta Hernandez; María Antonia García Castro; Mª Fe Brezmes-Valdivieso; Noelia Arenal Andrés; Silvia Rojo; Sonsoles Garcinuño Pérez |
| EPI_ISL_8920071, EPI_ISL_8920389, EPI_ISL_9303534, EPI_ISL_10495551 | SARS-CoV-2 testing team, National Institute of Infectious Diseases | Pathogen Genomics Center, National Institute of Infectious Diseases | Hazuka Y Furihata; Kentaro Itokawa; Makoto Kuroda; Masanori Hashino; Masumichi Saito; Naomi Nojiri; Nozomu Hanaoka; Rina Tanaka; Tsuguto Fujimoto; Tsuyoshi Sekizuka |
| EPI_ISL_9850777 | SEEBMO | Instituto Nacional de Saude (INSA) | Borges et al |
| EPI_ISL_11883434 | SMS MEDICAL COLLEGE,JAIPUR | NIV Influenza | Bharti Malhotra; Dinesh parsoya; Farah Deeba; Himanshu sharma; Neha Bhomia; Nita pal; Nivedita Gupta; Pragya D Yadav; Pratibha Sharma; Sudhir Bhandari; Swati Gautam; Varsha Potdar |
| EPI_ISL_9301654 | SOMDEJPRABOROMRACHINEENART NATAWEE HOSPITAL. | Prince of Songkhla University | Anusara Wongkotsila; Chanon Kongkamon; Pongsakorn Choochuen; Samonrapat Surasombatpattana; Sarunyou Chusri; Surakameth Mahasirimongkol; Surasak Sangkhathat; Thammasin Ingviya; Thanit Sila; Virasakdi Chongsuvivatwong; Wison Laochareonsuk |
| EPI_ISL_10271164 | SPZOZ K-KOŻLE | Wojewódzka Stacja Sanitarno-Epidemiologiczna w Katowicach | Adrian Miara; Agata Chajda; Beata Rozwadowska; Elżbieta Bartkowiak; Marta Albertyńska |
| EPI_ISL_10918897, EPI_ISL_11656659 | SYNLAB MVZ Weiden | Robert Koch Institute | |
| EPI_ISL_12631592 | SYNLAB Suomi | Expert Microbiology, National Institute for Health and Welfare | Aino Palva; Carita Savolainen-Kopra; Emma Saarinen; Erika Lindh; Haider al-Hello; Jani Halkilahti; Kirsi Liitsola; Niina Ikonen; Olli Vapalahti; Phuoc Truong; Päivi Laurila; Ravi Kant; Sari Hannula; Soile Blomqvist; Suvi Kyttänen; Teemu Smura; Tiina Hannunen |
| EPI_ISL_10932297, EPI_ISL_10932307, EPI_ISL_11503037 | Saratov Regional Clinical Hospital | WHO National Influenza Centre Russian Federation | Andrey Komissarov; Artem Fadeev; Daria Danilenko; Dmitry Lioznov; Elena Nabieva; Georgii Bazykin; Kirill Varchenko; Ksenia Safina; Kseniya Komissarova; Maria Pisareva; Mikhail Bakaev; Nikita Yolshin; Oula Mansour; Tamila Musaeva; Veronika Eder |
| EPI_ISL_11813240 | SataDiag | Expert Microbiology, National Institute for Health and Welfare | Aino Palva; Carita Savolainen-Kopra; Emma Saarinen; Erika Lindh; Haider al-Hello; Jani Halkilahti; Kirsi Liitsola; Niina Ikonen; Olli Vapalahti; Phuoc Truong; Päivi Laurila; Ravi Kant; Sari Hannula; Soile Blomqvist; Suvi Kyttänen; Teemu Smura; Tiina Hannunen |
| EPI_ISL_12972907 | Shahrekord University of Medical Sciences | National Influenza Center | A Nejati; Adel Abedi; J Yavarian; K Sadeghi; NZ Shafiei Jandaghi; Sevrin Zadheidar and T Mokhtari Azad; V Salimi |
| EPI_ISL_13234656 | Shizuoka Institute of Environment and Hygiene | Japan COVID-19 Open Data Consortium | Atsushi Toyoda; Hiromi Nagaoka; Hiroshi Mori; Ituro Inoue; Katsuyuki Ishigami; Ken Kurokawa; Masanori Arita; Satomi Asano; Shigeki Mitsunaga; Takatomo Fujisawa; Yasuhiro Tanizawa; Yono Arita |
| EPI_ISL_9301611, EPI_ISL_9301613, EPI_ISL_9301618 | Songkhla Hospital | Prince of Songkhla University | Anusara Wongkotsila; Chanon Kongkamon; Pongsakorn Choochuen; Samonrapat Surasombatpattana; Sarunyou Chusri; Surakameth Mahasirimongkol; Surasak Sangkhathat; Thammasin Ingviya; Thanit Sila; Virasakdi Chongsuvivatwong; Wison Laochareonsuk |
| EPI_ISL_9301688, EPI_ISL_9301689 | Songklanagarind hospital | Prince of Songkhla University | Anusara Wongkotsila; Chanon Kongkamon; Pongsakorn Choochuen; Samonrapat Surasombatpattana; Sarunyou Chusri; Surakameth Mahasirimongkol; Surasak Sangkhathat; Thammasin Ingviya; Thanit Sila; Virasakdi Chongsuvivatwong; Wison Laochareonsuk |
| EPI_ISL_12291218, EPI_ISL_12291238 | State Public Health Laboratory - INSACOG | NIV Influenza | Avudaiselvi Rathinasamy; Darez Ahamed; Devi Monika Ayyagari Venkata; Gurunathan Subramanian; Hemashree Kannan; Kalpana Raghu; Rajesh Kumar Manivannan; Raju Sivadoss; Sampath Palani; Selvavinayagam Sivaprakasam |
| EPI_ISL_10612213, EPI_ISL_11849183 | Swedish national genomic surveillance program of SARS-CoV-2 | The Public Health Agency of Sweden | Alma Brolund; Emmi Andersson; Maria Lind Karlberg; Maximilian Riess; Swedish national genomic surveillance program of SARS-CoV-2 |
| EPI_ISL_9698203, EPI_ISL_9698255, EPI_ISL_9963318, EPI_ISL_10444775, EPI_ISL_10444785, EPI_ISL_10444866, EPI_ISL_10444869, EPI_ISL_10444891, EPI_ISL_10444916 | | | |
| see above | Synlab Eesti OÜ | 1. Laboratory of Communicable Diseases (Estonia); 2. Eurofins Genomics Europe Sequencing GmbH | Abroi A.; Avi R.; Dotsenko L.; Epštein J.; Huik K.; Härma M-A.; Jaaniso E.; Kaarna K.; Kallas E.; Koppel I.; Kuzmin I.; Lahesaare A.; Lutsar I.; Metspalu M.; Milani L.; Naaber P.; Niglas H.; Oopkaup O.E.; Pauskar M.; Peterson H.; Päll T.; Ratnik K.; Raudvere U.; Reisberg T.; Sadikova O.; Sepp H.; Shablinskaja A.; Suija H.; Talas U.G.; Truusalu K. |
| EPI_ISL_10745657 | UAB "Baltic Medics" | National Public Health Surveillance Laboratory | Ana Steponkiene; Danas Baksa; Jelena Razmuk; Lukas Vasionis; Lukas Zemaitis; Migle Gabrielaite; Svajune Muralyte |
| EPI_ISL_9606453, EPI_ISL_9850775 | ULSNE - Braganca | Instituto Nacional de Saude (INSA) | Borges et al |
| EPI_ISL_10036476 | UMC Groningen, Clinical Virology, Department of Medical Microbiology and Infection Prevention | UMC Groningen, Clinical Virology, Department of Medical Microbiology and Infection Prevention | Alexander Friedrich; Coretta Van Leer-Buter; Erley Lizarazo-Forero; Hubert Niesters; Lilli Gard; Marjolein Knoester; Monika Fliss; Sigrid Rosema; Xuewei Zhou |
| EPI_ISL_9760530 | UW Virology Lab | UW Virology Lab | Alexander Greninger; Hong Xie; Isabel Arnould; Keith R Jerome; Meei-Li Huang; Pavitra Roychoudhury; Pooneh Hajian; Ricardo Perez; Saraswathi Sathees; Sean Ellis; Seffir T. Wendm; Shah Mohamed Bakhash |
| EPI_ISL_10953139 | Unidad de Investigación Biomédica de Zacatecas (UIBZ) | Microbial Genomics Laboratory | ; Alejandra García-Gasca; Alejandra Hernández-Terán; Alejandro Sánchez-Flores; Alfredo Herrera-Estrella; Alicia Ocaña-Mondragón; Andreu Comas-García; Angel Gustavo Salas-Lais; Antonio Loza Román; Bernardo Martínez-Miguel; Blanca Taboada; Brenda Irasema Maldonado-Meza; Bruno Gómez-Gil; Carla Ivón Herrera-Najera; Carlos F. Arias; Celia Boukadida; Clara Esperanza Santacruz-Tinoco; Concepción Grajales-Muñiz; Consorcio Mexicano de Vigilancia Genómica (CoViGen-Mex). Authors (in alphabetical order): Julio Elias Alvarado-Yaah; Cristóbal Cháidez-Quiróz; Célida Duque Molina; Célida Martínez-Rodríguez; Daniel Fregoso-Rueda; Daniel Lira Morales; Eduardo Becerril-Vargas; Fernando Fontove-Herrera; Fidencio Mejía-Nepomuceno; Francisco Pulido; Gloria Elena Espinosa-Ayala; Gloria María Molina-Salinas; Gloria Vazquez; Hector Esteban Paz-Juárez; Hector Montoya-Fuentes; Helen Haydee Fernanda Ramírez-Plascencia; Irvin González-López; Jean Pierre González; Jesús Hernández; Joel Armando Vázquez-Pérez.; Jorge Salas-Hernández; José Antonio Enciso-Moreno; José Arturo Martínez-Orozco; José Esteban Muñoz-Medina; José de Jesús Nuñez-Contreras; Juan Bautista Chale-Dzul; Julissa Enciso-Ibarra; Luis Alberto Ochoa-Carrera; Margarita Matías-Florentino; Mario Mújica-Sánchez; Marissa Perez-Garcia; María Guadalupe Santiago-Mauricio; María Guadalupe de Jesús Mireles-Rivera; Nelly Sélem-Mojica; Pavel Isa; Ricardo Ciria Merce; Ricardo Grande; Rosa María Gutiérrez Rios; Santiago Ávila-Ríos; Selene Zárate; Susana Lopez; Verónica Mata-Haro; Victor Eduardo García-Arias; Víctor Hugo Borja-Aburto |
| EPI_ISL_12650022 | Unilabs Laboratory Medicine | Norwegian Institute of Public Health, Department of Virology | Atiya R Ali; Debech Nadia; Engebretsen Serina Beate; Garcia Llorente Ignacio; Hilde Elshaug; Hilde Nordby Falkenhaug; Hilde Vollan; Jon Bråte; Kamilla Heddeland Instefjord; Karoline Bragstad; Kathrine Stene-Johansen; Line Victoria Moen; Marie Paulsen Madsen; Olav Hungnes; Pedersen Benedikte Nevjen; Rasmus Riis Kopperud |
| EPI_ISL_9075007, | University Clinical Hospital of | University of Sarajevo, Veterinary | Goletic S; Goletic T; Herceg I; Hodzic A; Jazic A; Nicevic M; Ostojic M; Sabic E; Skocibusic S; Softic A; Terzic I |

| | | | |
|---|---|---|---|
| EPI_ISL_9075015, EPI_ISL_9075021 | Mostar, Department of Microbiology and Molecular Diagnostics | Faculty, Laboratory for Molecular Diagnostic and Research | |
| EPI_ISL_8791223, EPI_ISL_9025461, EPI_ISL_9325219, EPI_ISL_10265762, EPI_ISL_10265780 | Universität Zürich | Institute of Medical Virology, University of Zurich | Alexandra Trkola; Annette Audigé; Catharine Aquino; Cyril Shah; Daniel Ehrsam; Gabriela Ziltener; Guido Bloemberg; Hubert Rehrauer; Isabel Stürmer; Joel Wirz; Jon Huder; Jürg Böni; Kevin Steiner; Maria Grünberg; Maryam Zaheri; Michael Huber; Riccarda Capaul; Stefan Schmutz; Verena Kufner; Weihong Qi |
| EPI_ISL_9025918, EPI_ISL_9325281, EPI_ISL_9622656, EPI_ISL_9622690, EPI_ISL_9622710, EPI_ISL_9881460 | UniversitätsSpital Zürich | Institute of Medical Virology, University of Zurich | Alexandra Trkola; Annette Audigé; Catharine Aquino; Cyril Shah; Daniel Ehrsam; Gabriela Ziltener; Guido Bloemberg; Hubert Rehrauer; Isabel Stürmer; Joel Wirz; Jon Huder; Jürg Böni; Kevin Steiner; Maria Grünberg; Maryam Zaheri; Michael Huber; Riccarda Capaul; Stefan Schmutz; Verena Kufner; Weihong Qi |
| EPI_ISL_10314277 | Universitätsklinikum Köln; Institut für Virologie | Robert Koch Institute | |
| EPI_ISL_11905699 | Urmia Reference Laboratory | National Influenza Center | A Nejati; Adel Abedi; J Yavarian; K Sadeghi; Marzieh Faraji-Zonouz and T Mokhtari Azad; NZ Shafiei Jandaghi; Nastaran Ghavami Sevrin Zadheidar; V Salimi |
| EPI_ISL_9402090, EPI_ISL_9402097 | VERMAAK | National Institute for Communicable Diseases of the National Health Laboratory Service | Amoako DG; Bhiman JN; Everatt J; Ismail A; Kekana D; Mahlangu B; Mnguni A; Mohale T; Ntuli N; Scheepers C; Wolter N |
| EPI_ISL_10851409 | Viesoji istaiga Klaipedos universitetine ligonine | National Public Health Surveillance Laboratory | Ana Steponkiene; Danas Baksa; Jelena Razmuk; Lukas Vasionis; Lukas Zemaitis; Migle Gabrielaite; Svajune Muralyte |
| EPI_ISL_11299840 | Viollier AG | Department of Biosystems Science and Engineering, ETH Zürich | Andrea Patrizia Salzmann; Chaoran Chen; Christian Beisel; Christiane Beckmann; Christoph Noppen; David Dreifuss; Elodie Burcklen; Franziska Singer; Henriette Kurth; Ina Nissen; Ivan Topolsky; Kim Philipp Jablonski; Lara Fuhrmann; Louis du Plessis; Matteo Carrara; Maurice Redondo; Mirjam Feldkamp; Natascha Santacroce; Niko Beerenwinkel; Olivier Kobel; Pelin Icer; Rebecca Denes; Sarah Nadeau; Shuqing Yu; Tanja Stadler; Tobias Schär |
| EPI_ISL_9353266, EPI_ISL_9353326, EPI_ISL_12172042, EPI_ISL_12172048, EPI_ISL_12172064 | Virology Laboratory, Scientific Department, Army Medical Center | Virology Laboratory, Scientific Department, Army Medical Center | Anella Monte; Anna Anselmo; Antonella Fortunato; Filippo Molinari; Florigio Lista; Francesco Giordani; Giancarlo Petralito; Giandomenico Cerreto; Giulia Campoli; Lucia Nicosia; Marzia Cavalli; Pietro Marco D'Angelo; Riccardo De Sanctis; Rossella Brandi; Silvia Fillo; Vanessa Vera Fain |
| EPI_ISL_9154644, EPI_ISL_9978176, EPI_ISL_9984839 | Vitomed Sp. z o.o. | 1. Academic Center for Pathomorphological and Genetic-Molecular Diagnostics ltd, Bialystok, Poland 2. National Institute of Public Health - National Institute of Hygiene, Warsaw, Poland | Anetta Sulewska; Jacek Nikliński; Janusz Dzięcioł; Joanna Kiśluk; Katarzyna Zacharczuk; Konrad Raczkowski; Magdalena Nowakowska; Małgorzata Sadkowska-Todys; Piotr Karabowicz; Piotr Majewski; Przemysław Biecek. Joanna Reszeć; Radosław Charkiewicz; Tomasz Wołkowicz |
| EPI_ISL_11924903 | West of Scotland Specialist Virology Centre, NHSGGC / MRC-University of Glasgow Centre for Virus Research | COVID-19 Genomics UK (COG-UK) Consortium | Alasdair MacLean; Ana da Silva Filipe; Andy Young; Antonia Ho; Daniel Mair; David L Robertson; Emily Goldstein; Emma Thomson; Gonzalo Yebra; Guy Mollett; Ioulia Tsatsani; James Shepherd; Jenna Nichols; Jessica Benkaroun; Jon Perkins; Jordan Ashworth; Joseph Hughes; Kathy Smollett; Kirsty Mangin; Kyriaki Nomikou; Lily Tong; Matthew Holden; Nicolas Suarez; Rachael Tomb; Rachel Blacow; Richard Orton; Rory Gunson; Sarah McDonald; Sharif Shaaban; Sreenu Vattipally |
| EPI_ISL_9414680 | White Buffalo, Inc | PSU Animal DiagnosticLaboratory | Meera Surendran Nair; Suresh Kuchipudi; Vivek Kapur |
| EPI_ISL_11934993 | Yourgene Health | COVID-19 Genomics UK (COG-UK) Consortium | UKHSA; Yourgene Health |
| EPI_ISL_9202992 | Zakład Patologii Nowotworów i Patomorfologii Centrum Onkologii im. prof. F. Łukaszczyka w Bydgoszczy | 1. Academic Center for Pathomorphological and Genetic-Molecular Diagnostics ltd, Bialystok, Poland 2. National Institute of Public Health - National Institute of Hygiene, Warsaw, Poland | Anetta Sulewska; Jacek Nikliński; Janusz Dzięcioł; Joanna Kiśluk; Katarzyna Zacharczuk; Konrad Raczkowski; Magdalena Nowakowska; Małgorzata Sadkowska-Todys; Piotr Karabowicz; Piotr Majewski; Przemysław Biecek. Joanna Reszeć; Radosław Charkiewicz; Tomasz Wołkowicz |
| EPI_ISL_9860127 | Zavod za javno zdravstvo Ličko-senjske županije | Hrvatski zavod za javno zdravstvo | Anita Jurić; Dragan Jurić; Irena Tabain; Ivana Ferenčak; Josipa Kuzle |
| EPI_ISL_9975863, EPI_ISL_9975913, EPI_ISL_9976348 | Zavod za javno zdravstvo Varaždinske županije | Hrvatski zavod za javno zdravstvo | Anita Jurić; Dragan Jurić; Irena Tabain; Ivana Ferenčak; Josipa Kuzle |
