## Supplementary material for "Tracing the international arrivals of SARS-CoV-2 Omicron variants after Aotearoa New Zealand reopened its border": BA.2.12.1 GISAID acknowledgements

We gratefully acknowledge the following Authors from the Originating laboratories responsible for obtaining the specimens, as well as the Submitting laboratories where the genome data were generated and shared via GISAID, on which this research is based.

All Submitters of data may be contacted directly via www.gisaid.org

Authors are sorted alphabetically.

Acknowledgement EPI_SET Identifier: EPI_SET_20220706us

| Accession ID | Originating Laboratory | Submitting Laboratory | Authors |
|---|---|---|---|
| EPI_ISL_12572794 | AREA DE SALUD FORTUNA | Inciensa, Instituto Costarricense de Investigación y Enseñanza en Nutrición y Salud | Adriana Godínez; Claudio Soto-Garita; Estela Cordero; Francisco Duarte; Gabriel Morales & Karolina Hall Loria; Hebleen Porras; José Luis Vargas; Mariela Gutiérrez; Melany Calderón; Sofia Herrera |
| EPI_ISL_13109457 | AREA DE SALUD GARABITO | Inciensa, Instituto Costarricense de Investigación y Enseñanza en Nutrición y Salud | Adriana Godínez; Claudio Soto-Garita; Estela Cordero; Francisco Duarte; Gabriel Morales; Hebleen Porras; José Luis Vargas; Mariela Gutiérrez; Melany Calderón; Natalia Bonilla & Andrea Moreno Carvajal; Sofia Herrera |
| EPI_ISL_13109453 | AREA DE SALUD PAVAS (COOPESALUD) | Inciensa, Instituto Costarricense de Investigación y Enseñanza en Nutrición y Salud | Adriana Godínez; Claudio Soto-Garita; Estela Cordero; Francisco Duarte; Gabriel Morales & Natalia Bonilla; Hebleen Porras; José Luis Vargas; Mariela Gutiérrez; Melany Calderón; Sofia Herrera |
| EPI_ISL_12935273 | AREA DE SALUD SANTA CRUZ | Inciensa, Instituto Costarricense de Investigación y Enseñanza en Nutrición y Salud | Adriana Godínez; Claudio Soto-Garita; Estela Cordero; Francisco Duarte; Gabriel Morales; Hebleen Porras; José Luis Vargas; Mariela Gutiérrez; Melany Calderón; Natalia Bonilla & Mónica Montoya; Sofia Herrera |
| EPI_ISL_13285186 | ASST GOM NIGUARDA | ASST Grande ospedale Metropolitano Niguarda | Alice Nava |
| EPI_ISL_12918611, EPI_ISL_13087666, EPI_ISL_13227223, EPI_ISL_13227233 | ASST MONZA | ASST MONZA | Sergio Maria Ivano Malandrin |
| EPI_ISL_12968865 | American Samoa DOH Clinical Laboratory Tafuna Family Health Center | State Laboratories Division, Hawaii State Department of Health | Ayana Garnet; Briana Ofilas; Cheryl-lynn Daquip; Cheyenne Barela; Daniel Strange; Drew Kuwazaki; Edward Desmond; Jeffrey Au; Mark Nagata; Pamela O'Brien; Remedios Gose; Samantha Cotter; Samantha Sruba |
| EPI_ISL_13297838 | Arcispedale Santa Maria Nuova Autoimmunità Allergologia e Biotecnologie Innovative | Istituto Zooprofilattico Sperimentale della Lombardia e dell'Emilia Romagna (IZSLER), Risk Analysis and Genomic Epidemiology Unit | Alessandro Zerbini; Erika Scaltriti; Ilaria Menozzi; Lucia Belloni; Marina Morganti; Stefania Croci; Stefano Pongolini |
| EPI_ISL_13199457 | Area of Virology, Serology and Virology Division (SAViD), New South Wales Health Pathology Randwick | Virology Research Laboratory; Area of Virology, Serology and Virology Division (SAViD), New South Wales Health Pathology Randwick | Foster, C.; Jean, T.; Rawlinson, W.; Van Hal, S.; Wong, M.; Yeang, M. |
| EPI_ISL_13276915, EPI_ISL_13276921, EPI_ISL_13276942 | Area of Virology, Serology and Virology Division (SAViD), New South Wales Health Pathology Randwick, Prince of Wales Hospital | Virology Research Laboratory, Area of Virology, Serology and Virology Division (SAViD), New South Wales Health Pathology Randwick, Prince Of Wales Hospital | Foster, C.; Jean, T.; Rawlinson, W.; Van Hal, S.; Wong, M.; Yeang, M. |
| EPI_ISL_12870016, EPI_ISL_12870018 | Area of Virology, Serology and Virology Division (SAViD), New South Wales Health Pathology Randwick, Prince of Wales Hospital | Virology Research Laboratory; Area of Virology, Serology and Virology Division (SAViD), New South Wales Health Pathology Randwick, Prince Of Wales Hospital | Foster, C.; Jean, T.; Rawlinson, W.; Van Hal, S.; Wong, M.; Yeang, M. |
| EPI_ISL_13059280, EPI_ISL_13059307 | Area of Virology, Serology and Virology Division (SAViD), New South Wales Health Pathology Randwick, Prince of Wales Hospital | Virology Research Laboratory; Area of Virology, Serology and Virology Division (SAViD), New South Wales Health Pathology Randwick, Prince of Wales Hospital | Foster, C.; Jean, T.; Rawlinson, W.; Van Hal, S.; Wong, M.; Yeang, M. |
| EPI_ISL_12808702 | Australian Clinical Labs (formerly Healthscope Pathology) | NSW Health Pathology - Institute of Clinical Pathology and Medical Research; Westmead Hospital; University of Sydney | Arnott A.; Draper J.; Gall M.; Martinez E.; Rockett R.; Sintchenko V.; on behalf of ICPMR |
| EPI_ISL_12606027 | Azienda Ospedaliero - Universitaria di Modena Policlinico - Virologia e Microbiologia Molecolare | Istituto Zooprofilattico Sperimentale della Lombardia e dell'Emilia Romagna (IZSLER), Risk Analysis and Genomic Epidemiology Unit | Erika Scaltriti; Giulia Fregni Serpini; Ilaria Menozzi; Marina Morganti; Monica Pecorari; Stefano Pongolini; William Gennari |
| EPI_ISL_12918854, EPI_ISL_13284343 | Azienda Sanitaria dell'Alto Adige - Laboratorio Aziendale di Microbiologia e Virologia | Azienda Sanitaria dell'Alto Adige | Irene Bianconi |
| EPI_ISL_13031916, EPI_ISL_13282591, EPI_ISL_13282682, EPI_ISL_13282705, EPI_ISL_13291939, EPI_ISL_13292088, EPI_ISL_13292168, EPI_ISL_13292473, EPI_ISL_13292511 | | | |
| see above | BioneXt Lab | Laboratoire national de sante, Microbiology, Microbial Genomics Platform | Anke Wienecke-Baldacchino; Catherine Ragimbeau; Elodie Solarino; Eric Hugoson; Fatu Djabi; Jessica Tapp; Lise Pignon; Raoul Salmon; Sibel Berger; Tamir Abdelrahman; Thibault Ferrandon; Virginie Jover |
| EPI_ISL_12966874, EPI_ISL_12966927, EPI_ISL_12967054, EPI_ISL_12967573, EPI_ISL_12967615, EPI_ISL_12968039 | British Columbia Centre For Disease Control | B.C. Centre for Disease Control Public Health Laboratory | Ana Pacagnella; Corrinne Ng; Dan Fornika; James Zlosnik; John Tyson; Kim Macdonald; Kimia Kamelian; Linda Hoang; Loretta Janz; Mel Krajden; Prystajecky Natalie; Robert Azana; Shannon Russell |
| EPI_ISL_12381897, EPI_ISL_12382570, EPI_ISL_12384337, EPI_ISL_12661665, EPI_ISL_12663137, EPI_ISL_12663617, EPI_ISL_12664541, EPI_ISL_12664673 | | | |
| see above | British Columbia Centre For Disease Control | BCCDC Public Health Laboratory | Ana Pacagnella; Corrinne Ng; Dan Fornika; James Zlosnik; John Tyson; Kim Macdonald; Kimia Kamelian; Linda Hoang; Loretta Janz; Mel Krajden; Prystajecky Natalie; Robert Azana; Shannon Russell |
| EPI_ISL_13068765, EPI_ISL_13068789, EPI_ISL_13068803, EPI_ISL_13068805, EPI_ISL_13068807, EPI_ISL_13068820, EPI_ISL_13068832, EPI_ISL_13223816, EPI_ISL_13223841 | | | |
| see above | CHUV | Laboratory of genomics and metagenomics, Institute of Microbiology, University Hospital Centre and University of Lausanne | Claire Bertelli; Damien Jacot; Gilbert Greub; Sébastien Aeby; Trestan Pillonel |
| EPI_ISL_13177319 | Centre Hospitalier Universitaire (CHU) Nimes | Centre Hospitalier Universitaire (CHU) Nîmes | Agathe Boudet; Milene Sasso; Sophie Bravo; Stephan Robin |
| EPI_ISL_13018096, EPI_ISL_13018097 | Centro de Investigación Biomédica de Occidente (CIBO) | Microbial Genomics Laboratory | ; Alejandra García-Gasca; Alejandra Hernández-Terán; Alejandro Sánchez-Flores; Alfredo Herrera-Estrella; Alicia Ocaña-Mondragón; Andreu Comas-García; Angel Gustavo Salas-Lais; Antonio Loza Román; Bernardo Martínez-Miguel; Blanca Taboada; Brenda Irasema Maldonado-Meza; Bruno Gómez-Gil; Carla Ivón Herrera-Najera; Carlos F. Arias; Celia Boukadida; Clara Esperanza Santacruz-Tinoco; Concepción Grajales-Muñiz; Consorcio Mexicano de Vigilancia Genómica (CoViGen-Mex). Authors (in alphabetical order): Julio Elias Alvarado-Yaah; Cristóbal Cháidez-Quiróz; Célida Duque Molina; Célida Martínez- Rodríguez; Daniel Fregoso-Rueda; Daniel Lira Morales; Eduardo Becerril-Vargas; Fernando Fontove-Herrera; Fidencio Mejía-Nepomuceno; Francisco Pulido; Gloria Elena Espinosa-Ayala; Gloria María Molina-Salinas; Gloria Vazquez; Hector Esteban Paz-Juárez; Hector Montoya-Fuentes; Helen Haydee Fernanda Ramírez-Plascencia; Irvin González-López; Jean Pierre González; Jesús Hernández; Joel Armando Vázquez-Pérez.; Jorge Salas-Hernández; José Antonio Enciso-Moreno; José Arturo Martínez-Orozco; José Esteban Muñoz-Medina; José de Jesús Nuñez-Contreras; Juan Bautista Chale-Dzul; Julissa Enciso-Ibarra; Luis Alberto Ochoa-Carrera; Margarita Matías-Florentino; Mario Mújica-Sánchez; Marissa Perez-Garcia; María Guadalupe Santiago-Mauricio; María Guadalupe de Jesús Mireles-Rivera; Nelly Sélem-Mojica; Pavel Isa; Ricardo Ciria Merce; Ricardo Grande; Rosa María Gutiérrez Rios; Santiago Ávila-Ríos; Selene Zárate; Susana Lopez; Verónica Mata-Haro; Victor Eduardo García-Arias; Víctor Hugo Borja-Aburto |

| | | | |
|---|---|---|---|
| EPI_ISL_13018124 | Centro de Investigación Biomédica del Noreste (CIBIN) | Microbial Genomics Laboratory | ; Alejandra García-Gasca; Alejandra Hernández-Terán; Alejandro Sánchez-Flores; Alfredo Herrera-Estrella; Alicia Ocaña-Mondragón; Andreu Comas-García; Angel Gustavo Salas-Lais; Antonio Loza Román; Bernardo Martínez-Miguel; Blanca Taboada; Brenda Irasema Maldonado-Meza; Bruno Gómez-Gil; Carla Ivón Herrera-Najera; Carlos F. Arias; Celia Boukadida; Clara Esperanza Santacruz-Tinoco; Concepción Grajales-Muñiz; Consorcio Mexicano de Vigilancia Genómica (CoViGen-Mex). Authors (in alphabetical order): Julio Elias Alvarado-Yaah; Cristóbal Cháidez-Quiróz; Célida Duque Molina; Célida Martínez- Rodríguez; Daniel Fregoso-Rueda; Daniel Lira Morales; Eduardo Becerril-Vargas; Fernando Fontove-Herrera; Fidencio Mejía-Nepomuceno; Francisco Pulido; Gloria Elena Espinosa-Ayala; Gloria María Molina-Salinas; Gloria Vazquez; Hector Esteban Paz-Juárez; Hector Montoya-Fuentes; Helen Haydee Fernanda Ramírez-Plascencia; Irvin González-López; Jean Pierre González; Jesús Hernández; Joel Armando Vázquez-Pérez.; Jorge Salas-Hernández; José Antonio Enciso-Moreno; José Arturo Martínez-Orozco; José Esteban Muñoz-Medina; José de Jesús Nuñez-Contreras; Juan Bautista Chale-Dzul; Julissa Enciso-Ibarra; Luis Alberto Ochoa-Carrera; Margarita Matías-Florentino; Mario Mújica-Sánchez; Marissa Perez-Garcia; María Guadalupe Santiago-Mauricio; María Guadalupe de Jesús Mireles-Rivera; Nelly Sélem-Mojica; Pavel Isa; Ricardo Ciria Merce; Ricardo Grande; Rosa María Gutiérrez Rios; Santiago Ávila-Ríos; Selene Zárate; Susana Lopez; Verónica Mata-Haro; Victor Eduardo García-Arias; Víctor Hugo Borja-Aburto |
| EPI_ISL_12916151, EPI_ISL_13074889, EPI_ISL_13075023, EPI_ISL_13075041, EPI_ISL_13075182 | Clinical Microbiology Laboratory, Tel Aviv Sourasky Medical Center | Clinical Microbiology Laboratory, Tel Aviv Sourasky Medical Center | Alon Ziv; Amos Adler; Goel Morad; Katya Levytskyi; Lior Handler; Matan Slutzkin; Ora Halutz; Orly Eshel |
| EPI_ISL_13295814 | Colorado Department of Public Health and Environment | Colorado Department of Public Health and Environment | Alexandria Rossheim; Arianna Smith; Diana Ir; Emily A. Travanty; Laura Bankers; Mandy Waters; Michael Martin; Molly C. Hetherington-Rauth; Shannon R. Matzinger |
| EPI_ISL_13035997 | Department für Labormedizin Abteilung III Bereich Molekulare Diagnostik Universitätsklinikum Halle(Saale) | Robert Koch Institute | |
| EPI_ISL_12632417, EPI_ISL_12895353, EPI_ISL_12895738, EPI_ISL_12993951, EPI_ISL_12994838, EPI_ISL_13030124, EPI_ISL_13049829, EPI_ISL_13067091, EPI_ISL_13090399, EPI_ISL_13127770, EPI_ISL_13127884, EPI_ISL_13177784, EPI_ISL_13177852, EPI_ISL_13178841, EPI_ISL_13178943, EPI_ISL_13241580, EPI_ISL_13280703, EPI_ISL_13299539, EPI_ISL_13299658, EPI_ISL_13299782 | | | |
| see above | Department of Bacteria, Parasites and Fungi, Statens Serum Institut, Copenhagen, Denmark | Statens Serum Institut Bioinformatics and Microbial Genomics | Danish Covid-19 Genome Consortium |
| EPI_ISL_13090220, EPI_ISL_13216471, EPI_ISL_13251688, EPI_ISL_13251692 | Department of Clinical Microbiology | GIGA Medical Genomics | Claire Gourzonès; Cécile Meex; Keith Durkin; Laurent Gillet; Maria Artesi; Marie-Pierre Hayette; Nadine Cambisano; Nathalie Renotte; Olivier Ek; Sébastien Bontems; Vincent Bours |
| EPI_ISL_12812530, EPI_ISL_12812534, EPI_ISL_12812562, EPI_ISL_13140325, EPI_ISL_13140329, EPI_ISL_13140331, EPI_ISL_13140342, EPI_ISL_13140466, EPI_ISL_13140497, EPI_ISL_13140514, EPI_ISL_13140531, EPI_ISL_13140542, EPI_ISL_13140543, EPI_ISL_13140547, EPI_ISL_13140557 | | | |
| see above | Department of Health Technology and Informatics, The Hong Kong Polytechnic University | Department of Health Technology and Informatics, The Hong Kong Polytechnic University | Alan Ka-Lun Wu; Alex Yat-Man Ho; Barry Kin-Chung Wong; Chloe Toi-Mei Chan; David Ho-Keung Shum; Gilman Kit-Hang Siu; Hiu-Yin Lao; Ivan Tak-Fai Wong; Jake Siu-Lun Leung; Kam-Tong Yip; Kenneth Siu-Sing Leung; Kingsley King-Gee Tam; Kitty Sau-Chun Fung; Kristine Luk; Lam-Kwong Lee; Miranda Chong-Yee Yau; Sandy Ka-Yee Chau; Shea Ping Yip; Tak-Lun Que; Timothy Ting-Leung Ng; Wing Cheong Yam; Wing-Hei Lo; Wing-Kin To; Yvette Wai-Man Lai |
| EPI_ISL_12471280, EPI_ISL_12869517, EPI_ISL_13132094 | Department of Medical Microbiology & Infection prevention, Amsterdam University Medical Centers location AMC | Department of Medical Microbiology & Infection prevention, Amsterdam University Medical Centers location AMC | Akke Cornelissen; Fokla Zorgdrager; Janke Schinkel; Jelle Koopsen; Judith den Uil; Marcel Jonges; Matthijs Welkers; Menno de Jong; Menno de Jong and Mariken van der Lubben on behalf of the Amsterdam Regional Genomic epidemiology and Outbreak Surveillance (ARGOS) consortium; Robin van Houdt; Sebastien Matamoros; Sjoerd Rebers; Sylvia Bruisten; Tjalling Leenstra and Mariken van der Lubben on behalf of the Amsterdam Regional Genomic epidemiology and Outbreak Surveillance (ARGOS) consortium |
| EPI_ISL_13181259 | Department of Medical Microbiology - section Molde, Molde Hospital | Norwegian Institute of Public Health, Department of Virology | Atiya R Ali; Debech Nadia; Engebretsen Serina Beate; Garcia Llorente Ignacio; Hilde Elshaug; Hilde Nordby Falkenhaug; Hilde Vollan; Jon Bråte; Kamilla Heddeland Instefjord; Karoline Bragstad; Kathrine Stene-Johansen; Line Victoria Moen; Marie Paulsen Madsen; Olav Hungnes; Pedersen Benedikte Nevjen; Rasmus Riis Kopperud |
| EPI_ISL_13182855 | Department of Medical Microbiology, Baerum Hospital, Vestre Viken Health Trust | Norwegian Institute of Public Health, Department of Virology | Atiya R Ali; Debech Nadia; Engebretsen Serina Beate; Garcia Llorente Ignacio; Hilde Elshaug; Hilde Nordby Falkenhaug; Hilde Vollan; Jon Bråte; Kamilla Heddeland Instefjord; Karoline Bragstad; Kathrine Stene-Johansen; Line Victoria Moen; Marie Paulsen Madsen; Olav Hungnes; Pedersen Benedikte Nevjen; Rasmus Riis Kopperud |
| EPI_ISL_12981760, EPI_ISL_12982778, EPI_ISL_13181514 | Department of Medical Microbiology, St. Olavs hospital | Norwegian Institute of Public Health, Department of Virology | Atiya R Ali; Debech Nadia; Engebretsen Serina Beate; Garcia Llorente Ignacio; Hilde Elshaug; Hilde Nordby Falkenhaug; Hilde Vollan; Jon Bråte; Kamilla Heddeland Instefjord; Karoline Bragstad; Kathrine Stene-Johansen; Line Victoria Moen; Marie Paulsen Madsen; Olav Hungnes; Pedersen Benedikte Nevjen; Rasmus Riis Kopperud |
| EPI_ISL_13182953 | Dept. of Medical Microbiology, Stavanger University Hospital, Helse Stavanger HF | Norwegian Institute of Public Health, Department of Virology | Atiya R Ali; Debech Nadia; Engebretsen Serina Beate; Garcia Llorente Ignacio; Hilde Elshaug; Hilde Nordby Falkenhaug; Hilde Vollan; Jon Bråte; Kamilla Heddeland Instefjord; Karoline Bragstad; Kathrine Stene-Johansen; Line Victoria Moen; Marie Paulsen Madsen; Olav Hungnes; Pedersen Benedikte Nevjen; Rasmus Riis Kopperud |
| EPI_ISL_13090981 | Dept. of Microbiology and Infection Control, Akershus University Hospital HF | Dept. of Microbiology and Infection Control, Akershus University Hospital HF | Alexander Hesselberg Løvestad; Divya Murugananthan; Hanne Berggreen; Hege Vangstein Aamot |
| EPI_ISL_13198059 | Diagnostyka Sp. z o. o. Lodz | 1. Academic Center for Pathomorphological and Genetic-Molecular Diagnostics ltd, Bialystok, Poland 2. National Institute of Public Health - National Institute of Hygiene, Warsaw, Poland | Anetta Sulewska; Jacek Niklinski; Janusz DzięcioL; Joanna Kiśluk; Katarzyna Zacharczuk; Konrad Raczkowski; MaLgorzata Sadkowska-Todys; Magdalena Nowakowska; Piotr Karabowicz; Piotr Majewski; PrzemysLaw Biecek. Joanna Reszeć; RadosLaw Charkiewicz; Tomasz WoLkowicz |
| EPI_ISL_12635082, EPI_ISL_13086514, EPI_ISL_13137582, EPI_ISL_13137913, EPI_ISL_13200241 | Division of Emerging Infectious Diseases, Bureau of Infectious Diseases Diagnosis Control, Korea Disease Control and Prevention Agency | Division of Emerging Infectious Diseases, Bureau of Infectious Diseases Diagnosis Control, Korea Disease Control and Prevention Agency | Ae Kyung Park; Chae Young Lee; Eun-Jin Kim; Hyuck Jin Lee; Il-Hwan Kim; Jeong-Ah Kim |
| EPI_ISL_13084086, EPI_ISL_13084087 | Division of Infectious Disease Diagnosis Control, Capital Regional Center for Disease Control and Prevention, Korea Disease Control and Prevention Agency, KDCA | Division of Emerging Infectious Diseases, Bureau of Infectious Diseases Diagnosis Control, Korea Disease Control and Prevention Agency | EunJung Lee; Jeong-Gu Nam; SaHyun Hong |
| EPI_ISL_13281606, EPI_ISL_13281618 | Division of Infectious Disease Diagnosis Control, Gyeongnam Regional Center for Disease Control and Prevention, Korea Disease Control and Prevention Agency, KDCA | Division of Emerging Infectious Diseases, Bureau of Infectious Diseases Diagnosis Control, Korea Disease Control and Prevention Agency | Byung Hak Kang; Dongchul Park; Seon Do Hwang |
| EPI_ISL_12932997 | Douglass Hanly Moir Pathology | NSW Health Pathology - Institute of Clinical Pathology and Medical Research; Westmead Hospital; University of Sydney | Arnott A.; Draper J.; Gall M.; Martinez E.; Rockett R.; Sintchenko V.; on behalf of ICPMR |
| EPI_ISL_13215555, EPI_ISL_13215597 | Edmonton Provincial Lab | Alberta Precision Labs (APL) | Buss E; Croxen M; Deo A; Dieu P; Ferrato C; Gill K; Koleva P; Li V; Lloyd C; Lynch T; Ma R; Murphy S; Pabbaraju K; Shideler S; Shokopoles S; Skitsko T; Thayer J; Tipples G; Wong A; Yu C; Zelyas N. |
| EPI_ISL_13034611 | Eurofins MVZ Labor Gelsenkirchen | Robert Koch Institute | |
| EPI_ISL_13134161, EPI_ISL_13294894 | Eurofins-NMDL | Eurofins-NMDL | Anco Molijn; Anne Vogel; Lisa Dreesens; Marvin Ruiter; Maurine Leversteijn-van Hall; Roy Masius; Simon Lansu |
| EPI_ISL_13104272, EPI_ISL_13247784, EPI_ISL_13247811 | Fondazione IRCCS Ca' Granda Ospedale Maggiore Policlinico | Fondazione IRCCS Ca' Granda Ospedale Maggiore Policlinico | Ferruccio Ceriotti; Sara Uceda Renteria |
| EPI_ISL_12223766, EPI_ISL_12613086, EPI_ISL_12613126, EPI_ISL_12953155, EPI_ISL_12953241, EPI_ISL_12953244, EPI_ISL_12953266, EPI_ISL_12953284, EPI_ISL_12953286, EPI_ISL_13305203, EPI_ISL_13305219, EPI_ISL_13305228, EPI_ISL_13305276, EPI_ISL_13307956, EPI_ISL_13310979, EPI_ISL_13311025, EPI_ISL_13311032, EPI_ISL_13311033, EPI_ISL_13311047 | | | |
| see above | Gandhi Medical College and Hospital (GMCH), Secunderabad | NIV Influenza | Abdul Majeed; Amrithesh Kumar Arun; D.R.Manisha Rani; Devendhar; Dr.G.Sushma Rajya Lakshmi; Dr.K.Nagamani; Dr.Sunitha Pakalapaty; Hajeera Osmani; Sahithya |
| EPI_ISL_12739971, EPI_ISL_12740011 | Genetica Molecular and Subdepartamento de Virologia ISP Chile | Instituto de Salud Publica de Chile | Andres Castillo; Barbara Parra; Constanza Campano; Ivan Ponce; Jorge Fernandez; Karen Orostica; Marcelo Rojas; Matias Pezoa; Patricia Bustos; Rodrigo Fasce |
| EPI_ISL_13040429 | Genomics for Life | Public Health Virology - Forensic and Scientific Services (PHV-FSS) | Chenwei Wang on behalf of Q-PHIRE Genomics |
| EPI_ISL_12832916 | HOSPITAL DE CIUDAD NEILY | Inciensa, Instituto Costarricense de | Adriana Godínez; Claudio Soto-Garita; Estela Cordero; Francisco Duarte; Gabriel Morales; Hebleen Porras; José Luis Vargas; Mariela Gutiérrez; Melany Calderón; Natalia Bonilla & Gabriela Marchena Jimenez; Sofia Herrera |

| | | | |
|---|---|---|---|
| | | Investigación y Enseñanza en Nutrición y Salud | |
| EPI_ISL_12935315, EPI_ISL_12935316 | HOSPITAL DR. RAFAEL A. CALDERON GUARDIA | Inciensa, Instituto Costarricense de Investigación y Enseñanza en Nutrición y Salud | Adriana Godínez; Claudio Soto-Garita; Estela Cordero; Francisco Duarte; Gabriel Morales; Hebleen Porras; José Luis Vargas; Mariela Gutiérrez; Melany Calderón; Natalia Bonilla & Ericka Madrigal Arias; Sofia Herrera |
| EPI_ISL_12832953 | HOSPITAL DR. TOMAS CASAS CASAJUS | Inciensa, Instituto Costarricense de Investigación y Enseñanza en Nutrición y Salud | Adriana Godínez; Claudio Soto-Garita; Estela Cordero; Francisco Duarte; Gabriel Morales; Hebleen Porras; José Luis Vargas; Mariela Gutiérrez; Melany Calderón; Natalia Bonilla & Antony Orozco Barquero; Sofia Herrera |
| EPI_ISL_12935330 | HOSPITAL GUAPILES | Inciensa, Instituto Costarricense de Investigación y Enseñanza en Nutrición y Salud | Adriana Godínez; Claudio Soto-Garita; Estela Cordero; Francisco Duarte; Gabriel Morales; Hebleen Porras; José Luis Vargas; Mariela Gutiérrez; Melany Calderón; Natalia Bonilla & Cesar Cerdas Quesada; Sofia Herrera |
| EPI_ISL_13209262, EPI_ISL_13209264, EPI_ISL_13209268, EPI_ISL_13209270, EPI_ISL_13209271, EPI_ISL_13209282, EPI_ISL_13209283 | | | |
| see above | HOSPITAL UNIVERSITARIO CENTRAL DE ASTURIAS | Laboratorio de Virología HUCA | ; Alba L; Alvarez-Arguelles ME; Boga JA; Costales I; Coto E; González-Alba JM; Gómez de Oña J; Martín-Rodríguez G; Melón S; Perez-Martínez Z; Rojo S; Sandoval M |
| EPI_ISL_13300448 | Hia Brest | Centre Hospitalier Universitaire (CHU) Brest | Adissa Tran Minoui; Christopher Payan; Léa Pilorgé; Simon Rambaud; Sophie Vallet |
| EPI_ISL_13018129 | Hospital Margarita Maza de Juárez | Microbial Genomics Laboratory | ; Alejandra García-Gasca; Alejandra Hernández-Terán; Alejandro Sánchez-Flores; Alfredo Herrera-Estrella; Alicia Ocaña-Mondragón; Andreu Comas-García; Angel Gustavo Salas-Lais; Antonio Loza Román; Bernardo Martínez-Miguel; Blanca Taboada; Brenda Irasema Maldonado-Meza; Bruno Gómez-Gil; Carla Ivón Herrera-Najera; Carlos F. Arias; Celia Boukadida; Clara Esperanza Santacruz-Tinoco; Concepción Grajales-Muñiz; Consorcio Mexicano de Vigilancia Genómica (CoViGen-Mex). Authors (in alphabetical order): Julio Elias Alvarado-Yaah; Cristóbal Cháidez-Quiróz; Célida Duque Molina; Célida Martínez- Rodríguez; Daniel Fregoso-Rueda; Daniel Lira Morales; Eduardo Becerril-Vargas; Fernando Fontove-Herrera; Fidencio Mejía-Nepomuceno; Francisco Pulido; Gloria Elena Espinosa-Ayala; Gloria María Molina-Salinas; Gloria Vazquez; Hector Esteban Paz-Juárez; Hector Montoya-Fuentes; Helen Haydee Fernanda Ramírez-Plascencia; Irvin González-López; Jean Pierre González; Jesús Hernández; Joel Armando Vázquez-Pérez.; Jorge Salas-Hernández; José Antonio Enciso-Moreno; José Arturo Martínez-Orozco; José Esteban Muñoz-Medina; José de Jesús Nuñez-Contreras; Juan Bautista Chale-Dzul; Julissa Enciso-Ibarra; Luis Alberto Ochoa-Carrera; Margarita Matías-Florentino; Mario Mújica-Sánchez; Marissa Perez-Garcia; María Guadalupe Santiago-Mauricio; María Guadalupe de Jesús Mireles-Rivera; Nelly Sélem-Mojica; Pavel Isa; Ricardo Ciria Merce; Ricardo Grande; Rosa María Gutiérrez Rios; Santiago Ávila-Ríos; Selene Zárate; Susana Lopez; Verónica Mata-Haro; Victor Eduardo García-Arias; Víctor Hugo Borja-Aburto |
| EPI_ISL_13105923 | Hospital Universitario 12 de Octubre | Hospital Universitario 12 de Octubre | Carmen Martín-Higuera; Esther Viedma; Irene Múñoz-Gallego; M.ª Dolores Folgueira; Mar Aguilera; Noelia Moral; Rafael Delgado; Sagrario Zurita |
| EPI_ISL_13304467, EPI_ISL_13304493, EPI_ISL_13304546, EPI_ISL_13304567, EPI_ISL_13304579 | ICMR-National Institute of Virology - INSACOG | NIV Influenza | Dr. Varsha Potdar and NIC Team |
| EPI_ISL_12982710 | Innlandet Hospital Trust, Division Lillehammer, Department for Medical Microbiology | Norwegian Institute of Public Health, Department of Virology | Atiya R Ali; Debech Nadia; Engebretsen Serina Beate; Garcia Llorente Ignacio; Hilde Elshaug; Hilde Nordby Falkenhaug; Hilde Vollan; Jon Bråte; Kamilla Heddeland Instefjord; Karoline Bragstad; Kathrine Stene-Johansen; Line Victoria Moen; Marie Paulsen Madsen; Olav Hungnes; Pedersen Benedikte Nevjen; Rasmus Riis Kopperud |
| EPI_ISL_13002093, EPI_ISL_13015511 | Institut für Immunologie und Genetik Kaiserslautern: Medizinisches Labor Dr. med. B. Thiele | Robert Koch Institute | |
| EPI_ISL_13183482, EPI_ISL_13183510, EPI_ISL_13183517, EPI_ISL_13183534 | Jessa | Jessa | Laura Vanstraelen et al. on behalf of the Jessa_cmdLab |
| EPI_ISL_13307011 | Kasturba Hospital Molecular Lab | NIV Influenza | Jayanthi Shastri; Vidushi Chitalia |
| EPI_ISL_12905607, EPI_ISL_12905610 | Kijabe Hospital | KEMRI-Wellcome Trust Research Programme,Kilifi | Agoti C.; D.J.Nokes; Githinji G.; Lambisia A.; Makori T.; Mburu M.W.; Mohamed K.S.; Morobe J.; Ndwiga L.; Ochola I; Ongera E.; de Laurent Z. |
| EPI_ISL_12854414, EPI_ISL_12854421, EPI_ISL_12854429, EPI_ISL_13032314, EPI_ISL_13032363 | Klinika za infektivne bolesti "Dr. Fran Mihaljević" | Hrvatski zavod za javno zdravstvo | Anita Jurić; Dragan Jurić; Irena Tabain; Ivana Ferenčak; Josipa Kuzle |
| EPI_ISL_13141704 | Klinikum Ernst von Bergmann gemeinnützige GmbH - stationärer Bereich | Robert Koch Institute | |
| EPI_ISL_12983339 | Klinisch Laboratorium ZNA | Klinisch Laboratorium ZNA | Verstrepen et al. |
| EPI_ISL_13192071 | LESP Sinaloa | Instituto de Diagnostico y Referencia Epidemiologicos (INDRE) | Abril Rodriguez-Maldonado; Ariadna Medina-Benitez; Claudia Wong-Arambula; Ernesto Ramirez-Gonzalez.; Fernando Gonzalez-Dominguez; Gisela Barrera-Badillo; Irma Lopez-Martinez; Joaquin Quiroz-Mercado; Lucia Hernandez-Rivas; Maribel Gonzalez-Villa; Natividad Cruz-Ortiz; Ruth Madera-Sandoval; Tatiana Nunez-Garcia; Vanessa Rivero-Arredondo |
| EPI_ISL_13192069 | LESP Tabasco | Instituto de Diagnostico y Referencia Epidemiologicos (INDRE) | Abril Rodriguez-Maldonado; Ariadna Medina-Benitez; Claudia Wong-Arambula; Ernesto Ramirez-Gonzalez.; Fernando Gonzalez-Dominguez; Gisela Barrera-Badillo; Irma Lopez-Martinez; Joaquin Quiroz-Mercado; Lucia Hernandez-Rivas; Maribel Gonzalez-Villa; Natividad Cruz-Ortiz; Ruth Madera-Sandoval; Tatiana Nunez-Garcia; Vanessa Rivero-Arredondo |
| EPI_ISL_12953088, EPI_ISL_13133552 | LKO | Jessa | Laura Vanstraelen et al. on behalf of the Jessa_cmdLab |
| EPI_ISL_13238512 | Labor Blackholm MVZ | Robert Koch Institute | |
| EPI_ISL_12678403, EPI_ISL_12678503 | Labor Mönchengladbach MVZ Dr. Stein + Kollegen GbR | Robert Koch Institute | |
| EPI_ISL_13244831 | Labor Prof. Dr. G. Enders MVZ GbR | Robert Koch Institute | |
| EPI_ISL_12702619, EPI_ISL_13292379 | Laboratoire national de sante, Microbiology, Virology | Laboratoire national de sante, Microbiology, Microbial Genomics Platform | Anke Wienecke-Baldacchino; Catherine Ragimbeau; Elodie Solarino; Eric Hugoson; Fatu Djabi; Jessica Tapp; Lise Pignon; Raoul Salmon; Sibel Berger; Tamir Abdelrahman; Thibault Ferrandon; Trung Nguyen Nguyen; Virginie Jover |
| EPI_ISL_13292055 | Laboratoires Reunis | Laboratoire national de sante, Microbiology, Microbial Genomics Platform | Anke Wienecke-Baldacchino; Bernard Weber; Catherine Ragimbeau; Elodie Solarino; Eric Hugoson; Fatu Djabi; Jessica Tapp; Lise Pignon; Raoul Salmon; Sibel Berger; Tamir Abdelrahman; Virginie Jover |
| EPI_ISL_12452939, EPI_ISL_13282603, EPI_ISL_13292283, EPI_ISL_13292284, EPI_ISL_13292341, EPI_ISL_13292426 | Laboratoires d'analyses medicales - KETTERTHILL | Laboratoire national de sante, Microbiology, Microbial Genomics Platform | Anke Wienecke-Baldacchino; Caroline Scheiber; Catherine Ragimbeau; Elodie Solarino; Eric Hugoson; Fatu Djabi; Jessica Tapp; Lise Pignon; Raoul Salmon; Serge Vedy; Sibel Berger; Tamir Abdelrahman; Virginie Jover |
| EPI_ISL_12837673, EPI_ISL_12837770 | Laboratoires d'analyses medicales - KETTERTHILL | Microbiology, Microbial Genomics Platform, LNS Laboratoire National De Santé | Anke Wienecke-Baldacchino; Caroline Scheiber; Catherine Ragimbeau; Elodie Solarino; Eric Hugoson; Fatu Djabi; Jessica Tapp; Lise Pignon; Raoul Salmon; Serge Vedy; Sibel Berger; Tamir Abdelrahman; Virginie Jover |
| EPI_ISL_13002244 | Laboratori de Referencia de Catalunya | Laboratori de Referencia de Catalunya | Bellosillo B.; Canal M.; Hernandez JJ.; Padilla E.; Ramirez A.; Vilas A. |
| EPI_ISL_13018078, EPI_ISL_13018086, EPI_ISL_13018100 | Laboratorio Central de Epidemiología (LCE) | Microbial Genomics Laboratory | ; Alejandra García-Gasca; Alejandra Hernández-Terán; Alejandro Sánchez-Flores; Alfredo Herrera-Estrella; Alicia Ocaña-Mondragón; Andreu Comas-García; Angel Gustavo Salas-Lais; Antonio Loza Román; Bernardo Martínez-Miguel; Blanca Taboada; Brenda Irasema Maldonado-Meza; Bruno Gómez-Gil; Carla Ivón Herrera-Najera; Carlos F. Arias; Celia Boukadida; Clara Esperanza Santacruz-Tinoco; Concepción Grajales-Muñiz; Consorcio Mexicano de Vigilancia Genómica (CoViGen-Mex). Authors (in alphabetical order): Julio Elias Alvarado-Yaah; Cristóbal Cháidez-Quiróz; Célida Duque Molina; Célida Martínez- Rodríguez; Daniel Fregoso-Rueda; Daniel Lira Morales; Eduardo Becerril-Vargas; Fernando Fontove-Herrera; Fidencio Mejía-Nepomuceno; Francisco Pulido; Gloria Elena Espinosa-Ayala; Gloria María Molina-Salinas; Gloria Vazquez; Hector Esteban Paz-Juárez; Hector Montoya-Fuentes; Helen Haydee Fernanda Ramírez-Plascencia; Irvin González-López; Jean Pierre González; Jesús Hernández; Joel Armando Vázquez-Pérez.; Jorge Salas-Hernández; José Antonio Enciso-Moreno; José Arturo Martínez-Orozco; José Esteban Muñoz-Medina; José de Jesús Nuñez-Contreras; Juan Bautista Chale-Dzul; Julissa Enciso-Ibarra; Luis Alberto Ochoa-Carrera; Margarita Matías-Florentino; Mario Mújica-Sánchez; Marissa Perez-Garcia; María Guadalupe Santiago-Mauricio; María Guadalupe de Jesús Mireles-Rivera; Nelly Sélem-Mojica; Pavel Isa; Ricardo Ciria Merce; Ricardo Grande; Rosa María Gutiérrez Rios; Santiago Ávila-Ríos; Selene Zárate; Susana Lopez; Verónica Mata-Haro; Victor Eduardo García-Arias; Víctor Hugo Borja-Aburto |
| EPI_ISL_13282998 | Laboratorio di Patologia Clinica, Ospedale San Paolo in Valloria, ASL 2 Liguria | U.O. Igiene, Ospedale Policlinico San Martino | Bruzzone Bianca; De Pace Vanessa; Domnich Alexander; Icardi Giancarlo on behalf of SARS-CoV-2 ITALIAN RESEARCH ENTERPRISE-(SCIRE) Collaborative Group; Lillo Flavia; Orsi Andrea; Randazzo Nadia; Ricucci Valentina; Stefanelli Federica |
| EPI_ISL_12594388, EPI_ISL_12620925, EPI_ISL_12758416, EPI_ISL_12889656, EPI_ISL_12889672, EPI_ISL_12974019, EPI_ISL_13080443, EPI_ISL_13118718, EPI_ISL_13151881, EPI_ISL_13161853, EPI_ISL_13219717, EPI_ISL_13309702, EPI_ISL_13314752, EPI_ISL_13315013, EPI_ISL_13315577 | | | |
| see above | Laboratory Corporation of | Centers for Disease Control and | Amanda Douglas; Amanda Suchanek; Andrea Throop; Ayla Burns; Benjamin Rambo-Martin; Bobbi Croy; Brian Krueger; Brian Norvell; Christopher Gulvick; Christos Petropoulos; Clinton Paden; Craig Lukasik; Dakota Howard; Debbie Boles; Dhwani Batra; Duncan MacCannell; Eyad Almasri; Goran Stevovic; Howard |

| | | | |
|---|---|---|---|
| | America | Prevention Division of Viral Diseases, Pathogen Discovery | Engler; Hrushikesh Deshmukh; Jake Humphrey; Jana Schroth; Jason Caravas; Joe Voshell; John Pruitt; Jonathan Meltzer; Jonathan Williams; Kimberly Wagner; Kristine Lacek; Lax Iyer; Lisa Pfefferle; Lyndon Tilson; Manoj Jain; Marcia Eisenberg; Mary Cristobal; Mary Williamson; Matthew Robinson; Matthew Schmerer; Michael Levandoski; Mike Sapeta; Mindy Nye; Minoo Agarwal; Mohan Kolli; Nuthawin Charoensri; Oren Cohen; Peter Cook; Prashant Gupta; Qian Zeng; Rama Ghatti; Scott Parker; Scott Ryan; Scott Sammons; Shatavia Morrison; Stanley Letovsky; Steven Ragan; Suresh Selvaraju; Susan Countryman; Susan Hicks; Suzanne Dale; Thomas Urban; Tim Kuphal; Tricia Zwiefelhofer; Tymeckia Kendall; Victoria Caban Figueroa; Vincent Drouillon; Yvette Unoarumhi |
| EPI_ISL_13019185, EPI_ISL_13256194 | Laboratory of Clinical Microbiology, Virology and Bioemergencies, ASST Fatebenefratelli Sacco - Sacco Hospital | Laboratory of Clinical Microbiology, Virology and Bioemergencies, ASST Fatebenefratelli Sacco - Sacco Hospital | Valeria Micheli |
| EPI_ISL_11984839, EPI_ISL_11984847, EPI_ISL_12471927, EPI_ISL_12610653, EPI_ISL_12875109, EPI_ISL_12875111, EPI_ISL_12875118, EPI_ISL_12875129, EPI_ISL_12875167, EPI_ISL_12875174, EPI_ISL_12875175, EPI_ISL_12875180, EPI_ISL_12875181, EPI_ISL_13228732, EPI_ISL_13228746, EPI_ISL_13228752, EPI_ISL_13228756, EPI_ISL_13228762, EPI_ISL_13238781, EPI_ISL_13238814, EPI_ISL_13238820, EPI_ISL_13250096, EPI_ISL_13250136, EPI_ISL_13281195 | | | |
| see above | Lifebrain Covid Labor GmbH | Lifebrain Covid Labor GmbH | Abhishek Mitra; Alexandra Wagner; Anna Edermayr; Felix Valentin Spiegel; Filip Sima; Florian Scharhauser; Hannes Hagen; Kristina Bavrka Kolenc; Lucia Castello; So Jung Han |
| EPI_ISL_12828082 | Limbach - MVZ Humangenetik Ulm | Robert Koch Institute | |
| EPI_ISL_12675667, EPI_ISL_13247268 | Limbach - MVZ Labor Dr. Volkmann & Kollegen | Robert Koch Institute | |
| EPI_ISL_12675694 | Limbach - MVZ Labor Westmecklenburg Schmudlach-Oswald-Kettermann & Kollegen | Robert Koch Institute | |
| EPI_ISL_13169074 | MARIANO MARCOS MEMORIAL HOSPITAL AND MEDICAL CENTER | Philippine Genome Center | Alethea R. de Guzman; Alyssa Joyce E. Telles; Anna Ong-Lim; Arianne A. Zamora; Benedict A. Maralit; Carlo M. Lapid; Celia Carlos; Cynthia P. Saloma; Devon Ray Pacial; Diomedes A. Cariño; Edsel Maurice Salvana; El King D. Morado; Elcid Aaron R. Pangilinan; Eva Maria Cutiongco-de la Paz; Francis A. Tablizo; Henrietta Marie Rodriguez; Jaime C. Montoya; Jan Michael C. Yap; Jarvin E. Nipales; Jo-Hannah S. Llames; John Michael Egana; John Q. Wong; Joshua Gregor A. Dizon; Joshua Jose Endozo; Juan Antonio R. Magalang; Karol Sophia Agape R. Padilla; Kris P. Punayan; Kristina Patriz Dela Cruz; Lindsay Clare D.L. Carandang; Ma. Exanil Plantig; Marc Edsel C. Ayes; Maria Rosario Singh-Vergeire; Maria Sofia L. Yangzon; Marielle M. Gamboa; Marissa Alejandria; Niña Francesca Bustamante; Razel Nikka M. Hao; Renato Jacinto Q. Mantaring; Rianna Patricia S. Cruz; Shiela Mae M. Araiza; Yvonne Valerie Austria; Zipporah Mariebelle R. Enriquez; Zyrel V. Mollejon |
| EPI_ISL_13086361 | MEPHI, Aix Marseille University | MEPHI, Aix Marseille University | Anthony LEVASSEUR |
| EPI_ISL_13001778, EPI_ISL_13241339 | MVZ Dr. Eberhard & Partner Dortmund | Robert Koch Institute | |
| EPI_ISL_13265071 | MVZ Labor Dr. Fenner und Kollegen (Standort Hamburg) | Robert Koch Institute | |
| EPI_ISL_12845818 | Maine Health and Environmental Testing Laboratory | Tewhey Lab, The Jackson Laboratory | Barter, M.; Dewey, H.; H. and Tewhey, R.; Iosue, F.; Lynch, R.; Matluk, N.; Munger |
| EPI_ISL_13235637, EPI_ISL_13235674 | Med. Labor Prof. Schenk Dr. Ansorge & Kollegen | Robert Koch Institute | |
| EPI_ISL_13202446 | Medical Microbiology Unit, Department for Laboratory Medicine, Drammen Hospital, Vestre Viken Health Trust | Norwegian Institute of Public Health, Department of Virology | Atiya R Ali; Debech Nadia; Engebretsen Serina Beate; Garcia Llorente Ignacio; Hilde Elshaug; Hilde Nordby Falkenhaug; Hilde Vollan; Jon Bråte; Kamilla Heddeland Instefjord; Karoline Bragstad; Kathrine Stene-Johansen; Line Victoria Moen; Marie Paulsen Madsen; Olav Hungnes; Pedersen Benedikte Nevjen; Rasmus Riis Kopperud |
| EPI_ISL_13028110, EPI_ISL_13312801, EPI_ISL_13312812, EPI_ISL_13312813 | Microvida | Microvida | Jaco J. Verweij; Joep J. J. M. Stohr; Suzan D. Pas |
| EPI_ISL_13253591 | NZOZ Białostockie Centrum Analiz Medycznych Sp. z o o | 1. Academic Center for Pathomorphological and Genetic-Molecular Diagnostics ltd, Bialystok, Poland 2. National Institute of Public Health - National Institute of Hygiene, Warsaw, Poland | Anetta Sulewska; Jacek Nikliński; Janusz Dzięcioł; Joanna Kiśluk; Katarzyna Zacharczuk; Konrad Raczkowski; Magdalena Nowakowska; Małgorzata Sadkowska-Todys; Piotr Karabowicz; Piotr Majewski; Przemysław Biecek. Joanna Reszeć; Radosław Charkiewicz; Tomasz Wołkowicz |
| EPI_ISL_12208048 | National Platform bis COVID ULB-IBC | National Platform bis COVID ULB-IBC | Arnaud Marchant; Benoit Haerlingen; Coralie Henin; Marie-Luce Delforge; Ricardo De Mendonça |
| EPI_ISL_12646114, EPI_ISL_12647046, EPI_ISL_12647174, EPI_ISL_12647189, EPI_ISL_13094097, EPI_ISL_13273959, EPI_ISL_13273968, EPI_ISL_13273971 | | | |
| see above | National Public Health Laboratory, National Centre for Infectious Diseases | National Public Health Laboratory, National Centre for Infectious Diseases | BeiBei Chen; Benny Yeo; Chen Shi Ling; Grace Ngan; Jesslin Tan; Lin Cui; Raymond Tzer Pin Lin; Royce Ang; Samuel Loo; Yichen Ding; Zhenyang Zhou |
| EPI_ISL_13186686, EPI_ISL_13252570, EPI_ISL_13252571, EPI_ISL_13252573, EPI_ISL_13252574, EPI_ISL_13252586, EPI_ISL_13252589, EPI_ISL_13252619, EPI_ISL_13252661, EPI_ISL_13252669, EPI_ISL_13252677, EPI_ISL_13252704, EPI_ISL_13252726, EPI_ISL_13252740, EPI_ISL_13252747, EPI_ISL_13252754, EPI_ISL_13252765, EPI_ISL_13252891, EPI_ISL_13298765, EPI_ISL_13298766, EPI_ISL_13298792, EPI_ISL_13298805, EPI_ISL_13298868, EPI_ISL_13298906 | | | |
| see above | National Virus Reference Laboratory | National Virus Reference Laboratory | Charlene Bennett; Cillian F De Gascun; Gabriel Gonzalez; Jonathan Dean; Michael Carr; Zoe Yandle |
| EPI_ISL_13251086 | NutriMedlab | National Institute of Public Health | Alexander Nagy; Helena Jirincova; Jan Moskalyk; Jaromira Vecerova; Timotej Suri |
| EPI_ISL_12983026, EPI_ISL_13180979, EPI_ISL_13181084 | Oslo University Hospital, Department of Microbiology | Norwegian Institute of Public Health, Department of Virology | Arvind Yegambaram Meenakshi Sundaram; Cathrine Fladeby; Garcia Llorente Ignacio; Gregor D. Gilfillan; Hilde Elshaug; Hilde Vollan; Jon Bråte; Kamilla Heddeland Instefjord; Karoline Bragstad; Kathrine Stene-Johansen; Line Victoria Moen; Lise Andresen; Mariann Nilsen; Mona Holberg-Petersen; Olav Hungnes; Pedersen Benedikte Nevjen; Pål Marius Bjørnstad; Rasmus Riis Kopperud; Teodora Plamenova Ribarska |
| EPI_ISL_13291713 | PR Public Health Lab | Centers for Disease Control and Prevention Division of Viral Diseases, Pathogen Discovery | Alex Burgin; Ben Rambo-Martin; Clinton Paden; Dakota Howard; Dave Wentworth; Dhwani Batra; Jasmine Padilla; Joseph Madden; Justin Lee; Kristen Knipe; Kristine Lacek; Mark Burroughs; Matthew Schmerer; Meghan Bentz; Mili Sheth; Peter Cook; Sam Shepard; Sarah Nobles; Vivien Dugan; Yvette Unoarumhi |
| EPI_ISL_12859909, EPI_ISL_13042826, EPI_ISL_13043400, EPI_ISL_13121528, EPI_ISL_13256963 | Pandemic Response Lab - NYC | Pandemic Response Lab, R&D | Alex Carpio; Cybill del Castillo; Haiping Hao; Isabel Fernandez Escapa; Jon Laurent; Melissa Hopkins; Michael Hammerling; Simran Chhabria; Simran Gupta; Sol Rey; Steven Chase; Tiara Rivera; William Ward |
| EPI_ISL_13040764 | Pathology Queensland and Forensic Scientific Services | Public Health Virology - Forensic and Scientific Services (PHV-FSS) | Chenwei Wang on behalf of Q-PHIRE Genomics |
| EPI_ISL_13103406, EPI_ISL_13104100 | Provincial Laboratory for Public Health (ProvLab) - North | Alberta Precision Labs (APL) | Buss E; Croxen M; Deo A; Dieu P; Ferrato C; Gill K; Koleva P; Li V; Lloyd C; Lynch T; Ma R; Murphy S; Pabbaraju K; Shideler S; Shokopoles S; Skitsko T; Thayer J; Tipples G; Wong A; Yu C; Zelyas N. |
| EPI_ISL_12869449, EPI_ISL_12869477, EPI_ISL_12869500, EPI_ISL_13044485, EPI_ISL_13132054, EPI_ISL_13132085, EPI_ISL_13271194, EPI_ISL_13271216, EPI_ISL_13271246 | | | |
| see above | Public Health Laboratory, Public Health Service Amsterdam, The Netherlands | Department of Medical Microbiology & Infection prevention, Amsterdam University Medical Centers location AMC | Akke Cornelissen; Fokla Zorgdrager; Gini van Rijckevorsel; Janke Schinkel; Jelle Koopsen; Judith den Uil; Marcel Jonges; Matthijs Welkers; Menno de Jong; Menno de Jong and Mariken van der Lubben on behalf of the Amsterdam Regional Genomic epidemiology and Outbreak Surveillance (ARGOS) consortium; Patrick Habermehl; Robin van Houdt; Sebastien Matamoros; Sjoerd Rebers; Sylvia Bruisten; Tjalling Leenstra and Mariken van der Lubben on behalf of the Amsterdam Regional Genomic epidemiology and Outbreak Surveillance (ARGOS) consortium |
| EPI_ISL_13111253 | Public health laboratories Jerusalem | Israel Central Virology laboratory | Danit Sofer; Efrat Dahan Bucris; Ella Mendelson; Evan Nachum; Hagar Morad; Julia Vainer; Maya Davidovich; Michal Mandelboim; Michal Zak; Miranda Geva; Neta Zuckerman; Or Zilbertzan; Oran Erster; Orna Mor; Rona Grossman |
| EPI_ISL_12582173, EPI_ISL_12583321, EPI_ISL_13001342, EPI_ISL_13028243, EPI_ISL_13048796, EPI_ISL_13066298, EPI_ISL_13066330, EPI_ISL_13066345, EPI_ISL_13107947, EPI_ISL_13108332, EPI_ISL_13131454, EPI_ISL_13201660, EPI_ISL_13201824, EPI_ISL_13244003, EPI_ISL_13244065, EPI_ISL_13244115, EPI_ISL_13244149, EPI_ISL_13244180, EPI_ISL_13244230 | | | |
| see above | Respiratory Virus Unit, Microbiology Services Colindale, Public Health England | COVID-19 Genomics UK (COG-UK) Consortium | PHE Covid Sequencing Team |
| EPI_ISL_13242611, EPI_ISL_13242616 | Rosalind Franklin Laboratory | Wellcome Sanger Institute for the COVID-19 Genomics UK (COG-UK) Consortium | Cordelia Langford; David K. Jackson; Dominic Kwiatkowski; Donald Fraser; Ewan Harrison; Ian Johnston; Jeffrey Barrett; John Sillitoe on behalf of the Wellcome Sanger Institute COVID-19 Surveillance Team; Rob Howes; Roberto Amato; Sonia Goncalves; Suki Lee; The Rosalind Franklin Laboratory and Alex Alderton |
| EPI_ISL_13282978 | S.C. Laboratorio Analisi, Ospedale di Lavagna, ASL 4 Liguria | U.O. Igiene, Ospedale Policlinico San Martino | Bandettini Roberto; Bruzzone Bianca; De Pace Vanessa; Domnich Alexander; Icardi Giancarlo on behalf of SARS-CoV-2 ITALIAN RESEARCH ENTERPRISE-(SCIRE) Collaborative Group; Orsi Andrea; Randazzo Nadia; Ricucci Valentina; Stefanelli Federica |
| EPI_ISL_13292599, EPI_ISL_13293034, EPI_ISL_13300919, EPI_ISL_13300921 | SARS-CoV-2 Sequencing Castilla y Leon-Spain Consortium | SARS-CoV-2 Sequencing Castilla y Leon-Spain Consortium | Antonio Orduña-Domingo; Carlos Fuster Foz; Carmen Aldea-Mansilla; Carmen Gimeno Crespo; David Abad; Gabriel March Rosello; Gregoria Megías Lobón; Jose María Eiros Bouza; M. Isabel Fernandez-Natal; Marta Dominguez-Gil; Marta Hernandez; María Antonia García Castro; Mª Fe Brezmes-Valdivieso; Noelia Arenal Andrés; Silvia Rojo; Sonsoles Garcinuño Pérez |

| | | | |
|---|---|---|---|
| EPI_ISL_12806962, EPI_ISL_12807012, EPI_ISL_12807046, EPI_ISL_12807091, EPI_ISL_12807162, EPI_ISL_13027401, EPI_ISL_13027571, EPI_ISL_13027577, EPI_ISL_13027612, EPI_ISL_13217440, EPI_ISL_13217505, EPI_ISL_13217673, EPI_ISL_13217703 | | | |
| see above | SARS-CoV-2 testing team, National Institute of Infectious Diseases | Pathogen Genomics Center, National Institute of Infectious Diseases | Hazuka Y Furihata; Kentaro Itokawa; Makoto Kuroda; Masanori Hashino; Masumichi Saito; Naomi Nojiri; Nozomu Hanaoka; Rina Tanaka; Tsuguto Fujimoto; Tsuyoshi Sekizuka |
| EPI_ISL_10943953 | SMS MEDICAL COLLEGE,JAIPUR | NIV Influenza | Bharti Malhotra; Dinesh parsoya; Farah Deeba; Himanshu sharma; Neha Bhomia; Nita pal; Nivedita Gupta; Pragya D Yadav; Pratibha Sharma; Sudhir Bhandari; Swati Gautam; Varsha Potdar |
| EPI_ISL_12903141 | SYNLAB | University Hospital Brno, CMBG | Bezdicek Matej; Dolejska Monika; Kristyna Dufkova; Lengerova Martina; Svaton Jan |
| EPI_ISL_11312517, EPI_ISL_12902035, EPI_ISL_13112098 | Shamir Medical Center (Asaf Harofe) | Shamir Medical Center (Asaf Harofe) | Abu Hamad Ramzia; Adina Bar Chaim; Alona Frenkel; Anna Vishnevsky; Chen Weiner; Nir Rainy; Patricia Benveniste-Lekovitz; Reut Sorek Abramovich; Yevgeni Yegorov |
| EPI_ISL_13040526 | Sullivan Nicolaides Pathology | Public Health Virology - Forensic and Scientific Services (PHV-FSS) | Chenwei Wang on behalf of Q-PHIRE Genomics |
| EPI_ISL_13066241, EPI_ISL_13133873 | Synlab Eesti OÜ | 1. Laboratory of Communicable Diseases (Estonia); 2. Eurofins Genomics Europe Sequencing GmbH | Abroi A.; Avi R.; Dotsenko L.; Epštein J.; Hoidmets D.; Huik K.; Härma M-A.; Jaaniso E.; Kaarna K.; Kallas E.; Koppel I.; Kuzmin I.; Lahesaare A.; Lutsar I.; Metspalu M.; Milani L.; Naaber P.; Niglas H.; Oopkaup O.E.; Pauskar M.; Peterson H.; Päll T.; Ratnik K.; Raudvere U.; Reisberg T.; Sadikova O.; Sepp H.; Shablinskaja A.; Suija H.; Talas U.G.; Truusalu K. |
| EPI_ISL_13198051 | Szpital Wojewodzki im. KardynaLa Stefana Wyszynskiego w Lomży | 1. Academic Center for Pathomorphological and Genetic-Molecular Diagnostics ltd, Bialystok, Poland 2. National Institute of Public Health - National Institute of Hygiene, Warsaw, Poland | Anetta Sulewska; Jacek Niklinski; Janusz DzięcioL; Joanna Kiśluk; Katarzyna Zacharczuk; Konrad Raczkowski; MaLgorzata Sadkowska-Todys; Magdalena Nowakowska; Piotr Karabowicz; Piotr Majewski; PrzemysLaw Biecek. Joanna Reszeć; RadosLaw Charkiewicz; Tomasz WoLkowicz |
| EPI_ISL_12980664, EPI_ISL_13273628 | Tokyo Metropolitan Institute of Public Health | Tokyo Metropolitan Institute of Public Health | Ai Suzuki; Akane Negishi; Arisa Amano; Fumi Kasuya; Hirofumi Miyake; Kenji Sadamasu; Kenshirou Kuroki; Mami Nagashima; Maya Isogai; Ryota Kumagai; Sachiko Harada; Takushi Fujiwara |
| EPI_ISL_12870115, EPI_ISL_13223359, EPI_ISL_13223378 | U.O. Microbiologia Laboratorio Unico Centro Servizi - AUSL della Romagna | U.O. Microbiologia, Laboratorio Unico Centro Servizi - AUSL della Romagna | Giorgio Dirani |
| EPI_ISL_12825301, EPI_ISL_12825326 | UMC Groningen, Clinical Virology, Department of Medical Microbiology and Infection Prevention | UMC Groningen, Clinical Virology, Department of Medical Microbiology and Infection Prevention | Alexander Friedrich; Coretta Van Leer-Buter; Erley Lizarazo-Forero; Hubert Niesters; Lilli Gard; Marjolein Knoester; Monika Fliss; Sigrid Rosema; Xuewei Zhou |
| EPI_ISL_13046845, EPI_ISL_13103105 | UW Virology Lab | UW Virology Lab | Alexander Greninger; Hong Xie; Isabel Arnould; Keith R Jerome; Pavitra Roychoudhury; Pooneh Hajian; Sarai Luna; Saraswathi Sathees; Sean Ellis; Seffir T. Wendm; Shah Mohamed Bakhash |
| EPI_ISL_13018126, EPI_ISL_13018127, EPI_ISL_13018142 | Unidad de Investigación Médica de Yucatán (UIMY) | Microbial Genomics Laboratory | ; Alejandra García-Gasca; Alejandra Hernández-Terán; Alejandro Sánchez-Flores; Alfredo Herrera-Estrella; Alicia Ocaña-Mondragón; Andreu Comas-García; Angel Gustavo Salas-Lais; Antonio Loza Román; Bernardo Martínez-Miguel; Blanca Taboada; Brenda Irasema Maldonado-Meza; Bruno Gómez-Gil; Carla Ivón Herrera-Najera; Carlos F. Arias; Celia Boukadida; Clara Esperanza Santacruz-Tinoco; Concepción Grajales-Muñiz; Consorcio Mexicano de Vigilancia Genómica (CoViGen-Mex). Authors (in alphabetical order): Julio Elias Alvarado-Yaah; Cristóbal Cháidez-Quiróz; Célida Duque Molina; Célida Martínez- Rodríguez; Daniel Fregoso-Rueda; Daniel Lira Morales; Eduardo Becerril-Vargas; Fernando Fontove-Herrera; Fidencio Mejía-Nepomuceno; Francisco Pulido; Gloria Elena Espinosa-Ayala; Gloria María Molina-Salinas; Gloria Vazquez; Hector Esteban Paz-Juárez; Hector Montoya-Fuentes; Helen Haydee Fernanda Ramírez-Plascencia; Irvin González-López; Jean Pierre González; Jesús Hernández; Joel Armando Vázquez-Pérez.; Jorge Salas-Hernández; José Antonio Enciso-Moreno; José Arturo Martínez-Orozco; José Esteban Muñoz-Medina; José de Jesús Nuñez-Contreras; Juan Bautista Chale-Dzul; Julissa Enciso-Ibarra; Luis Alberto Ochoa-Carrera; Margarita Matías-Florentino; Mario Mújica-Sánchez; Marissa Perez-Garcia; María Guadalupe Santiago-Mauricio; María Guadalupe de Jesús Mireles-Rivera; Nelly Sélem-Mojica; Pavel Isa; Ricardo Ciria Merce; Ricardo Grande; Rosa María Gutiérrez Rios; Santiago Ávila-Ríos; Selene Zárate; Susana Lopez; Verónica Mata-Haro; Victor Eduardo García-Arias; Víctor Hugo Borja-Aburto |
| EPI_ISL_12983143 | University Hospital of Northern Norway, Department for Microbiology and Infectious Disease Control | Norwegian Institute of Public Health, Department of Virology | Atiya R Ali; Debech Nadia; Engebretsen Serina Beate; Garcia Llorente Ignacio; Hilde Elshaug; Hilde Nordby Falkenhaug; Hilde Vollan; Jon Bråte; Kamilla Heddeland Instefjord; Karoline Bragstad; Kathrine Stene-Johansen; Line Victoria Moen; Marie Paulsen Madsen; Olav Hungnes; Pedersen Benedikte Nevjen; Rasmus Riis Kopperud |
| EPI_ISL_13182714 | Vestfold Hospital, Toensberg, Department of Microbiology | Norwegian Institute of Public Health, Department of Virology | Atiya R Ali; Debech Nadia; Engebretsen Serina Beate; Garcia Llorente Ignacio; Hilde Elshaug; Hilde Nordby Falkenhaug; Hilde Vollan; Jon Bråte; Kamilla Heddeland Instefjord; Karoline Bragstad; Kathrine Stene-Johansen; Line Victoria Moen; Marie Paulsen Madsen; Olav Hungnes; Pedersen Benedikte Nevjen; Rasmus Riis Kopperud |
| EPI_ISL_12737315, EPI_ISL_13133561 | ZOL | Jessa | Laura Vanstraelen et al. on behalf of the Jessa_cmdLab; Rita Smets et al. on behalf of the Jessa_cmdLab |
| EPI_ISL_12879758 | ZOTZ KLIMAS MVZ Düsseldorf-Centrum GbR ÜBAG für Labormedizin, Genetik, Zytologie, Pathologie | Center of Medical Microbiology, Virology, and Hospital Hygiene, University of Duesseldorf | Alexander Dilthey; Andreas Walker; Daniel Strelow; Jessica Nicolai; Jörg Timm; Katrin Hoffmann; Klaus Pfeffer; Lisanna Hülse; Malte Kohns Vasconcelos; Maximilian Damagnez; Nadine Lübke; Patrick Finzer; Rainer Zotz; Tobias Wienemann; Torsten Houwaart |
| EPI_ISL_13238014, EPI_ISL_13268353 | labopart - Medizinische Laboratorien Dresden | Robert Koch Institute | |
