## Supplementary material for "Tracing the international arrivals of SARS-CoV-2 Omicron variants after Aotearoa New Zealand reopened its border": BA.2 GISAID acknowledgements

We gratefully acknowledge the following Authors from the Originating laboratories responsible for obtaining the specimens, as well as the Submitting laboratories where the genome data were generated and shared via GISAID, on which this research is based.

All Submitters of data may be contacted directly via www.gisaid.org

Authors are sorted alphabetically.

| Accession ID | Originating Laboratory | Submitting Laboratory | Authors |
|---|---|---|---|
| EPI_ISL_12177518, EPI_ISL_12570911 | A.O. PERUGIA | A.O. PERUGIA | Bicchieraro G; Bondi P; Camilloni B; Cappelletti E; Ciurnelli R; Lepri E; Lucheroni F; Mencacci A; Spaccapelo R |
| EPI_ISL_12838823 | ALIFE HEALTH | Instituto Nacional de Salud-Dirección de Investigación en Salud Pública | Beatriz de Arco; Carlos Franco-Muñoz; Diego A. Álvarez-Díaz; Diego Andrés Prada; Dioselina Peláez-Carvajal; Gerardo Santamaría; Héctor Alejandro Ruiz-Moreno; Jhonnatan Reales-González; Jorge Rivera; Julián Naizaque; Katherine Laiton-Donato; Marcela Mercado-Reyes.; Martha Lucia Ospina Martínez; María T. Herrera-Sepúlveda; Paola Rojas-Estevez; Sheryll Corchuelo; Tatiana Cobos |
| EPI_ISL_13059818, EPI_ISL_13059823, EPI_ISL_13059835 | ALIFE HEALTH | Instituto de Investigación de Recursos Biológicos Alexander von Humboldt | Daniel Martínez Vargas; Mailyn Gonzalez; Nicolás D. Franco-Sierra; Vanessa Otero-Jiménez |
| EPI_ISL_12832931 | AREA DE SALUD BARVA | Inciensa, Instituto Costarricense de Investigación y Enseñanza en Nutrición y Salud | Adriana Godínez; Claudio Soto-Garita; Estela Cordero; Francisco Duarte; Gabriel Morales; Hebleen Porras; José Luis Vargas; Mariela Gutiérrez; Melany Calderón; Natalia Bonilla & Sharon Peñaranda Chanto; Sofia Herrera |
| EPI_ISL_13109480 | AREA DE SALUD COTO BRUS | Inciensa, Instituto Costarricense de Investigación y Enseñanza en Nutrición y Salud | Adriana Godínez; Claudio Soto-Garita; Estela Cordero; Francisco Duarte; Gabriel Morales & Natalia Bonilla; Hebleen Porras; José Luis Vargas; Mariela Gutiérrez; Melany Calderón; Sofia Herrera |
| EPI_ISL_12935269 | AREA DE SALUD SANTA CRUZ | Inciensa, Instituto Costarricense de Investigación y Enseñanza en Nutrición y Salud | Adriana Godínez; Claudio Soto-Garita; Estela Cordero; Francisco Duarte; Gabriel Morales; Hebleen Porras; José Luis Vargas; Mariela Gutiérrez; Melany Calderón; Natalia Bonilla & María José Villegas Bermúdez; Sofia Herrera |
| EPI_ISL_12784048, EPI_ISL_12784049 | AZ St.Jan Brugge-Oostende | AZ St.Jan Brugge-Oostende | Jorn Hellemans; Marie Madeleine Chabert-Consen; Marijke Reynders; Melissa Provoost; Merijn Vanhee; Sofie Mahboob |
| EPI_ISL_9829939, EPI_ISL_9829944, EPI_ISL_9829952, EPI_ISL_9829955, EPI_ISL_9829964, EPI_ISL_10104461, EPI_ISL_10407059, EPI_ISL_10407068, EPI_ISL_10993984, EPI_ISL_10993999, EPI_ISL_10994012, EPI_ISL_10994016, EPI_ISL_10994033, EPI_ISL_10994034, EPI_ISL_10994039 | | | |
| see above | Allergy, Immunology and Cell Biology Unit (AICBU) | Allergy, Immunology and Cell Biology Unit (AICBU) | Ayesha Wijesinghe; Chandima Jeewandara; Deshni Jayathilaka; Dinuka Ariyaratne; Diyanath Ranasinghe; Dumni Guasinghe; Farha Bary; Gathsaurie Neelika Malavige; Heshan Kuruppu; Tibutius Thanesh |
| EPI_ISL_13337990 | Area De Salud Colorado | Inciensa, Instituto Costarricense de Investigación y Enseñanza en Nutrición y Salud | Adriana Godínez; Claudio Soto-Garita; Estela Cordero; Francisco Duarte; Gabriel Morales; Hebleen Porras; José Luis Vargas; Mariela Gutiérrez; Melany Calderón; Natalia Bonilla & Shirley Rojas Ramírez; Sofia Herrera |
| EPI_ISL_13337847 | Area de salud corredores | Inciensa, Instituto Costarricense de Investigación y Enseñanza en Nutrición y Salud | Adriana Godínez; Claudio Soto-Garita; Estela Cordero; Francisco Duarte; Gabriel Morales; Hebleen Porras; José Luis Vargas; Mariela Gutiérrez; Melany Calderón; Natalia Bonilla & Gabriela Marchena Jimenez; Sofia Herrera |
| EPI_ISL_13337834 | Area de salud la cruz | Inciensa, Instituto Costarricense de Investigación y Enseñanza en Nutrición y Salud | Adriana Godínez; Claudio Soto-Garita; Estela Cordero; Francisco Duarte; Gabriel Morales & Natalia Bonilla; Hebleen Porras; José Luis Vargas; Mariela Gutiérrez; Melany Calderón; Sofia Herrera |
| EPI_ISL_11852963 | Associazione Donatori di Sangue Teramo | Istituto Zooprofilattico Sperimentale dell'Abruzzo e Molise "G. Caporale" | Ancora M; Calistri P; Cammà C; Caporale M; Curini V; Delli Compagni E; Di Domenico M; Di Lollo Valeria; Di Pasquale A; Lorusso A; Mangone I; Marcacci M; Puglia I; Rinaldi A; Savini G; Scialabba S |
| EPI_ISL_11073123, EPI_ISL_12024284, EPI_ISL_12313884, EPI_ISL_12650435, EPI_ISL_12727797 | Austrian Agency for Health and Food Safety (AGES) | Bergthaler laboratory, CeMM Research Center for Molecular Medicine of the Austrian Academy of Sciences | Alberto Alises; Andreas Bergthaler; Anna Schedl; Bekir Erguner; Benedikt Agerer; Christoph Bock; Fabian Amman; Jan Laine; Lukas Endler; Martin Senekowitsch; Matthew Thornton; Michael Schuster; Michelle Chan; Petr Triska; Thomas Penz |
| EPI_ISL_11269099 | Aversi Clinic | Department for Virology, Molecular Biology and Genome Research, R. G. Lugar Center for Public Health Research, National Center for Disease Control and Public Health (NCDC) of Georgia. | Adam Kotorashvili; Amiran Gamkrelidze.; Ana Papkiauri; Ann Machablishvili; Anna Kasradze; Davit Tsaguria; Ekaterine Khmaladze; Ekaterine Zangaladze; Ekaterine Zhghenti; Giorgi Gogoladze; Giorgi Tomashvili; Gvantsa Brachveli; Gvantsa Chanturia; Irma Burjanadze; Ketevan Sidamonidze; Khatuna Zakhashvili; Lela Sabadze; Lela Urushadze; Magda Dgebuadze; Maia Alkhazashvili; Mari Gavashelidze; Mariam Zakalashvili; Marine Murtskhvaladze; Meri Pantsulaia; Nato Kotaria; Nino Berishvili; Paata Imnadze; Roena Sukhiashvili; Salome Javashvili; Tamar Jashiashvili; Tata Imnadze; Tea Tevdoradze |
| EPI_ISL_12946141, EPI_ISL_13100329 | Basurto University Hospital: Clinical Microbiology Laboratory | Basurto University Hospital: Clinical Microbiology Laboratory | Estibaliz Ugalde Zarraga; José Luis Díaz de Tuesta del Arco; Mikel Urrutikoetxea-Gutiérrez; Mª Carmen Nieto Toboso |
| EPI_ISL_10969316 | Biomedical Centre Martin, Jessenius Faculty of Medicine in Martin, Comenius University | Laboratory of Genomics and Bioinformatics, Comenius University Science Park | Andrea Hornáková; Anna Kaliňáková; Barbora Kotvasová; Diana Rusňáková; Dušan Loderer; Ivana Kašubová; Jaroslav Budiš; Katarína Janíková; Lucia Ševčíková; Marián Grendár; Miroslav Böhmer; Mária Škereňová; Pavol Mišenko; Terézia Vrabľová; Tomáš Szemes |
| EPI_ISL_10442995 | Botswana Harvard HIV Reference Laboratory | Botswana Harvard HIV Reference Laboratory | Boitumelo Zuze; Dorcas Maruapula; Joseph Makhema; Kgomotso Moruisi; Legodile Kooepile; Mosepele Mosepele; Mphaphi B. Mbulawa; Ontlametse T. Bareng; Pamela Smith-Lawrence; Roger Shapiro; Sefetogi Ramaologa; Shahin Lockman; Shirley Johane; Sikhulile Moyo; Simani Gaseitsiwe; Thongbotho Mphoyakgosi; Wonderful T. Choga |
| EPI_ISL_13168976, EPI_ISL_13168981 | CENTRO DE ESPECIALIDADES DERMATOLÓGICAS | Laboratorio Central de Salud Pública | Analía Rojas; Andrea Gómez de la Fuente; Cynthia Vazquez; César Cantero; César Ojeda; Emmanuel Céspedes; Fátima Fleitas; Ivana Fernández; Juan Torales; Julio Barrios; Maria Liz Gamarra; María Jose Duarte; Sandra González; Shirley Villalba; Tania Alfonzo; Wilson Benítez. |
| EPI_ISL_12285404 | CERBALLIANCE ARTOIS | CERBA HealthCare | Bénédicte Roquebert; Laura Verdurme; Mathilde Roussel; Sabine Trombert; Stéphanie Haim-Boukobza |
| EPI_ISL_9550429 | COVID-19 Detection Lab, Chattogram Veterinary and Animal Sciences University | COVID-19 Detection Lab, Chattogram Veterinary and Animal Sciences University | 56. Tanvir Ahmad Nizami; Barna Goswami; Eaftekhar Ahmed Rana; Goutam Buddha Das; Iffat Jahan; Md. Ahasan Habib; Md. Murshed Hasan Sarkar; Md. Salim Khan; Md. Sirazul Islam; Mohammad Mohi Uddin; Paritosh Kumar Biswas; Pronesh Dutta; Sanjana Fatema Chowdhury; Shahina Akter; Sharmin Chowdhury; Showti Raheel Nase; Tanjina Akhter Banu; Tridip Das |
| EPI_ISL_12279304, EPI_ISL_12428361, EPI_ISL_12693978 | COVID-19 Diagnostic Lab, BADAS | Genomic Research Laboratory, BSMMU | Laila Anjuman Banu; Mahmud Hossain; Md.Sharfuddin Ahmed; Nahid Azmin; Zahid Hassan |
| EPI_ISL_12607716, EPI_ISL_12607741, EPI_ISL_12607756, EPI_ISL_12607779, EPI_ISL_12607813 | CPHL,MOH,EGYPT | CPHL | Abd Monaem Adel; Amel nagiub; Amer Sayed; Amr Kandil.; Dalia Ramadan; Galal Mahmoud; Marwa Abd EL fatah; Marwa Saleh; Mohamed Hassany; Mohamed Kamal; Nancy el guindy; Ramy Galal; Rasha Zaater; Shymaa s. Ahmed; Wael H. Roshdy; salma sayed |
| EPI_ISL_13134340 | Cantacuzino National Military-Medical Institute, Viral Respiratory Infections Laboratory | Cantacuzino National Military-Medical Institute, Viral Respiratory Infections Laboratory | Catalina Pascu; Luiza Ustea; Mihaela Lazar; Mihaela Oprea; Sorin Dinu; Vitencu Oana |
| EPI_ISL_12024394, EPI_ISL_12144619, EPI_ISL_12896424 | Center for Virology, Medical University of Vienna | Bergthaler laboratory, CeMM Research Center for Molecular Medicine of the Austrian Academy of Sciences | Alberto Alises; Andreas Bergthaler; Anna Schedl; Christoph Bock; Fabian Amman; Lukas Endler; Matthew Thornton; Michael Schuster; Michelle Chan; Petr Triska |
| EPI_ISL_11698098, EPI_ISL_11698102 | Center of Excellence in Clinical Virology, Faculty of Medicine, Chulalongkorn University | Center of Excellence in Clinical Virology, Faculty of Medicine, Chulalongkorn University | Amornmas Kongkliang; Jira Chansaenroj; Jiratchaya Puenpa; Kamolthip Atsawawaranunt; Nutsada Saengdao; Oraphan Mayuramart; Pakkaporn Panwijitkul; Pattaraporn Nimsamer; Patthaya Rattanakomol; Sunchai Payungporn; Suwanna Petto; Vichan Pawun; Vorthon Sawaswong; Yong Poovorawan |
| EPI_ISL_13332960, EPI_ISL_13333002, EPI_ISL_13333011, EPI_ISL_13333023, EPI_ISL_13333027, EPI_ISL_13333037 | Centers for Disease Control and Prevention, Dengue Branch | Centers for Disease Control and Prevention, Dengue Branch | Betzabel Flores; Gabriela Paz-Bailey; Gilberto A. Santiago; Glenda Gonzalez; Jorge L. Munoz-Jordan; Keyla Charriez |
| EPI_ISL_13003004 | Centre Hospitalier Universitaire Clermont-Ferrand | CHU Clermont-Ferrand, service de virologie | Bisseux Maxime; Combes Patricia; Henquell Cecile; Mirand Audrey |
| EPI_ISL_13350769 | Centro Medico La Costa | Laboratorio Central de Salud Pública | Analía Rojas; Andrea Gómez de la Fuente; Cynthia Vazquez; César Cantero; César Ojeda; Emmanuel Céspedes; Fátima Fleitas; Ivana Fernández; Juan Torales; Julio Barrios; Maria Liz Gamarra; María Jose Duarte; Sandra González; Shirley Villalba; Tania Alfonzo; Wilson Benítez. |

| | | | |
|---|---|---|---|
| EPI_ISL_12934866 | Centro de Investigación Biomédica del Noreste (CIBIN) | Instituto de Biotecnología de la UNAM | ; Alejandra García-Gasca; Alejandra Hernández-Terán; Alejandro Sánchez-Flores; Alfredo Herrera-Estrella; Alicia Ocaña-Mondragón; Andreu Comas-García; Angel Gustavo Salas-Lais; Antonio Loza Román; Bernardo Martínez-Miguel; Blanca Taboada; Brenda Irasema Maldonado-Meza; Bruno Gómez-Gil; Carla Ivón Herrera-Najera; Carlos F. Arias; Celia Boukadida; Clara Esperanza Santacruz-Tinoco; Concepción Grajales-Muñiz; Consorcio Mexicano de Vigilancia Genómica (CoViGen-Mex). Authors (in alphabetical order): Julio Elias Alvarado-Yaah; Cristóbal Cháidez-Quiróz; Célida Duque Molina; Célida Martínez- Rodríguez; Daniel Fregoso-Rueda; Daniel Lira Morales; Eduardo Becerril-Vargas; Fernando Fontove-Herrera; Fidencio Mejía-Nepomuceno; Francisco Pulido; Gloria Elena Espinosa-Ayala; Gloria María Molina-Salinas; Gloria Vazquez; Hector Esteban Paz-Juárez; Hector Montoya-Fuentes; Helen Haydee Fernanda Ramírez-Plascencia; Irvin González-López; Jean Pierre González; Jesús Hernández; Joel Armando Vázquez-Pérez).; Jorge Salas-Hernández; José Antonio Enciso-Moreno; José Arturo Martínez-Orozco; José Esteban Muñoz-Medina; José de Jesús Nuñez-Contreras; Juan Bautista Chale-Dzul; Julissa Enciso-Ibarra; Kathia Elizabeth Tapia-Diaz; Luis Alberto Ochoa-Carrera; Margarita Matías-Florentino; Mario Mújica-Sánchez; Marissa Perez-Garcia; María Guadalupe Santiago-Mauricio; María Guadalupe de Jesús Mireles-Rivera; Nelly Sélem-Mojica; Pavel Isa; Ricardo Ciria Merce; Ricardo Grande; Rosa María Gutiérrez Rios; Santiago Ávila-Ríos; Selene Zárate; Susana Lopez; Verónica Mata-Haro; Victor Eduardo García-Arias; Víctor Hugo Borja-Aburto |
| EPI_ISL_10993343 | Cianjur Public Health Laboratory | West Java Health Laboratory; School of Life Sciences and Technology, Institut Teknologi Bandung | Azzania Fibriani; Cut Nur Cinthia Alamanda; Ema Rahmawati; Karimatu Khoirunnisa; Miftahul Faridl; Rifky Waluyajati Rachman; Rini Robiani; Ryan Bayusantika Ristandi |
| EPI_ISL_11832515, EPI_ISL_13135431, EPI_ISL_13135599 | Clinical Emergency County Hospital, Craiova | Cantacuzino National Military-Medical Institute, Viral Respiratory Infections Laboratory | Carmen Cherciu; Catalina Pascu; Luiza Ustea; Mihaela Lazar; Mihaela Oprea; Pascu Catalina; Sorin Dinu; Vitencu Oana |
| EPI_ISL_12415946 | Clinical Hospital of Infectious Diseases, Cluj-Napoca | Cantacuzino National Military-Medical Institute, Viral Respiratory Infections Laboratory | Catalina Pascu; Luiza Ustea; Mihaela Lazar; Mihaela Oprea; Sorin Dinu; Vitencu Oana |
| EPI_ISL_13017266 | Cliniques universitaires Saint-Luc | UCLouvain/IREC/MBLG-CTMA | Benoit Kabamba Mukadi; Bertrand Bearzatto; Jean-Luc Gala; Valentin Coste |
| EPI_ISL_12666259 | Colsanitas Keralty | Instituto de Investigación de Recursos Biológicos Alexander von Humboldt | Daniel Martínez Vargas; Mailyn Gonzalez; Nicolás D. Franco-Sierra |
| EPI_ISL_11149531, EPI_ISL_11149533, EPI_ISL_11149552, EPI_ISL_11149643 | DSMRC | Defence Services Medical Research Centre (DSMRC) | Aung Myo Nyi; Aung Sitt Hmue; Khine Zaw Oo; Ko Ko Lwin; Kyaw Moe Htike; Kyaw Zawl Lin; Kyee Myint; Phyo Kyaw Aung; Thet Wai Oo; Zaw Win Htun |
| EPI_ISL_11027040 | Department for Virology, Molecular Biology and Genome Research, R. G. Lugar Center for Public Health Research, National Center for Disease Control and Public Health (NCDC) of Georgia. | Department for Virology, Molecular Biology and Genome Research, R. G. Lugar Center for Public Health Research, National Center for Disease Control and Public Health (NCDC) of Georgia. | Adam Kotorashvili; Amiran Gamkrelidze.; Ana Papkiauri; Ann Machablishvili; Anna Kasradze; Davit Tsaguria; Ekaterine Khmaladze; Ekaterine Zangaladze; Ekaterine Zhghenti; Giorgi Gogoladze; Giorgi Tomashvili; Gvantsa Brachveli; Gvantsa Chanturia; Irma Burjanadze; Ketevan Sidamonidze; Khatuna Zakhashvili; Lela Sabadze; Lela Urushadze; Magda Dgebuadze; Maia Alkhazashvili; Mari Gavashelidze; Mariam Zakalashvili; Marine Murtskhvaladze; Meri Pantsulaia; Nato Kotaria; Nino Berishvili; Paata Imnadze; Roena Sukhiashvili; Salome Javashvili; Tamar Jashiashvili; Tata Imnadze; Tea Tevdoradze |
| EPI_ISL_13352745, EPI_ISL_13352747, EPI_ISL_13352772, EPI_ISL_13352778 | Department of Clinical Laboratory | Department of Clinical Laboratory | Jin Xu; Mei Zeng; Yuanyun Ao |
| EPI_ISL_12417480, EPI_ISL_12417518, EPI_ISL_12417533, EPI_ISL_12417571, EPI_ISL_12417581, EPI_ISL_12417591, EPI_ISL_12417780, EPI_ISL_12417944 | | | |
| see above | Department of Health Technology and Informatics, The Hong Kong Polytechnic University | Department of Health Technology and Informatics, The Hong Kong Polytechnic University | Alan Ka-Lun Wu; Alex Yat-Man Ho; Barry Kin-Chung Wong; Chloe Toi-Mei Chan; David Ho-Keung Shum; Gilman Kit-Hang Siu; Hiu-Yin Lao; Ivan Tak-Fai Wong; Jake Siu-Lun Leung; Kam-Tong Yip; Kenneth Siu-Sing Leung; Kingsley King-Gee Tam; Kitty Sau-Chun Fung; Kristine Luk; Lam-Kwong Lee; Miranda Chong-Yee Yau; Sandy Ka-Yee Chau; Shea Ping Yip; Tak-Lun Que; Timothy Ting-Leung Ng; Wing Cheong Yam; Wing-Hei Lo; Wing-Kin To; Yvette Wai-Man Lai |
| EPI_ISL_13088333 | Department of Laboratory Services, National Virology Reference Laboratory | Clinical Molecular Diagnostic Laboratory For Infectious Disease, Department of Laboratory Services, Microbial Genomic Services | Amal Nabihah Ahmad; Faezah Fariha Abd Latif; Haziq Momin; Izzati Azhar; Nor Azian Hafneh; Nur Amirah Ibarahim; Zainun Zaini |
| EPI_ISL_11804454, EPI_ISL_11804489, EPI_ISL_11804580, EPI_ISL_11804613, EPI_ISL_12702064 | Department of Microbiology, National Institute of Public Health of Kosovo | Department of Microbiology, National Institute of Public Health of Kosovo | Aferdita Kuqi-Hyseni; Blendi Jerliu; Donjeta Hajdari; Nazmi Mehmeti; Pranvera Abazi; Robert Ramadani; Xhevat Jakupi; Zana Deva |
| EPI_ISL_9856176, EPI_ISL_10774006, EPI_ISL_10774010 | Department of Pathology and Laboratory Medicine, AKUH Laboratories, Karachi, Pakistan | Department of Pathology and Laboratory Medicine, AKUH Laboratories, Karachi, Pakistan | A. Kanji; A. Nasir; A. Samreen; A.R. Bukhari; AKU; Health Security Partners; Islamabad; J. Ashraf; P.Thielen; Pakistan; R. Hasan; U.B. Aamir; USA; USA); University Research Council Grant; WHO; Z. AzizUllah; Z. Hasan (Funding support NIH Fogarty Training Grant |
| EPI_ISL_10337127, EPI_ISL_10337406 | Department of Virology, National Veterinary Research Institute | Department of Omics Analysis, National Veterinary Research Institute | Bomba Arkadiusz; Domanska-Blicharz Katarzyna; Iwan Ewelina; Katarzyna Tluscik; Lisowska Anna; Niemczuk Krzysztof; Opolska Justyna; Orlowska Anna; Rola Jerzy; Smreczak Marcin; Trebas Pawel |
| EPI_ISL_8799568, EPI_ISL_8799569, EPI_ISL_8932859, EPI_ISL_8933054 | Dhulikhel Hospital, Kathmandu University Hospital | Molecular and Genomics Research Lab, Dhulikhel Hospital, Kathmandu University Hospital | Dipesh Tamrakar; Nishan Katuwal; Rajeev Shrestha; Surendra Kumar Madhup |
| EPI_ISL_11750210 | Diagnostyka Sp. z o.o. Gdańsk | Wojewódzka Stacja Sanitarno-Epidemiologiczna w Gdańsku | Barbara Skórczewska; Barbara Zawadzka; Gabriela Rutkowska; Marta Piszczyk |
| EPI_ISL_12750355, EPI_ISL_12899516 | Dr. Yoshitaka Tamura Department of Clinical Laboratory, Osaka Habikino Medical Center | Osaka Women's and Children's Hospital Department of Developmental Medicine, Research Institute, Osaka Women's and Children's Hospital | Itaru Yanagihara; Yukiko Nakura |
| EPI_ISL_10875757, EPI_ISL_10875764, EPI_ISL_10875767, EPI_ISL_10875777, EPI_ISL_10875781, EPI_ISL_10875786 | EWARN's Lab | Ministry of Health Turkey | Yasir Elferruh |
| EPI_ISL_12871556 | Fasa University of Medical Sciences | National Influenza Center | A Nejati; Adel Abedi; J Yavarian; K Sadeghi; Mrzieh Faraji-Zonouzand T Mokhtari Azad; NZ Shafiei Jandaghi; Nastaran Ghavami; Sevrin Zadheidar; V Salimi |
| EPI_ISL_12832821 | Fusco Lab | Maness Lab | Arnaud Drouin; Dahlene Fusco; Joshua Katz; Lori Rowe; Matthew Moreida; Nicholas Maness |
| EPI_ISL_8605841 | GENOME CENTER, Jashore University of Science and Technology. | GENOME CENTER, Jashore University of Science and Technology. | Hassan M. Al-Emran; Iqbal Kabir Jahid; Md. Ali Ahasan Setu; Md. Anwar Hossain; Md. Rasel Parvez; Md. Shaminur Rahman Prosanto Kumar Das; Md. Shazid Hasan; Shovon Lal Sarkar; Toukir Ahammed |
| EPI_ISL_11018136, EPI_ISL_11050899 | Genome Analysis Center, Yamanashi Central Hospital | Genome Analysis Center, Yamanashi Central Hospital | Yosuke Hirotsu; Yuki Nagakubo; Yuki Nagakubo and Yosuke Hirotsu |
| EPI_ISL_13169680 | HE-HOSPITAL DEL CANCER | Laboratorio Central de Salud Pública | Analía Rojas; Andrea Gómez de la Fuente; Cynthia Vazquez; César Cantero; César Ojeda; Emmanuel Céspedes; Fátima Fleitas; Ivana Fernández; Juan Torales; Julio Barrios; Maria Liz Gamarra; María Jose Duarte; Sandra González; Shirley Villalba; Tania Alfonzo; Wilson Benítez. |
| EPI_ISL_12172435 | HOSPITAL DR. TOMAS CASAS CASAJUS | Inciensa, Instituto Costarricense de Investigación y Enseñanza en Nutrición y Salud | Adriana Godínez; Claudio Soto-Garita; Estela Cordero; Francisco Duarte; Gabriel Morales & Antony Orozco Barquero; Hebleen Porras; José Luis Vargas; Mariela Gutiérrez; Melany Calderón; Sofia Herrera |
| EPI_ISL_12838819 | HOSPITAL UNIVERSITARIO FUNDACION SANTA FE DE BOGOTA | Instituto Nacional de Salud-Dirección de Investigación en Salud Pública | Beatriz de Arco; Carlos Franco-Muñoz; Diego A. Álvarez-Díaz; Diego Andrés Prada; Dioselina Peláez-Carvajal; Gerardo Santamaría; Héctor Alejandro Ruiz-Moreno; Jhonnatan Reales-González; Jorge Rivera; Julián Naizaque; Katherine Laiton-Donato; Marcela Mercado-Reyes.; Martha Lucia Ospina Martínez; María T. Herrera-Sepúlveda; Paola Rojas-Estevez; Sheryll Corchuelo; Tatiana Cobos |
| EPI_ISL_11006098 | Ha Nam | Bachmai Hospital | Doanh Khuong; Dung Le; Lan Pham; Linh Le; Nga Pham; Ngan Le; Phuong Truong; Van Vu; Vuong Bui |
| EPI_ISL_13025766 | Hoa Binh General Hospital | Bachmai Hospital | Doanh Khuong; Dung Le; Lan Pham; Linh Le; Nga Pham; Ngan Le; Phuong Truong; Van Vu; Vuong Bui |
| EPI_ISL_12157760, EPI_ISL_12157762 | Hopital | National Reference Center for Viruses of Respiratory Infections, Institut Pasteur, Paris | Angela Brisebarre; Camille Capel; Christophe Malabat; Corinne Maufrais; Etienne Simon-Lorière; Frédéric Lemoine; Julien Fumey; Laurence FAGOUR; Louise Lefrançois; Marion Barbet; Maud Vanpeene; Méline Bizard; Slim El Khiari; Sylvie Behillil; Sylvie Van der Werf; Vincent Enouf |
| EPI_ISL_12717510 | Hospital Universitario Marqués de Valdecilla | Servicio de Microbiologia. Hospital Universitario Marqués de Valdecilla | Elena Bolado Conde; Javier Freire Salinas; Jesús Agüero Balbín; Jesús Rodríguez Lozano; Jorge Calvo Montes; Mónica Gozalo Margüello; Sergio Garcia Fernandez |
| EPI_ISL_13337845 | Hospital de ciudad neily | Inciensa, Instituto Costarricense de Investigación y Enseñanza en | Adriana Godínez; Claudio Soto-Garita; Estela Cordero; Francisco Duarte; Gabriel Morales; Hebleen Porras; José Luis Vargas; Mariela Gutiérrez; Melany Calderón; Natalia Bonilla & Gabriela Marchena Jimenez; Sofia Herrera |

| | | | |
|---|---|---|---|
| | | Nutrición y Salud | |
| EPI_ISL_13350774 | Hospital del Cancer-INCAN | Laboratorio Central de Salud Pública | Analía Rojas; Andrea Gómez de la Fuente; Cynthia Vazquez; César Cantero; César Ojeda; Emmanuel Céspedes; Fátima Fleitas; Ivana Fernández; Juan Torales; Julio Barrios; Maria Liz Gamarra; María Jose Duarte; Sandra González; Shirley Villalba; Tania Alfonzo; Wilson Benítez. |
| EPI_ISL_12637345 | ICMT APARTADO | Laboratorio Departamental de Salud Publica de Antioquia | Ana Victoria Valencia Duarte; Cristian Arbey Velarde Hoyos; Gloria Isabel Escobar; Idabely Betancur Ortiz; Juan Pablo Isaza Agudelo |
| EPI_ISL_13107536 | INCMNSZ | Instituto Nacional de Medicina Genomica | Cedro-Tanda A; Cruz-Islas Jazmin; Escobar-Arrazola MA; Garnica-Lopez Dora; Herrera-Montalvo LA.; Hidalgo-Miranda A; Mendoza-Vargas A; Ramirez-Vega O; Rangel-DeLeon D; Reyes-Grajeda JP; Rubio-Alvarado PV; Yair Alfaro-Mora |
| EPI_ISL_9738913, EPI_ISL_10994857, EPI_ISL_10994862 | Indira Gandhi Memorial Hospital | Indira Gandhi Memorial Hospital | D. Fathmath Nazla Rafeeq; Dr. Ibrahim Afzal; Mr. Ibrahim Nishan Ahmed; Ms. Aishath Shuhudha; Ms. Aminath Shazleena Abdul Rahman; Ms. Soafy Mohamed |
| EPI_ISL_11732770 | Institut für Lebensmittelsicherheit, Veterinärmedizin, Umwelt | Institut für Lebensmittelsicherheit, Veterinärmedizin, Umwelt | Gunther Vogl |
| EPI_ISL_11878815, EPI_ISL_12650139, EPI_ISL_12896521 | Institute for Water Quality and Resource Management, Technical University Vienna | Bergthaler laboratory, CeMM Research Center for Molecular Medicine of the Austrian Academy of Sciences | Alberto Alises; Andreas Bergthaler; Anna Schedl; Bekir Erguner; Benedikt Agerer; Christoph Bock; Fabian Amman; Jan Laine; Lukas Endler; Martin Senekowitsch; Matthew Thornton; Michael Schuster; Michelle Chan; Petr Triska; Thomas Penz |
| EPI_ISL_12024316, EPI_ISL_12024321, EPI_ISL_12727976 | Institute of Legal Medicine, Medical University of Innsbruck | Bergthaler laboratory, CeMM Research Center for Molecular Medicine of the Austrian Academy of Sciences | Alberto Alises; Andreas Bergthaler; Anna Schedl; Christoph Bock; Fabian Amman; Lukas Endler; Matthew Thornton; Michael Schuster; Michelle Chan; Petr Triska |
| EPI_ISL_12153603, EPI_ISL_12153605, EPI_ISL_12153606, EPI_ISL_12153607, EPI_ISL_12153613, EPI_ISL_12153617, EPI_ISL_12153618, EPI_ISL_12153620, EPI_ISL_12153625 | | | |
| see above | Institute of Medical Microbiology and Hospital Hygiene | Institute of Medical Microbiology and Hospital Hygiene | Aljoscha Tersteegen; Prof. Dr. Achim Kaasch |
| EPI_ISL_12156899, EPI_ISL_12400912 | Institute of Molecular and Translational Medicine / Laboratory of Experimental Medicine, Faculty of Medicine and Dentistry, Palacky University | Institute of Molecular and Translational Medicine / Laboratory of Experimental Medicine, Faculty of Medicine and Dentistry, Palacky University | Barbora Blumová; Hana Jaworek; Kateřina Kubáňová; Marián Hajdúch; Rastislav Slavkovský; Vladimíra Koudeláková |
| EPI_ISL_13107322 | Instituto Nacional de Medicina Genomica | Instituto Nacional de Medicina Genomica | Cedro-Tanda A; Escobar-Arrazola MA; Garnica-Lopez Dora; Herrera-Montalvo LA.; Hidalgo-Miranda A; Mendoza-Vargas A; Ramirez-Vega O; Rangel-DeLeon D; Reyes-Grajeda JP; Yair Alfaro-Mora |
| EPI_ISL_11994764, EPI_ISL_11994767, EPI_ISL_11994771, EPI_ISL_11994776, EPI_ISL_11994781 | Iressef Genomics lab | IRESSEF | Abdou PADANE; Ambroise AHOUIDI; Aminata DIA; Aminata MBOUP; Astou Gaye GAYE; Barada CISSE; Birahim Piere NDIAYE; Cyrille Diedhiou; Diabou Diagne; Gora LO; Khadim GUEYE; Moustapha MBOW; Nafisatou LEYE; Ndeye Coumba Toure KANE; Papa Alassane DIAW; Samba Ndiour; Seni Ndiaye; Souleymane MBOUP; Yacine DIA |
| EPI_ISL_12657276 | Jurisdiccion Sanitaria No.1 | LESP Aguascalientes | Anastacio Palacios-Marmolejo; Antonio Hernandez-Mejia; Brian Muñoz-Gomez; David Hernandez-Wancho; Magally Tristan-Leon; Mirella Paredes-Salas; Miriam Lozano-Gamboa; Monica Ortiz-Palos; Sabel Hernandez-Zavala; Susana Morales-Moran. Brenda Ortiz-Jasso |
| EPI_ISL_11323578 | Kasturba Hospital Molecular Lab | Kasturba hospital WGS | Jayanthi Shastri; Vidushi Chitalia |
| EPI_ISL_11664731 | Kasturba Hospital Molecular Lab | NIV Influenza | Jayanthi Shastri; Vidushi Chitalia |
| EPI_ISL_9512091 | Kasturba Hospital Molecular Lab - INSACOG | Kasturba hospital WGS | Jayanthi Shastri; Vidushi Chitalia |
| EPI_ISL_12813250, EPI_ISL_12813251 | Klinisch Laboratorium ZNA | Klinisch Laboratorium ZNA | Verstrepen et al. |
| EPI_ISL_13037531 | Krajská nemocnice T.Bati, a.s. | Institute of Molecular and Translational Medicine / Laboratory of Experimental Medicine, Faculty of Medicine and Dentistry, Palacky University | Barbora Blumová; Hana Jaworek; Kateřina Kubáňová; Marián Hajdúch; Rastislav Slavkovský; Vladimíra Koudeláková |
| EPI_ISL_12061332, EPI_ISL_12175820, EPI_ISL_12175862, EPI_ISL_12299416, EPI_ISL_13064574, EPI_ISL_13064624 | Kuala Lumpur International Airport | Institute for Medical Research, Infectious Disease Research Centre, National Institutes of Health, Ministry of Health Malaysia | Anasir MI; G.Adypatti NM; Jamaluddin MS; Kalyanasundram J; Kamel K; MatRahim N; Nawi MH; Suib FA; Suppiah J; Thayan R |
| EPI_ISL_13168949 | LAB PRIV CYRLAB | Laboratorio Central de Salud Pública | Analía Rojas; Andrea Gómez de la Fuente; Cynthia Vazquez; César Cantero; César Ojeda; Emmanuel Céspedes; Fátima Fleitas; Ivana Fernández; Juan Torales; Julio Barrios; Maria Liz Gamarra; María Jose Duarte; Sandra González; Shirley Villalba; Tania Alfonzo; Wilson Benítez. |
| EPI_ISL_11586749 | LABORATORIUM BADAŃ KLINICZNYCH WSSE w OPOLU | Wojewódzka Stacja Sanitarno-Epidemiologiczna w Katowicach | Adrian Miara; Agata Chajda; Beata Rozwadowska; Dorota Liberska; Elzbieta Bartkowiak; Hubert Okla; Marta Albertynska; Natalia Kuczera |
| EPI_ISL_11892333 | LBA - SITE CASTELJALOUX | CERBA HealthCare | Bénédicte Roquebert; Laura Verdurme; Mathilde Roussel; Sabine Trombert; Stéphanie Haim-Boukobza |
| EPI_ISL_12140289 | LBBMS BIO POLE ANTILLES | CERBA HealthCare | Bénédicte Roquebert; Laura Verdurme; Mathilde Roussel; Sabine Trombert; Stéphanie Haim-Boukobza |
| EPI_ISL_9749785 | LESP Jalisco | Instituto de Diagnostico y Referencia Epidemiologicos (INDRE) | Abril Rodriguez-Maldonado; Ariadna Medina-Benitez; Claudia Wong-Arambula; Ernesto Ramirez-Gonzalez.; Fernando Gonzalez-Dominguez; Gisela Barrera-Badillo; Irma Lopez-Martinez; Joaquin Quiroz-Mercado; Lucia Hernandez-Rivas; Maribel Gonzalez-Villa; Natividad Cruz-Ortiz; Sergio Rangel-Guerrero; Tatiana Nunez-Garcia; Vanessa Rivero-Arredondo |
| EPI_ISL_13094338 | LOMWRU | LOMWRU/Microbiology Laboratory, Mahosot Hospital | Audrey Dubot-Pérès; Elizabeth Ashley; Elizabeth Batty; Manila Souksavanh; Manivanh Vongsouvath; Matthew T. Robinson; Siribun Panapruksachat |
| EPI_ISL_9618959 | Lab Klinik Utama Karunia | National Institute of Health Research and Development | Arie Ardiansyah Nugraha; Fajar Nur Sulistiyohadi; Hana Apsari Pawestri; Hartanti Dian Ikawati; Kartika Dewi Puspa; Nelly Puspandari; Nur Ika Hariastuti; Putri Widia; Subangkit; Vivi Setiawaty |
| EPI_ISL_11021578, EPI_ISL_12157642, EPI_ISL_12637726, EPI_ISL_13283347 | Labo Analyses Med | National Reference Center for Viruses of Respiratory Infections, Institut Pasteur, Paris | Angela Brisebarre; Camille Capel; Christophe Malabat; Corinne Maufrais; Etienne Simon-Lorière; Frédéric Lemoine; Isabelle SOURON; Julien Fumey; Louise Lefrançois; Marc BIRON; Marie-Hélène GLAUDON LOUVEAU DE LA GUIGNERAYE; Marion Barbet; Maud Vanpeene; Méline Bizard; Nicolas GUIGUE; Slim El Khiari; Sylvie Behillil; Sylvie Van der Werf; Vincent Enouf |
| EPI_ISL_11450489, EPI_ISL_12349816, EPI_ISL_12349820, EPI_ISL_12349827 | Laboratori d'anàlisis clíniques, Hospital Nostra Senyora de Meritxell | LBM de CHU de Toulouse, Hôpitaux de Toulouse | Garcia M et al.; Lobaco C; Rendon M |
| EPI_ISL_12906839, EPI_ISL_12906840 | Laboratorio Central de Salud Publica | Laboratorio de Biología Molecular, Instituto de Medicina Regional on behalf of 'Proyecto Argentino Interinstitucional de genomica de SARS-CoV-2' (PAIS Consortium) | Andrea Ayala; Bettina Brusés; Erica Struss; Esteban Paredes; Griselda Oria; Horacio Lucero.; Javier Mussin; Laura Formichelli; Melina Lorenzini Campos; Raúl Maximiliano Acevedo; Verónica Gómez; Victoria Femenías |
| EPI_ISL_13350750 | Laboratorio Central de Salud Pública | Laboratorio Central de Salud Pública | Analía Rojas; Andrea Gómez de la Fuente; Cynthia Vazquez; César Cantero; César Ojeda; Emmanuel Céspedes; Fátima Fleitas; Ivana Fernández; Juan Torales; Julio Barrios; Maria Liz Gamarra; María Jose Duarte; Sandra González; Shirley Villalba; Tania Alfonzo; Wilson Benítez. |
| EPI_ISL_13020757, EPI_ISL_13228829, EPI_ISL_13228831 | Laboratorio Central de Saude Publica do Estado de Santa Catarina (LACEN/SC) | Laboratory of Respiratory Viruses and Measles, Oswaldo Cruz Institute, FIOCRUZ | Alice Sampaio Rocha; Bruna Mendonça da Silva; Darcita Buerger Rovaris; Elisa Cavalcante Pereira; Fernando Motta; Ighor Arantes; Jéssica Graça Macedo de Carvalho; Larissa Macedo Pinto; Luciana Appolinario; Marilda Siqueira on behalf of the Fiocruz COVID-19 Genomic Surveillance Network; Paola Resende; Sandra Bianchini Fernandes; Victor Guimaraes |
| EPI_ISL_12680632 | Laboratorio Central de Saude Publica do Estado do Parana (LACEN/PR) | Laboratory of Respiratory Viruses and Measles, Oswaldo Cruz Institute, FIOCRUZ | Alice Sampaio Rocha; Bruna Mendonça da Silva; Elisa Cavalcante Pereira; Fernando Motta; Ighor Arantes; Irina Riediger; Jéssica Graça Macedo de Carvalho; Larissa Macedo Pinto; Luciana Appolinario; Marilda Siqueira on behalf of the Fiocruz COVID-19 Genomic Surveillance Network; Paola Resende; Victor Guimaraes |
| EPI_ISL_13350777 | Laboratorio Curie SRL | Laboratorio Central de Salud Pública | Analía Rojas; Andrea Gómez de la Fuente; Cynthia Vazquez; César Cantero; César Ojeda; Emmanuel Céspedes; Fátima Fleitas; Ivana Fernández; Juan Torales; Julio Barrios; Maria Liz Gamarra; María Jose Duarte; Sandra González; Shirley Villalba; Tania Alfonzo; Wilson Benítez. |
| EPI_ISL_12543211 | Laboratorio Nacional de Salud (LNS), Guatemala | Inciensa, Instituto Costarricense de Investigación y Enseñanza en Nutrición y Salud | Claudia Estrada; César Conde; Laboratorio de Genómica INCIENSA; Selene González |
| EPI_ISL_13081535, EPI_ISL_13081537, EPI_ISL_13081539, | Laboratorio Nacional de Salud (LNS), Guatemala | Laboratorio Nacional de Salud (LNS), Guatemala | Calan K; García G; M; Mendoza L; Perez |

| | | | |
|---|---|---|---|
| EPI_ISL_13096537, EPI_ISL_13096541, EPI_ISL_13096550 | | | |
| EPI_ISL_12694830, EPI_ISL_12694862, EPI_ISL_12694865, EPI_ISL_13221697, EPI_ISL_13221706, EPI_ISL_13221709, EPI_ISL_13221730, EPI_ISL_13221731 | | | |
| see above | Laboratorio de Infectologia y Virologia Molecular | Laboratory of Molecular Virology, School of Medicine, Pontificia Universidad Catolica de Chile | Ana Maria Contreras; Andres E. Munoz-Marcos; Carlos Palma; Catalina Pardo-Roa; Constanza Maldonado; Eileen Serrano; Erick Salinas; Estefany Poblete; Eugenia L. Fuentes Luppichini; Francisco Melo; Jennifer Angulo; Jorge Levican; Leonardo I. Almonacid; M. Belen Leyton; Marcela Ferres; Maria Jose Avendano; María José Avendaño; Rafael A. Medina.; Tamara Garcia-Salum |
| EPI_ISL_9791182 | Laboratorium Diagnostyki Medycznej | Wojewódzka Stacja Sanitarno-Epidemiologiczna w Gdańsku | Barbara Skórczewska; Gabriela Rutkowska; Karolina Smarzyńska |
| EPI_ISL_11701733 | Laboratorium Medyczne GynCentrum – Oddział Sosnowiec | Wojewódzka Stacja Sanitarno-Epidemiologiczna w Katowicach | Adrian Miara; Agata Chajda; Beata Rozwadowska; Dorota Liberska; Elzbieta Bartkowiak; Hubert Okla; Marta Albertynska; Natalia Kuczera |
| EPI_ISL_11269129 | Laboratory Med Diagnostics | Department for Virology, Molecular Biology and Genome Research, R. G. Lugar Center for Public Health Research, National Center for Disease Control and Public Health (NCDC) of Georgia. | Adam Kotorashvili; Amiran Gamkrelidze.; Ana Papkiauri; Ann Machablishvili; Anna Kasradze; Davit Tsaguria; Ekaterine Khmaladze; Ekaterine Zangaladze; Ekaterine Zhghenti; Giorgi Gogoladze; Giorgi Tomashvili; Gvantsa Brachveli; Gvantsa Chanturia; Irma Burjanadze; Ketevan Sidamonidze; Khatuna Zakhashvili; Lela Sabadze; Lela Urushadze; Magda Dgebuadze; Maia Alkhazashvili; Mari Gavashelidze; Mariam Zakalashvili; Marine Murtskhvaladze; Meri Pantsulaia; Nato Kotaria; Nino Berishvili; Paata Imnadze; Roena Sukhiashvili; Salome Javashvili; Tamar Jashiashvili; Tata Imnadze; Tea Tevdoradze |
| EPI_ISL_11269132 | Laboratory Pyramid | Department for Virology, Molecular Biology and Genome Research, R. G. Lugar Center for Public Health Research, National Center for Disease Control and Public Health (NCDC) of Georgia. | Adam Kotorashvili; Amiran Gamkrelidze.; Ana Papkiauri; Ann Machablishvili; Anna Kasradze; Davit Tsaguria; Ekaterine Khmaladze; Ekaterine Zangaladze; Ekaterine Zhghenti; Giorgi Gogoladze; Giorgi Tomashvili; Gvantsa Brachveli; Gvantsa Chanturia; Irma Burjanadze; Ketevan Sidamonidze; Khatuna Zakhashvili; Lela Sabadze; Lela Urushadze; Magda Dgebuadze; Maia Alkhazashvili; Mari Gavashelidze; Mariam Zakalashvili; Marine Murtskhvaladze; Meri Pantsulaia; Nato Kotaria; Nino Berishvili; Paata Imnadze; Roena Sukhiashvili; Salome Javashvili; Tamar Jashiashvili; Tata Imnadze; Tea Tevdoradze |
| EPI_ISL_10645403, EPI_ISL_11704808 | Laboratory for COVID19 diagnostics, Clinical Centre of Serbia | Institute of microbiology and Immunology, Faculty of Medicine, University of Belgrade | Jankovic, M.; Jovanovic, T.; Knezevic, A.; Milicevic, O.; Sekler, M.; Tesovic, B.; Vidanovic, D. |
| EPI_ISL_12236180, EPI_ISL_12337045, EPI_ISL_12345902 | Laboratory for Molecular Diagnostics,IPHMN | Eurofins Genomics Europe Sequencing GmbH | Danijela Vujošević; Marija Govedarica; Rejhan Hot |
| EPI_ISL_9441481, EPI_ISL_11166339, EPI_ISL_11166342, EPI_ISL_11166343 | Laboratory of Genetics and Personalized Medicine, Zan Mitrev Clinic | Laboratory of Genetics and Personalized Medicine, Zan Mitrev Clinic | Gjorgjievska M; Kungulovski G; Kungulovski G. et al.; et al. |
| EPI_ISL_11798846 | Laboratory of Genomics and Bioinformatics, Comenius University Science Park | Laboratory of Genomics and Bioinformatics, Comenius University Science Park | Anna Kaliňáková; Barbora Kotvasová; Diana Rusňáková; Jakub Styk; Jaroslav Budiš; Lucia Ševčíková; Michaela Jakubková Forgáčová; Miroslav Böhmer; Nikola Lipková; Pavol Mišenko; Silvia Bokorová; Tatiana Sedláčková; Terézia Vrabľová; Tomáš Szemes |
| EPI_ISL_13653682 | Laboratory of Respiratory Viruses and Measles, Oswaldo Cruz Institute, FIOCRUZ | Laboratory of Respiratory Viruses and Measles, Oswaldo Cruz Institute, FIOCRUZ | Alice Sampaio Rocha; Bruna Mendonça da Silva; Elisa Cavalcante Pereira; Fernando Motta; Ighor Arantes; Jéssica Graça Macedo de Carvalho; Larissa Macedo Pinto; Luciana Appolinario; Marilda Siqueira on behalf of the Fiocruz COVID-19 Genomic Surveillance Network; Paola Resende; Victor Guimaraes |
| EPI_ISL_12978303 | Laboratório de Microbiologia Molecular - Universidade FEEVALE | Laboratório de Microbiologia Molecular - Universidade FEEVALE | Alana Witt Hansen; Fernando Rosado Spilki; Fágner Henrique Heldt; Juliana Schons Gularte; Juliane Deise Fleck; Mariana Soares da Silva; Matheus Nunes Weber; Meriane Demoliner; Micheli Filippi; Paula Rodrigues de Almeida; Viviane Girardi; Vyctoria Malayhka |
| EPI_ISL_13169087 | MANILA DOCTORS HOSPITAL | Philippine Genome Center | Alethea R. de Guzman; Alyssa Joyce E. Telles; Anna Ong-Lim; Arianne A. Zamora; Benedict A. Maralit; Carlo M. Lapid; Celia Carlos; Cynthia P. Saloma; Devon Ray Pacial; Diomedes A. Cariño; Edsel Maurice Salvana; El King D. Morado; Elcid Aaron R. Pangilinan; Eva Maria Cutiongco-de la Paz; Francis A. Tablizo; Henrietta Marie Rodriguez; Jaime C. Montoya; Jan Michael C. Yap; Jarvin E. Nipales; Jo-Hannah S. Llames; John Michael Egana; John Q. Wong; Joshua Gregor A. Dizon; Joshua Jose Endozo; Juan Antonio R. Magalang; Karol Sophia Agape R. Padilla; Kris P. Punayan; Kristina Patriz Dela Cruz; Lindsay Clare D.L. Carandang; Ma. Exanil Plantig; Marc Edsel C. Ayes; Maria Rosario Singh-Vergeire; Maria Sofia L. Yangzon; Marielle M. Gamboa; Marissa Alejandria; Niña Francesca Bustamante; Razel Nikka M. Hao; Renato Jacinto Q. Mantaring; Rianna Patricia S. Cruz; Shiela Mae M. Araiza; Yvonne Valerie Austria; Zipporah Mariebelle R. Enriquez; Zyrel V. Mollejon |
| EPI_ISL_11523594 | Medical Genetics Laboratory, Regional Centre of Medical Genetics, Emergency County Hospital Craiova | Medical Genetics Laboratory, Regional Centre of Medical Genetics, Emergency County Hospital Craiova | Adina Dragos; Ana-Maria Buga; Anca-Lelia (Riza) Costache; Andrei Pirvu; Elena Plesea; Ioana Streata; Mihai Cucu; Mihai Ioana; Monica Cara; Razvan Plesea; Stefania Dorobantu |
| EPI_ISL_12006316, EPI_ISL_12006363, EPI_ISL_12006364 | Medical Genetics, Pamukkale University | Medical Genetics, Pamukkale University | Sari, T.; Tokgun, O. |
| EPI_ISL_12559611 | Medyczne Laboratorium Diagnostyczne, Szpital Miejski nr 4 w Gliwicach Sp. z.o.o | Wojewódzka Stacja Sanitarno-Epidemiologiczna w Katowicach | Adrian Miara; Agata Chajda; Beata Rozwadowska; Dorota Liberska; ElZbieta Bartkowiak; Hubert Okla; Marta AlbertyNska; Natalia Kuczera |
| EPI_ISL_12315253 | Microbiology Department, University Hospital Donostia | Microbiology Department, University Hospital Donostia | Cilla G.; Marimon JM; Martin-Peñaranda T; Montes M; Piñeiro L; Sorarrain A; Vallejo P |
| EPI_ISL_12130670, EPI_ISL_12484402, EPI_ISL_12484670, EPI_ISL_12484857 | Microbiology Department. Complexo Hospitalario Universitario de Vigo | Microbiology Department. Complexo Hospitalario Universitario de Vigo | Microbiology Department. Complexo Hospitalario Universitario de Vigo |
| EPI_ISL_12420377, EPI_ISL_13043517, EPI_ISL_13043700 | Molecular Biology Laboratory, Vidant Medical Center | Brody Integrative Genomics Core, East Carolina University | Changhong Yin; Heather Duncan; James Woodward; John T. Fallon; Kimberly P. Briley; Weihua Huang |
| EPI_ISL_12875335, EPI_ISL_12875336, EPI_ISL_12875340, EPI_ISL_12875371, EPI_ISL_12875386, EPI_ISL_13032234, EPI_ISL_13032235, EPI_ISL_13032269, EPI_ISL_13032270, EPI_ISL_13032282 | | | |
| see above | Molecular Genetic Monitoring Group | Molecular Genetic Monitoring Group | Anna S. Gladkikh; Areg A.Totolian; Ekaterina O. Klyuchnikova; Valerya A. Sbarzaglia; Vladimir G. Dedkov |
| EPI_ISL_13259123, EPI_ISL_13259130 | NIH | National Institute of Hygiene | Abderrahman Bimouhen; Fatima El Falaki; Hassan Ihazmade; Hicham Oumzil; Zakia Regragui |
| EPI_ISL_13289770, EPI_ISL_13289786, EPI_ISL_13289787, EPI_ISL_13289790, EPI_ISL_13289793 | NIH-Rabat | NIC at National Institute of Hygiene | Abderrahman Bimouhen; Fatima El Falaki; Hassan Ihazmade; Hicham Oumzil; Mohammed Rajaoui; Zakia Regragui |
| EPI_ISL_12643819, EPI_ISL_12643823, EPI_ISL_12791912, EPI_ISL_12791923, EPI_ISL_13078896, EPI_ISL_13345968 | NL-Dr. Leonard A. Miller Centre for Health Services | Newfoundland and Labrador - Eastern Health | Ana Duggan; Anna Majer; Anneliese Landgraff; CanCOGeN's metadata curation team; Christopher Corkum; Darian Hole; Elsie Grudeski; Gary Van Domselaar; Geoffrey Woodland; George Zahariadis; Grace Seo; Jennifer Tanner; Kirsten Biggar; Madison Chapel; Morag Graham; Natalie Knox; Nathalie Bastien; Phil Andrews; Philip Mabon; Public Health Agency of Canada CanCOGeN team; Rhiannon Huzarewich; Robert Needle; Russell Mandes; Shari Tyson; Timothy Booth; Yan Li; Yang Yu |
| EPI_ISL_13138232, EPI_ISL_13138351, EPI_ISL_13138352, EPI_ISL_13138609, EPI_ISL_13138625, EPI_ISL_13138640, EPI_ISL_13138695, EPI_ISL_13179231, EPI_ISL_13180592, EPI_ISL_13180788 | | | |
| see above | National Center of Infectious and Parasitic Diseases | National Center of Infectious and Parasitic Diseases | Alexiev et al |
| EPI_ISL_9342570, EPI_ISL_10893985, EPI_ISL_11526215 | National Health Laboratory | Botswana Harvard HIV Reference Laboratory | Boitumelo Zuze; Botshelo Radibe; Dorcas Maruapula; Joseph Makhema; Keoratile Ntshambiwa; Kgomotso Moruisi; Legodile Kooepile; Mosepele Mosepele; Mphaphi B. Mbulawa; Ontlametse T. Bareng; Pamela Smith-Lawrence; Roger Shapiro; Sefetogi Ramaologa; Shahin Lockman; Shirley Johane; Sikhulile Moyo; Simani Gaseitsiwe; Thongbotho Mphoyakgosi; Wonderful T. Choga |
| EPI_ISL_11674429, EPI_ISL_12587875, EPI_ISL_12872672 | National Health Laboratory Services, Virology | National Health Laboratory Services, Virology | Ashlyn S. C. Davis; Florette K. Treurnicht; Kathleen Subramoney; Nkhensani Mtileni; Selebogo F. Maoko |
| EPI_ISL_11209670, EPI_ISL_11209674, EPI_ISL_11209679, EPI_ISL_11209697, EPI_ISL_11209699 | National Influenza Center - Rafic Hariri University Hospital | National Influenza Center Lebanon Rafik Hariri University Hospital | Alissar Zalghout; Amirtharaj Francis; Hanan Abbass; Lina Mroueh; Mona Albuaini; Nada Ghosn; Nisrine Jammal |
| EPI_ISL_12603513 | National Institute of Infectious Diseases-Prof. Dr. Matei Bals Molecular Diagnostics Laboratory | National Institute of Infectious Diseases-Prof. Dr. Matei Bals Molecular Diagnostics Laboratory | Corina Casangiu; Dan Otelea; Leontina Banica; Marius Surleac; Ovidiu Vlaicu; Simona Paraschiv |

| | | | |
|---|---|---|---|
| EPI_ISL_9696521, EPI_ISL_10067998 | National Institute of Public Health | Virology Unit, Institut Pasteur du Cambodge | Cecile Troupin; Chau Darapheak; Chin Savuth; Erik A Karlsson; Jurre Y Siegers; Kraing Sidonn; Leakhena Pum; Ly Sovann; Sophoannadedh Rath; Veasna Duong; Yi Sengdoeurn |
| EPI_ISL_12252832 | National Public Health Laboratory | National Public Health Laboratory | Kamal Hisham Bin Kamarul Zaman; Muhamad Syamim Bin Roslan; Noriah Binti Mohd Yusof; Nur Hazliza Binti Salleh; Rehan Shuhada Binti Abu Bakar; Selvanesan A/L Sengol; Yu Kie A/P Chem |
| EPI_ISL_10796087, EPI_ISL_10804816, EPI_ISL_10805960 | National Virology Reference Laboratory | Microbial Genomic Services | Haziq Momin; Nor Azian Hafneh; Zainun Zaini |
| EPI_ISL_13020806 | Nghe An General Hospital | Bachmai Hospital | Doanh Khuong; Dung Le; Lan Pham; Linh Le; Nga Pham; Ngan Le; Phuong Truong; Van Vu; Vuong Bui |
| EPI_ISL_11936110, EPI_ISL_13091544 | OSPEDALE CIVILE TERAMO - CENTRO TRASFUSIONALE | Istituto Zooprofilattico Sperimentale dell'Abruzzo e Molise "G. Caporale" | Ancora M; Calistri P; Cammà C; Caporale M; Curini V; Delli Compagni E; Di Domenico M; Di Lollo Valeria; Di Pasquale A; Lorusso A; Mangone I; Marcacci M; Puglia I; Rinaldi A; Savini G; Scialabba S |
| EPI_ISL_13372020 | Oblastni nemocnice Trutnov a.s. | University Hospital Hradec Kralove | Helena Parova; Lenka Rysava; Marketa Gancarcikova; Monika Berankova |
| EPI_ISL_12806352 | Orion Laboratories | Orion Laboratories | Andrew Campagna; Keisha Simoneaux |
| EPI_ISL_12635835 | Ospedale Civile Teramo - Centro Trasfusionale | Istituto Zooprofilattico Sperimentale dell'Abruzzo e Molise "G. Caporale" | Ancora M; Calistri P; Cammà C; Caporale M; Curini V; Delli Compagni E; Di Domenico M; Di Lollo Valeria; Di Pasquale A; Lorusso A; Mangone I; Marcacci M; Puglia I; Rinaldi A; Savini G; Scialabba S |
| EPI_ISL_12851716, EPI_ISL_13331828, EPI_ISL_13331838, EPI_ISL_13331857 | Outre Mer | Institut Pasteur | Angela Brisebarre; Camille Capel; Christophe Malabat; Corinne Maufrais; Etienne Simon-Lorière; Frédéric Lemoine; Julien Fumey; Louise Lefrançois; Marc BIRON; Marie-Hélène GLAUDON LOUVEAU DE LA GUIGNERAYE; Marion Barbet; Maud Vanpeene; Méline Bizard; Slim El Khiari; Sylvaine BASTIAN; Sylvie Behillil; Sylvie Van der Werf; Vincent Enouf |
| EPI_ISL_11759616, EPI_ISL_11894151, EPI_ISL_11997193, EPI_ISL_12179886, EPI_ISL_12465580, EPI_ISL_12499883, EPI_ISL_13283356, EPI_ISL_13324157 | | | |
| see above | Outre Mer | National Reference Center for Viruses of Respiratory Infections, Institut Pasteur, Paris | Angela Brisebarre; Camille Capel; Christophe Malabat; Corinne Maufrais; Etienne Simon-Lorière; François NESTOUR; Frédéric Lemoine; Julien Fumey; Laurence FAGOUR; Louise Lefrançois; Marc BIRON; Marie-Hélène GLAUDON LOUVEAU DE LA GUIGNERAYE; Marion Barbet; Maud Vanpeene; Méline Bizard; Slim El Khiari; Stéphanie GUYOMARD-RABENIRINA; Sylvie Behillil; Sylvie Van der Werf; Vincent Enouf |
| EPI_ISL_13169212 | PHILIPPINE GENERAL HOSPITAL (PGH) | Philippine Genome Center | Alethea R. de Guzman; Alyssa Joyce E. Telles; Anna Ong-Lim; Arianne A. Zamora; Benedict A. Maralit; Carlo M. Lapid; Celia Carlos; Cynthia P. Saloma; Devon Ray Pacial; Diomedes A. Cariño; Edsel Maurice Salvana; El King D. Morado; Elcid Aaron R. Pangilinan; Eva Maria Cutiongco-de la Paz; Francis A. Tablizo; Henrietta Marie Rodriguez; Jaime C. Montoya; Jan Michael C. Yap; Jarvin E. Nipales; Jo-Hannah S. Llames; John Michael Egana; John Q. Wong; Joshua Gregor A. Dizon; Joshua Jose Endozo; Juan Antonio R. Magalang; Karol Sophia Agape R. Padilla; Kris P. Punayan; Kristina Patriz Dela Cruz; Lindsay Clare D.L. Carandang; Ma. Exanil Plantig; Marc Edsel C. Ayes; Maria Rosario Singh-Vergeire; Maria Sofia L. Yangzon; Marielle M. Gamboa; Marissa Alejandria; Niña Francesca Bustamante; Razel Nikka M. Hao; Renato Jacinto Q. Mantaring; Rianna Patricia S. Cruz; Shiela Mae M. Araiza; Yvonne Valerie Austria; Zipporah Mariebelle R. Enriquez; Zyrel V. Mollejon |
| EPI_ISL_13312858 | PO AVEZZANO - U.O.C. Laboratorio Analisi | Istituto Zooprofilattico Sperimentale dell'Abruzzo e Molise "G. Caporale" | Ancora M; Calistri P; Cammà C; Caporale M; Curini V; Delli Compagni E; Di Domenico M; Di Lollo Valeria; Di Pasquale A; Lorusso A; Mangone I; Marcacci M; Puglia I; Rinaldi A; Savini G; Scialabba S |
| EPI_ISL_13345346 | Palapye Primary Hospital Laboratory | Botswana Harvard HIV Reference Laboratory | Boitumelo Zuze; Botshelo Radibe; Dorcas Maruapula; Joseph Makhema; Keoratile Ntshambiwa; Kgomotso Moruisi; Legodile Kooepile; Lynnette N. Bhebhe; Mosepele Mosepele; Mphaphi B. Mbulawa; Ontlametse T. Bareng; Pamela Smith-Lawrence; Patrick T. Mokgethi; Roger Shapiro; Sefetogi Ramaologa; Shahin Lockman; Sikhulile Moyo; Simani Gaseitsiwe; Thongbotho Mphoyakgosi; Wonderful T. Choga |
| EPI_ISL_12139787 | Pardubicka nemocnice | University Hospital Hradec Kralove | Helena Kovarikova; Lenka Rysava; Marketa Gancarcikova; Monika Berankova |
| EPI_ISL_11742048 | Pathocare Pathology Laboratory, Vadodara | Gujarat Biotechnology Research Centre | Akhilesh Modi; Apurvasinh Puvar; Bhadreshsinh Gohil; Chaitanya Joshi; Disha Vora; Janvi Raval; Jaykumar Rangani; Madhvi Joshi; Nimesh Patel; Nitin Savaliya; Nitin Shukla; Priyank Chavda; Ramesh Pandit; Roshani Mishra; Sonal Sharma; Tasnim Trivedi; Viral Patel; Zarna Patel |
| EPI_ISL_12206505 | Province Public Health Laboratory | Province Public Health laboratory | Amrendra Mishra; Jitendra Sah; Manmohan Mishra; Shravan Kumar Mishra |
| EPI_ISL_11370184, EPI_ISL_11370186, EPI_ISL_11370187, EPI_ISL_11402758 | Provincial Public Health Reference Lab, Punjab AIDS Control Program, Lahore | Provincial Public Health Reference Lab, Punjab AIDS Control Program, Lahore | Abida Bano; Fuzail Ahmad; Hasnain Javed; Laiba; Warda Fatima |
| EPI_ISL_13069004 | R SOLVAY 27 | Institut de Pathologie et Genetique (IPG) | Jérémie Gras; Pascale Hilbert |
| EPI_ISL_10816832, EPI_ISL_10816834, EPI_ISL_10816835, EPI_ISL_10816836, EPI_ISL_10816837, EPI_ISL_10816843 | Recherche | National Reference Center for Viruses of Respiratory Infections, Institut Pasteur, Paris | Angela Brisebarre; Camille Capel; Christophe Malabat; Corinne Maufrais; Etienne Simon-Lorière; Frédéric Lemoine; Julien Fumey; Louise Lefrançois; Marion Barbet; Maud Vanpeene; Méline Bizard; Olivier DEJOUX; Slim El Khiari; Sylvie Behillil; Sylvie Van der Werf; Vincent Enouf |
| EPI_ISL_12727219 | Regional Medical Sciences Center 1 Chiang Mai | National Institute of Health, Department of Medical Sciences, Ministry of Public Health, Thailand | Archawin Rojanawiwat; Natchaya Khiadsang; Nuttida Thongpramul; Pakorn Piromtong; Pilailuk Okada; Sirikanda Wimol; Siripaporn Phuygun; Sunthareeya Waicharoen; Suratchana Mitrat; Thanutsapa Thanadachakul |
| EPI_ISL_12141415 | Regional Medical Sciences Center 12/1 Trang | National Institute of Health, Department of Medical Sciences, Ministry of Public Health, Thailand | Archawin Rojanawiwat; Natchaya Khiadsang; Nuttida Thongpramul; Pakorn Piromtong; Pilailuk Okada; Sirikanda Wimol; Siripaporn Phuygun; Sunthareeya Waicharoen; Suratchana Mitrat; Thanutsapa Thanadachakul |
| EPI_ISL_11179283, EPI_ISL_11179518 | Regional Medical Sciences Center 6 Chonburi | Medical Genomic Centre,Medical Life Sciences Institute,Department of Medical Sciences, Ministry of Public Health, Thailand | Archawin Rojanawiwat; Jirapha Pakdee; Natthakul Bunneang; Nuanjun Wichukchinda; Penpitcha Thawong; Pilailuk Akkapaiboon Okada; Pundharika Piboonsiri; Surakameth Mahasirimongkol; Waritta Sawaengdee |
| EPI_ISL_11986011 | S.C. Laboratorio Analisi, ASL 3 Liguria | U.O. Igiene, Ospedale Policlinico San Martino | Bruzzone Bianca; De Pace Vanessa; Domnich Alexander; Icardi Giancarlo on behalf of SARS-CoV-2 ITALIAN RESEARCH ENTERPRISE-(SCIRE) Collaborative Group; Orsi Andrea; Randazzo Nadia; Ricucci Valentina; Spitaleri Antonino; Stefanelli Federica |
| EPI_ISL_11864810 | SALUD DIGNA | Instituto Nacional de Medicina Genomica | Abraham Campos-Romero; Cedro-Tanda A; Escobar-Arrazola MA; Garcia-Garcia FE; Garnica-Lopez Dora; Herrera-Montalvo LA.; Hidalgo-Miranda A; Luna-Ruiz Marco; Mendoza-Vargas A; Moreno-Camacho José Luis; Ramirez-Vega O; Rangel-DeLeon D; Reyes-Grajeda JP; Rodriguez-Gallegos Jorge; Sanchez-Xochipa S; Yair Alfaro-Mora |
| EPI_ISL_12685438, EPI_ISL_12685498, EPI_ISL_12727363 | SC (UCO) Igiene e Sanità Pubblica, ASUGI, Trieste | SC (UCO) Igiene e Sanità Pubblica, ASUGI, Trieste | Basaglia G; Busetti M; D'Agaro P; Fontana F; Forciniti G; Koncan R; Pipan C; Piscianz E; Segat L |
| EPI_ISL_9859616, EPI_ISL_11438687, EPI_ISL_11438717, EPI_ISL_11438720 | SI «Public Health Center of MHU» | The Institute of Molecular Biology and Genetics of NASU | M.Tukalo et al. |
| EPI_ISL_13169039 | SOUTHERN ISABELA MEDICAL CENTER MOLECULAR DIAGNOSTIC & RESEARCH LABORATORY (SIMC - MDRL) | Philippine Genome Center | Alethea R. de Guzman; Alyssa Joyce E. Telles; Anna Ong-Lim; Arianne A. Zamora; Benedict A. Maralit; Carlo M. Lapid; Celia Carlos; Cynthia P. Saloma; Devon Ray Pacial; Diomedes A. Cariño; Edsel Maurice Salvana; El King D. Morado; Elcid Aaron R. Pangilinan; Eva Maria Cutiongco-de la Paz; Francis A. Tablizo; Henrietta Marie Rodriguez; Jaime C. Montoya; Jan Michael C. Yap; Jarvin E. Nipales; Jo-Hannah S. Llames; John Michael Egana; John Q. Wong; Joshua Gregor A. Dizon; Joshua Jose Endozo; Juan Antonio R. Magalang; Karol Sophia Agape R. Padilla; Kris P. Punayan; Kristina Patriz Dela Cruz; Lindsay Clare D.L. Carandang; Ma. Exanil Plantig; Marc Edsel C. Ayes; Maria Rosario Singh-Vergeire; Maria Sofia L. Yangzon; Marielle M. Gamboa; Marissa Alejandria; Niña Francesca Bustamante; Razel Nikka M. Hao; Renato Jacinto Q. Mantaring; Rianna Patricia S. Cruz; Shiela Mae M. Araiza; Yvonne Valerie Austria; Zipporah Mariebelle R. Enriquez; Zyrel V. Mollejon |
| EPI_ISL_13169032 | SUPERCARE MEDICAL SERVICES, INC. | Philippine Genome Center | Alethea R. de Guzman; Alyssa Joyce E. Telles; Anna Ong-Lim; Arianne A. Zamora; Benedict A. Maralit; Carlo M. Lapid; Celia Carlos; Cynthia P. Saloma; Devon Ray Pacial; Diomedes A. Cariño; Edsel Maurice Salvana; El King D. Morado; Elcid Aaron R. Pangilinan; Eva Maria Cutiongco-de la Paz; Francis A. Tablizo; Henrietta Marie Rodriguez; Jaime C. Montoya; Jan Michael C. Yap; Jarvin E. Nipales; Jo-Hannah S. Llames; John Michael Egana; John Q. Wong; Joshua Gregor A. Dizon; Joshua Jose Endozo; Juan Antonio R. Magalang; Karol Sophia Agape R. Padilla; Kris P. Punayan; Kristina Patriz Dela Cruz; Lindsay Clare D.L. Carandang; Ma. Exanil Plantig; Marc Edsel C. Ayes; Maria Rosario Singh-Vergeire; Maria Sofia L. Yangzon; Marielle M. Gamboa; Marissa Alejandria; Niña Francesca Bustamante; Razel Nikka M. Hao; Renato Jacinto Q. Mantaring; Rianna Patricia S. Cruz; Shiela Mae M. Araiza; Yvonne Valerie Austria; Zipporah Mariebelle R. Enriquez; Zyrel V. Mollejon |
| EPI_ISL_13134834 | SURA | Laboratorio Departamental de Salud Publica de Antioquia | Ana Victoria Valencia Duarte; Cristian Arbey Velarde Hoyos; Gloria Isabel Escobar; Idabely Betancur Ortiz; Juan Pablo Isaza Agudelo |
| EPI_ISL_13300419 | SYNLAB ANGEL DIAGNOSTICA | Universidad del Valle | Andres Castillo; Beatriz Parra & Programa Nacional de Caracterización Genómica de SARS-CoV-2; David Valencia; Diana López-Alvarez; Erica M. Aristizabal; Flor Saa; Melissa Solarte; Nelson Rivera Franco |
| EPI_ISL_12871530 | Sari University of Medical Sciences | National Influenza Center | A Nejati; Adel Abedi and T Mokhtari Azad; J Yavarian; K Sadeghi; Marzieh Fraji-Zonouz; NZ Shafiei Jandaghi; Nastaran Ghavami; Sevrin Zadheidar; V Salimi |
| EPI_ISL_11750302 | Sayang Cianjur General Hospital Laboratory | West Java Health Laboratory; School of Life Sciences and Technology, Institut Teknologi Bandung | Azzania Fibriani; Cut Nur Cinthia Alamanda; Ema Rahmawati; Hadiana; Karimatu Khoirunnisa; Miftahul Faridl; Rifky Waluyajati Rachman; Rini Robiani; Ryan Bayusantika Ristandi |
| EPI_ISL_12970541 | Semnan University of Medical Sciences | National Influenza Center | A Nejati; Adel Abedi; J Yavarian; K Sadeghi; Mrzieh Faraji-Zonouzand T Mokhtari Azad; NZ Shafiei Jandaghi; Nastaran Ghavami; Sevrin Zadheidar; V Salimi |
| EPI_ISL_12139809, EPI_ISL_12139871, EPI_ISL_12144237, EPI_ISL_12184894, EPI_ISL_12185039 | Servicio Microbiología H.U. Dr. Negrín | Servicio Microbiología H.U. Dr. Negrín | Ana Bordes Benítez; Francisco Javier Chamizo López; Francisco Javier Chamizo López Ana Bordes Benítez |
| EPI_ISL_12871526, EPI_ISL_12970531 | Shahrekord University of Medical Sciences | National Influenza Center | A Nejati; Adel Abedi; Adel Abedi and T Mokhtari Azad; J Yavarian; K Sadeghi; Marzieh Fraji-Zonouz; NZ Shafiei Jandaghi; Nastaran Ghavami; Sevrin Zadheidar; V Salimi; and T Mokhtari Azad |

| | | | |
|---|---|---|---|
| EPI_ISL_9698125, EPI_ISL_9963286, EPI_ISL_10444842, EPI_ISL_10798736, EPI_ISL_11287432, EPI_ISL_11737003, EPI_ISL_11739553, EPI_ISL_11739584, EPI_ISL_13029278, EPI_ISL_13066207 | | | |
| see above | Synlab Eesti OÜ | 1. Laboratory of Communicable Diseases (Estonia); 2. Eurofins Genomics Europe Sequencing GmbH | Abroi A.; Avi R.; Dotsenko L.; Epštein J.; Hoidmets D.; Huik K.; Härma M-A.; Jaaniso E.; Kaarna K.; Kallas E.; Koppel I.; Kuzmin I.; Lahesaare A.; Lutsar I.; Metspalu M.; Milani L.; Naaber P.; Niglas H.; Oopkaup O.E.; Pauskar M.; Peterson H.; Päll T.; Ratnik K.; Raudvere U.; Reisberg T.; Sadikova O.; Sepp H.; Shablinskaja A.; Suija H.; Talas U.G.; Truusalu K. |
| EPI_ISL_11586788 | Szpital Uniwersytecki w Krakowie | Wojewódzka Stacja Sanitarno-Epidemiologiczna w Katowicach | Adrian Miara; Agata Chajda; Beata Rozwadowska; Dorota Liberska; Elzbieta Bartkowiak; Hubert Okla; Marta Albertynska; Natalia Kuczera |
| EPI_ISL_11147430 | TSGH-CP molecular lab | TSGH-CP molecular lab | Cherng-Lih Perng; Chien-Wen Chen; Chih-Kai Chang; Feng-Yee Chang; Hsing-Yi Chung; Hung-Sheng Shang; Jung-Chung Lin; Kuo-Ming Yeh; Kuo-Sheng Hung; Ming-Jr JIAN; Sheng-Kang Chiu; Shih-Hung Tsai; Tien-Yao Chang |
| EPI_ISL_13322005 | Thai Binh General Hospital | Bachmai Hospital | Doanh Khuong; Dung Le; Lan Pham; Linh Le; Nga Pham; Ngan Le; Phuong Truong; Van Vu; Vuong Bui |
| EPI_ISL_11583310 | Thai Binh hospital | Bachmai Hospital | Doanh Khuong; Dung Le; Lan Pham; Linh Le; Nga Pham; Ngan Le; Phuong Truong; Van Vu; Vuong Bui |
| EPI_ISL_12248962, EPI_ISL_12870044 | Tokyo Medical and Dental University | Tokyo Medical and Dental University | Akinori Kimura; Hiroaki Takeuchi; Kousuke Tanimoto; Yoko Nukui; Yoshiaki Gu; Yukie Tanaka |
| EPI_ISL_11221479 | Turkish Society of Internal Medicine | Gen Era Diagnostics Corporation Life Science Research and Molecular Diagnostics | Serhat Ünal |
| EPI_ISL_12195111, EPI_ISL_12212702 | USL1 | A.O. PERUGIA | Bicchieraro G; Bondi P; Cappelletti E; Ciurnelli R; Lepri E; Lucheroni F; Malagigi V; Spaccapelo R; Tacconi P |
| EPI_ISL_12771335 | Unit Forensik, Hospital Kepala Batas | iPROMISE, UiTM | Ariza Adnan; Fadzilah Mohd Nor; Lim Wai Feng; Mohd Asif Mohd Sukri; Mohd Nur Fakhruzzaman Noorizhab; Mohd Zaki Salleh; Sazzli Shahlan Kassim; Siti Farah Alwani Mohd Nawi; Siti Hamimah Sheikh Abdul Kadir; Teh Lay Kek; Wang Seok Mui |
| EPI_ISL_10440805, EPI_ISL_11159932, EPI_ISL_12862020 | University Hospital Hradec Kralove | University Hospital Hradec Kralove | Helena Kovarikova; Lenka Rysava; Marketa Gancarcikova; Monika Berankova |
| EPI_ISL_11010632 | Università degli Studi di Perugia | Istituto Zooprofilattico Sperimentale dell'Abruzzo e Molise "G. Caporale" | Ancora M; Biagetti M; Calistri P; Camilloni B; Cammà C; Curini V; Delli Compagni E; Di Domenico M; Di Pasquale A; Giammarioli M; Lorusso A; Mangone I; Marcacci M; Mencacci A; Puglia I; Rinaldi A; Savini G; Scialabba S |
| EPI_ISL_13172633, EPI_ISL_13173728, EPI_ISL_13173897 | VICENTE SOTTO MEMORIAL MEDICAL CENTER (VSMMC) | Philippine Genome Center | Alethea R. de Guzman; Alyssa Joyce E. Telles; Anna Ong-Lim; Arianne A. Zamora; Benedict A. Maralit; Carlo M. Lapid; Celia Carlos; Cynthia P. Saloma; Devon Ray Pacial; Diomedes A. Cariño; Edsel Maurice Salvana; El King D. Morado; Elcid Aaron R. Pangilinan; Eva Maria Cutiongco-de la Paz; Francis A. Tablizo; Henrietta Marie Rodriguez; Jaime C. Montoya; Jan Michael C. Yap; Jarvin E. Nipales; Jo-Hannah S. Llames; John Michael Egana; John Q. Wong; Joshua Gregor A. Dizon; Joshua Jose Endozo; Juan Antonio R. Magalang; Karol Sophia Agape R. Padilla; Kris P. Punayan; Kristina Patriz Dela Cruz; Lindsay Clare D.L. Carandang; Ma. Exanil Plantig; Marc Edsel C. Ayes; Maria Rosario Singh-Vergeire; Maria Sofia L. Yangzon; Marielle M. Gamboa; Marissa Alejandria; Niña Francesca Bustamante; Razel Nikka M. Hao; Renato Jacinto Q. Mantaring; Rianna Patricia S. Cruz; Shiela Mae M. Araiza; Yvonne Valerie Austria; Zipporah Mariebelle R. Enriquez; Zyrel V. Mollejon |
| EPI_ISL_10283481, EPI_ISL_10284065, EPI_ISL_10730607, EPI_ISL_10730610, EPI_ISL_10910333, EPI_ISL_11106023, EPI_ISL_11106084, EPI_ISL_11106669, EPI_ISL_11106789, EPI_ISL_11412329, EPI_ISL_11412331 | | | |
| see above | Virology Laboratory, NAPH, Moldova | Virology Laboratory, NAPH, Moldova | Apostol Mariana; Cataraga Alina; Colac Svetlana; Olga Burduniuc |
| EPI_ISL_10314780, EPI_ISL_10635697, EPI_ISL_10635740, EPI_ISL_11023851, EPI_ISL_12089737, EPI_ISL_12089792, EPI_ISL_12089851 | | | |
| see above | Virology Unit, Institut Pasteur du Cambodge | Virology Unit, Institut Pasteur du Cambodge | Cecile Troupin; Chau Darapheak; Chin Savuth; Erik A Karlsson; Jurre Y Siegers; Kraing Sidonn; Leakhena Pum; Ly Sovann; Sophoannadedh Rath; Veasna Duong; Yi Sengdoeurn |
| EPI_ISL_10847043 | Vitomed Sp. z o.o. | Wojewódzka Stacja Sanitarno-Epidemiologiczna w Katowicach | Adrian Miara; Agata Chajda; Beata Rozwadowska; Elżbieta Bartkowiak; Hubert Okla; Marta Albertyńska; Natalia Kuczera |
| EPI_ISL_11789460 | Đà Nẵng, Vietnam | Department of Microbiology and Immunology, Pasteur Institute in Nha Trang | Do Thai Hung; Hoang Tien Thanh; Huynh Kim Mai; Nguyen Bao Trieu; Nguyen Dinh Luong; Nguyen Duc Duy; Trinh Hoang Long; Vu Thi Ngoc |
