## Supplementary material for "Tracing the international arrivals of SARS-CoV-2 Omicron variants after Aotearoa New Zealand reopened its border": BA.4 GISAID acknowledgements

We gratefully acknowledge the following Authors from the Originating laboratories responsible for obtaining the specimens, as well as the Submitting laboratories where the genome data were generated and shared via GISAID, on which this research is based.

All Submitters of data may be contacted directly via www.gisaid.org

Authors are sorted alphabetically.

Acknowledgement EPI_SET Identifier: EPI_SET_20220706bz

| Accession ID | Originating Laboratory | Submitting Laboratory | Authors |
|---|---|---|---|
| EPI_ISL_12273990, EPI_ISL_12274060, EPI_ISL_12307643, EPI_ISL_12476998, EPI_ISL_12520048, EPI_ISL_12763752, EPI_ISL_12763807, EPI_ISL_12765602, EPI_ISL_12903392, EPI_ISL_12903602 | | | |
| see above | AMPATH | National Institute for Communicable Diseases of the National Health Laboratory Service | Amoako DG; Bhiman JN; Everatt J; Ismail A; Kekana D; Mahlangu B; Mnguni A; Mohale T; Ntuli N; Scheepers C; Wolter N |
| EPI_ISL_12790124 | AOUC Azienda Ospedaliero-Universitaria Careggi | Microbiology and Virology Unit, Florence Careggi University Hospital; Department of Experimental and Clinical Medicine, University of Florence, Florence, Italy | Gianmaria Rossolini |
| EPI_ISL_12722930 | AREA DE SALUD HEREDIA-CUBUJUQUI - CLINICA DR. FRANCISCO BOLAÑOS | Inciensa, Instituto Costarricense de Investigación y Enseñanza en Nutrición y Salud | Adriana Godínez; Claudio Soto-Garita; Estela Cordero; Francisco Duarte; Gabriel Morales & Natalia Bonilla; Hebleen Porras; José Luis Vargas; Mariela Gutiérrez; Melany Calderón; Sofia Herrera |
| EPI_ISL_12935378 | AREA DE SALUD MATA REDONDA-HOSPITAL - CLINICA DR. MORENO CAÑAS | Inciensa, Instituto Costarricense de Investigación y Enseñanza en Nutrición y Salud | Adriana Godínez; Claudio Soto-Garita; Estela Cordero; Francisco Duarte; Gabriel Morales & Natalia Bonilla; Hebleen Porras; José Luis Vargas; Mariela Gutiérrez; Melany Calderón; Sofia Herrera |
| EPI_ISL_12278927 | ASP Reggio Calabria Polo Sanitario Nord - Dr.ssa Fiorillo | SOC Microbiologia e Virologia - AO Pugliese-Ciaccio | Pasquale Minchella |
| EPI_ISL_12780020 | ASST GOM NIGUARDA | ASST Grande ospedale Metropolitano Niguarda | Alice Nava |
| EPI_ISL_12401707, EPI_ISL_12401714, EPI_ISL_12401721, EPI_ISL_12496098, EPI_ISL_12496100, EPI_ISL_12496101, EPI_ISL_12496103, EPI_ISL_12496106, EPI_ISL_12496108, EPI_ISL_12496111, EPI_ISL_12496114, EPI_ISL_12496123, EPI_ISL_12496124, EPI_ISL_12563697 | | | |
| see above | ASST MONZA | ASST MONZA | Sergio Maria Ivano Malandrin |
| EPI_ISL_12758092 | AZ Klina | AZ Klina | Carl Vael; Lynsey Berckmans |
| EPI_ISL_13124152 | Ampath | National Institute for Communicable Diseases of the National Health Laboratory Service | Amoako DG; Bhiman JN; Everatt J; Ismail A; Kekana D; Mahlangu B; Mnguni A; Mohale T; Ntuli N; Scheepers C; Wolter N |
| EPI_ISL_12206542 | Area of Virology, Serology and Virology Division (SAViD), New South Wales Health Pathology Randwick | Virology Research Laboratory; Area of Virology, Serology and Virology Division (SAViD), New South Wales Health Pathology Randwick | Foster, C.; Jean, T.; Rawlinson, W.; Van Hal, S.; Wong, M.; Yeang, M. |
| EPI_ISL_12578776, EPI_ISL_12578777 | Area of Virology, Serology and Virology Division (SAViD), New South Wales Health Pathology Randwick, Prince Of Wales Hospital | Area of Virology, Serology and Virology Division (SAViD), New South Wales Health Pathology Randwick, Prince Of Wales Hospital | Foster, C.; Jean, T.; Rawlinson, W.; Van Hal, S.; Wong, M.; Yeang, M. |
| EPI_ISL_12679229 | Australian Clinical Labs (formerly Healthscope Pathology) | NSW Health Pathology - Institute of Clinical Pathology and Medical Research; Westmead Hospital; University of Sydney | Arnott A.; Draper J.; Gall M.; Martinez E.; Rockett R.; Sintchenko V.; on behalf of ICPMR |
| EPI_ISL_12727829, EPI_ISL_12896303, EPI_ISL_12896305, EPI_ISL_12896306, EPI_ISL_12896308, EPI_ISL_12896310, EPI_ISL_12896313 | | | |
| see above | Austrian Agency for Health and Food Safety (AGES) | Bergthaler laboratory, CeMM Research Center for Molecular Medicine of the Austrian Academy of Sciences | Alberto Alises; Andreas Bergthaler; Anna Schedl; Christoph Bock; Fabian Amman; Lukas Endler; Matthew Thornton; Michael Schuster; Michelle Chan; Petr Triska |
| EPI_ISL_12421048 | BIO VSM LAB | Department of Virology, Henri Mondor University Hospital, Assistance Publique Hôpitaux de Paris, Université Paris-Est Créteil, INSERM U955 | Alexandre Soulier; Christophe Rodriguez; Elisabeth Trawinski; Guillaume Gricourt; Jean-Michel Pawlotsky; Melissa N'Debi; Slim Fourati; Vanessa Demontant |
| EPI_ISL_12837202, EPI_ISL_12837262 | BioneXt LAB - Laboratoire d'analyses médicales | Microbiology, Microbial Genomics Platform, LNS Laboratoire National De Santé | Anke Wienecke-Baldacchino; Catherine Ragimbeau; Elodie Solarino; Eric Hugoson; Fatu Djabi; Jessica Tapp; Lise Pignon; Raoul Salmon; Sibel Berger; Tamir Abdelrahman; Thibault Ferrandon; Virginie Jover |
| EPI_ISL_12527983 | Bioscientia Labor Wermsdorf | Robert Koch Institute | |
| EPI_ISL_12119232 | Botswana Harvard HIV Reference Laboratory | Botswana Harvard HIV Reference Laboratory | Boitumelo Zuze; Dorcas Maruapula; Joseph Makhema; Kgomotso Moruisi; Legodile Kooepile; Mosepele Mosepele; Mphaphi B. Mbulawa; Ontlametse T. Bareng; Pamela Smith-Lawrence; Roger Shapiro; Sefetogi Ramaologa; Shahin Lockman; Shirley Johane; Sikhulile Moyo; Simani Gaseitsiwe; Thongbotho Mphoyakgosi; Wonderful T. Choga |
| EPI_ISL_12968476, EPI_ISL_13318092 | British Columbia Centre For Disease Control | B.C. Centre for Disease Control Public Health Laboratory | Ana Pacagnella; Corrinne Ng; Dan Fornika; James Zlosnik; John Tyson; Kim Macdonald; Kimia Kamelian; Linda Hoang; Loretta Janz; Mel Krajden; Prystajecky Natalie; Robert Azana; Shannon Russell |
| EPI_ISL_12665547, EPI_ISL_12665652 | British Columbia Centre For Disease Control | BCCDC Public Health Laboratory | Ana Pacagnella; Corrinne Ng; Dan Fornika; James Zlosnik; John Tyson; Kim Macdonald; Kimia Kamelian; Linda Hoang; Loretta Janz; Mel Krajden; Prystajecky Natalie; Robert Azana; Shannon Russell |
| EPI_ISL_12682132, EPI_ISL_12791446 | Broad Institute Clinical Research Sequencing Platform | Infectious Disease Program, Broad Institute of Harvard and MIT | Adams, G.; B.L.; B.W.; Bauer, M.; Birren; Blumenstiel, B.; Brown, C.; Carter, A.; Chaluvadi, S.; D.J.; DeFelice, M.; DeRuff, K.; Dodge, S.; Gabriel, S.; Gallagher, G.; Gladden-Young, A.; Granger, B.; J.E.; K.J.; Lagerborg, K.; Larkin, K.; Lee, M.; Lemieux; Lennon, N.; Loreth, C.; Madoff, L.; McGovern, S.; Meldrim, J.; Normandin, E.; P.C.; Park; Pearlman, L.; Reilly, S.; Rudy, M.; Sabeti; Siddle; Smole, S.; Tomkins-Tinch, C.; Vicente, G.; and MacInnis |
| EPI_ISL_13276608, EPI_ISL_13276610, EPI_ISL_13276613, EPI_ISL_13276614, EPI_ISL_13276621, EPI_ISL_13276622, EPI_ISL_13276623, EPI_ISL_13276624, EPI_ISL_13276625, EPI_ISL_13276633, EPI_ISL_13276634, EPI_ISL_13276636 | | | |
| see above | Bumrungrad International Hospital | National Institute of Health, Department of Medical Sciences, Ministry of Public Health, Thailand | Archawin Rojanawiwat; Natchaya Khiadsang; Nuttida Thongpramul; Pakorn Piromtong; Pilailuk Okada; Sirikanda Wimol; Siripaporn Phuygun; Sunthareeya Waicharoen; Suratchana Mitrat; Thanutsapa Thanadachakul |
| EPI_ISL_12954867, EPI_ISL_12954868, EPI_ISL_12954870, EPI_ISL_12954873, EPI_ISL_12954877, EPI_ISL_12954878, EPI_ISL_12954879, EPI_ISL_12954880, EPI_ISL_12954881, EPI_ISL_12954882, EPI_ISL_12954883, EPI_ISL_12954884, EPI_ISL_12954887, EPI_ISL_12954888, EPI_ISL_12954889, EPI_ISL_12954890, EPI_ISL_12954891, EPI_ISL_12954892, EPI_ISL_12954895, EPI_ISL_12954896, EPI_ISL_12954897, EPI_ISL_12954900, EPI_ISL_12954903 | | | |
| see above | CENTRAL HEALTH LABORATORY | National Institute for Communicable Diseases of the National Health Laboratory Service | Amoako DG; Bhiman JN; Everatt J; Ismail A; Kekana D; Mahlangu B; Mnguni A; Mohale T; Ntuli N; Scheepers C; Wolter N |
| EPI_ISL_12688770, EPI_ISL_12688820, EPI_ISL_12688839 | CH. E. MULLER | Department of Virology, Henri Mondor University Hospital, Assistance Publique Hôpitaux de Paris, Université Paris-Est Créteil, INSERM U955 | Alexandre Soulier; Christophe Rodriguez; Elisabeth Trawinski; Guillaume Gricourt; Jean-Michel Pawlotsky; Melissa N'Debi; Slim Fourati; Vanessa Demontant |
| EPI_ISL_12763787 | CHRIS HANI BARAGWANATH LABORATORY | National Institute for Communicable Diseases of the National Health Laboratory Service | Amoako DG; Bhiman JN; Everatt J; Ismail A; Kekana D; Mahlangu B; Mnguni A; Mohale T; Ntuli N; Scheepers C; Wolter N |
| EPI_ISL_13068754 | CHUV | Laboratory of genomics and metagenomics, Institute of Microbiology, University Hospital Centre and University of Lausanne | Claire Bertelli; Damien Jacot; Gilbert Greub; Sébastien Aeby; Trestan Pillonel |
| EPI_ISL_13331190, EPI_ISL_13331210 | Central Health Laboratory, Ministry of Health and Wellness, Mauritius | Central Health Laboratory, Ministry of Health and Wellness, Mauritius | Bahadoor BS; Mathur H; Ramuth M Janoo N; Sonoo J; Sujeewon C; Ubheeram J |
| EPI_ISL_12456544, EPI_ISL_12633207, EPI_ISL_12636562, EPI_ISL_12637066, EPI_ISL_12637330 | Clinical Microbiology Laboratory, Tel Aviv Sourasky Medical Center | Clinical Microbiology Laboratory, Tel Aviv Sourasky Medical Center | Alon Ziv; Amos Adler; Goel Morad; Katya Levytskyi; Lior Handler; Matan Slutzkin; Ora Halutz; Orly Eshel |

| | | | |
|---|---|---|---|
| EPI_ISL_12701860 | Clinical Microbiology, Infection Prevention and Control | Section for Molecular Diagnostics | Björn Hallström; Jonas Björkman |
| EPI_ISL_12660480, EPI_ISL_13017314 | Clinique Saint-Pierre Ottignies | UCLouvain/IREC/MBLG-CTMA | Benoit Kabamba Mukadi; Bertrand Bearzatto; Jean-Luc Gala; Valentin Coste |
| EPI_ISL_12226685, EPI_ISL_12226709, EPI_ISL_12660422, EPI_ISL_12660425 | Cliniques universitaires Saint-Luc | UCLouvain/IREC/MBLG-CTMA | Benoit Kabamba Mukadi; Bertrand Bearzatto; Jean-Luc Gala; Valentin Coste |
| EPI_ISL_13020144 | Colorado Department of Public Health and Environment | Colorado Department of Public Health and Environment | Alexandria Rossheim; Arianna Smith; Diana Ir; Emily A. Travanty; Laura Bankers; Mandy Waters; Michael Martin; Molly C. Hetherington-Rauth; Shannon R. Matzinger |
| EPI_ISL_13363318 | DASA | DASA | Adriano Bonaldi; Annelise Lopes; Bianca Cota; Camila Romano; Cristina Oliveira; Jose Levi; Keila Orneles; Laryssa Sassi; Luciane Sussuchi; Paulo Pierry; Rodrigo Guarischi; Rodrigo Salazar |
| EPI_ISL_12248508, EPI_ISL_12286266, EPI_ISL_12317495, EPI_ISL_12401348, EPI_ISL_12435606, EPI_ISL_12472206, EPI_ISL_12473024, EPI_ISL_12533737, EPI_ISL_12558347, EPI_ISL_12558354, EPI_ISL_12558608, EPI_ISL_12609354, EPI_ISL_12610106, EPI_ISL_12632184, EPI_ISL_12632471, EPI_ISL_12632539, EPI_ISL_12648815, EPI_ISL_12648960, EPI_ISL_12648962, EPI_ISL_12649098, EPI_ISL_12727163, EPI_ISL_12811576 | | | |
| see above | Department of Bacteria, Parasites and Fungi, Statens Serum Institut, Copenhagen, Denmark | Statens Serum Institut Bioinformatics and Microbial Genomics | Danish Covid-19 Genome Consortium |
| EPI_ISL_12812558, EPI_ISL_12812568 | Department of Health Technology and Informatics, The Hong Kong Polytechnic University | Department of Health Technology and Informatics, The Hong Kong Polytechnic University | Alan Ka-Lun Wu; Alex Yat-Man Ho; Barry Kin-Chung Wong; Chloe Toi-Mei Chan; David Ho-Keung Shum; Gilman Kit-Hang Siu; Hiu-Yin Lao; Ivan Tak-Fai Wong; Jake Siu-Lun Leung; Kam-Tong Yip; Kenneth Siu-Sing Leung; Kingsley King-Gee Tam; Kitty Sau-Chun Fung; Kristine Luk; Lam-Kwong Lee; Miranda Chong-Yee Yau; Sandy Ka-Yee Chau; Shea Ping Yip; Tak-Lun Que; Timothy Ting-Leung Ng; Wing Cheong Yam; Wing-Hei Lo; Wing-Kin To; Yvette Wai-Man Lai |
| EPI_ISL_13088328 | Department of Laboratory Services, National Virology Reference Laboratory | Clinical Molecular Diagnostic Laboratory For Infectious Disease, Department of Laboratory Services, Microbial Genomic Services | Amal Nabihah Ahmad; Faezah Fariha Abd Latif; Haziq Momin; Izzati Azhar; Nor Azian Hafneh; Nur Amirah Ibarahim; Zainun Zaini |
| EPI_ISL_12471284, EPI_ISL_12715948 | Department of Medical Microbiology & Infection prevention, Amsterdam University Medical Centers location AMC | Department of Medical Microbiology & Infection prevention, Amsterdam University Medical Centers location AMC | Akke Cornelissen; Fokla Zorgdrager; Janke Schinkel; Jelle Koopsen; Judith den Uil; Marcel Jonges; Matthijs Welkers; Menno de Jong; Robin van Houdt; Sebastien Matamoros; Sjoerd Rebers; Sylvia Bruisten; Tjalling Leenstra and Mariken van der Lubben on behalf of the Amsterdam Regional Genomic epidemiology and Outbreak Surveillance (ARGOS) consortium |
| EPI_ISL_13278440 | Department of Veterinary Science and Department of Virology I, National Institute of Infectious Diseases | Research Center for Influenza and Respiratory Viruses, National Institute of Infectious Disease | Hideka Miura; Hideki Ebihara; Hideki Hasegawa; Ikuyo Takayama; Kaya Miyazaki; Ken Maeda; Mutsuyo Takayama-Ito; Seiichiro Fujisaki; Shiho Nagata; Shinji Watanabe; Shuetsu Fukushi; Takahiro Maeki; Tsukasa Yamamoto; Yudai Kuroda |
| EPI_ISL_13024747, EPI_ISL_13086515 | Division of Emerging Infectious Diseases, Bureau of Infectious Diseases Diagnosis Control, Korea Disease Control and Prevention Agency | Division of Emerging Infectious Diseases, Bureau of Infectious Diseases Diagnosis Control, Korea Disease Control and Prevention Agency | Ae Kyung Park; Chae Young Lee; Eun-Jin Kim; Hyuck Jin Lee; Il-Hwan Kim; Jeong-Ah Kim |
| EPI_ISL_12751210 | Dr. Mustafa, Dr. Richter Labor für medizinisch-chemische und mikrobiologische Diagnostik GmbH, Abteilung Molekularbiologie | Dr. Mustafa, Dr. Richter Labor für medizinisch-chemische und mikrobiologische Diagnostik GmbH, Abteilung Molekularbiologie | Alexander Gamisch; Maria Elisabeth Mustafa |
| EPI_ISL_12782902, EPI_ISL_12783249, EPI_ISL_12953365, EPI_ISL_12953676, EPI_ISL_12953692, EPI_ISL_12953764, EPI_ISL_12953785, EPI_ISL_12953795, EPI_ISL_12953796 | | | |
| see above | Dutch COVID-19 response team | National Institute for Public Health and the Environment (RIVM) | Adam Meijer; Afke Vogelzang; AnneMarie van den Brandt; Annelies Kroneman; Bas van der Veer; Chantal Reusken; Dennis Schmitz; Dirk Eggink; Florian Zwagemaker; Harry Vennema; Ivo van Walle; Jeroen Cremer; Jil Kocken; Jordy de Bakker; Karim Hajji; Kim Freriks; Linda van Someren; Lisa Wijsman; Lynn Aarts; Ryanne Jaarsma; Sanne Bos; Sharon van den Brink; on behalf of the national COVID-19 response team |
| EPI_ISL_12832192, EPI_ISL_12832194 | Edmonton Provincial Lab | Alberta Precision Labs (APL) | Buss E; Croxen M; Deo A; Dieu P; Ferrato C; Gill K; Granger D; Koleva P; Li V; Lloyd C; Lynch T; Ma R; Murphy S; Pabbaraju K; Rotich S; Shideler S; Shokopoles S; Skitsko T; Thayer J; Tipples G; Wong A; Yu C; Zelyas N. |
| EPI_ISL_12624687, EPI_ISL_12624817, EPI_ISL_12714036, EPI_ISL_12714066 | Enfer | Enfer | Elaine M. Kenny; Suzie Coughlan |
| EPI_ISL_12589338, EPI_ISL_12706344, EPI_ISL_12706389, EPI_ISL_12706401 | Eurofins-NMDL | Eurofins-NMDL | Anco Molijn; Anne Vogel; Lisa Dreesens; Marvin Ruiter; Maurine Leversteijn-van Hall; Roy Masius; Simon Lansu |
| EPI_ISL_13344356 | Fundación Cardiovascular de Colombia Zona Franca | Molecular Genetics and Antimicrobial Resistance - UGRA, Universidad El Bosque | Alexandra Parada; Jinnethe Reyes; Lorena Diaz; Marcela Mercado; Mauricio Pacheco; Nicolas Forero; Sandra Rincon; Yordy Rodriguez |
| EPI_ISL_12688305 | Genetica Molecular and Subdepartamento de Virologia ISP Chile | Instituto de Salud Publica de Chile | Andres Castillo; Barbara Parra; Constanza Campano; Ivan Ponce; Jorge Fernandez; Karen Orostica; Marcelo Rojas; Matias Pezoa; Patricia Bustos; Rodrigo Fasce |
| EPI_ISL_12954183 | Good Sherperd Hospital | National Institute for Communicable Diseases of the National Health Laboratory Service | Amoako DG; Bhiman JN; Everatt J; Ismail A; Kekana D; Mahlangu B; Maphalala G; Mnguni A; Mohale T; Ntuli N; Scheepers C; Wolter N |
| EPI_ISL_12475182, EPI_ISL_12475185 | HOSPITAL UNIVERSITARIO SON ESPASES | HOSPITAL UNIVERSITARIO SON ESPASES | Dr. Antonio Oliver; Dr. Carla López-Causapé; Dr. Gabriel Cabot; Hospital Universitario Son Espases; on behalf of Servicio de Microbiología |
| EPI_ISL_12339502 | Haaglanden Medisch Centrum | Leiden University Medical Center | Stefan Boers |
| EPI_ISL_12638056, EPI_ISL_12740321, EPI_ISL_12786084, EPI_ISL_12786412 | Helix | Centers for Disease Control and Prevention Division of Viral Diseases, Pathogen Discovery | Benjamin Rambo-Martin; Christopher Gulvick; Clinton Paden; Dakota Howard; Dhwani Batra; Duncan MacCannell; Erisa Sula; Helix CA; Jason Caravas; Kristine Lacek; Matthew Schmerer; Peter Cook; Scott Sammons; Shatavia Morrison; Tymeckia Kendall; Victoria Caban Figueroa; Yvette Unoarumhi |
| EPI_ISL_12605801 | Histopath | NSW Health Pathology - Institute of Clinical Pathology and Medical Research; Westmead Hospital; University of Sydney | Arnott A.; Draper J.; Gall M.; Martinez E.; Rockett R.; Sintchenko V.; on behalf of ICPMR |
| EPI_ISL_12582288, EPI_ISL_13372671 | Hopital | National Reference Center for Viruses of Respiratory Infections, Institut Pasteur, Paris | Angela Brisebarre; Aurélie GUIGON; Camille Capel; Christophe Malabat; Corinne Maufrais; Etienne Simon-Lorière; Frédéric Lemoine; Julien Fumey; Louise Lefrançois; Marion Barbet; Maud Vanpeene; Méline Bizard; Slim El Khiari; Sylvie Behillil; Sylvie Van der Werf; Vincent Enouf |
| EPI_ISL_12703161 | Hospital Center Luxembourg | Laboratoire national de sante, Microbiology, Microbial Genomics Platform | Anke Wienecke-Baldacchino; Catherine Ragimbeau; Elodie Solarino; Eric Hugoson; Fatu Djabi; Jessica Tapp; Lise Pignon; Raoul Salmon; Sibel Berger; Tamir Abdelrahman; Trung Nguyen Nguyen; Virginie Jover |
| EPI_ISL_12687970 | Hospital General Universitario Gregorio Marañón | Hospital General Universitario Gregorio Marañón | Cristina Rodriguez-Grande; Daniel Peñas Utrilla; Darío García de Viedma; Jorge Rodríguez-Grande; Julia Suárez; Laura Pérez-Lago; Marta Herranz Martin; Patricia Muñoz; Pedro Sola Campoy; Pilar Catalán; Rosalía Palomino Cabrera; Sergio Buenestado Serrano |
| EPI_ISL_12717878, EPI_ISL_13014083 | Institute for Infectious Diseases | Institute for Infectious Diseases, University of Bern | Alban Ramette; Christian Baumann; Cora Sägesser; Franziska Suter-Riniker; Lea Stauber; Loïc Borcard; Miguel A Terrazos Miani; Pascal Bittel; Peter Keller; Sonja Gempeler; Stefan Neuenschwander; Stephen L Leib |
| EPI_ISL_12483994 | Jessa | Jessa | Rita Smets et al. on behalf of the Jessa_cmdLab |
| EPI_ISL_12963450, EPI_ISL_12965004 | Kaiser Permanente Southern California | Helix | Helix; Kaiser Permanente Southern California |
| EPI_ISL_12730831, EPI_ISL_12749785 | LABM SAINT BENOIT | Laboratoire de virologie, CNR arbovirus Associé, Chu de la Réunion | Anne-Julie Gourdé; Etienne Frumence; Marie-Christine Jaffar Bandjee; Nicolas M'nemosyme; Nicolas Traversier; Rubens Lhonneur |
| EPI_ISL_11994300 | LABM VIALATTE ESPACE SANTE | CERBA HealthCare | Bénédicte Roquebert; Laura Verdurme; Mathilde Roussel; Sabine Trombert; Stéphanie Haim-Boukobza |
| EPI_ISL_12755936 | LABORATOIRE BIO-VAL | CNR Virus des Infections Respiratoires - France SUD | Antonin Bal; Bruno Lina; Bruno Simon; Gregory Destras; Gwendolyne Burfin; Hadrien Regue; Laurence Josset; Martine Valette; Quentin Semanas; Theophile Boyer |
| EPI_ISL_12699259, EPI_ISL_12755669 | LABORATOIRE NOVELAB | CNR Virus des Infections Respiratoires - France SUD | Antonin Bal; Bruno Lina; Bruno Simon; Gregory Destras; Gwendolyne Burfin; Hadrien Regue; Laurence Josset; Martine Valette; Quentin Semanas; Theophile Boyer |
| EPI_ISL_13321776 | LPA BESANCON | Department of Virology, Henri Mondor University Hospital, Assistance Publique Hôpitaux de Paris, Université Paris-Est Créteil, INSERM U955 | Alexandre Soulier; Christophe Rodriguez; Elisabeth Trawinski; Guillaume Gricourt; Jean-Michel Pawlotsky; Melissa N'Debi; Slim Fourati; Vanessa Demontant |

| | | | |
|---|---|---|---|
| EPI_ISL_13017778 | Labo Analyses Med | Institut Pasteur | Angela Brisebarre; Camille Capel; Christophe Malabat; Corinne Maufrais; Domitille LEMAN; Etienne Simon-Lorière; Frédéric Lemoine; Julien Fumey; Louise Lefrançois; Marion Barbet; Maud Vanpeene; Méline Bizard; Slim El Khiari; Sylvie Behillil; Sylvie Van der Werf; Vincent Enouf |
| EPI_ISL_12714440 | Labo Analyses Med - Ocealab - Le Tenerio | National Reference Center for Viruses of Respiratory Infections, Institut Pasteur, Paris | Angela Brisebarre; Camille Capel; Christophe Malabat; Corinne Maufrais; Etienne Simon-Lorière; Frédéric Lemoine; Julien Fumey; Karine MICHEZ; Louise Lefrançois; Marion Barbet; Maud Vanpeene; Méline Bizard; Slim El Khiari; Sylvie Van der Werf; Vincent Enouf |
| EPI_ISL_13002482 | Labor Dr. Spranger | Robert Koch Institute | |
| EPI_ISL_13014545 | Labor ZOTZ\|KLIMAS; MVZ Düsseldorf-Centrum | Robert Koch Institute | |
| EPI_ISL_12907221 | Laboratoire MAYMAT | Department of Virology, Henri Mondor University Hospital, Assistance Publique Hôpitaux de Paris, Université Paris-Est Créteil, INSERM U955 | Alexandre Soulier; Christophe Rodriguez; Elisabeth Trawinski; Guillaume Gricourt; Jean-Michel Pawlotsky; Melissa N'Debi; Slim Fourati; Vanessa Demontant |
| EPI_ISL_12295715, EPI_ISL_12914362, EPI_ISL_12914364, EPI_ISL_13055526 | Laboratoire de santé publique du Québec | Laboratoire de santé publique du Québec | Guillaume Bourque; Ioannis Ragoussis; Jesse Shapiro; Mark Lathrop and Judith Fafard on behalf of the CoVSeQ research group; Sandrine Moreira |
| EPI_ISL_12837461, EPI_ISL_12837532 | Laboratoire national de sante, Microbiology, Virology | Microbiology, Microbial Genomics Platform, LNS Laboratoire National De Santé | Anke Wienecke-Baldacchino; Catherine Ragimbeau; Elodie Solarino; Eric Hugoson; Fatu Djabi; Jessica Tapp; Lise Pignon; Raoul Salmon; Sibel Berger; Tamir Abdelrahman; Trung Nguyen Nguyen; Virginie Jover |
| EPI_ISL_13031783 | Laboratoires Reunis | Laboratoire national de sante, Microbiology, Microbial Genomics Platform | Anke Wienecke-Baldacchino; Bernard Weber; Catherine Ragimbeau; Elodie Solarino; Eric Hugoson; Fatu Djabi; Jessica Tapp; Lise Pignon; Raoul Salmon; Sibel Berger; Tamir Abdelrahman; Virginie Jover |
| EPI_ISL_12837868, EPI_ISL_12837880 | Laboratoires Reunis | Microbiology, Microbial Genomics Platform, LNS Laboratoire National De Santé | Anke Wienecke-Baldacchino; Bernard Weber; Catherine Ragimbeau; Elodie Solarino; Eric Hugoson; Fatu Djabi; Jessica Tapp; Lise Pignon; Raoul Salmon; Sibel Berger; Tamir Abdelrahman; Virginie Jover |
| EPI_ISL_12512307, EPI_ISL_12512309 | Laboratorio CQRC | CQRC_QUALITY CONTROL CHEMICAL BIOLOGICAL RISK_AOOR Villa Sofia Cervello Palermo | Broccolo F.; Brunacci G.; Contino F.; Di Gaudio F. |
| EPI_ISL_12169006, EPI_ISL_12169353, EPI_ISL_12756256, EPI_ISL_12757576 | Laboratory Corporation of America | Centers for Disease Control and Prevention Division of Viral Diseases, Pathogen Discovery | Amanda Douglas; Amanda Suchanek; Andrea Throop; Ayla Burns; Benjamin Rambo-Martin; Bobbi Croy; Brian Krueger; Brian Norvell; Christopher Gulvick; Christos Petropoulos; Clinton Paden; Craig Lukasik; Dakota Howard; Debbie Boles; Dhwani Batra; Duncan MacCannell; Eyad Almasri; Goran Stevovic; Howard Engler; Hrushikesh Deshmukh; Jake Humphrey; Jana Schroth; Jason Caravas; Joe Voshell; John Pruitt; Jonathan Meltzer; Jonathan Williams; Kimberly Wagner; Kristine Lacek; Lax Iyer; Lisa Pfefferle; Lyndon Tilson; Manoj Jain; Marcia Eisenberg; Mary Cristobal; Mary Williamson; Matthew Robinson; Matthew Schmerer; Michael Levandoski; Mike Sapeta; Mindy Nye; Minoo Agarwal; Mohan Kolli; Nuthawin Charoensri; Oren Cohen; Peter Cook; Prashant Gupta; Qian Zeng; Rama Ghatti; Scott Parker; Scott Ryan; Scott Sammons; Shatavia Morrison; Stanley Letovsky; Steven Ragan; Suresh Selvaraju; Susan Countryman; Susan Hicks; Suzanne Dale; Thomas Urban; Tim Kuphal; Tricia Zwiefelhofer; Tymeckia Kendall; Victoria Caban Figueroa; Vincent Drouillon; Yvette Unoarumhi |
| EPI_ISL_11984862, EPI_ISL_12292992, EPI_ISL_12293148, EPI_ISL_12293149, EPI_ISL_12293160, EPI_ISL_12293161, EPI_ISL_12293281, EPI_ISL_12293283, EPI_ISL_12293292, EPI_ISL_12472109, EPI_ISL_12472111, EPI_ISL_12472129, EPI_ISL_12472136, EPI_ISL_12610583, EPI_ISL_12610598, EPI_ISL_12610609 | | | |
| see above | Lifebrain Covid Labor GmbH | Lifebrain Covid Labor GmbH | Abhishek Mitra; Alexandra Wagner; Anna Edermayr; Felix Valentin Spiegel; Filip Sima; Florian Scharhauser; Hannes Hagen; Kristina Bavrka Kolenc; Lucia Castello; So Jung Han |
| EPI_ISL_11604519, EPI_ISL_12478907, EPI_ISL_12482009, EPI_ISL_12515844, EPI_ISL_12515952, EPI_ISL_12548001, EPI_ISL_12550282, EPI_ISL_12553282, EPI_ISL_12555769, EPI_ISL_12570336, EPI_ISL_12630820, EPI_ISL_12630919, EPI_ISL_12630959, EPI_ISL_12647695, EPI_ISL_13130112 | | | |
| see above | Lighthouse Lab in Glasgow | Wellcome Sanger Institute for the COVID-19 Genomics UK (COG-UK) Consortium | Anna Dominiczak and Alex Alderton; Carol Clugston; Cordelia Langford; David Gray; David K. Jackson; Dominic Kwiatkowski; Ewan Harrison; Harper VanSteenhouse; Ian Johnston; Jeffrey Barrett; John Sillitoe on behalf of the Wellcome Sanger Institute COVID-19 Surveillance Team; Roberto Amato; Sonia Goncalves; Yumi Kasai |
| EPI_ISL_12675976 | Limbach - MVZ Clotten Labor Freiburg Labor Dr. Haas Dr. Raif & Kollegen GbR | Robert Koch Institute | |
| EPI_ISL_12671280 | Limbach - MVZ Labor Dr. Volkmann & Kollegen | Robert Koch Institute | |
| EPI_ISL_13003774 | Limbach - MVZ Labor Ravensburg Labor Dr. Gärtner | Robert Koch Institute | |
| EPI_ISL_12821530 | Limbach - MVZ Labor Westmecklenburg Schmudlach-Oswald-Kettermann & Kollegen | Robert Koch Institute | |
| EPI_ISL_12755245, EPI_ISL_12755263, EPI_ISL_12892789 | MIRIALIS CLUSES BECHET | CNR Virus des Infections Respiratoires - France SUD | Antonin Bal; Bruno Lina; Bruno Simon; Gregory Destras; Gwendolyne Burfin; Hadrien Regue; Laurence Josset; Martine Valette; Quentin Semanas; Theophile Boyer |
| EPI_ISL_12733100 | MVZ Labor Dr. Limbach & Kollegen GbR | Robert Koch Institute | |
| EPI_ISL_12844498 | Max von Pettenkofer Institute, Virology, National Reference Center for Retroviruses, LMU Munich | Laboratory for Functional Genome Analysis (LAFUGA), Gene Center of the LMU Munich | Alexander Graf; Helmut Blum; Max Muenchhoff; Oliver Keppler; Stefan Krebs |
| EPI_ISL_12954184 | Mbabane Government Hospital | National Institute for Communicable Diseases of the National Health Laboratory Service | Amoako DG; Bhiman JN; Everatt J; Ismail A; Kekana D; Mahlangu B; Maphalala G; Mnguni A; Mohale T; Ntuli N; Scheepers C; Wolter N |
| EPI_ISL_12954185, EPI_ISL_12954186 | Mbabane Public Health Unit | National Institute for Communicable Diseases of the National Health Laboratory Service | Amoako DG; Bhiman JN; Everatt J; Ismail A; Kekana D; Mahlangu B; Maphalala G; Mnguni A; Mohale T; Ntuli N; Scheepers C; Wolter N |
| EPI_ISL_12767686 | Michigan Department of Health and Human Services, Bureau of Laboratories | Michigan Department of Health and Human Services, Bureau of Laboratories | Blankenship HM; Riner D; Soehnlen MK |
| EPI_ISL_12628241 | Microbiologia CATLAB | Can Ruti SARS-CoV-2 Sequencing Hub (HUGTiP/IrsiCaixa/IGTP) | Alexia París; Ana Blanco; Andreu Coello; Antoni E Bordoy; Bonaventura Clotet; David Panisello; Francesc Catala-Moll; Gemma Clara; Ignacio Blanco; Laia Soler; Marc Noguera-Julian; Montserrat Giménez; Pere-Joan Cardona; Pilar Armengol; Roger Paredes; Sara González; Verónica Saludes; and Elisa Martró on behalf of the Can Ruti SARS-CoV-2 Sequencing Hub |
| EPI_ISL_12979827, EPI_ISL_12979841 | Microbiological Diagnostic Unit - Public Health Laboratory (MDU-PHL) | Microbiological Diagnostic Unit - Public Health Laboratory (MDU-PHL) | Horan, K.; N.L.; Seemann, T.; Sherry |
| EPI_ISL_12849528 | Microbiological Diagnostic Unit - Public Health Laboratory (MDU-PHL), The Peter Doherty Institute for Infection and Immunity | Microbiological Diagnostic Unit - Public Health Laboratory (MDU-PHL), The Peter Doherty Institute for Infection and Immunity | Horan, K.; N.L.; Seemann, T.; Sherry |
| EPI_ISL_12850230 | Microbiological Diagnostic Unit - Public Health Laboratory (MDU-PHL), The Peter Dororthy Institute for Infection and Immunity | Microbiological Diagnostic Unit - Public Health Laboratory (MDU-PHL), The Peter Dororthy Institute for Infection and Immunity | Horan, K.; N.L.; Seemann, T.; Sherry |
| EPI_ISL_12628235, EPI_ISL_12628250, EPI_ISL_12628267, EPI_ISL_12628303 | Microbiology Department, Laboratori Clínic Metropolitana Nord. Hospital Universitari Germans Trias i Pujol | Can Ruti SARS-CoV-2 Sequencing Hub (HUGTiP/IrsiCaixa/IGTP) | Alexia París; Ana Blanco; Andreu Coello; Antoni E Bordoy; Bonaventura Clotet; David Panisello; Francesc Catala-Moll; Gemma Clara; Ignacio Blanco; Laia Soler; Marc Noguera-Julian; Montserrat Giménez; Pere-Joan Cardona; Pilar Armengol; Roger Paredes; Sara González; Verónica Saludes; and Elisa Martró on behalf of the Can Ruti SARS-CoV-2 Sequencing Hub |
| EPI_ISL_12642823 | Microbiology Department. Complexo Hospitalario Universitario de Vigo | Microbiology Department. Complexo Hospitalario Universitario de Vigo | Microbiology Department. Complexo Hospitalario Universitario de Vigo |
| EPI_ISL_12845550 | National Health Laboratory Services | CERI, Centre for Epidemic Response and Innovation, Stellenbosch University and KRISP, KZN Research Innovation and Sequencing Platform, UKZN. | Anyaneji UJ; Giandhari J; Maharaj A; Moir M; Naidoo Y; Nokukhanya Mdlalose; Pillay S; San JE; Sanko TJ; Tegally H; Tshiabuila D; Van Wyk S; Wilkinson E; de Oliveira T |
| EPI_ISL_12043264, EPI_ISL_12474407 | National Health Laboratory Services, Virology | National Health Laboratory Services, Virology | Ashlyn S. C. Davis; Florette K. Treurnicht; Kathleen Subramoney; Nkhensani Mtileni |
| EPI_ISL_12252953 | National Platform bis UMONS / Jolimont | National Platform bis UMONS / Jolimont | Caroline Debecker; Clothilde Claus; Eric Tarantino; Florian Juszczak; Gautier Detry; Laetitia Gheysen; Ruddy Wattiez |
| EPI_ISL_12533200, EPI_ISL_12835584 | National Platform bis UMONS/Jolimont | National Platform bis UMONS/Jolimont | Caroline Debecker; Clothilde Claus; Eric Tarantino; Florian Juszczak; Gautier Detry; Laetitia Gheysen; Ruddy Wattiez |
| EPI_ISL_12647216, EPI_ISL_12647217, EPI_ISL_12689375 | National Public Health Laboratory, National Centre for Infectious Diseases | National Public Health Laboratory, National Centre for Infectious Diseases | BeiBei Chen; Benny Yeo; Chen Shi Ling; Grace Ngan; Jesslin Tan; Lin Cui; Raymond Tzer Pin Lin; Royce Ang; Samuel Loo; Yichen Ding; Zhenyang Zhou |

| | | | |
|---|---|---|---|
| EPI_ISL_13186295, EPI_ISL_13186587, EPI_ISL_13280019, EPI_ISL_13280073, EPI_ISL_13280158, EPI_ISL_13280186, EPI_ISL_13280229, EPI_ISL_13280310, EPI_ISL_13280312 | | | |
| see above | National Virus Reference Laboratory | National Virus Reference Laboratory | Charlene Bennett; Cillian F De Gascun; Gabriel Gonzalez; Jonathan Dean; Michael Carr; Zoe Yandle |
| EPI_ISL_12623503 | New Brunswick - Vitalite Health Network | New Brunswick - Vitalite Health Network | Allain E.; Chacko S.; Crapoulet N.; Desnoyers G.; Garceau R.; Lacroix J.; Lyons P.; Shaw W. |
| EPI_ISL_12841626 | New Brunswick - Vitalite Health Network, Dr. Georges-L.-Dumont University Hospital Centre | New Brunswick - Vitalite Health Network, Dr. Georges-L.-Dumont University Hospital Centre | Allain E.; Chacko S.; Crapoulet N.; Desnoyers G.; Garceau R.; Lacroix J.; Lyons P.; Shaw W. |
| EPI_ISL_13079038 | Noble Hospital Samples | INSACOG-IISER Pune | ; Aurnab Ghose; Joy Merwin Monteiro; Krishanpal Karmodiya |
| EPI_ISL_12853572, EPI_ISL_12853587, EPI_ISL_12895004, EPI_ISL_12895028 | Originating lab: Wales Specialist Virology Centre Sequencing lab: Pathogen Genomics Unit | Public Health Wales Microbiology Cardiff Wales Specialist Virology Centre | Alec Birchley; Alexander Adams; Amy Gaskin; Angela Marchbank; Bree Gatica-Wilcox; Catherine Moore; Jason Coombes; Joanne Watkins; Joel Southgate; Johnathan Evans; Laura Gifford; Lauren Gilbert; Lee Graham; Malorie Perry; Matthew Bull; Nicole Pacchiarini; Sally Corden; Sara Kumziene-Summerhayes; Sara Rey; Sarah Taylor; Simon Cottrell; Sophie Jones; Tom Connor |
| EPI_ISL_13010930, EPI_ISL_13010931 | Outre Mer | National Reference Center for Viruses of Respiratory Infections, Institut Pasteur, Paris | Angela Brisebarre; Camille Capel; Christophe Malabat; Corinne Maufrais; Didier MATTERA; Etienne Simon-Lorière; Frédéric Lemoine; Julien Fumey; Louise Lefrançois; Marion Barbet; Maud Vanpeene; Méline Bizard; Slim El Khiari; Sylvie Van der Werf; Vincent Enouf |
| EPI_ISL_13048417, EPI_ISL_13048521 | PORT ELIZABETH LABORATORY | National Institute for Communicable Diseases of the National Health Laboratory Service | Amoako DG; Bhiman JN; Everatt J; Ismail A; Kekana D; Mahlangu B; Mnguni A; Mohale T; Ntuli N; Scheepers C; Wolter N |
| EPI_ISL_12012887 | PathCare, Cape Town | Division of Medical Virology, National Health Laboratory Service (NHLS), Tygerberg Hospital / Stellenbosch University | Gert van Zyl; Jean Maritz; Nadine Cronje; Petra Raimond; Shannon Wilson; Tongai Maponga; Wolfgang Preiser |
| EPI_ISL_12626263, EPI_ISL_12626309, EPI_ISL_12770929, EPI_ISL_12911598 | PathWest Laboratory Medicine WA | PathWest Laboratory Medicine WA Microbial Surveillance Unit | PathWest Laboratory Medicine WA Microbial Surveillance Unit |
| EPI_ISL_12871857 | Pathcare | CERI, Centre for Epidemic Response and Innovation, Stellenbosch University and KRISP, KZN Research Innovation and Sequencing Platform, UKZN. | Anyaneji UJ; Claassen M; Giandhari J; Maharaj A; Maponga T; Moir M; Naidoo Y; Pillay S; Preiser W; San JE; Sanko TJ; Stander T; Tegally H; Tshiabuila D; Van Wyk S; Wilkinson E; Wilson S; de Oliveira T; van Zyl G |
| EPI_ISL_12905595 | Pathologist Lancet Kenya | KEMRI-Wellcome Trust Research Programme,Kilifi | Agoti C.; D.J.Nokes; Githinji G.; Lambisia A.; Makori T.; Mburu M.W.; Mohamed K.S.; Morobe J.; Mukadam R; Munoko A.; Ndwiga L.; Ngari C.; Ochola I; Ongera E.; de Laurent Z. |
| EPI_ISL_12605042, EPI_ISL_12605043 | Pathology North - Royal North Shore Hospital - NSW Health Pathology | NSW Health Pathology - Institute of Clinical Pathology and Medical Research; Westmead Hospital; University of Sydney | Arnott A.; Draper J.; Gall M.; Martinez E.; Rockett R.; Sintchenko V.; on behalf of ICPMR |
| EPI_ISL_12613687, EPI_ISL_12648066 | Plateforme de testing Namuroise | Plateforme de testing Namuroise | Bleret Leonore; Degosserie Jonathan; Denis Olivier; Gilliard Nicolas; Lesly Nyinkeu Kemamen; Louise Janssens; Maschietto Céline; Mullier François; Otto Gaetan; Pellet Nathan; Renguet Edith |
| EPI_ISL_12683708 | Platform BIS UZA/UAntwerpen | Labo Klinische Biologie, UZA | Basil Britto Xavier; Christine Lammens; Herman Goossens; Ines Verbesselt; Jasmine Coppens; Kathleen Holemans; Marie Le Mercier; Silke Liers; Veerle Matheeussen |
| EPI_ISL_12571627, EPI_ISL_12715870, EPI_ISL_12715916, EPI_ISL_12715923 | Public Health Laboratory, Public Health Service Amsterdam, The Netherlands | Department of Medical Microbiology & Infection prevention, Amsterdam University Medical Centers location AMC | Akke Cornelissen; Fokla Zorgdrager; Janke Schinkel; Jelle Koopsen; Judith den Uil; Marcel Jonges; Matthijs Welkers; Menno de Jong; Robin van Houdt; Sebastien Matamoros; Sjoerd Rebers; Sylvia Bruisten; Tjalling Leenstra and Mariken van der Lubben on behalf of the Amsterdam Regional Genomic epidemiology and Outbreak Surveillance (ARGOS) consortium |
| EPI_ISL_13149254, EPI_ISL_13149259, EPI_ISL_13149261, EPI_ISL_13149273, EPI_ISL_13149279, EPI_ISL_13149280, EPI_ISL_13149284, EPI_ISL_13149291, EPI_ISL_13149293, EPI_ISL_13149297, EPI_ISL_13149303, EPI_ISL_13149305, EPI_ISL_13149318 | | | |
| see above | Public Health Laboratory: COVID-19 Lab | International Livestock Research Institute | Collins Muli; Daniel Ouso; Edward Kiritu; Edward O. Abworo; Gilbert Kibet; Gugu Maphalala; Mncedisi Hlophe; Nomcebo Phungwayo; Patrick Amoth; Paul Dobi; Samuel O. Oyola; Shebbar Osiany; Siphesihle Langwenya; Sonal P. Henson; Susan Kamalizeni; Vishvanath Nene |
| EPI_ISL_12266791, EPI_ISL_12466376, EPI_ISL_12627205 | Public Health Ontario Laboratory | Public Health Ontario Laboratory | Aimin Li; Alex Marchand-Austin; Andre Villegas; Anna Puzinovici; Ashleigh Sullivan; Brandon Ye; Candice Schreiber; Carla Duncan; Christina Rampertab; Christine Seah; Claudia Chu; Dean Maxwell; Dhiraj Gaglani; Doonia Bajovic; Esther Nagai; Fatemeh Shaeri; Fatima Merza; Grace Jeong; Hadia Hussain; Himeshi Samarsinghe; Jacob Afelskie; Jason Iraheta; Jesse Wang; John Palmer; Karthikeyan Sivaraman; Kirby Cronin; Lisa Kim; Lisa McTaggart; Maria Mariscal; Mark Horsman; Narisha Shakuralli; Nataliya Potapova; Natasha Sing; Nobish Varghese; Philip Banh; Rachelle DiTullio; Rebecca Azzaro; Rima Palencia; Samir N Patel; Sarah Teatero; Semra Tibebu; Sophie Yu; Surendra Kumar; Sushma Kavikondala; Vincent Su Bin Cha; Zarah Rajaei |
| EPI_ISL_12932497 | Queensland Medical Laboratories | PHV-FSS | Chenwei Wang on behalf of Q-PHIRE Genomics |
| EPI_ISL_12660069, EPI_ISL_12874244, EPI_ISL_12874769, EPI_ISL_13056807 | Quest Diagnostics Incorporated | Centers for Disease Control and Prevention Division of Viral Diseases, Pathogen Discovery | A. Gerasimova; A. Perez; B. Anderson; Benjamin Rambo-Martin; Christopher Gulvick; Clinton Paden; Dakota Howard; Dhwani Batra; Duncan MacCannell; Erisa Sula; F. Lacbawan; I. Shlyakhter; Jason Caravas; K. Livingston; Kristine Lacek; L. Bernstein; M. Hua; Matthew Schmerer; P. Tanpaiboon; Peter Cook; R. Kagan; R. Owen; R. Rolando; S. Rosenthal; Scott Sammons; Shatavia Morrison; Tymeckia Kendall; Victoria Caban Figueroa; Y. Liu; Yvette Unoarumhi |
| EPI_ISL_13089278 | ROMILLY DYNALAB | Department of Virology, Henri Mondor University Hospital, Assistance Publique Hôpitaux de Paris, Université Paris-Est Créteil, INSERM U955 | Alexandre Soulier; Christophe Rodriguez; Elisabeth Trawinski; Guillaume Gricourt; Jean-Michel Pawlotsky; Melissa N'Debi; Slim Fourati; Vanessa Demontant |
| EPI_ISL_12704014 | Regional Virus Laboratory, Belfast Health and Social Care Trust; and: Genomics Core Technology Unit, Queen's University Belfast. | COVID-19 Genomics UK (COG-UK) Consortium | Alan; Alison Watt; Arun Mahesh; BHSCT]: Conall McCaughey; Ciara Cox; Clara Radulescu; David Simpson; Deborah Lavin; Derek Fairley; Evan Troendle; Fiona Rogan; James McKenna; Jana Gazdova; Julia Miskelly; Mairead Connor; Miao Tang; QUB]: Marc Fuchs; Rice; Sarah Sonner; Stephen Bridgett; Susan Feeney; Syed Umbreen; Tanya Curran; Timofey Skvortsov; Zoltan Molnar; [Genomics Core Technology Unit; [Regional Virus Laboratory |
| EPI_ISL_12152675, EPI_ISL_12241466, EPI_ISL_12469666, EPI_ISL_12469747, EPI_ISL_12469892, EPI_ISL_12470002, EPI_ISL_12514770, EPI_ISL_12515007, EPI_ISL_12515119, EPI_ISL_12606310, EPI_ISL_12606668, EPI_ISL_12606838, EPI_ISL_12606881, EPI_ISL_12606913, EPI_ISL_12667094, EPI_ISL_12694146, EPI_ISL_12698346 | | | |
| see above | Rosalind Franklin Laboratory | Wellcome Sanger Institute for the COVID-19 Genomics UK (COG-UK) Consortium | Cordelia Langford; David K. Jackson; Dominic Kwiatkowski; Donald Fraser; Ewan Harrison; Ian Johnston; Jeffrey Barrett; John Sillitoe on behalf of the Wellcome Sanger Institute COVID-19 Surveillance Team; Rob Howes; Roberto Amato; Sonia Goncalves; Suki Lee; The Rosalind Franklin Laboratory and Alex Alderton |
| EPI_ISL_12442764, EPI_ISL_13045384, EPI_ISL_13045388, EPI_ISL_13045498, EPI_ISL_13045602, EPI_ISL_13045673, EPI_ISL_13045755 | | | |
| see above | SA Pathology | SA Pathology | Caitlin Selway; Chuan Kok Lim; Ivan Bastian; Lex Leong; Mark Turra |
| EPI_ISL_12724716, EPI_ISL_12807071, EPI_ISL_13026924 | SARS-CoV-2 testing team, National Institute of Infectious Diseases | Pathogen Genomics Center, National Institute of Infectious Diseases | Hazuka Y Furihata; Kentaro Itokawa; Makoto Kuroda; Masanori Hashino; Masumichi Saito; Naomi Nojiri; Nozomu Hanaoka; Rina Tanaka; Tsuguto Fujimoto; Tsuyoshi Sekizuka |
| EPI_ISL_12959393 | SD Public Health Laboratory | Centers for Disease Control and Prevention Division of Viral Diseases, Pathogen Discovery | Alex Burgin; Ben Rambo-Martin; Clinton Paden; Dakota Howard; Dave Wentworth; Dhwani Batra; Jasmine Padilla; Joseph Madden; Justin Lee; Kristen Knipe; Kristine Lacek; Mark Burroughs; Matthew Schmerer; Meghan Bentz; Mili Sheth; Peter Cook; Sam Shepard; Sarah Nobles; Vivien Dugan; Yvette Unoarumhi |
| EPI_ISL_12856291 | SELAS BC-LAB | Department of Virology, Henri Mondor University Hospital, Assistance Publique Hôpitaux de Paris, Université Paris-Est Créteil, INSERM U955 | Alexandre Soulier; Christophe Rodriguez; Elisabeth Trawinski; Guillaume Gricourt; Jean-Michel Pawlotsky; Melissa N'Debi; Slim Fourati; Vanessa Demontant |
| EPI_ISL_12335014 | SYNLAB MVZ Leverkusen | Robert Koch Institute | |
| EPI_ISL_12520814, EPI_ISL_12674190, EPI_ISL_12733204 | SYNLAB MVZ Weiden | Robert Koch Institute | |
| EPI_ISL_12871407, EPI_ISL_12877262, EPI_ISL_12877455, EPI_ISL_12902154 | Shamir Medical Center (Asaf Harofe) | Shamir Medical Center (Asaf Harofe) | Abu Hamad Ramzia; Adina Bar Chaim; Anna Vishnevsky; Chen Weiner; Nir Rainy; Patricia Benveniste-Lekovitz; Reut Sorek Abramovich; Yevgeni Yegorov |
| EPI_ISL_13248991 | Sonic - Labor Staber Dresden (Klipphausen) | Robert Koch Institute | |
| EPI_ISL_12605102 | St Vincent's Pathology (SydPath) | NSW Health Pathology - Institute of Clinical Pathology and Medical Research; Westmead Hospital; University of Sydney | Arnott A.; Draper J.; Gall M.; Martinez E.; Rockett R.; Sintchenko V.; on behalf of ICPMR |
| EPI_ISL_12587465 | Stadtspital Triemli | Institute of Medical Virology, University of Zurich | Alexandra Trkola; Annette Audigé; Catharine Aquino; Cyril Shah; Daniel Ehrsam; Gabriela Ziltener; Guido Bloemberg; Hubert Rehrauer; Isabel Stürmer; Joel Wirz; Jon Huder; Jürg Böni; Kevin Steiner; Maria Grünberg; Maryam Zaheri; Michael Huber; Riccarda Capaul; Stefan Schmutz; Verena Kufner; Weihong Qi |
| EPI_ISL_12785355 | State Public Health Laboratory O/o DPH&PM, Chennai | CSIR-NEERI, Nagpur Covid-19 Testing Lab | Krishna Khairnar et al. |
| EPI_ISL_13184869, | Swedish national genomic | The Public Health Agency of Sweden | Alma Brolund; Emmi Andersson; Maria Lind Karlberg; Swedish national genomic surveillance program of SARS-CoV-2 |

| | | | |
|---|---|---|---|
| EPI_ISL_13185323, EPI_ISL_13185598 | surveillance program of SARS-CoV-2 | | |
| EPI_ISL_12898417 | Switch Health | National Microbiology Laboratory (NML) | Adrian Zetner; Anna Majer; Anneliese Landgraff; CanCOGeN's metadata curation team; Carmen Lia Murall; Chanchal Yadav; Connor Chato; Darian Hole; Elsie Grudeski; Emily Haidl; Gary Van Domselaar; Gordon Jolly; Grace Seo; Jeff Tuff; Jennifer Tanner; Katherine Eaton; Kirsten Biggar; Kristyn Burak; Madison Chapel; Morag Graham; Natalie Knox; Nathalie Bastien; Philip Mabon; Public Health Agency of Canada's CCGP and Scientific Informatics Services team; Rhiannon Huzarewich; Russell Mandes; Shari Tyson; Timothy Booth; Yan Li |
| EPI_ISL_13029523 | Synlab Eesti OÜ | 1. Laboratory of Communicable Diseases (Estonia); 2. Eurofins Genomics Europe Sequencing GmbH | Abroi A.; Avi R.; Dotsenko L.; Epštein J.; Hoidmets D.; Huik K.; Härma M-A.; Jaaniso E.; Kaarna K.; Kallas E.; Koppel I.; Kuzmin I.; Lahesaare A.; Lutsar I.; Metspalu M.; Milani L.; Naaber P.; Niglas H.; Oopkaup O.E.; Pauskar M.; Peterson H.; Päll T.; Ratnik K.; Raudvere U.; Reisberg T.; Sadikova O.; Sepp H.; Shablinskaja A.; Suija H.; Talas U.G.; Truusalu K. |
| EPI_ISL_13207909 | Taksin hospital | Medical Genomic Centre,Medical Life Sciences Institute,Department of Medical Sciences, Ministry of Public Health, Thailand | Archawin Rojanawiwat; Jirapha Pakdee; Naphatcha Thawong; Natthakul Bunneang; Nuanjun Wichukchinda; Pilailuk Akkapaiboon Okada; Pundharika Piboonsiri; Surakameth Mahasirimongkol; Waritta Sawaengdee |
| EPI_ISL_12771764, EPI_ISL_12771765 | Temporary Specimen Collection Centre at the AsiaWorld-Expo | Hong Kong Department of Health | Alan K.L. Tsang; Edman T.K. Lam; Ken H.L. Ng; Patricia K. L. Leung; Peter C.W. Yip; Rickjason C.W. Chan |
| EPI_ISL_12607382, EPI_ISL_12607392, EPI_ISL_12607398, EPI_ISL_12607412, EPI_ISL_12749708 | U.O. Microbiologia Laboratorio Unico Centro Servizi - AUSL della Romagna | U.O. Microbiologia, Laboratorio Unico Centro Servizi - AUSL della Romagna | Giorgio Dirani |
| EPI_ISL_12252797 | UMC Utrecht | UMC Utrecht | Anne Wensing; Joris Schoonderwoerd; Rob Schuurman |
| EPI_ISL_13330459 | UNILIANS BIOGROUP Décines | Department of Virology, Henri Mondor University Hospital, Assistance Publique Hôpitaux de Paris, Université Paris-Est Créteil, INSERM U955 | Alexandre Soulier; Christophe Rodriguez; Elisabeth Trawinski; Guillaume Gricourt; Jean-Michel Pawlotsky; Melissa N'Debi; Slim Fourati; Vanessa Demontant |
| EPI_ISL_12529661 | UO Microbiologia, IRCCS Az.Ospedaliero-Universitaria di Bologna, Policlinico di S.Orsola | Unità Operativa di Microbiologia, IRCSS Policlinico di Sant'Orsola, Azienda Ospedaliero Universitaria di Bologna | Giada Rossini |
| EPI_ISL_12696584 | USC Clinical Lab | Los Angeles County Public Health Laboratories | J. Garrigues et al. |
| EPI_ISL_12956393 | University Hospitals of Geneva, Laboratory of Virology | HUG, Laboratory of Virology and the Health2030 Genome Center | Aline Mamin; Ana Rita Goncalves; Cedric Howald; Deborah Penet; Francisco Perez; Henri Pegeot; Ioannis Xenarios; Keith Harshman; Laurent Kaiser; Lorenzo Cerutti; Melyssa Elies; Samuel Cordey |
| EPI_ISL_13050826 | Usansolo-Galdakao University Hospital | Usansolo-Galdakao University Hospital | Ana Gual-de-Torrella; Izaskun Alejo-Cancho; Mikel Urrutikoetxea-Gutierrez |
| EPI_ISL_12130073, EPI_ISL_12340151, EPI_ISL_12846026 | Viollier AG | Department of Biosystems Science and Engineering, ETH Zürich | Andrea Patrizia Salzmann; Chaoran Chen; Christian Beisel; Christiane Beckmann; Christoph Noppen; David Dreifuss; Elodie Burcklen; Franziska Singer; Henriette Kurth; Ina Nissen; Ivan Topolsky; Kim Philipp Jablonski; Lara Fuhrmann; Louis du Plessis; Matteo Carrara; Maurice Redondo; Mirjam Feldkamp; Natascha Santacroce; Niko Beerenwinkel; Olivier Kobel; Pelin Icer; Rebecca Denes; Sarah Nadeau; Sebastian Kurscheid; Shuqing Yu; Tanja Stadler; Tobias Schär |
| EPI_ISL_12651663, EPI_ISL_12709399, EPI_ISL_12780237, EPI_ISL_12895107, EPI_ISL_12895207 | West of Scotland Specialist Virology Centre, NHSGGC / MRC-University of Glasgow Centre for Virus Research | COVID-19 Genomics UK (COG-UK) Consortium | Alasdair MacLean; Ana da Silva Filipe; Andy Young; Antonia Ho; Daniel Mair; David L Robertson; Emily Goldstein; Emma Thomson; Gonzalo Yebra; Guy Mollett; Ioulia Tsatsani; James Shepherd; Jenna Nichols; Jessica Benkaroun; Jon Perkins; Jordan Ashworth; Joseph Hughes; Kathy Smollett; Kirsty Mangin; Kyriaki Nomikou; Lily Tong; Matthew Holden; Nicolas Suarez; Rachael Tomb; Rachel Blacow; Richard Orton; Rory Gunson; Sarah McDonald; Sharif Shaaban; Sreenu Vattipally |
| EPI_ISL_12713156 | Worcester Hospital wc WOC | NHLS/UCT | Arash Iranzadeh; Carolyn Williamson; Diana Hardie; Gert Marais; Innocent Mudau; Luicer Olubayo; Marvin Hsiao; Nokuzola Mbhele; Rageema Joseph; Stephen Korsman |
| EPI_ISL_12739598 | Yale Clinical Virology Lab | Grubaugh Lab - Yale School of Public Health | Anne Hahn; Bony De Kumar; Chaney Kalinich; Chantal Vogels; Christopher Castaldi; David Ferguson; David Peaper; Irina Tikhonova; Kendall Billig; Kien Pham; Mallery Breban; Marie L. Landry; Nathan Grubaugh; Nicholas Chen; Nicholas Kerantzas; Rebecca Earnest; Tobias Koch; Wade Schulz |
| EPI_ISL_12580061 | ZOTZ KLIMAS MVZ Düsseldorf-Centrum GbR ÜBAG für Labormedizin, Genetik, Zytologie, Pathologie | Center of Medical Microbiology, Virology, and Hospital Hygiene, University of Duesseldorf | Alexander Dilthey; Andreas Walker; Daniel Strelow; Jessica Nicolai; Jörg Timm; Katrin Hoffmann; Klaus Pfeffer; Lisanna Hülse; Malte Kohns Vasconcelos; Maximilian Damagnez; Nadine Lübke; Patrick Finzer; Rainer Zotz; Tobias Wienemann; Torsten Houwaart |
