## Supplementary material for "Tracing the international arrivals of SARS-CoV-2 Omicron variants after Aotearoa New Zealand reopened its border": BA.5 GISAID acknowledgements

We gratefully acknowledge the following Authors from the Originating laboratories responsible for obtaining the specimens, as well as the Submitting laboratories where the genome data were generated and shared via GISAID, on which this research is based.

All Submitters of data may be contacted directly via www.gisaid.org

Authors are sorted alphabetically.

| Accession ID | Originating Laboratory | Submitting Laboratory | Authors |
| --- | --- | --- | --- |
| EPI_ISL_13227147 | A.O. PERUGIA | A.O. PERUGIA | Bicchieraro G; Bondi P; Camilloni B; Cappelletti E; Ciurnelli R; Lepri E; Lucheroni F; Mencacci A; Spaccapelo R |
| EPI_ISL_13323342 | A.S.L. TO3 | Fondazione del Piemonte per l'Oncologia IRCCS | Antonino Sottile; Giorgio Giardina; Paola Marino; Silvia Brossa |
| EPI_ISL_13108779 | ABC Algarve | Instituto Nacional de Saude Doutor Ricardo Jorge (INSA) | Borges et al |
| EPI_ISL_12763815 | AMPATH | National Institute for Communicable Diseases of the National Health Laboratory Service | Amoako DG; Bhiman JN; Everatt J; Ismail A; Kekana D; Mahlangu B; Mnguni A; Mohale T; Ntuli N; Scheepers C; Wolter N |
| EPI_ISL_13322873 | AOPD | Istituto Zooprofilattico Sperimentale delle Venezie | Adelaide Milani; Alessia Schivo; Alice Fusaro; Ambra Pastori; Angela Salomoni; Annalisa Salviato; Antonia Ricci; Calogero Terregino; Edoardo Giussani; Elisa Palumbo; Erika Giorgia Quaranta; Isabella Monne |
| EPI_ISL_13176072 | AOUIVR | Unità Operativa Complessa di Microbiologia e Virologia Azienda Ospedaliera Universitaria Integrata di Verona; Istituto Zooprofilattico Sperimentale delle Venezie | Adelaide Milani; Alessia Schivo; Alice Fusaro; Angela Salomoni; Annalisa Salviato; Antonia Ricci; Calogero Terregino; Davide Gibellini; Edoardo Giussani; Elisa Palumbo; Erika Giorgia Quaranta; Giona Turri; Isabella Monne; Monica Castellucci; Nicoletta Medaina |
| EPI_ISL_13371594, EPI_ISL_13371618 | ARION GENETICA | Instituto Nacional de Medicina Genomica | Cedro-Tanda A; Escobar-Arrazola MA; Garnica-Lopez Dora; Herrera-Montalvo LA.; Hidalgo-Miranda A; Mendoza-Vargas A; Ramirez-Vega O; Rangel-DeLeon D; Reyes-Grajeda JP; Roldan-Castillo Magaly; Uribe-Figueroa Laura; Verea Jazmin; Yair Alfaro-Mora |
| EPI_ISL_13019093, EPI_ISL_13019106 | ARS Algarve - Laboratorio Laura Ayres | Instituto Nacional de Saude Doutor Ricardo Jorge (INSA) | Borges et al |
| EPI_ISL_12981999, EPI_ISL_13181342, EPI_ISL_13181797 | Akershus University Hospital, Department for Microbiology and Infectious Disease Control | Norwegian Institute of Public Health, Department of Virology | Atiya R Ali; Debech Nadia; Engebretsen Serina Beate; Garcia Llorente Ignacio; Hilde Elshaug; Hilde Nordby Falkenhaug; Hilde Vollan; Jon Bråte; Kamilla Heddeland Instefjord; Karoline Bragstad; Kathrine Stene-Johansen; Line Victoria Moen; Marie Paulsen Madsen; Olav Hungnes; Pedersen Benedikte Nevjen; Rasmus Riis Kopperud |
| EPI_ISL_13202249 | Algemeen Klinisch Labo | Labo Klinische Biologie, UZA | Basil Britto Xavier; Christine Lammens; Herman Goossens; Ines Verbesselt; Jasmine Coppens; Kathleen Holemans; Marie Le Mercier; Silke Liers; Veerle Matheeussen |
| EPI_ISL_12917780 | Ampath | National Institute for Communicable Diseases of the National Health Laboratory Service | Amoako DG; Bhiman JN; Everatt J; Ismail A; Kekana D; Mahlangu B; Mnguni A; Mohale T; Ntuli N; Scheepers C; Wolter N |
| EPI_ISL_13337865 | Area de salud barva (coopesiba) | Inciensa, Instituto Costarricense de Investigación y Enseñanza en Nutrición y Salud | Adriana Godínez; Claudio Soto-Garita; Estela Cordero; Francisco Duarte; Gabriel Morales; Hebleen Porras; José Luis Vargas; Mariela Gutiérrez; Melany Calderón; Natalia Bonilla & Sharon Peñaranda Chanto; Sofia Herrera |
| EPI_ISL_13337850 | Area de salud pavas (coopesalud) | Inciensa, Instituto Costarricense de Investigación y Enseñanza en Nutrición y Salud | Adriana Godínez; Claudio Soto-Garita; Estela Cordero; Francisco Duarte; Gabriel Morales & Natalia Bonilla; Hebleen Porras; José Luis Vargas; Mariela Gutiérrez; Melany Calderón; Sofia Herrera |
| EPI_ISL_13340450 | Austrian Agency for Health and Food Safety (AGES) | Bergthaler laboratory, CeMM Research Center for Molecular Medicine of the Austrian Academy of Sciences | Alberto Alises; Andreas Bergthaler; Anna Schedl; Christoph Bock; Fabian Amman; Lukas Endler; Matthew Thornton; Michael Schuster; Michelle Chan; Petr Triska |
| EPI_ISL_13134758 | BP Healthcare Group | Institute for Medical Research, Infectious Disease Research Centre, National Institutes of Health, Ministry of Health Malaysia | Anasir MI; G.Adypatti NM; Jamaluddin MS; Kalyanasundram J; Kamel K; MatRahim N; Nawi MH; Suib FA; Suppiah J; Thayan R |
| EPI_ISL_13106453, EPI_ISL_13177148, EPI_ISL_13340742 | Basurto University Hospital: Clinical Microbiology Laboratory | Basurto University Hospital: Clinical Microbiology Laboratory | Estibaliz Ugalde Zarraga; José Luis Díaz de Tuesta del Arco; Mikel Urrutikoetxea-Gutiérrez; Mª Carmen Nieto Toboso |
| EPI_ISL_13031192, EPI_ISL_13032045, EPI_ISL_13282427 | BioneXt Lab | Laboratoire national de sante, Microbiology, Microbial Genomics Platform | Anke Wienecke-Baldacchino; Catherine Ragimbeau; Elodie Solarino; Eric Hugoson; Fatu Djabi; Jessica Tapp; Lise Pignon; Raoul Salmon; Sibel Berger; Tamir Abdelrahman; Thibault Ferrandon; Virginie Jover |
| EPI_ISL_13344815, EPI_ISL_13344818, EPI_ISL_13344826, EPI_ISL_13344838, EPI_ISL_13344839 | Botswana Harvard AIDS Institute Partnership | Botswana Harvard HIV Reference Laboratory | Boitumelo Zuze; Botshelo Radibe; Dorcas Maruapula; Joseph Makhema; Keoratile Ntshambiwa; Kgomotso Moruisi; Legodile Kooepile; Mosepele Mosepele; Mphaphi B. Mbulawa; Ontlametse T. Bareng; Pamela Smith-Lawrence; Patrick T. Mokgethi; Roger Shapiro; Sefetogi Ramaologa; Shahin Lockman; Sikhulile Moyo; Simani Gaseitsiwe; Thongbotho Mphoyakgosi; Wonderful T. Choga |
| EPI_ISL_13158767, EPI_ISL_13159538, EPI_ISL_13159551, EPI_ISL_13276609, EPI_ISL_13276611, EPI_ISL_13276629, EPI_ISL_13276630 | | | |
| see above | Bumrungrad International Hospital | National Institute of Health, Department of Medical Sciences, Ministry of Public Health, Thailand | Archawin Rojanawiwat; Natchaya Khiadsang; Nuttida Thongpramul; Pakorn Piromtong; Pilailuk Okada; Sirikanda Wimol; Siripaporn Phuygun; Sunthareeya Waicharoen; Suratchana Mitrat; Thanutsapa Thanadachakul |
| EPI_ISL_12954894, EPI_ISL_12954905 | CENTRAL HEALTH LABORATORY | National Institute for Communicable Diseases of the National Health Laboratory Service | Amoako DG; Bhiman JN; Everatt J; Ismail A; Kekana D; Mahlangu B; Mnguni A; Mohale T; Ntuli N; Scheepers C; Wolter N |
| EPI_ISL_13253710 | CERBALLIANCE OISE | CERBA HealthCare | Bénédicte Roquebert; Laura Verdurme; Mathilde Roussel; Sabine Trombert; Stéphanie Haim-Boukobza |
| EPI_ISL_12731318 | CERBALLIANCE PORT | Laboratoire de virologie, CNR arbovirus Associé, Chu de la Réunion | Anne-Julie Gourdé; Etienne Frumence; Marie-Christine Jaffar Bandjee; Nicolas M'nemosyme; Nicolas Traversier; Rubens Lhonneur |
| EPI_ISL_13253757 | CERBALLIANCE PYRENEES | CERBA HealthCare | Bénédicte Roquebert; Laura Verdurme; Mathilde Roussel; Sabine Trombert; Stéphanie Haim-Boukobza |
| EPI_ISL_13229725, EPI_ISL_13229728 | CH Porto - H Sto Antonio | Instituto Nacional de Saude Doutor Ricardo Jorge (INSA) | Borges et al |
| EPI_ISL_12721258, EPI_ISL_13229798 | CH Setubal | Instituto Nacional de Saude Doutor Ricardo Jorge (INSA) | Borges et al |
| EPI_ISL_12863086 | CH Tamega e Sousa | Instituto Nacional de Saude Doutor Ricardo Jorge (INSA) | Borges et al |
| EPI_ISL_13229615 | CH Tondela Viseu | Instituto Nacional de Saude Doutor Ricardo Jorge (INSA) | Borges et al |
| EPI_ISL_13053240 | CHAZ Laboratory | Churches Health Association of Zambia (CHAZ) Laboratory | CHAZ Lab Staff; Chipango. C; Muyombo. A; Sandala. D; Shempela. D; Sikalima. J |
| EPI_ISL_13229591, EPI_ISL_13229600 | CHTMAD | Instituto Nacional de Saude Doutor Ricardo Jorge (INSA) | Borges et al |
| EPI_ISL_13257781 | Center for laboratory medicine, Clinical Center of Vojvodina | Institute of Molecular Genetics and Genetic Engineering University of Belgrade | Andriana Lazic; Anita Skakic; Bojan Ristivojevic; Ivana Moric; Jelena Stojčević Maletić; Katarina Novovic; Maja Tolinacki; Marija Cumbo; Milka Malesevic; Mina Peric; Mirjana Novkovic; Natasa Radakovic; Natasa Stevanovic; Sandra Vojnovic; Sofija Nesic; Stefan Stanovcic; Valentina Djordjevic |
| EPI_ISL_13341550, EPI_ISL_13360209, EPI_ISL_13360226 | Central Health Laboratory , Victoria Hospital, Ministry of Health and Wellness, Mauritius | CERI, Centre for Epidemic Response and Innovation, Stellenbosch University and KRISP, KZN Research Innovation and Sequencing Platform, UKZN. | Anyaneji UJ; Bahadoor BS; Claassen M; Giandhari J; Issack M; Jannoo N; Maharaj A; Maponga T; Mathur H; Moir M; Naidoo Y; Pillay S; Preiser W; Ramuth M; San JE; Sanko TJ; Sankon TJ; Sonoo J; Stander T; Tegally H; Tshiabuila D; Ubheeram A; Van Wyk S; Wilkinson E; Wilson S; de Oliveira T; van Zyl G |
| EPI_ISL_13243355 | Centrum Medyczne MEDYK Zaklad Diagnostyki Medycznej | Wojewodzka Stacja Sanitarno-Epidemiologiczna w Rzeszowie, Laboratorium Diagnostyki Medycznej | Anna Nowakowska; Karolina Ostrowska; Katarzyna Wilk; Marzena Baranowska |
| EPI_ISL_13337902 | Clinica biblica | Inciensa, Instituto Costarricense de Investigación y Enseñanza en Nutrición y Salud | Adriana Godínez; Claudio Soto-Garita; Estela Cordero; Francisco Duarte; Gabriel Morales & Natalia Bonilla; Hebleen Porras; José Luis Vargas; Mariela Gutiérrez; Melany Calderón; Sofia Herrera |
| EPI_ISL_13323849, EPI_ISL_13323984 | Clinical Microbiology Laboratory, Tel Aviv Sourasky Medical Center | Clinical Microbiology Laboratory, Tel Aviv Sourasky Medical Center | Alon Ziv; Amos Adler; Goel Morad; Katya Levytskyi; Lior Handler; Matan Slutzkin; Ora Halutz; Orly Eshel |
| EPI_ISL_13129404, EPI_ISL_13242132 | Clinical Microbiology, Infection Prevention and Control | Section for Molecular Diagnostics | Björn Hallström; Jonas Björkman |
| EPI_ISL_12954164 | Coronavirus Homecare | National Institute for Communicable Diseases of the National Health Laboratory Service | Amoako DG; Bhiman JN; Everatt J; Ismail A; Kekana D; Mahlangu B; Maphalala G; Mnguni A; Mohale T; Ntuli N; Scheepers C; Wolter N |

| | | | |
|---|---|---|---|
| EPI_ISL_12918515 | D'Almeida Clinic wc DAL | NHLS/UCT | Arash Iranzadeh; Carolyn Williamson; Diana Hardie; Gert Marais; Innocent Mudau; Luicer Olubayo; Marvin Hsiao; Nokuzola Mbhele; Rageema Joseph; Stephen Korsman |
| EPI_ISL_12291987, EPI_ISL_12728612, EPI_ISL_12895296, EPI_ISL_13049308, EPI_ISL_13049625, EPI_ISL_13157845, EPI_ISL_13177782, EPI_ISL_13178663, EPI_ISL_13201568, EPI_ISL_13241448, EPI_ISL_13299517 | | | |
| see above | Department of Bacteria, Parasites and Fungi, Statens Serum Institut, Copenhagen, Denmark | Statens Serum Institut Bioinformatics and Microbial Genomics | Danish Covid-19 Genome Consortium |
| EPI_ISL_13140517 | Department of Health Technology and Informatics, The Hong Kong Polytechnic University | Department of Health Technology and Informatics, The Hong Kong Polytechnic University | Alan Ka-Lun Wu; Alex Yat-Man Ho; Barry Kin-Chung Wong; Chloe Toi-Mei Chan; David Ho-Keung Shum; Gilman Kit-Hang Siu; Hiu-Yin Lao; Ivan Tak-Fai Wong; Jake Siu-Lun Leung; Kam-Tong Yip; Kenneth Siu-Sing Leung; Kingsley King-Gee Tam; Kitty Sau-Chun Fung; Kristine Luk; Lam-Kwong Lee; Miranda Chong-Yee Yau; Sandy Ka-Yee Chau; Shea Ping Yip; Tak-Lun Que; Timothy Ting-Leung Ng; Wing Cheong Yam; Wing-Hei Lo; Wing-Kin To; Yvette Wai-Man Lai |
| EPI_ISL_13182857 | Department of Medical Microbiology, Baerum Hospital, Vestre Viken Health Trust | Norwegian Institute of Public Health, Department of Virology | Atiya R Ali; Debech Nadia; Engebretsen Serina Beate; Garcia Llorente Ignacio; Hilde Elshaug; Hilde Nordby Falkenhaug; Hilde Vollan; Jon Bråte; Kamilla Heddeland Instefjord; Karoline Bragstad; Kathrine Stene-Johansen; Line Victoria Moen; Marie Paulsen Madsen; Olav Hungnes; Pedersen Benedikte Nevjen; Rasmus Riis Kopperud |
| EPI_ISL_13328006 | Department of Medical Microbiology, St. Olavs hospital | Norwegian Institute of Public Health, Department of Virology | Atiya R Ali; Debech Nadia; Engebretsen Serina Beate; Garcia Llorente Ignacio; Hilde Elshaug; Hilde Nordby Falkenhaug; Hilde Vollan; Jon Bråte; Kamilla Heddeland Instefjord; Karoline Bragstad; Kathrine Stene-Johansen; Line Victoria Moen; Marie Paulsen Madsen; Olav Hungnes; Pedersen Benedikte Nevjen; Rasmus Riis Kopperud |
| EPI_ISL_13351801 | Department of Microbiology and Infection Control, Akershus University Hospital HF | Department of Microbiology and Infection Control, Akershus University Hospital HF | Alexander Hesselberg Løvestad; Divya Murugananthan; Hanne Berggreen; Hege Vangstein Aamot |
| EPI_ISL_13109539, EPI_ISL_13109540, EPI_ISL_13302865 | Department of Virology, National Institute of Health, Islamabad, Pakistan | Department of Virology, National Institute of Health, Islamabad, Pakistan | Aamer Ikram; Massab Umair; Muhammad Ammar; Muhammad Salman; Nazish Badar; Qasim Malik; Syed Adnan Haider; Zaira Rehman |
| EPI_ISL_12972952 | Dianalabs SA | Genesupport | Geraldine Jost; Katia Jaton; Nadia Liassine; Tanguy ARAUD |
| EPI_ISL_13102657, EPI_ISL_13102658, EPI_ISL_13102683, EPI_ISL_13254037, EPI_ISL_13254040, EPI_ISL_13254058, EPI_ISL_13342160 | | | |
| see above | Directorate of Public Health and Preventive Medicine | CDFD-INSACOG | Arunkumar Karunanidhi; Ashwin Dalal; Asmita Gupta; Avudaiselvi Rathinasamy; Darez Ahamed; Devi Monika Ayyagari Venkata; Divya Vashisht; Gurunathan Subramanian; Hemashree Kannan; Kalpana Raghu; Murali Bashyam; Rajesh Kumar Manivannan; Raju Sivadoss; Rupin Shelake; Sampath Palani; Selvavinayagam Sivaprakasam; Vinay Donipadi |
| EPI_ISL_13025341, EPI_ISL_13086516, EPI_ISL_13353282, EPI_ISL_13353404, EPI_ISL_13353627 | Division of Emerging Infectious Diseases, Bureau of Infectious Diseases Diagnosis Control, Korea Disease Control and Prevention Agency | Division of Emerging Infectious Diseases, Bureau of Infectious Diseases Diagnosis Control, Korea Disease Control and Prevention Agency | Ae Kyung Park; Chae Young Lee; Eun-Jin Kim; Hyuck Jin Lee; Il-Hwan Kim; Jeong-Ah Kim |
| EPI_ISL_12703375, EPI_ISL_12903896, EPI_ISL_13209520 | Dr. Mustafa, Dr. Richter Labor für medizinisch-chemische und mikrobiologische Diagnostik GmbH, Abteilung Molekularbiologie | Dr. Mustafa, Dr. Richter Labor für medizinisch-chemische und mikrobiologische Diagnostik GmbH, Abteilung Molekularbiologie | Alexander Gamisch; Maria Elisabeth Mustafa |
| EPI_ISL_13051401, EPI_ISL_13051430, EPI_ISL_13243915, EPI_ISL_13243973, EPI_ISL_13243977, EPI_ISL_13243990, EPI_ISL_13363222 | | | |
| see above | Dr. Risch Ostschweiz AG | Dr Risch Laboratory | Dominique Fabien Hilti; Faina Wehrli; Lorenz Risch; Martin Risch; Nadia Wohlwend; Sinem Kas; Thomas Bodmer |
| EPI_ISL_12783845, EPI_ISL_13144618, EPI_ISL_13214669, EPI_ISL_13332292, EPI_ISL_13332293, EPI_ISL_13332294, EPI_ISL_13332295, EPI_ISL_13332296, EPI_ISL_13332297, EPI_ISL_13332299, EPI_ISL_13332300, EPI_ISL_13332355, EPI_ISL_13332375 | | | |
| see above | Dutch COVID-19 response team | National Institute for Public Health and the Environment (RIVM) | Adam Meijer; Afke Vogelzang; AnneMarie van den Brandt; Annelies Kroneman; Bas van der Veer; Chantal Reusken; Dennis Schmitz; Dirk Eggink; Florian Zwagemaker; Harry Vennema; Ivo van Walle; Jeroen Cremer; Jil Kocken; Jordy de Bakker; Karim Hajji; Kim Freriks; Linda van Someren; Lisa Wijsman; Lynn Aarts; Ryanne Jaarsma; Sanne Bos; Sharon van den Brink; on behalf of the national COVID-19 response team |
| EPI_ISL_13160184 | EORLA | Kingston Health Sciences Centre | Calvin Sjaarda; Drew Roberts; Henry Wong; Jacob Whalen; Nick Buchner; Phung Ta; Prameet Sheth; Sheri Levesque |
| EPI_ISL_13353762 | Elizabeth Glaser Pediatric AIDS Foundation | KEMRI-Wellcome Trust Research Programme,Kilifi | Agoti C.; D.J.Nokes; Githinji G.; Lambisia A.; Makori T.; Mburu M.W.; Mohamed K.S.; Morobe J.; Ndwiga L.; Ochola I; Ongera E.; de Laurent Z. |
| EPI_ISL_13223992 | Enfer | Enfer | Elaine M. Kenny; Suzie Coughlan |
| EPI_ISL_13294882 | Eurofins-NMDL | Eurofins-NMDL | Anco Molijn; Anne Vogel; Lisa Dreesens; Marvin Ruiter; Maurine Leversteijn-van Hall; Roy Masius; Simon Lansu |
| EPI_ISL_13228364 | Faculty Hospital Bulovka, Department of Clinical Microbiology | Charles University, Faculty of Science, BIOCEV, OMICS Genomics | Alžběta Bučková; Blanka Hamplová; Ingrid Poláková; Jiří Novák; Nela Václavíková; Ruth Tachezy; Sebastian Cristian Treitli; Vladimír Hampl; Zoltán Füssy; Štěpánka Hrdá |
| EPI_ISL_13163536 | Fondation Congolaise pour la recherche medicale (FCRM), Francine Ntoumi | Fondation Congolaise pour la Recherche Médicale | Dr Batchi-Bouyou Armel Landry; Dr. Jean Claude Djontu; Mfoutou Mapanguy Claujens Chastel; Prof. Francine Ntoumi |
| EPI_ISL_13311036, EPI_ISL_13346730, EPI_ISL_13346731, EPI_ISL_13346748 | Gandhi Medical College and Hospital (GMCH), Secunderabad | NIV Influenza | A.Rajender goud; Abdul Majeed; Amrithesh Kumar Arun; D.R.Manisha Rani; Devendhar; Dr.G.Sushma Rajya Lakshmi; Dr.K.Nagamani; Dr.Sunitha Pakalapaty; Hajeera Osmani; Sahithya |
| EPI_ISL_12982341, EPI_ISL_12982342, EPI_ISL_12982343, EPI_ISL_13154708, EPI_ISL_13154710, EPI_ISL_13154971, EPI_ISL_13155270, EPI_ISL_13155302, EPI_ISL_13155321, EPI_ISL_13164925, EPI_ISL_13164932, EPI_ISL_13164941, EPI_ISL_13164983, EPI_ISL_13164991, EPI_ISL_13165708, EPI_ISL_13165719 | | | |
| see above | Genetica Molecular and Subdepartamento de Virologia ISP Chile | Instituto de Salud Publica de Chile | Andres Castillo; Barbara Parra; Constanza Campano; Ivan Ponce; Jorge Fernandez; Karen Orostica; Marcelo Rojas; Matias Pezoa; Patricia Bustos; Paulo covarrubias; Rodrigo Fasce |
| EPI_ISL_13229452, EPI_ISL_13368867 | H Fernando Fonseca | Instituto Nacional de Saude Doutor Ricardo Jorge (INSA) | Borges et al |
| EPI_ISL_13142445, EPI_ISL_13142450, EPI_ISL_13206776 | HB TERREO EMERG CONV QUARTOS | Instituto Butantan | ; Alex Ranieri Lima; Antonio Jorge Martins; Claudia Renata dos Santos Barros; David Schlesinger; Debora Botequio Moretti; Dimas Tadeu Covas; Elaine Cristina Marqueze; Elaine Vieira Santos; Evandra Strazza Rodrigues; Gabriela Ribeiro; Heidge Fukumasu; Jayme Augusto de Souza-Neto; Luiz Alcantara; Luiz Lehmann Coutinho; Maria Carolina Elias; Maurício Lacerda Nogueira; Raul Machado Neto; Rejane Maria Tommasini Grotto; Ricardo Haddad; Sandra Coccuzzo Sampaio Vessoni; Simone Kashima; Svetoslav Nanev Slavov; Vincent Louis Viala |
| EPI_ISL_13150091 | HOPITAL DE MONTLUCON | CHU Clermont-Ferrand, service de virologie | Bisseux Maxime; Combes Patricia; Henquell Cecile; Mirand Audrey |
| EPI_ISL_13069827 | HOSPITAL DR. WILLIAM ALLEN | Inciensa, Instituto Costarricense de Investigación y Enseñanza en Nutrición y Salud | Adriana Godínez; Claudio Soto-Garita; Estela Cordero; Francisco Duarte; Gabriel Morales & Natalia Bonilla; Hebleen Porras; José Luis Vargas; Mariela Gutiérrez; Melany Calderón; Sofia Herrera |
| EPI_ISL_13114125 | HOSPITAL UNIVERSITARIO CENTRAL DE ASTURIAS | Laboratorio de Virología HUCA | ; Alba L; Alvarez-Arguelles ME; Boga JA; Costales I; Coto E; González-Alba JM; Gómez de Oña J; Martín-Rodríguez G; Melón S; Perez-Martínez Z; Rojo S; Sandoval M |
| EPI_ISL_12933029, EPI_ISL_13198799, EPI_ISL_13337436 | Histopath | NSW Health Pathology - Institute of Clinical Pathology and Medical Research; Westmead Hospital; University of Sydney | Arnott A.; Draper J.; Gall M.; Martinez E.; Rockett R.; Sintchenko V.; on behalf of ICPMR |
| EPI_ISL_13369822, EPI_ISL_13369826, EPI_ISL_13369827, EPI_ISL_13369828, EPI_ISL_13369861 | Home Quarantine Taskforce | Hong Kong Department of Health | Alan K.L. Tsang; Edman T.K. Lam; Ken H.L. Ng; Patricia K. L. Leung; Rickjason C.W. Chan |
| EPI_ISL_13283230, EPI_ISL_13283380, EPI_ISL_13283861, EPI_ISL_13372852 | Hopital | National Reference Center for Viruses of Respiratory Infections, Institut Pasteur, Paris | Angela Brisebarre; Camille Capel; Christophe Malabat; Corinne Maufrais; Etienne Simon-Lorière; Frédéric Lemoine; Julien Fumey; L COURDAVAULT; Laura DJAMDJIAN; Laurence FAGOUR; Louise Lefrançois; Marion Barbet; Maud Vanpeene; Méline Bizard; Slim El Khiari; Sylvie Van der Werf; Vincent Enouf |
| EPI_ISL_13002450 | Hospital Universitari Dr. Josep Trueta | Institut d'Investigació Biomèdica de Girona \| Hospital Universitari Dr. Josep Trueta | Bernat del Olmo; Mel·lina Pinsach; Meritxell Deulofeu; Nuria Esther Neto; Paula Costa |
| EPI_ISL_13340685 | Institute for Water Quality and Resource Management, Technical University Vienna | Bergthaler laboratory, CeMM Research Center for Molecular Medicine of the Austrian Academy of Sciences | Alberto Alises; Andreas Bergthaler; Anna Schedl; Christoph Bock; Fabian Amman; Lukas Endler; Matthew Thornton; Michael Schuster; Michelle Chan; Petr Triska |
| EPI_ISL_13368512, EPI_ISL_13368516, EPI_ISL_13368536 | Institute of Microbiology and Immunology, Faculty of Medicine, University of Ljubljana | Institute of Microbiology and Immunology, Faculty of Medicine, University of Ljubljana | Alen Suljič; Andraž Celar; Doroteja Vlaj; Mario Poljak; Miša Korva; Patricija Pozvek; Samo Zakotnik; Tatjana Avšič – Županc; Tina Gabrovšek; Tina Živič; Tomaž Mark Zorec; Špela Pleh |
| EPI_ISL_13118911 | Islab, Pohjois-Karjalan aluelaboratorio | Expert Microbiology, National Institute for Health and Welfare | Carita Savolainen-Kopra; Erika Lindh; Haider al-Hello; Jani Halkilahti; Kirsi Liitsola; Niina Ikonen; Niko Tervo; Olli Vapalahti; Pekka Ellonen; Phuoc Truong; Päivi Laurila; Ravi Kant; Sari Hannula; Soile Blomqvist; Teemu Smura |
| EPI_ISL_13203753, EPI_ISL_13203755, EPI_ISL_13203845, EPI_ISL_13203852, EPI_ISL_13203877, EPI_ISL_13203887 | Islab, Pohjois-Savon aluelaboratorio | Expert Microbiology, National Institute for Health and Welfare | Carita Savolainen-Kopra; Erika Lindh; Haider al-Hello; Jani Halkilahti; Kirsi Liitsola; Niina Ikonen; Niko Tervo; Olli Vapalahti; Pekka Ellonen; Phuoc Truong; Päivi Laurila; Ravi Kant; Sari Hannula; Soile Blomqvist; Teemu Smura |

| | | | |
|---|---|---|---|
| EPI_ISL_13353748 | KEMRI-Wellcome Trust Research Programme,Kilifi | KEMRI-Wellcome Trust Research Programme,Kilifi | Agoti C.; D.J.Nokes; Githinji G.; Lambisia A.; Makori T.; Mburu M.W.; Mohamed K.S.; Morobe J.; Ndwiga L.; Ochola I; Ongera E.; de Laurent Z. |
| EPI_ISL_13334887 | Kaiser Permanente Southern California | Helix | Helix; Kaiser Permanente Southern California |
| EPI_ISL_13371724, EPI_ISL_13371772, EPI_ISL_13371783 | Karolinska University Hospital Huddinge | Karolinska University Hospital | Annika Tiveljung Lindell; Henning Onsbring; Jan Albert; Karina Hentrich; Lynda Eneh; Maria Ropat; Martin Ekman; Natalija Gerasimcik; Robert Dyrdak; Sandra Broddesson; Shambhu Ganeshappa Aralaguppe; Tanja Normark; Tobias Allander; Valtteri Wirta; Zhibing Yun |
| EPI_ISL_12864060, EPI_ISL_13131104, EPI_ISL_13313980, EPI_ISL_13314006 | Karolinska University Hospital Solna | Karolinska University Hospital | Annika Tiveljung Lindell; Henning Onsbring; Jan Albert; Karina Hentrich; Lynda Eneh; Maria Ropat; Martin Ekman; Natalija Gerasimcik; Robert Dyrdak; Sandra Broddesson; Shambhu Ganeshappa Aralaguppe; Tanja Normark; Tobias Allander; Valtteri Wirta; Zhibing Yun |
| EPI_ISL_13064563 | Kuala Lumpur International Airport | Institute for Medical Research, Infectious Disease Research Centre, National Institutes of Health, Ministry of Health Malaysia | Anasir MI; G.Adypatti NM; Jamaluddin MS; Kalyanasundram J; Kamel K; MatRahim N; Nawi MH; Suib FA; Suppiah J; Thayan R |
| EPI_ISL_13342510 | LABORATOIRE CREAVALLEE | CNR Virus des Infections Respiratoires - France SUD | Antonin Bal; Bruno Lina; Bruno Simon; Gregory Destras; Gwendolyne Burfin; Hadrien Regue; Laurence Josset; Martine Valette; Quentin Semanas; Theophile Boyer |
| EPI_ISL_13176333 | LABORATOIRE d'ANALYSES de BIOLOGIE MEDICALES | CNR Virus des Infections Respiratoires - France SUD | Antonin Bal; Bruno Lina; Bruno Simon; Gregory Destras; Gwendolyne Burfin; Hadrien Regue; Laurence Josset; Martine Valette; Quentin Semanas; Theophile Boyer |
| EPI_ISL_12767816 | LATE - Laboratório de Técnicas Especiais - Hospital Israelita Albert Einstein | LATE - Laboratório de Técnicas Especiais - Hospital Israelita Albert Einstein | Alexandre Hideaki Takara; Ana Paula Moreira Salles; Anelisie da Silva Santos; Deyvid Amgarten; Erick Gustavo Dorlass; Fernanda de Mello Malta; João Renato Rebello Pinho; Luiz Vicente Rizzo; Marcio Anunciacao Menezes; Pedro Henrique Sebe Rodrigues; Raquel Riyuzo |
| EPI_ISL_13183074 | Lab voor klinische biologie | Lab voor klinische biologie | Bruno Verhasselt; Hannelore Hamerlinck; May-Linh Truong |
| EPI_ISL_13363991, EPI_ISL_13364015 | Labkesda DKI | National Quality Control Laboratory of Drug and Food | Hana Apsari Pawestri; M. Erdiansyah; Nurul Azizah; Sri Utaminingsih |
| EPI_ISL_13351624 | Labkesda Kabupaten Batang | National Institute of Health Research and Development | Arie Ardiansyah Nugraha; Fajar Nur Sulistiyohadi; Hana Apsari Pawestri; Hartanti Dian Ikawati; IGM Wirabrata; Kartika Dewi Puspa; Nelis Imaningsih; Nur Ika Hariastuti; Putri Widia Astuti; Subangkit |
| EPI_ISL_13091520, EPI_ISL_13283371, EPI_ISL_13362924, EPI_ISL_13372706, EPI_ISL_13372838 | Labo Analyses Med | National Reference Center for Viruses of Respiratory Infections, Institut Pasteur, Paris | Angela Brisebarre; Camille Capel; Christophe Malabat; Corinne Maufrais; Etienne Simon-Lorière; Frédéric Lemoine; J.M CORCY; Julien Fumey; Karine MICHEZ; Louise Lefrançois; Marie-Hélène GLAUDON LOUVEAU DE LA GUIGNERAYE; Marion Barbet; Maud Vanpeene; Méline Bizard; Slim El Khiari; Sylvie Van der Werf; Vincent Enouf |
| EPI_ISL_13241273 | Labor Lübeck bzw. Laborärztliche Gemeinschaftspraxis Lübeck | Robert Koch Institute | |
| EPI_ISL_13343334 | Labor team w AG | Department of Biosystems Science and Engineering, ETH Zürich | Andreas Grutsch; Andreas Lindauer; Chaoran Chen; Christian Beisel; David Dreifuss; Elodie Burcklen; Franziska Singer; Ina Nissen; Ivan Topolsky; Kim Philipp Jablonski; Lara Fuhrmann; Louis du Plessis; Matteo Carrara; Mirjam Feldkamp; Monika Bucher; Natascha Santacroce; Niko Beerenwinkel; Pelin Burcak Icer; Rebecca Denes; Rebekka Pohl; Sarah Nadeau; Shuqing Yu; Tanja Stadler; Tobias Schär |
| EPI_ISL_13140449, EPI_ISL_13140452 | Laboratoire Rodolphe Mérieux (INRB Goma) | Pathogen Genomics Lab, National Institute for Biomedical Research (INRB) | Allison Black; Amuri Aziza; Andrew Rambaut; Catherine Pratt; Daniel Mukadi; Eddy Kinganda-Lusamaki; Edith Nkwembe; Emmanuel Lokilo Lofiko; Francisca Muyembe Mawete; Gradi Luakanda; Hervé Viala; Ian Goodfellow; James Hadfield; Jean Claude Makangara; Jean-Jacques Muyembe Tamfum; Josh Quick; Kristian Andersen; Matthias Pauthner; Michael Wiley; Michel Mbimbi; Nick Loman; Placide Mbala-Kingebeni; Steve Ahuka-Mundeke; Trevor Bedford |
| EPI_ISL_13303541, EPI_ISL_13303746, EPI_ISL_13304033 | Laboratoire de santé publique du Québec | Laboratoire de santé publique du Québec | Guillaume Bourque; Ioannis Ragoussis; Jesse Shapiro; Mark Lathrop and Judith Fafard on behalf of the CoVSeQ research group; Sandrine Moreira |
| EPI_ISL_13282483 | Laboratoire national de sante, Microbiology, Virology | Laboratoire national de sante, Microbiology, Microbial Genomics Platform | Anke Wienecke-Baldacchino; Catherine Ragimbeau; Elodie Solarino; Eric Hugoson; Fatu Djabi; Jessica Tapp; Lise Pignon; Raoul Salmon; Sibel Berger; Tamir Abdelrahman; Trung Nguyen Nguyen; Virginie Jover |
| EPI_ISL_13292140 | Laboratoires Reunis | Laboratoire national de sante, Microbiology, Microbial Genomics Platform | Anke Wienecke-Baldacchino; Bernard Weber; Catherine Ragimbeau; Elodie Solarino; Eric Hugoson; Fatu Djabi; Jessica Tapp; Lise Pignon; Raoul Salmon; Sibel Berger; Tamir Abdelrahman; Virginie Jover |
| EPI_ISL_13292035, EPI_ISL_13292343, EPI_ISL_13292366 | Laboratoires d'analyses medicales - KETTERTHILL | Laboratoire national de sante, Microbiology, Microbial Genomics Platform | Anke Wienecke-Baldacchino; Caroline Scheiber; Catherine Ragimbeau; Elodie Solarino; Eric Hugoson; Fatu Djabi; Jessica Tapp; Lise Pignon; Raoul Salmon; Serge Vedy; Sibel Berger; Tamir Abdelrahman; Virginie Jover |
| EPI_ISL_12838778 | Laboratorio Central de Saude Publica do Estado do Rio de Janeiro (LACEN/RJ) | Laboratory of Respiratory Viruses and Measles, Oswaldo Cruz Institute, FIOCRUZ | Alice Sampaio Rocha; Andrea Cony Cavalcanti; Bruna Mendonça da Silva; Elisa Cavalcante Pereira; Fernando Motta; Ighor Arantes; Jéssica Graça Macedo de Carvalho; Larissa Macedo Pinto; Luciana Appolinario; Marilda Siqueira on behalf of the Fiocruz COVID-19 Genomic Surveillance Network; Paola Resende; Victor Guimaraes |
| EPI_ISL_13142047, EPI_ISL_13314030, EPI_ISL_13314041, EPI_ISL_13314213 | Laboratorio de Referencia Nacional de Virus Respiratorios. Centro Nacional de Salud Publica. Instituto Nacional de Salud Peru. | Laboratorio de Referencia Nacional de Virus Respiratorios. Centro Nacional de Salud Publica. Instituto Nacional de Salud Peru. | Alicia Nuñez Llanos; Carlos Padilla Rojas; Estela Huaman Angeles; Francisco Ascue Orosco; Gloria Arotinco Garayar.; Henri Bailon Calderon; Iris Silva Molina; Jorge Giraldo Chavez; Joseph Huayra Niquen; Karla Vasquez Cajachahua; Kelly Izarra Rojas; Lely Solari Zerpa; Lilian Huarca Balbin; Lisbet Roxana Inga Angulo; Luis Barcena Flores; Luren Sevilla Castañeda; Marco Galarza Perez; Maria Villar Saavedra; Nancy Rojas Serrano; Omar Caceres Rey; Orson Mestanza Millones; Princesa Medrano Alhuay; Priscila Lope Pari; Steve Acedo Lazo; Veronica Hurtado Vela; Victor Jimenez Vasquez; Wendy Lizarraga Olivares |
| EPI_ISL_12889805, EPI_ISL_13152605 | Laboratory Corporation of America | Centers for Disease Control and Prevention Division of Viral Diseases, Pathogen Discovery | Amanda Douglas; Amanda Suchanek; Andrea Throop; Ayla Burns; Benjamin Rambo-Martin; Bobbi Croy; Brian Krueger; Brian Norvell; Christopher Gulvick; Christos Petropoulos; Clinton Paden; Craig Lukasik; Dakota Howard; Debbie Boles; Dhwani Batra; Duncan MacCannell; Eyad Almasri; Goran Stevovic; Howard Engler; Hrushikesh Deshmukh; Jake Humphrey; Jana Schroth; Jason Caravas; Joe Voshell; John Pruitt; Jonathan Meltzer; Jonathan Williams; Kimberly Wagner; Kristine Lacek; Lax Iyer; Lisa Pfefferle; Lyndon Tilson; Manoj Jain; Marcia Eisenberg; Mary Cristobal; Mary Williamson; Matthew Robinson; Matthew Schmerer; Michael Levandoski; Mike Sapeta; Mindy Nye; Minoo Agarwal; Mohan Kolli; Nuthawin Charoensri; Oren Cohen; Peter Cook; Prashant Gupta; Qian Zeng; Rama Ghatti; Scott Parker; Scott Ryan; Scott Sammons; Shatavia Morrison; Stanley Letovsky; Steven Ragan; Suresh Selvaraju; Susan Countryman; Susan Hicks; Suzanne Dale; Thomas Urban; Tim Kuphal; Tricia Zwiefelhofer; Tymeckia Kendall; Victoria Caban Figueroa; Vincent Drouillon; Yvette Unoarumhi |
| EPI_ISL_13013557, EPI_ISL_13229149, EPI_ISL_13229156, EPI_ISL_13229163, EPI_ISL_13229172 | Laboratory of Genomics and Bioinformatics, Comenius University Science Park | Laboratory of Genomics and Bioinformatics, Comenius University Science Park | Anna Kaliňáková; Barbora Kotvasová; Diana Rusňáková; Jakub Styk; Jaroslav Budiš; Lucia Ševčíková; Michaela Jakubková Forgáčová; Miroslav Böhmer; Nikola Lipková; Pavol Mišenko; Silvia Bokorová; Tatiana Sedláčková; Terézia Vrabľová; Tomáš Szemes |
| EPI_ISL_12561210, EPI_ISL_12561211, EPI_ISL_12561212, EPI_ISL_12561213 | Landspítali, Department of Clinical Microbiology | Landspítali, Department of Clinical Microbiology | Arsalan Amirfallah; Freyja Valsdóttir; Zarko Urosevic |
| EPI_ISL_12875191, EPI_ISL_12875272, EPI_ISL_13249983, EPI_ISL_13250051 | Lifebrain Covid Labor GmbH | Lifebrain Covid Labor GmbH | Alexandra Wagner; Anna Edermayr; Filip Sima; Kristina Bavrka Kolenc; Lucia Castello |
| EPI_ISL_13157142, EPI_ISL_13185833, EPI_ISL_13231687, EPI_ISL_13311807 | Lighthouse Lab in Glasgow | Wellcome Sanger Institute for the COVID-19 Genomics UK (COG-UK) Consortium | Anna Dominiczak and Alex Alderton; Carol Clugston; Cordelia Langford; David Gray; David K. Jackson; Dominic Kwiatkowski; Ewan Harrison; Harper VanSteenhouse; Ian Johnston; Jeffrey Barrett; John Sillitoe on behalf of the Wellcome Sanger Institute COVID-19 Surveillance Team; Roberto Amato; Sonia Goncalves; Yumi Kasai |
| EPI_ISL_13033928 | Limbach - MVZ Labor Ravensburg Labor Dr. Gärtner | Robert Koch Institute | |
| EPI_ISL_13137139, EPI_ISL_13355257 | MANILA DOCTORS HOSPITAL | Research Institute for Tropical Medicine | Alexander Sadiasa; Amalea Dulcene D. Nicolasora; Angela Kae T. Chang; Anne Pauline A. Alpino; Ardiane Ysabelle M. Dolor; Catalino S. Demetria; Charalyn Babida; Claudette Lee S. Navarro; Criselda T. Bautista; Dodge R. Lim; Francisco Gerardo M. Polotan; Gerald Ivan S. Sotelo; Jefferson Earl J. Halog; Joseph Hughes; Joy Mariette L. Parayray; Katie Hampson; Kirstyn Brunker; Lei Lanna M. Dancel; Ma. Angelica A. Tujan; Ma. Ricci R. Gomez; Mayan Uy-Lumandas; Othoniel Jan T. Onza; Simon Daldry; Timothy John R. Dizon; Vina Lea F. Arguelles; Yao-Tsun Li |
| EPI_ISL_13247839 | MDI Limbach Berlin GmbH; MVZ Labor Berlin | Robert Koch Institute | |
| EPI_ISL_13332769, EPI_ISL_13332770, EPI_ISL_13332771, EPI_ISL_13332773, EPI_ISL_13332774, EPI_ISL_13332775 | MRC/UVRI & LSHTM Uganda Research Unit | MRC/UVRI & LSHTM Uganda Research Unit | Bernard Mpairwe; Joseph Mugisha; Matthew Cotten; My V.T. Phan; Robert Newton |
| EPI_ISL_13269831 | MVZ Dr. Eberhard & Partner Dortmund | Robert Koch Institute | |
| EPI_ISL_13268448 | MVZ Labor Krone GbR | Robert Koch Institute | |
| EPI_ISL_13359967 | Mater Pathology, South Brisbane—Mater Hospital Brisbane | Public Health Virology - Forensic and Scientific Services (PHV-FSS) | Chenwei Wang on behalf of Q-PHIRE Genomics |
| EPI_ISL_12954406 | Mbabane Public Health Unit | National Institute for Communicable Diseases of | Amoako DG; Bhiman JN; Everatt J; Ismail A; Kekana D; Mahlangu B; Maphalala G; Mnguni A; Mohale T; Ntuli N; Scheepers C; Wolter N |

| | | | |
|---|---|---|---|
| | | the National Health Laboratory Service | |
| EPI_ISL_12954176 | Mbabane city council | National Institute for Communicable Diseases of the National Health Laboratory Service | Amoako DG; Bhiman JN; Everatt J; Ismail A; Kekana D; Mahlangu B; Maphalala G; Mnguni A; Mohale T; Ntuli N; Scheepers C; Wolter N |
| EPI_ISL_13238615 | Medizinische Laboratorien Düsseldorf | Robert Koch Institute | |
| EPI_ISL_12915494 | Microbiology Department, Laboratori Clínic Metropolitana Nord. Hospital Universitari Germans Trias i Pujol | Can Ruti SARS-CoV-2 Sequencing Hub (HUGTiP/IrsiCaixa/IGTP) | Alexia París; Ana Blanco; Andreu Coello; Antoni E Bordoy; Bonaventura Clotet; David Panisello; Francesc Catala-Moll; Gemma Clara; Ignacio Blanco; Laia Soler; Marc Noguera-Julian; Montserrat Giménez; Pere-Joan Cardona; Pilar Armengol; Roger Paredes; Sara González; Verónica Saludes; and Elisa Martró on behalf of the Can Ruti SARS-CoV-2 Sequencing Hub |
| EPI_ISL_13229987, EPI_ISL_13230291, EPI_ISL_13312308, EPI_ISL_13312310 | Microbiology Department. Complexo Hospitalario Universitario de Vigo | Microbiology Department. Complexo Hospitalario Universitario de Vigo | Cabrera JJ; Cortizo S; Daviña C; Gonzalez-Dominguez M; Martinez L; Pena I; Perez-Castro S; Potel C; Rey S; Vasallo FJ; del-Campo V |
| EPI_ISL_13113071, EPI_ISL_13113222, EPI_ISL_13176029 | Ministry of Health Turkey | Ministry of Health Turkey | Arzu İrvem; Cemal Kazezoğlu; Feride Velaei; Gülay Korukluoğlu; Gültekin Ünal; Meral Kaya; Rabia Can Sarınoğlu; Serap Demir Tekol; Şemsinur Karabela |
| EPI_ISL_12972929, EPI_ISL_12972930, EPI_ISL_12972931, EPI_ISL_12972932, EPI_ISL_12972933 | Molecular Biology and Virology | Molecular Biology and Virology | Dr.Moh''D Borhan Al-Zghoul; Dr.Mustafa Ababneh; Mohammad Alboom |
| EPI_ISL_13244309 | Molecular Genetic Monitoring Group | Molecular Genetic Monitoring Group | Anna S. Gladkikh; Areg A.Totolian; Ekaterina O. Klyuchnikova; Valerya A. Sbarzaglia; Vladimir G. Dedkov |
| EPI_ISL_13259127, EPI_ISL_13259128 | NIH | National Institute of Hygiene | Abderrahman Bimouhen; Fatima El Falaki; Hassan Ihazmade; Hicham Oumzil; Zakia Regragui |
| EPI_ISL_12837921 | Nastavni zavod za javno zdravstvo Splitsko- Dalmatinske županije | Hrvatski zavod za javno zdravstvo | Anita Jurić; Dragan Jurić; Irena Tabain; Ivana Ferenčak; Josipa Kuzle |
| EPI_ISL_12845590 | National Health Laboratory Services | CERI, Centre for Epidemic Response and Innovation, Stellenbosch University and KRISP, KZN Research Innovation and Sequencing Platform, UKZN. | Anyaneji UJ; Giandhari J; Maharaj A; Moir M; Naidoo Y; Nokukhanya Mdlalose; Pillay S; San JE; Sanko TJ; Tegally H; Tshiabuila D; Van Wyk S; Wilkinson E; de Oliveira T |
| EPI_ISL_11763529, EPI_ISL_12097409 | National Health Laboratory Services | CERI, Centre for Epidemic Response and Innvoation, Stellenbosch University and KRISP, KZN Research Innovation and Sequencing Platform, UKZN. | Anyaneji UJ; Giandhari J; Maharaj A; Mdlalose N; Moir M; Naicker D; Naidoo Y; Nokukhanya Mdlalose; Pillay S; San JE; Tegally H; Tshiabuila D; Van Wyk S; Wilkinson E; de Oliveira T |
| EPI_ISL_12474479 | National Health Laboratory Services, Virology | National Health Laboratory Services, Virology | Ashlyn S. C. Davis; Florette K. Treurnicht; Kathleen Subramoney; Nkhensani Mtileni |
| EPI_ISL_13345427 | National Laboratory for Health, Environment and Food, OMM, Maribor | NLZOH (National Laboratory for Health, Environment and Food) | Aleksander Mahnic; Alenka Štorman; Andrej Golle; Kaja Tominc; Leon Marič; Maja Rupnik; Maša Jarčič; Mojca Cimerman; Nika Gobec; Nika Volmajer; Sabina Mlakar; Sandra Janezic; Tanja Vrabič; Tjaša Žohar Čretnik; Urška Dobovišek |
| EPI_ISL_13102212 | National Platform bis COVID ULB-IBC | National Platform bis COVID ULB-IBC | Arnaud Marchant; Coralie Henin; Lionel Schiavolin; Marie-Luce Delforge; Mathilde Le Garrec |
| EPI_ISL_12835609 | National Platform bis UMONS/Jolimont | National Platform bis UMONS/Jolimont | Caroline Debecker; Clothilde Claus; Eric Tarantino; Florian Juszczak; Gautier Detry; Laetitia Gheysen; Ruddy Wattiez |
| EPI_ISL_13094168, EPI_ISL_13150735, EPI_ISL_13150742, EPI_ISL_13259913 | National Public Health Laboratory, National Centre for Infectious Diseases | National Public Health Laboratory, National Centre for Infectious Diseases | BeiBei Chen; Benny Yeo; Chen Shi Ling; Grace Ngan; Jesslin Tan; Lin Cui; Raymond Tzer Pin Lin; Royce Ang; Samuel Loo; Yichen Ding; Zhenyang Zhou |
| EPI_ISL_13186247, EPI_ISL_13186619, EPI_ISL_13186641, EPI_ISL_13252794, EPI_ISL_13252801, EPI_ISL_13298834 | National Virus Reference Laboratory | National Virus Reference Laboratory | Charlene Bennett; Cillian F De Gascun; Gabriel Gonzalez; Jonathan Dean; Michael Carr; Zoe Yandle |
| EPI_ISL_12954169 | Nhlangano Health Centre | National Institute for Communicable Diseases of the National Health Laboratory Service | Amoako DG; Bhiman JN; Everatt J; Ismail A; Kekana D; Mahlangu B; Maphalala G; Mnguni A; Mohale T; Ntuli N; Scheepers C; Wolter N |
| EPI_ISL_13066528, EPI_ISL_13298415, EPI_ISL_13326086, EPI_ISL_13326108, EPI_ISL_13362168 | Originating lab: Wales Specialist Virology Centre Sequencing lab: Pathogen Genomics Unit | Public Health Wales Microbiology Cardiff Wales Specialist Virology Centre | Alec Birchley; Alexander Adams; Amy Gaskin; Angela Marchbank; Bree Gatica-Wilcox; Catherine Moore; Jason Coombes; Joanne Watkins; Joel Southgate; Johnathan Evans; Laura Gifford; Lauren Gilbert; Lee Graham; Malorie Perry; Matthew Bull; Nicole Pacchiarini; Sally Corden; Sara Kumziene-Summerhayes; Sara Rey; Sarah Taylor; Simon Cottrell; Sophie Jones; Tom Connor |
| EPI_ISL_12851724, EPI_ISL_13140805, EPI_ISL_13140809, EPI_ISL_13331766, EPI_ISL_13331779 | Outre Mer | Institut Pasteur | Angela Brisebarre; Camille Capel; Christophe Malabat; Corinne Maufrais; Etienne Simon-Lorière; Frédéric Lemoine; Julien Fumey; Louise Lefrançois; Marie-Hélène GLAUDON LOUVEAU DE LA GUIGNERAYE; Marion Barbet; Maud Vanpeene; Méline Bizard; Slim El Khiari; Sylvie Behillil; Sylvie Van der Werf; Vincent Enouf |
| EPI_ISL_13324310 | Outre Mer | National Reference Center for Viruses of Respiratory Infections, Institut Pasteur, Paris | Angela Brisebarre; Camille Capel; Christophe Malabat; Corinne Maufrais; Etienne Simon-Lorière; Frédéric Lemoine; Julien Fumey; Laurence FAGOUR; Louise Lefrançois; Marion Barbet; Maud Vanpeene; Méline Bizard; Slim El Khiari; Sylvie Van der Werf; Vincent Enouf |
| EPI_ISL_13137131 | PASIG CITY CHILDREN'S HOSPITAL | Research Institute for Tropical Medicine | Alexander Sadiasa; Amalea Dulcene D. Nicolasora; Angela Kae T. Chang; Anne Pauline A. Alpino; Ardiane Ysabelle M. Dolor; Catalino S. Demetria; Charalyn Babida; Claudette Lee S. Navarro; Criselda T. Bautista; Dodge R. Lim; Francisco Gerardo M. Polotan; Gerald Ivan S. Sotelo; Jefferson Earl J. Halog; Joseph Hughes; Joy Mariette L. Parayray; Katie Hampson; Kirstyn Brunker; Lei Lanna M. Dancel; Ma. Angelica A. Tujan; Ma. Ricci R. Gomez; Mayan Uy-Lumandas; Othoniel Jan T. Onza; Simon Daldry; Timothy John R. Dizon; Vina Lea F. Arguelles; Yao-Tsun Li |
| EPI_ISL_13355613 | PCR Laboratory, Divisional Head Quarters Teaching Hospital, Mirpur, AJ&K | Department of Virology, National Institute of Health, Islamabad, Pakistan | Aamer Ikram; Massab Umair; Muhammad Ammar; Muhammad Salman; Nazish Badar; Qasim Ali and Najma Majeed; Syed Adnan Haider; Zaira Rehman |
| EPI_ISL_13369867 | PRENETICS LIMITED | Hong Kong Department of Health | Alan K.L. Tsang; Edman T.K. Lam; Ken H.L. Ng; Patricia K. L. Leung; Rickjason C.W. Chan |
| EPI_ISL_13198072 | PSSE Lowicz | 1. Academic Center for Pathomorphological and Genetic-Molecular Diagnostics ltd, Bialystok, Poland 2. National Institute of Public Health - National Institute of Hygiene, Warsaw, Poland | Anetta Sulewska; Jacek Niklinski; Janusz DzięcioL; Joanna Kiśluk; Katarzyna Zacharczuk; Konrad Raczkowski; MaLgorzata Sadkowska-Todys; Magdalena Nowakowska; Piotr Karabowicz; Piotr Majewski; PrzemysLaw Biecek. Joanna Reszeć; RadosLaw Charkiewicz; Tomasz WoLkowicz |
| EPI_ISL_13371984 | Pardubicka nemocnice | University Hospital Hradec Kralove | Helena Parova; Lenka Rysava; Marketa Gancarcikova; Monika Berankova |
| EPI_ISL_12871865 | Pathcare | CERI, Centre for Epidemic Response and Innovation, Stellenbosch University and KRISP, KZN Research Innovation and Sequencing Platform, UKZN. | Anyaneji UJ; Claassen M; Giandhari J; Maharaj A; Maponga T; Moir M; Naidoo Y; Pillay S; Preiser W; San JE; Sanko TJ; Stander T; Tegally H; Tshiabuila D; Van Wyk S; Wilkinson E; Wilson S; de Oliveira T; van Zyl G |
| EPI_ISL_13231360 | Pathologist Lancet Kenya | KEMRI-Wellcome Trust Research Programme,Kilifi | Agoti C.; D.J.Nokes; Githinji G.; Lambisia A.; Makori T.; Mburu M.W.; Mohamed K.S.; Morobe J.; Mukadam R; Munoko A.; Ndwiga L.; Ngari C.; Ochola I; Ongera E.; de Laurent Z. |
| EPI_ISL_13107189 | Pathology Queensland and Forensic Scientific Services | Public Health Virology - Forensic and Scientific Services (PHV-FSS) | Son Nguyen on behalf of Q-PHIRE Genomics |
| EPI_ISL_13359899 | Plateforme de testing Namuroise | Plateforme de testing Namuroise | Degosserie Jonathan; Denis Olivier; Drugmand Jonathan; Janssens Louise; Laurent Hélène; Maschietto Céline; Mullier François; Otto Gaetan; Renguet Edith |
| EPI_ISL_13251640 | Platform BIS UZA/UAntwerpen | Labo Klinische Biologie, UZA | Basil Britto Xavier; Christine Lammens; Herman Goossens; Ines Verbesselt; Jasmine Coppens; Kathleen Holemans; Marie Le Mercier; Silke Liers; Veerle Matheeussen |
| EPI_ISL_13177706 | Public Health Authority of the Slovak Republic | Laboratory of Genomics and Bioinformatics, Comenius University Science Park | Anna Kaliňáková; Barbora Kotvasová; Diana Rusňáková; Elena Tichá; Jaroslav Budiš; Lucia Ševčíková; Miroslav Böhmer; Pavol Mišenko; Terézia Vrabľová; Tomáš Szemes |
| EPI_ISL_13149262, EPI_ISL_13149283, EPI_ISL_13149290, EPI_ISL_13149292, EPI_ISL_13149299, EPI_ISL_13149302, EPI_ISL_13149306, EPI_ISL_13149324 | | | |
| see above | Public Health Laboratory: COVID-19 Lab | International Livestock Research Institute | Collins Muli; Daniel Ouso; Edward Kiritu; Edward O. Abworo; Gilbert Kibet; Gugu Maphalala; Mncedisi Hlophe; Nomcebo Phungwayo; Patrick Amoth; Paul Dobi; Samuel O. Oyola; Shebbar Osiany; Siphesihle Langwenya; Sonal P. Henson; Susan Kamalizeni; Vishvanath Nene |

| | | | |
|---|---|---|---|
| EPI_ISL_13053559, EPI_ISL_13053622, EPI_ISL_13135210, EPI_ISL_13270696, EPI_ISL_13270939 | Public Health Ontario Laboratory | Public Health Ontario Laboratory | Aimin Li; Alex Marchand-Austin; Andre Villegas; Anna Puzinovici; Ashleigh Sullivan; Brandon Ye; Candice Schreiber; Carla Duncan; Christina Rampertab; Christine Seah; Claudia Chu; Dean Maxwell; Dhiraj Gaglani; Doonia Bajovic; Esther Nagai; Fatemeh Shaeri; Fatima Merza; Grace Jeong; Hadia Hussain; Himeshi Samarsinghe; Jacob Afelskie; Jason Iraheta; Jesse Wang; John Palmer; Karthikeyan Sivaraman; Kirby Cronin; Lisa Kim; Lisa McTaggart; Maria Mariscal; Mark Horsman; Narisha Shakuralli; Nataliya Potapova; Natasha Sing; Nobish Varghese; Philip Banh; Rachelle DiTullio; Rebecca Azzaro; Rima Palencia; Samir N Patel; Sarah Teatero; Semra Tibebu; Sophie Yu; Surendra Kumar; Sushma Kavikondala; Vincent Su Bin Cha; Zarah Rajaei |
| EPI_ISL_13249923, EPI_ISL_13249972 | Puskesmas Pasirkaliki | West Java Health Laboratory; School of Life Sciences and Technology, Institut Teknologi Bandung | Azzania Fibriani; Cut Nur Cinthia Alamanda; Ema Rahmawati; Hadiana; Karimatu Khoirunnisa; Miftahul Faridl; Rifky Waluyajati Rachman; Rini Robiani; Ryan Bayusantika Ristandi |
| EPI_ISL_13261955, EPI_ISL_13291443, EPI_ISL_13316310 | Quest Diagnostics Incorporated | Centers for Disease Control and Prevention Division of Viral Diseases, Pathogen Discovery | A. Gerasimova; A. Perez; B. Anderson; Benjamin Rambo-Martin; Christopher Gulvick; Clinton Paden; Dakota Howard; Dhwani Batra; Duncan MacCannell; Erisa Sula; F. Lacbawan; I. Shlyakhter; Jason Caravas; K. Livingston; Kristine Lacek; L. Bernstein; M. Hua; Matthew Schmerer; P. Tanpaiboon; Peter Cook; R. Kagan; R. Owen; R. Rolando; S. Rosenthal; Scott Sammons; Shatavia Morrison; Tymeckia Kendall; Victoria Caban Figueroa; Y. Liu; Yvette Unoarumhi |
| EPI_ISL_12704285, EPI_ISL_12704369, EPI_ISL_12810659, EPI_ISL_12810724, EPI_ISL_13157528 | Regional Virus Laboratory, Belfast Health and Social Care Trust; and: Genomics Core Technology Unit, Queen's University Belfast. | COVID-19 Genomics UK (COG-UK) Consortium | Alan; Alison Watt; Arun Mahesh; BHSCT]: Conall McCaughey; Ciara Cox; Clara Radulescu; David Simpson; Deborah Lavin; Derek Fairley; Evan Troendle; Fiona Rogan; James McKenna; Jana Gazdova; Julia Miskelly; Mairead Connor; Miao Tang; QUB]: Marc Fuchs; Rice; Sarah Sonner; Stephen Bridgett; Susan Feeney; Syed Umbreen; Tanya Curran; Timofey Skvortsov; Zoltan Molnar; [Genomics Core Technology Unit; [Regional Virus Laboratory |
| EPI_ISL_13298307 | Respiratory Virus Unit, Microbiology Services Colindale, Public Health England | COVID-19 Genomics UK (COG-UK) Consortium | PHE Covid Sequencing Team |
| EPI_ISL_12838667, EPI_ISL_12920374, EPI_ISL_13106591, EPI_ISL_13231623, EPI_ISL_13242371, EPI_ISL_13311352, EPI_ISL_13311641, EPI_ISL_13311704, EPI_ISL_13338284 | | | |
| see above | Rosalind Franklin Laboratory | Wellcome Sanger Institute for the COVID-19 Genomics UK (COG-UK) Consortium | Cordelia Langford; David K. Jackson; Dominic Kwiatkowski; Donald Fraser; Ewan Harrison; Ian Johnston; Jeffrey Barrett; John Sillitoe on behalf of the Wellcome Sanger Institute COVID-19 Surveillance Team; Rob Howes; Roberto Amato; Sonia Goncalves; Suki Lee; The Rosalind Franklin Laboratory and Alex Alderton |
| EPI_ISL_13249967 | Rumah Sakit TK II Dustira | West Java Health Laboratory; School of Life Sciences and Technology, Institut Teknologi Bandung | Azzania Fibriani; Cut Nur Cinthia Alamanda; Ema Rahmawati; Hadiana; Karimatu Khoirunnisa; Miftahul Faridl; Rifky Waluyajati Rachman; Rini Robiani; Ryan Bayusantika Ristandi |
| EPI_ISL_13262714, EPI_ISL_13371523, EPI_ISL_13371628, EPI_ISL_13371646 | SALUD DIGNA | Instituto Nacional de Medicina Genomica | Abraham Campos-Romero; Cedro-Tanda A; Escobar-Arrazola MA; Garcia-Garcia FE; Garnica-Lopez Dora; Herrera-Montalvo LA.; Hidalgo-Miranda A; Luna-Ruiz Marco; Mendoza-Vargas A; Moreno-Camacho José Luis; Ramirez-Vega O; Rangel-DeLeon D; Reyes-Grajeda JP; Rodriguez-Gallegos Jorge; Sanchez-Xochipa S; Yair Alfaro-Mora |
| EPI_ISL_13292855, EPI_ISL_13301003 | SARS-CoV-2 Sequencing Castilla y Leon-Spain Consortium | SARS-CoV-2 Sequencing Castilla y Leon-Spain Consortium | Antonio Orduña-Domingo; Carlos Fuster Foz; Carmen Aldea-Mansilla; Carmen Gimeno Crespo; David Abad; Gabriel March Rosello; Gregoria Megías Lobón; Jose María Eiros Bouza; M. Isabel Fernandez-Natal; Marta Dominguez-Gil; Marta Hernandez; María Antonia García Castro; Mª Fe Brezmes-Valdivieso; Noelia Arenal Andrés; Silvia Rojo; Sonsoles Garcinuño Pérez |
| EPI_ISL_13027465, EPI_ISL_13167988, EPI_ISL_13217382, EPI_ISL_13217556, EPI_ISL_13217726, EPI_ISL_13217784 | SARS-CoV-2 testing team, National Institute of Infectious Diseases | Pathogen Genomics Center, National Institute of Infectious Diseases | Hazuka Y Furihata; Kentaro Itokawa; Makoto Kuroda; Masanori Hashino; Masumichi Saito; Naomi Nojiri; Nozomu Hanaoka; Rina Tanaka; Tsuguto Fujimoto; Tsuyoshi Sekizuka |
| EPI_ISL_13372121 | SC (UCO) Igiene e Sanità Pubblica, ASUGI, Trieste | SC (UCO) Igiene e Sanità Pubblica, ASUGI, Trieste | Basaglia G; Busetti M; D'Agaro P; Fontana F; Forciniti G; Koncan R; Pipan C; Piscianz E; Segat L |
| EPI_ISL_12863427, EPI_ISL_13369107 | SESARAM | Instituto Nacional de Saude Doutor Ricardo Jorge (INSA) | Borges et al |
| EPI_ISL_12903160, EPI_ISL_13076019 | SYNLAB | University Hospital Brno, CMBG | Bezdicek Matej; Dolejska Monika; Kristyna Dufkova; Lengerova Martina; Svaton Jan |
| EPI_ISL_12190609, EPI_ISL_13143504 | SYNLAB MVZ Weiden | Robert Koch Institute | |
| EPI_ISL_13155840 | Servicio Virosis Respiratorias-Departamento Virología-INEI | Instituto Nacional Enfermedades Infecciosas C.G.Malbran | Avaro M.; Baumeister E.; Benedetti E.; Campos J.; Cisterna D.; Dattero ME; De Belder D.; Haim MS.; Mallou F.; Molina V.; Perandones C.; Poklepovich T.; Pontoriero A.; Russo M.; Sanchez Loria J.; Tuduri E. |
| EPI_ISL_13250911 | Servicio de Microbiologia Hospital Ramon y Cajal | Servicio de Microbiologia Hospital Ramon y Cajal | Galan JC; Martinez-García L.; Ponce-Alonso M |
| EPI_ISL_13302920, EPI_ISL_13303076, EPI_ISL_13313392, EPI_ISL_13313420, EPI_ISL_13313628, EPI_ISL_13316622, EPI_ISL_13316807 | | | |
| see above | Shamir Medical Center (Asaf Harofe) | Shamir Medical Center (Asaf Harofe) | Abu Hamad Ramzia; Adina Bar Chaim; Anna Vishnevsky; Chen Weiner; Nir Rainy; Patricia Benveniste-Lekovitz; Reut Sorek Abramovich; Yevgeni Yegorov |
| EPI_ISL_13208025 | Sirindhorn Hospital | Medical Genomic Centre,Medical Life Sciences Institute,Department of Medical Sciences, Ministry of Public Health, Thailand | Archawin Rojanawiwat; Jirapha Pakdee; Naphatcha Thawong; Natthakul Bunneang; Nuanjun Wichukchinda; Pilailuk Akkapaiboon Okada; Pundharika Piboonsiri; Surakameth Mahasirimongkol; Waritta Sawaengdee |
| EPI_ISL_13339041 | Stadtspital Triemli | Department of Biosystems Science and Engineering, ETH Zürich | Alexandra Trkola; Chaoran Chen; Christian Beisel; David Dreifuss; Elodie Burcklen; Franziska Singer; Guido Bloemberg; Ina Nissen; Ivan Topolsky; Kevin Steiner; Kim Philipp Jablonski; Lara Fuhrmann; Louis du Plessis; Maryam Zaheri; Matteo Carrara; Michael Huber; Mirjam Feldkamp; Natascha Santacroce; Niko Beerenwinkel; Pelin Burcak Icer; Rebecca Denes; Riccarda Capaul; Sarah Nadeau; Shuqing Yu; Stefan Schmutz; Tanja Stadler; Tobias Schär; Verena Kufner |
| EPI_ISL_13273973 | SuperCare Medical Services, Inc | Research Institute for Tropical Medicine | Alexander Sadiasa; Amalea Dulcene D. Nicolasora; Angela Kae T. Chang; Anne Pauline A. Alpino; Ardiane Ysabelle M. Dolor; Catalino S. Demetria; Charalyn Babida; Claudette Lee S. Navarro; Criselda T. Bautista; Dodge R. Lim; Francisco Gerardo M. Polotan; Gerald Ivan S. Sotelo; Jefferson Earl J. Halog; Joseph Hughes; Joy Mariette L. Parayray; Katie Hampson; Kirstyn Brunker; Lei Lanna M. Dancel; Ma. Angelica A. Tujan; Ma. Ricci R. Gomez; Mayan Uy-Lumandas; Othoniel Jan T. Onza; Simon Daldry; Timothy John R. Dizon; Vina Lea F. Arguelles; Yao-Tsun Li |
| EPI_ISL_13184257 | Swedish national genomic surveillance program of SARS-CoV-2 | The Public Health Agency of Sweden | Alma Brolund; Emmi Andersson; Maria Lind Karlberg; Swedish national genomic surveillance program of SARS-CoV-2 |
| EPI_ISL_13313928 | Switch Health | National Microbiology Laboratory (NML) | Adrian Zetner; Anna Majer; Anneliese Landgraff; CanCOGeN's metadata curation team; Carmen Lia Murall; Chanchal Yadav; Connor Chato; Darian Hole; Elsie Grudeski; Emily Haidl; Gary Van Domselaar; Gordon Jolly; Grace Seo; Jeff Tuff; Jennifer Tanner; Katherine Eaton; Kirsten Biggar; Kristyn Burak; Madison Chapel; Morag Graham; Natalie Knox; Nathalie Bastien; Philip Mabon; Public Health Agency of Canada's CCGP and Scientific Informatics Services team; Rhiannon Huzarewich; Russell Mandes; Shari Tyson; Timothy Booth; Yan Li |
| EPI_ISL_13029236, EPI_ISL_13029256, EPI_ISL_13066210, EPI_ISL_13133837, EPI_ISL_13133857, EPI_ISL_13133875, EPI_ISL_13133904 | | | |
| see above | Synlab Eesti OÜ | 1. Laboratory of Communicable Diseases (Estonia); 2. Eurofins Genomics Europe Sequencing GmbH | Abroi A.; Avi R.; Dotsenko L.; Epštein J.; Hoidmets D.; Huik K.; Härma M-A.; Jaaniso E.; Kaarna K.; Kallas E.; Koppel I.; Kuzmin I.; Lahesaare A.; Lutsar I.; Metspalu M.; Milani L.; Naaber P.; Niglas H.; Oopkaup O.E.; Pauskar M.; Peterson H.; Päll T.; Ratnik K.; Raudvere U.; Reisberg T.; Sadikova O.; Sepp H.; Shablinskaja A.; Suija H.; Talas U.G.; Truusalu K. |
| EPI_ISL_13369821 | Temporary Specimen Collection Centre at the AsiaWorld-Expo | Hong Kong Department of Health | Alan K.L. Tsang; Edman T.K. Lam; Ken H.L. Ng; Patricia K. L. Leung; Rickjason C.W. Chan |
| EPI_ISL_12954411 | The Luke Commission Hospital | National Institute for Communicable Diseases of the National Health Laboratory Service | Amoako DG; Bhiman JN; Everatt J; Ismail A; Kekana D; Mahlangu B; Maphalala G; Mnguni A; Mohale T; Ntuli N; Scheepers C; Wolter N |
| EPI_ISL_13156680 | Tokyo Metropolitan Institute of Public Health | Tokyo Metropolitan Institute of Public Health | Ai Suzuki; Akane Negishi; Arisa Amano; Fumi Kasuya; Hirofumi Miyake; Kenji Sadamasu; Kenshiro Kuroki; Mami Nagashima; Maya Isogai; Ryota Kumagai; Sachiko Harada; Takushi Fujiwara |
| EPI_ISL_13369104 | ULS Castelo Branco | Instituto Nacional de Saude Doutor Ricardo Jorge (INSA) | Borges et al |
| EPI_ISL_13345811, EPI_ISL_13345825, EPI_ISL_13345829, EPI_ISL_13345841 | Unilabs Eskilstuna Laboratorium | Unilabs Eskilstuna Laboratorium | Emma Arvidsson |
| EPI_ISL_13372013, EPI_ISL_13372032, EPI_ISL_13372036 | University Hospital Hradec Kralove | University Hospital Hradec Kralove | Helena Parova; Lenka Rysava; Marketa Gancarcikova; Monika Berankova |
| EPI_ISL_13202480, EPI_ISL_13202485 | University Hospital Ostrava | University Hospital Ostrava | Chorzempa; Matějová; Špulerová |
| EPI_ISL_13202591 | University Hospitals of Geneva, Laboratory of Virology | HUG, Laboratory of Virology and the Health2030 Genome Center | Aline Mamin; Ana Rita Goncalves; Cedric Howald; Deborah Penet; Francisco Perez; Henri Pegeot; Ioannis Xenarios; Keith Harshman; Laurent Kaiser; Lorenzo Cerutti; Melyssa Elies; Samuel Cordey |
| EPI_ISL_13286168 | Utah Public Health Laboratory | Utah Public Health Laboratory | Erin L. Young; John Arnn; Kelly F. Oakeson; Olinto Linares-Perdomo; Pooja Gupta; Tom Iverson |
| EPI_ISL_13343324, EPI_ISL_13343396, EPI_ISL_13343462 | Viollier AG | Department of Biosystems Science and Engineering, ETH Zürich | Andrea Patrizia Salzmann; Chaoran Chen; Christian Beisel; Christiane Beckmann; Christoph Noppen; David Dreifuss; Elodie Burcklen; Franziska Singer; Henriette Kurth; Ina Nissen; Ivan Topolsky; Kim Philipp Jablonski; Lara Fuhrmann; Louis du Plessis; Matteo Carrara; Maurice Redondo; Mirjam Feldkamp; Natascha Santacroce; Niko Beerenwinkel; Olivier Kobel; Pelin Burcak Icer; Rebecca Denes; Sarah Nadeau; Sebastian Kurscheid; Shuqing Yu; Tanja Stadler; Tobias Schär |
| EPI_ISL_13228472 | ZNPHL | CHAZ Laboratory | CHAZ Lab Staff; Chipango. C; Muyombo. A; Sandala. D; Shempela. D; Sikalima. J |
